# Genetic architecture of lumbar spinal stenosis

**DOI:** 10.1101/2024.10.16.24315641

**Authors:** Ville Salo, Juhani Määttä, Jasmin Takala, Anni Heikkilä, FinnGen, Ene Reimann, Reedik Mägi, Estonian Biobank Research Team, Kadri Reis, Abdelrahman G.Elhanas, Anu Reigo, Priit Palta, Tõnu Esko, Ville Leinonen, Jaro Karppinen, Eeva Sliz, Johannes Kettunen

**Author notes:** A list of FinnGen authors and their affiliations available at the end of the paper. A list of Estonian Biobank Research Team authors and their affiliations available at the end of the paper. These authors contributed equally to this work (Eeva Sliz, Johannes Kettunen).

## Abstract

**Introductory paragraph:** Over 100 million people worldwide suffer from lumbar spinal stenosis (LSS) with increasing incidence with ageing population, yet little is known about the LSS genetic background. Given the high cost of treating LSS, a deeper understanding of LSS pathogenesis may eventually result in the development of novel preventative and treatment methods, potentially leading to reductions in related societal costs. Our aim is to gain a better understanding of the genetic components underlying LSS. In the FinnGen, Estonian, and UK biobanks, we conduct a genome-wide association study (GWAS) of LSS and merge the results in the genome-wide meta-analysis. In addition to the seven known risk loci, our meta-analysis reveals 47 loci that have not been associated with LSS in previous studies. Many downstream analyses and multiple candidate genes discovered from the LSS-associated loci suggest that spinal degeneration plays a major role in the pathogenesis of LSS.

## Main text

Lumbar spinal stenosis (LSS) narrows the lumbar central spinal canal or foramina causing symptoms mainly to buttocks and lower extremities^1,2^. Term central stenosis is used if the whole central spinal canal is compressed, and foraminal or lateral stenosis if the single nerve root exiting the neural foramina is compressed^1^. LSS usually develops with ageing when lumbar structures degenerate, including changes in intervertebral discs, facet joints, ligamentum flavum, and vertebrae. In addition, spondylolisthesis can cause or aggravate LSS^1^. Even though the majority of LSS cases have a degenerative origin, in a smaller subgroup of patients, LSS arises from developmental or congenital factors^2,3^. Naturally, developmental LSS can predispose individuals to degenerative LSS as they age^3^. Developmental causes include a congenitally small central canal and Paget disease, which leads to atypical bone growth^1,3^.

The prevalence of LSS has been estimated to be around 11% in the general population and 20% among people over 60 years, varying greatly with age, and with the majority of cases being asymptomatic^1,4^. LSS becomes clinically significant when it causes symptoms affecting daily life. The main symptoms include discomfort, pain, fatigue, paresthesia, and/or numbness in the lower extremities or the buttocks, which exacerbate during activities such as walking, frequently called as neurological claudication^1,2,4^. Low back pain can be present or absent^2^. The symptoms are typically relieved during sitting or forward flexion^1,2^. Diagnosis is based on patient history and symptoms, and clinical examination, with confirmation through imaging, usually magnetic resonance imaging (MRI) or computed tomography (CT)^1^. Non-surgical management, such as physiotherapy, oral medication, and activity modification, is usually the first-line treatment. Surgical treatment is warranted if the symptoms are severe or non-surgical care does not improve the symptoms enough^1,2^.

In this study, we used the International Classification of Diseases (ICD)-10 code M48.0 to characterize LSS in the genome-wide meta-analysis. Data from FinnGen, the Estonian Biobank, and the UK Biobank were analyzed, comprising 30 269 cases and 739 414 controls (Table S1). A total of 54 loci with at least one genome-wide significant (P<5×10^−8^) variant were associated with LSS in the meta-analysis. Forty-seven of the loci were previously unreported (Table 1, Supplementary Data), and we also successfully replicated seven loci previously associated with LSS (Fig. 1A, Table S2-S4 Fig. S1)^5–7^. However, 17 of the newly identified loci had been previously associated with highly related phenotypes, such as back pain (BP) or lumbar disc herniations (LDH) (Table 1). In the conditional analysis, we also detected secondary signals at four loci (Table S2). At locus 12p11.22, we observed several secondary signals, and the association signal extended beyond the 2MB window used to define loci (Supplementary Data 29).

**Table 1.**
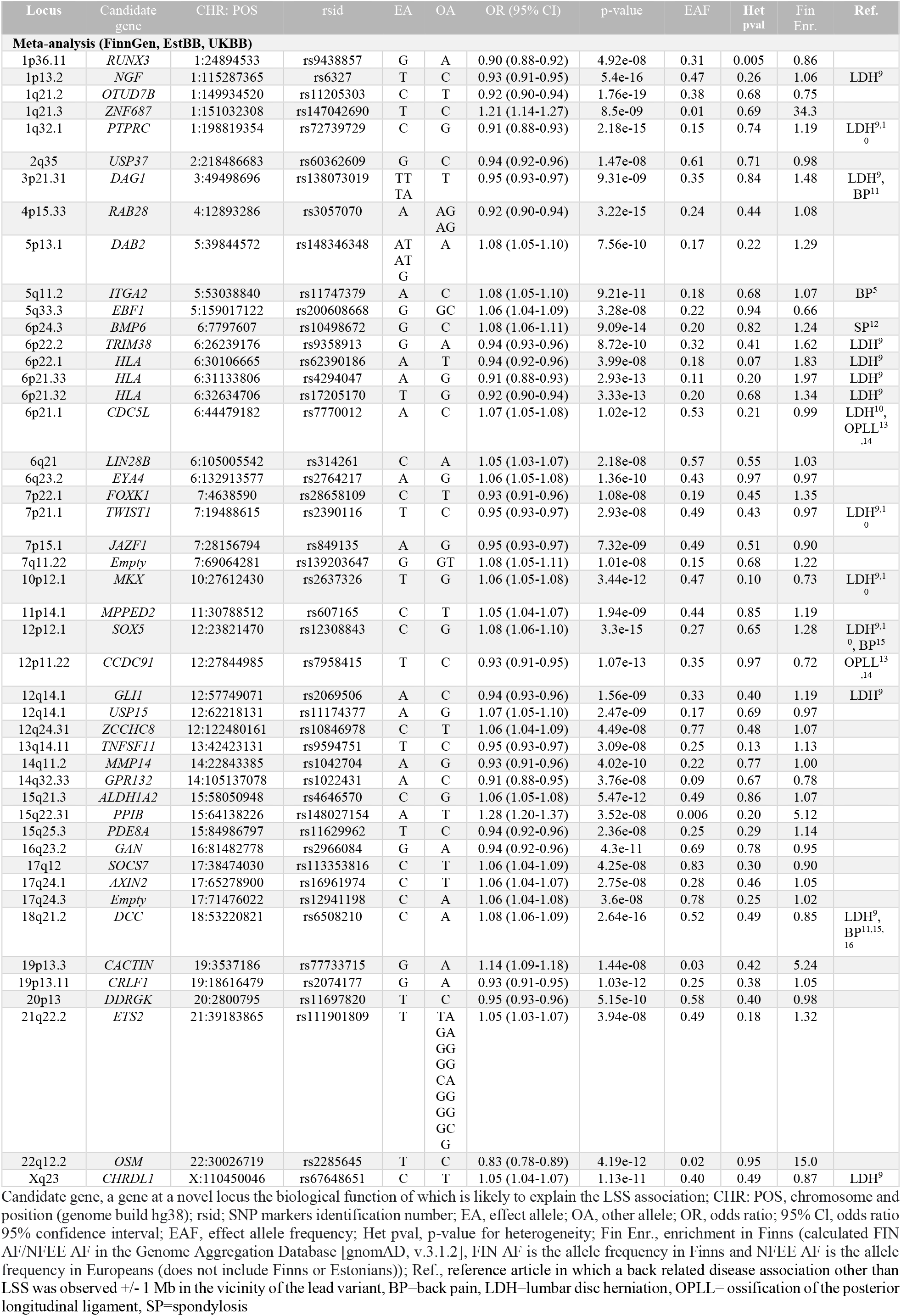
The lead variants at 47 novel LSS-associated (p<5×10^−8^) loci. In the meta-analysis, there were a total of 30 269 cases and 739 414 controls from FinnGen, the Estonian Biobank, and the UK Biobank. All genome-wide significantly associated loci observed in the meta-analysis are presented in Table S2.

**Fig. 1.**
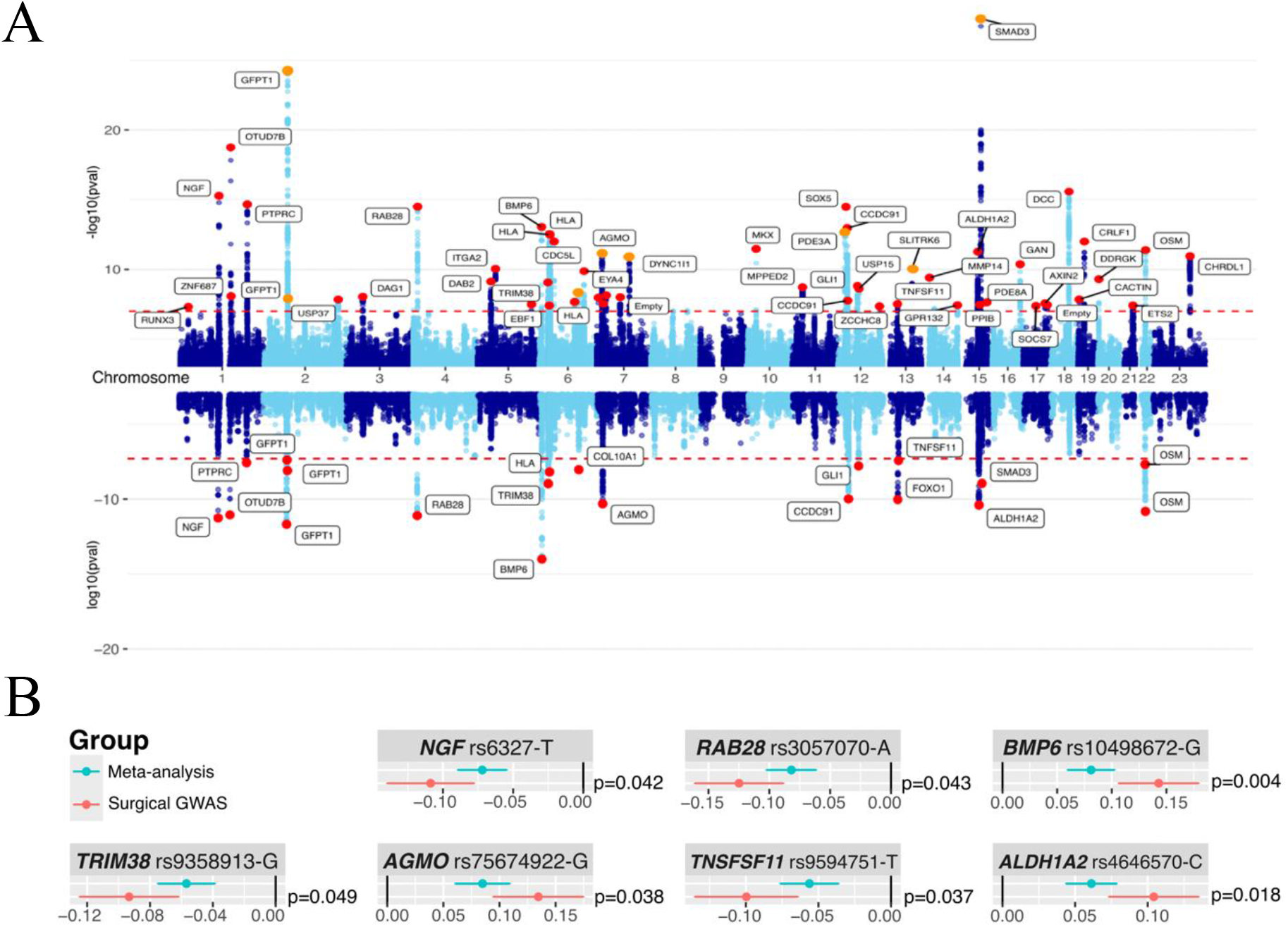
**A)** Above, the Manhattan plot of the associations detected to be associated with LSS in the meta-analysis of 30 269 cases and 739 414 controls. Previously reported loci are indicated with orange color, while 47 novel loci that we observed are highlighted in red. Candidate genes possibly explaining the LSS associations were used as loci identifiers. The red dashed line depicts the genome-wide significance limit (p<5×10^−8^). Below, the Manhattan plot of the associations identified in the surgical GWAS conducted in FinnGen (8627 LSS cases requiring surgery and 270 773 controls). **B)** A comparison of the effect estimates of the lead variants discovered in the original meta-analysis (Meta-analysis, ICD-10:M48.0 [green]) and in sensitivity analysis with patients requiring surgery (Surgical GWAS, *Nomesco v1*.*15*: ABC36, ABC56, ABC66, ABC99, NAG61, NAG62, NAG63, NAG66, NAG67, and NAG99 [red]). Significant differences (p<0.05) in the effect estimates were observed for 7 variants. Dots indicate effect size and vertical lines are the corresponding 95% confidence intervals. For effect differences statistical comparison, we used a two-tailed test, using group-specific effect estimates of the variants and the corresponding standard errors *((Effect_Meta-Effect_Surg)/ sqrt(standarderror_Meta*^*2*^*+standarderror_Surg*^*2*^)), *p-value=2*(1-diff)*. The effect size comparisons of all variants are given in Table S5, with a graphical illustration shown in Fig. S2.

We estimated LD score regression-derived SNP-based heritability to be 0.18 (standard error [SE] = 0.0098), suggesting genetic factors account for 18% of the common variation in LSS risk. The genomic inflation factor lambda (1.40) suggested inflation in the test statistics. Considering the observed intercept value of 1.11, the inflation is likely attributed to a polygenic signal and residual population stratification not fully controlled by principal component adjustments^8^. Based on FinnGen data alone, heritability was estimated to be 21% [SE]=0.0141, lambda 1.34, intercept 1.13. Our heritability estimates were considerably higher compared to the previous SNP-based heritability estimate of 10.1% [SE]=0.0412, lambda 1.03, intercept 0.99 (https://nealelab.github.io/UKBB_ldsc/h2_summary_M13_SPINSTENOSIS.html), also based on FinnGen. Our estimates were calculated using a larger sample, which is likely the main reason for the difference.

In the sensitivity analysis performed at FinnGen, focusing on LSS patients requiring surgical treatment, we identified 20 genome-wide significant loci (Fig. 1A, Table 2). The surgical cases were required to have procedure codes of *Nomesco v1*.*15*: ABC36, ABC56, ABC66, ABC99, NAG61, NAG62, NAG63, NAG66, NAG67, and NAG99 (see details in Table S1). We compared the original meta-analysis with the sensitivity analysis on LSS patients requiring surgery and observed differences in the effect estimates for seven variants, indicating larger effect estimates in the surgical cohort (Fig. 1B, Fig. S2, Table S5).

**Table 2.**
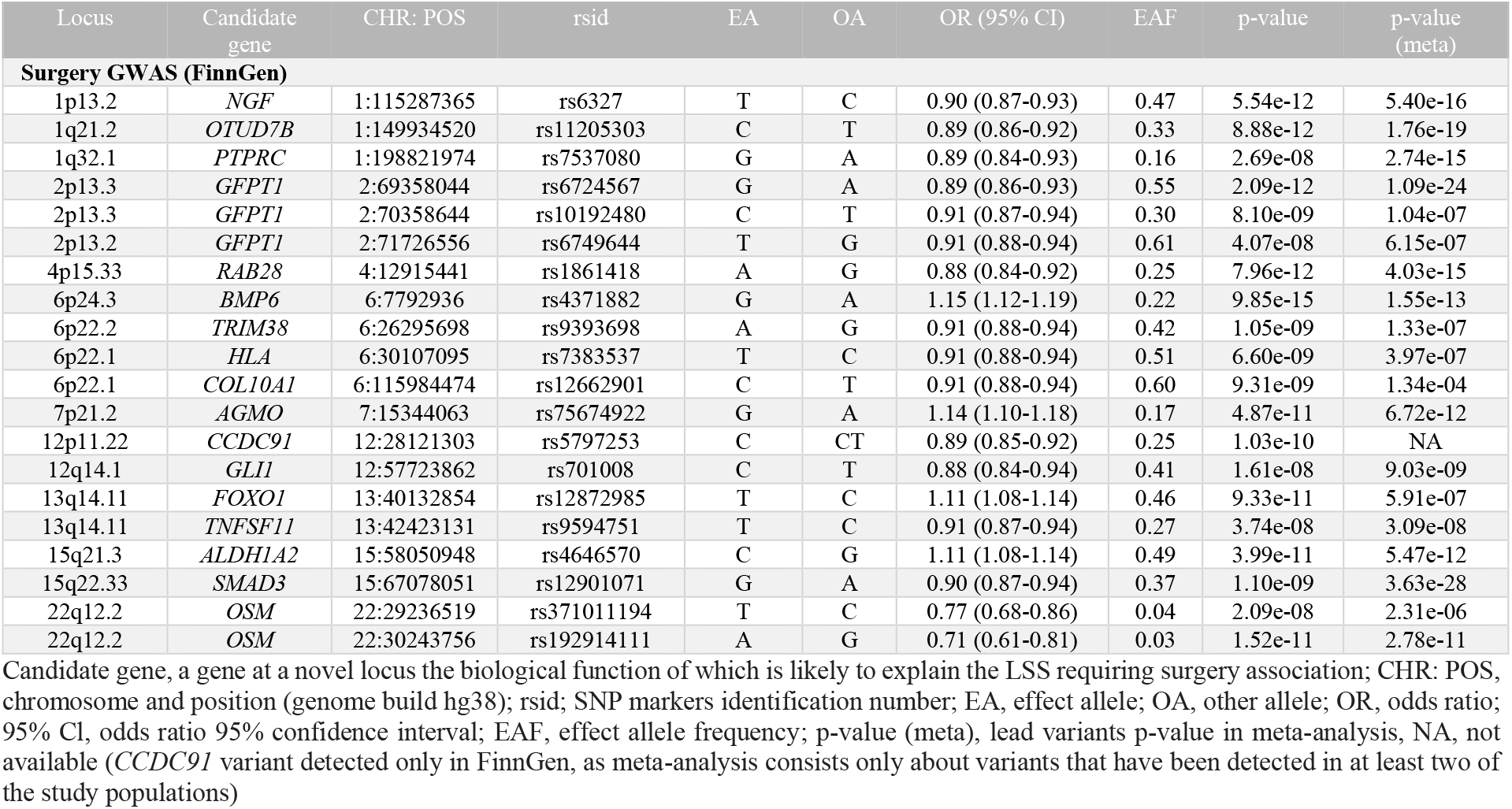
The lead variants at 20 loci associated (p<5×10^−8^) with LSS requiring surgical treatment. The surgery GWAS involved patients with LSS who had undergone surgery (*NOMESCO version 1.15*, ABC36, ABC56, ABC66, ABC99, NAG61, NAG62, NAG63, NAG66, NAG67, and NAG99). A total of 8627 cases and 270 773 controls were included in the surgery GWAS. LSS cases that lacked documentation of the surgical codes were excluded from the analysis. Patients who had undergone surgery but did not have a LSS diagnosis were excluded, as they likely had an acute injury.

LSS is suggested to be caused by degenerative changes in the spine^17^, and we identified several potential genes that may account for these degenerative changes. In our PheWAS analysis, we found that variants at LSS-associated loci were also associated with numerous other spine-related diseases, such as spondylosis and lumbar disc herniations (Table S3).

Of particular interest was the locus close to *BMP6* (*bone morphogenetic protein 6*). *BMP6* has previously been associated with spondylosis^12^ and osteoarthritis^18^ and has been found to play an important role in bone and cartilage formation and repair^19^. At this locus, the effect estimate was significantly larger in the LSS surgery GWAS compared to the primary LSS meta-analysis (Fig. 1B). Other examples of interesting LSS-associated loci include the ones near *CDC5L* (*cell division cycle 5 like*) and *CCDC91* (*coiled-coil domain containing 91*), which have previously been associated with the ossification of the posterior longitudinal ligament (OPLL)^13,14^. OPLL is characterized by ectopic bone formation within the posterior longitudinal ligament in the spinal canal. OPLL may also contribute to the pathogenesis of LSS, as ossified ligament can compress the spinal cord or nerve roots^20,21^. Another interesting LSS-associated locus is the one near *MMP14* (*matrix metallopeptidase 14*). The lead variant is a missense variant (rs1042704, p.D273N) known to significantly reduce collagen catabolism^22^, which might have an impact on spinal degeneration, where modifications in collagen structures are often observed^23^. However, based on our PheWAS results (Table S3) and the previously reported associations with Dupuytren contracture^24^ and adhesive capsulitis^25^, the *MMP14* gene may not be a suitable target for translational research, as the rs1042704 variant appears to protect from LSS while increasing the risk of the aforementioned disorders.

The loci we observed in surgical GWAS were largely overlapping with the loci we observed in the meta-analysis. Only the locus near *FOXO1 (forkhead box O1)* was specific to the surgical GWAS (Table S3).

Multiple genes that we identified as potential candidate genes at the novel LSS-associated loci were also highlighted by the GCTA-fastBAT analysis and the MAGMA gene-based test (Table 1, Fig. 2A, Table S6). The pathway responsible for sequence-specific DNA binding showed the most significant enrichments in the MAGMA gene-set analysis (Fig. 2B). Gene-set analysis also highlighted pathways associated with ossification and pathways controlling biosynthetic processes and differentiation. Additionally, enrichments were found in many gene-sets related to the nervous system, such as the pathway related to the binding of neurotrophin receptors. There were no tissues identified in the MAGMA tissue expression analysis (Fig. S3) that exhibited a positive correlation between associations with LSS and tissue-specific gene expression profiles. The lack of the relevant tissues— namely bone and cartilage—in the Genotype-Tissue Expression (GTEx) dataset utilized for these analyses is likely the reason for the null result.

**Fig. 2.**
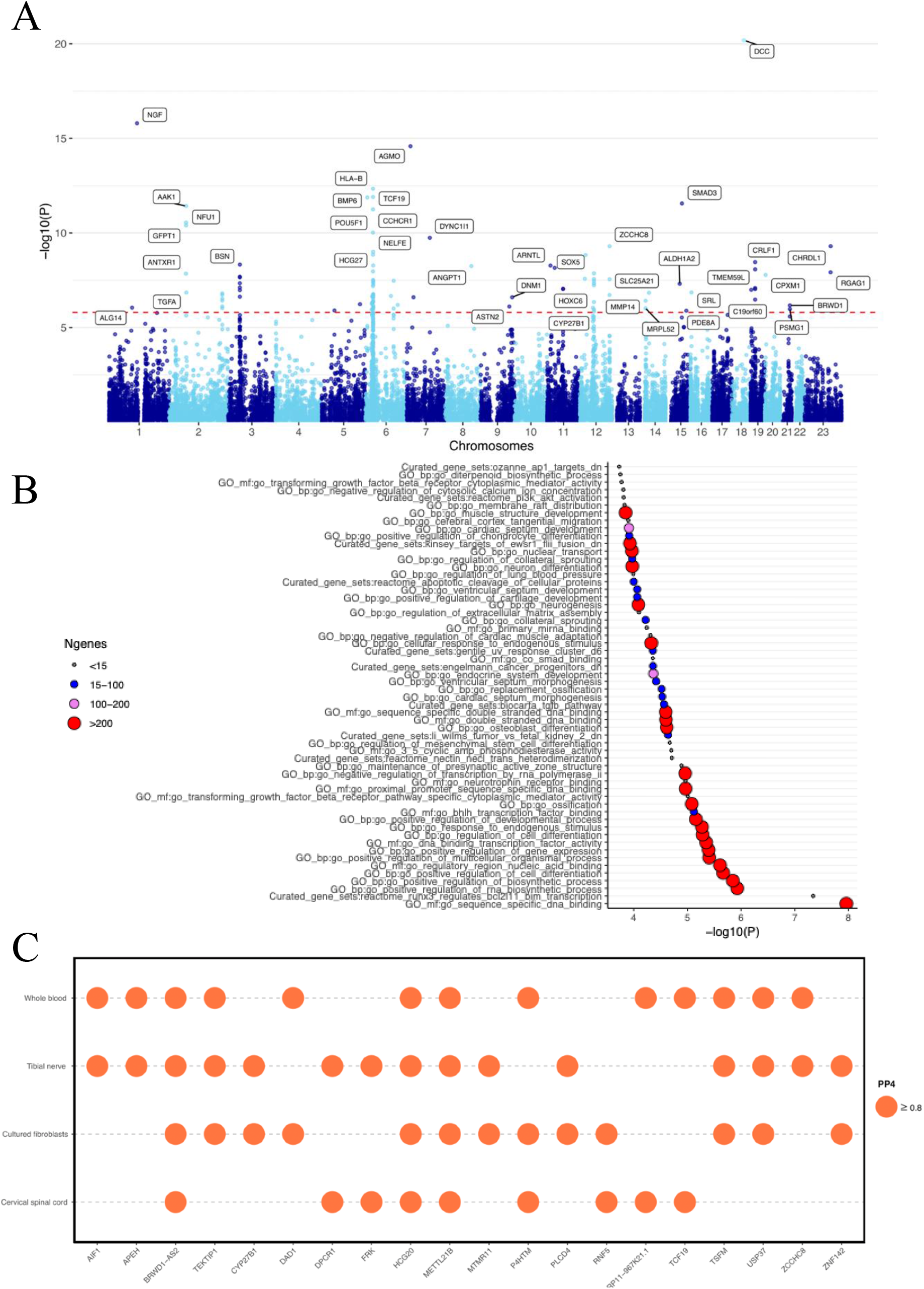
**A)** Results of the MAGMA^26^ gene-based test in a Manhattan plot. X-axis chromosomes, y-axis -log10(p-value). **B)** MAGMA gene-set enrichment analysis. The plot shows significantly enriched pathways (pFDR < 0.05), curated gene sets, and GO-annotations ranked by significance (-log10(P-value)). The circle size refers to the size of the gene set. Small gray <15, blue 15–100, violet 100-200, and red >200 genes. The analysis was done using FUMA^27^, and the gene sets and GO annotations included in the analysis are from MSigDB^28^. **C)** Colocalizations between LSS GWAS and eQTL signals were estimated with approximate Bayesian factor analyses, implemented with the ‘coloc.abf’ function available in the ‘coloc’ R-library (Table S7). The ‘biomaRt’ R-library was used to extract genes from Emsembl archives^29^ within a 1 MB (± 500000 bases) window surrounding each lead variant. Colocalizations were investigated for 754 genes that were available in GTEx. Variant-gene expression associations for selected tissues (whole blood, cultured fibroblasts, tibial nerve, and cervical spinal cord) were downloaded from GTEx v8. Colocalizations with posterior probabilities ≥ 0.8 for the variant were considered significant^30^. Only genes with at least two significant colocalizations are shown in the plot; all significant colocalizations are listed in Table S7.

Colocalizations between LSS GWAS and gene expression signals (eQTL) were observed in each selected tissue, namely whole blood, cultured fibroblasts, tibial nerve, and cervical spinal cord. We found evidence of the colocalization of the expression of the 85 genes and LSS signals in at least one tissue (posterior probability [PP] for the shared variant ≥ 0.8, Table S7), and 20 colocalizations were observed in at least two or more of the tissues investigated (Fig. 2C). For example, significant colocalizations between LSS association signals and *BRWD1-AS2* (*BRWD1 antisense RNA2*), *HCG20* (*HLA complex group 20*) and *METTL21B* (*EEF1A lysine methyltransferase 3*, also known as *EEF1AKMT3*) in all studied tissues (Fig. 2C, Table S7).

LSS usually manifests later in life^1^. Before our study, an age-of-onset analysis for genetic variants was not possible since the genetic background of the LSS has not been investigated extensively and there have been limitations in the availability of large, longitudinal datasets. At each locus associated with LSS, we examined the differences in the cumulative incidences between homozygotes for the lead variants and calculated the cumulative incidence of LSS diagnoses. Around age 40, the number of LSS diagnoses started to accumulate, with the majority of diagnoses occurring between the ages of 50 and 70. All lead variants showed a similar trend in terms of the accumulation of LSS diagnoses, with the lead variant genotypes showing only slight variation and cumulative morbidities following the sample prevalence value (6.5% for LSS in FinnGen, Table S8). We observed that for five LSS-associated variants the cumulative incidence differed between homozygotes already at the age of 40 (Table S8). These lead variants located near the *OTUD7B* (*OTU deubiquitinase 7B*), *USP37* (*ubiquitin specific peptidase 37*), *CDC5L* (*cell division cycle 5 like*), *SOX5* (*SRY-box transcription factor 5*) and *SMAD3* (*SMAD family member 3*) genes (Fig. S4). With the majority of the other variants, the difference in the cumulative LSS diagnoses between homozygotes manifested at a later age, but with some variants no significant difference was observed at all (Table S8). We carried out the same analysis in the Estonian Biobank and discovered that diagnoses were accumulating for the variants in a similar manner to that observed in FinnGen (Fig. S4).

Significant genetic correlations were found between 517 traits and LSS (Fig. 3, Table S9). Back pain (rg=0.60, pFDR=5.05^-109^) showed the most significant positive genetic correlation in terms of the lowest p-value, and the diagnosis of M54 Dorsalgia (rg=0.75, pFDR=7.40^-29^) showed the largest significant genetic correlation. There was also a positive genetic correlation between LSS and many pain-related endpoints, including leg pain on walking (rg=0.57, pFDR=1.89e-35). Positive genetic correlations were also observed for general health- and mood-related traits, such as overall health rating (rg=0.40, pFDR=2.25^-65^) and probable major depressive disorder (rg=0.44, pFDR=8.26^-16^).

**Fig. 3.**
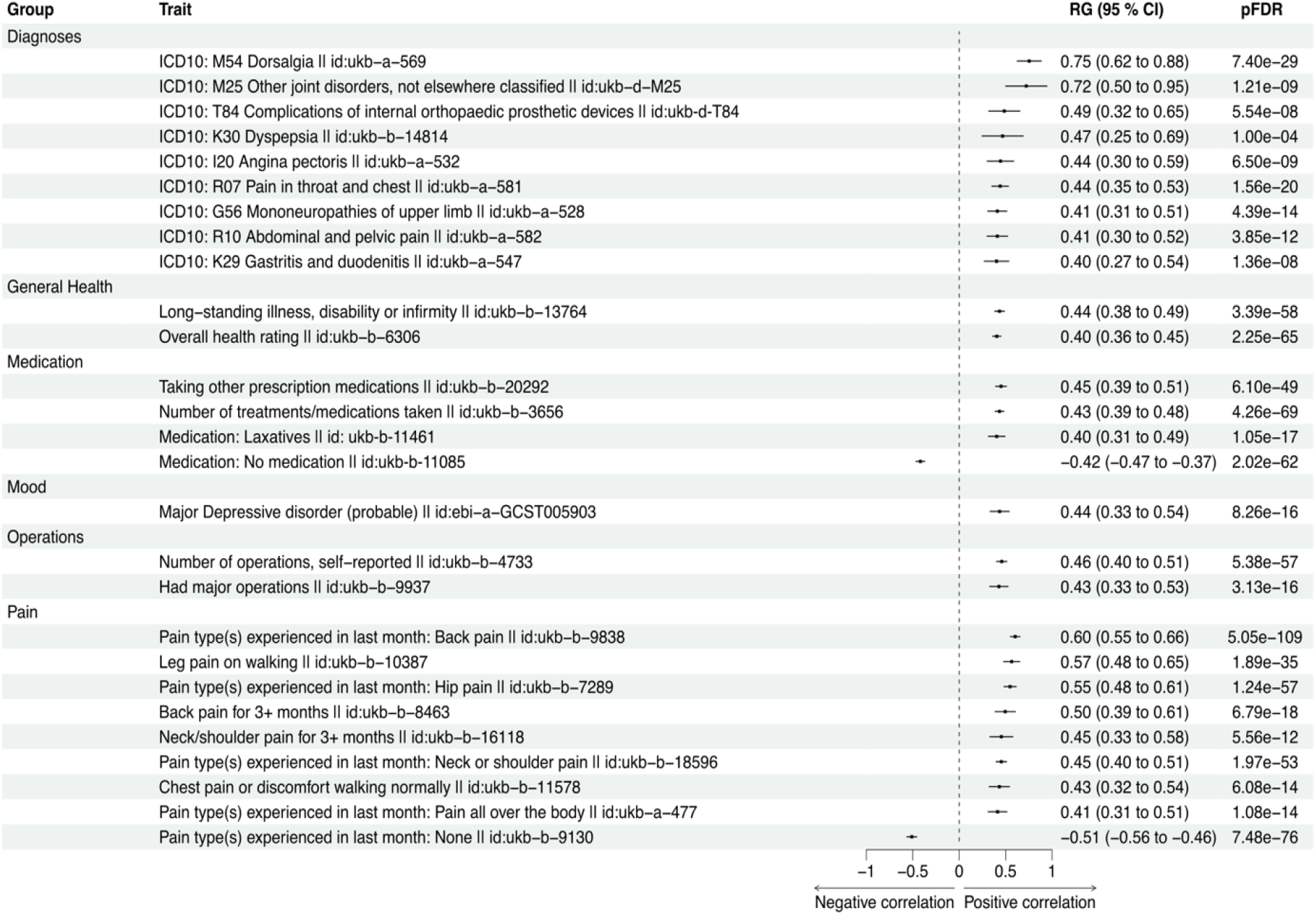
Genetic correlations were calculated using LDSC-software^8,31^. All traits were extracted from the GWAS database provided by the MRC Integrative Epidemiology Unit (IEU). Only the strongest observed (rg< -0.4 & rg> 0.4) correlations with a significant false discovery corrected p-value (p_FDR_ < 0.05) are shown in the figure. RG, genetic correlation coefficient value; pFDR, false discovery rate-corrected p-value. Genetic correlations for all 517 phenotypes can be seen in Table S9.

In Mendelian randomization, we uncovered potential causal relationships between several factors and LSS (Fig. 4A, Fig. S5.1-4, Table S10, Table S11). The most significant potential causal relationships were observed between whole body fat-free mass and higher LSS risk (OR=1.78, pFDR=2.22e-21, Fig. S5.1) and between body mass index (BMI) and higher LSS risk (OR=1.55, pFDR=2.22e-21, Fig. S5.2). The only outcome for which we found LSS to be potentially causal was back pain (Beta=0.02, pFDR=0.0001, Fig. 4B, Fig. S6, Table S10). Pain is the main reason for seeking care and often also the main symptom^2^. Although pleiotropy was not observed, some causal estimates were heterogeneous (Table S10), so these results should be interpreted cautiously. It appears that individual variants do not drive the observed causal relationships, as all causal estimates in the leave-out analyses consistently point in the same direction (Fig. S7.1-4, Fig. S8).

**Fig. 4.**
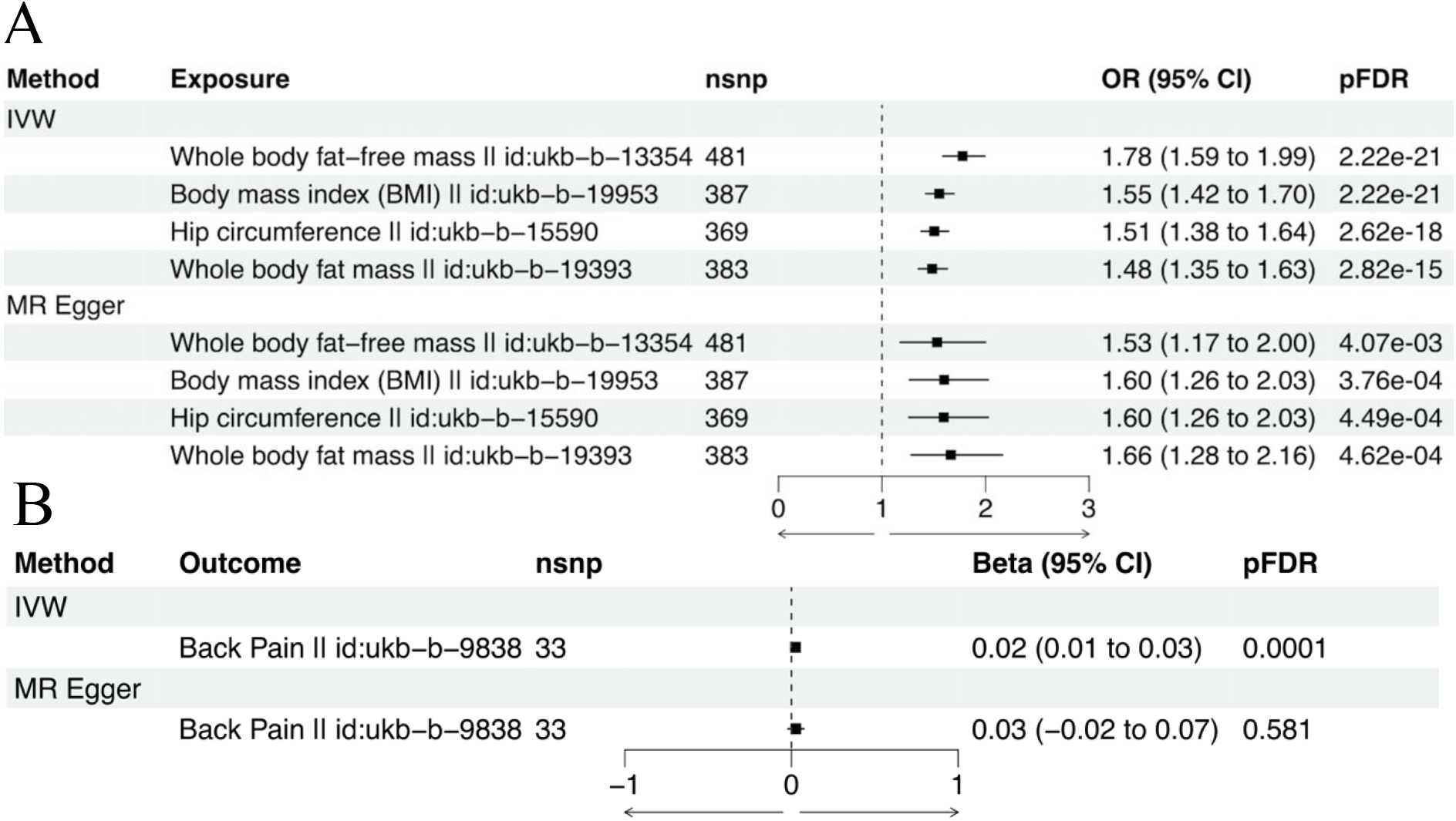
**A)** Exposures potentially causal for LSS, and **B)** outcomes that LSS was potentially found to be causal for. The analysis was performed using the TwoSampleMR R library and data from the present study and MRC-IEU database. The inverse variance weighted (IVW) model was our primary analysis, for which statistical significance was considered at false discovery rate corrected p-value (pFDR < 0.05). We also performed a sensitivity analysis using MR Egger. nsnp, number of SNPs; OR (95% CI), odds ratio and its 95% confidence interval; Beta (95% CI), beta estimate and its 95% confidence interval; pFDR, false discovery rate-corrected p-value.

Our study has strengths and limitations. Multiple genome-wide significant associations with LSS were discovered, facilitated by integrating data from three large biobanks, which resulted in a substantial sample size. Divergences in the proportional frequency of LSS cases among the sample groups incorporated in the meta-analysis indicate possible inconsistencies in the ability of biobanks to recognize patients with LSS. It is also important to note that only people with European ancestry were included in our study. Both the frequency of LSS surgical treatment and the type of surgery performed can differ greatly between countries^2^. In our research, we have used surgical codes that are used in Finland for the surgical treatment of LSS patients^32^. The eQTL analyses would benefit from incorporating bone and cartilage data, which are crucial tissues in LSS pathogenesis. Since our work only uses computational methods, future functional studies would enhance our results.

In conclusion, by identifying a significant number of novel loci and performing multiple downstream analyses, we were able to expand the current understanding of the genetic basis of LSS. Our findings highlighted the significance of spinal degeneration as a major risk factor for lumbar spinal stenosis. Since we found that the number of diagnoses for LSS increased only after the age of 50, our study’s age-of-onset analyses lend credence to the notion that LSS is an age-related condition. However, our analysis of cumulative LSS incidence patterns revealed variants that significantly impact the risk of LSS morbidity as early as 40 years of age.

Additionally, numerous loci associated with LSS requiring surgery underlined pathways that can lead to a more severe LSS. Taken together, our work contributes to our understanding of the genetic origins of LSS and may open the door to future research into novel therapies and preventative strategies for LSS.

## Supporting information

Regional association plots

## Data Availability

The individual-level data are available under restricted access for legal and ethical reasons. Formal approval for the researchers is needed to access the data: please see https://www.finngen.fi/en/access_results for more details. Individual-level data access and genotype data is managed by the Finnish Biobank Cooperative at the Fingenious portal [https://site.fingenious.fi/en/]). We will make the summary statistics of this study available through the NHGRI-EBI GWAS Catalog upon publication.

## Methods

### Study populations FinnGen

FinnGen strives to advance our understanding of disease mechanisms and the efficacy of medical care. The studies aim is to identify links between individual genetic differences and diseases. The FinnGen project has the necessary ethical and prior permits for biobank research (Supplementary Note), and all persons who have provided a research sample are aware of the intended use of the samples and have given their written consent to biobank research either in connection with sample donation or when participating in older research projects, the materials of which have been transferred to Finnish biobanks with the written consent of Fimea. We used *The International Classification of Diseases Tenth Revision* (ICD-10) code M48.0 and *Ninth Revision* (ICD-9) codes 7230A, 7240A, and 7240B (Table S1) to characterize cases in FinnGen. Patients without a record of these ICD codes were classified as controls, and participants who were diagnosed with other dorsopathies were excluded (M40-M54, M76). Patient data were obtained from the Hospital Discharge Registry and the Cause of Death Registry however, patients whose registration data was only available in the primary care registry were excluded from the analysis. The FinnGen (R9-version) data used in the study included 18 989 LSS cases and 270 964 controls.

### Estonian Biobank (EstBB)

The Estonian Biobank cohort is a volunteer-based sample of the Estonian resident adult population (aged ≥18 years)^33^. Estonians represent 83%, Russians 14%, and other nationalities 3% of all participants. The current number of participants is > 205,000 and represents a large proportion, > 15 % of the Estonian adult population, making it ideally suited to population-based studies. General practitioners (GPs) and medical personnel in the special recruitment offices have recruited participants throughout the country. At baseline, the GPs performed a standardized health examination of the participants, who also donated blood samples for DNA, white blood cells and plasma tests and filled out a 16-module questionnaire on health-related topics such as lifestyle, diet and clinical diagnoses described in WHO ICD-10. A significant part of the cohort has whole genome sequencing (3000), whole exome sequencing (2500), genome-wide single nucleotide polymorphism (SNP) array data (200 000) and/or NMR metabolome data (200 000) available. In the meta-analysis, 72 679 participants from the Estonian Biobank were included; 6019 of these were LSS cases, and 65 258 were controls.

### UK Biobank

The UK biobank^34^ cohort comprises of samples acquired between 2006-2010. Hundreds of thousands of people aged 40-69 from across Britain provided samples. In our research, we used a subset of European ancestry, consisting of 5134 LSS patients and 401 917 controls from the UK biobank, more precisely summary statistics from the PanUKBB project.

### Genotyping, imputation & quality control FinnGen

Genotyping was performed using Illumina and Affymetrix DNA microarrays. Genotype data were quality controlled to exclude variants with a low Hardy-Weinberg equilibrium (HWE) p-value (<1×10^−6^), minor allele count (MAC) below three, and high missingness (cut-off 2%), as well as individuals with high genotype missingness (cut-off 5%), high levels of heterozygosity (±4 SD), non-Finnish ancestry, and individuals whose sex did not match the genotype data. Eagle 2.3.5 was used in prephasing, were 20000 was used as a threshold of conditioning haplotypes. Genotype imputation was done with Beagle 4.1 (version 08Jun17.d8b), and Finnish SISu v3 was used as a reference panel. Whole imputation protocol has been described at doi.org/10.17504/protocols.io.nmndc5e. For post-imputation quality control, we excluded variants with imputation information less than 0.6.

### EstBB

Illumina Human CoreExome, OmniExpress, 370CNV BeadChip and GSA arrays were used for genotyping. Individuals were excluded from the analysis if their call-rate was < 95% or sex defined using X chromosome heterozygosity estimates didn’t match phenotypic data. Quality control included filtering on the basis of sample call rate (< 98%), heterozygosity (> mean ± 3SD), genotype and phenotype sex discordance, cryptic relatedness (IBD > 20%) and outliers from the European descent based on the MDS plot in comparison with HapMap reference samples. SNP quality filtering included call rate (<99%), MAF (<1%) and extreme deviation from Hardy–Weinberg equilibrium (P < 1 × 10−4). SHAPEIT2 was used for prephasing, while the imputation was performed with the Estonian-specific reference panel^35^ and IMPUTE2^36^ with default settings. Association testing was carried out with snptest-2.5.2, adjusting for 4 PCs, arrays, current age, and sex (when relevant). Variants with call-rate < 95%, MAF < 1% or HWE p-value < 1e-4 (autosomal variants only) and indels were excluded.

### GWAS

The Regenie (version 2.2.4) program^37^ was used to conduct GWAS using an additive genetic model in FinnGen and EstBB, modifying each phenotype for age, sex, the first 10 genetic principal components and genotyping arrays. The sensitivity analysis was performed using Regenie and the same covariates as above.

### FinnGen-based sensitivity analysis on surgical LSS

Additionally, we conducted sensitivity analysis in FinnGen, with the aim of identifying genomic regions associated with more severe LSS that required surgical treatment. The sensitivity analysis was done using patients with LSS who had undergone surgery (*NOMESCO version 1*.*15*, ABC36, ABC56, ABC66, ABC99, NAG61, NAG62, NAG63, NAG66, NAG67, and NAG99)^32^, with the control group defined similarly to the meta-analysis. A total of 8627 cases and 270 773 controls were included in this analysis. LSS cases that lacked documentation of studied surgical codes were not included in the analysis. The same policy was followed in cases of patients who underwent surgery but did not have a LSS diagnosis because it is likely that these patients had an acute injury.

### Meta-analysis (FinnGen, EstBB, UKBB)

The inverse-variance weighted meta-analysis was carried out using Python software (https://github.com/FINNGEN/META_ANALYSIS/). Variant data from the Estonian and UK Biobanks were converted from hg19 to hg38 before being meta-analyzed using the Picard liftover (http://broadinstitute.io/picard). Meta-analysis data consists only SNPs that have been observed in at least two of the datasets. In total, there were 769 683 participants in the meta-analysis, of which there were 30 269 cases and 739 414 controls.

### FinnGen+EstBB meta-analysis

The METAL software^38^ was used to do an inverse variance-weighted fixed-effect meta-analysis of the GWAS results from FinnGen and EstBB. Prior to the meta-analysis, variant data from the Estonian Biobank were converted from the hg19 to the hg38 prior to the meta-analysis. Summary statistics from the FinnGen+EstBB meta-analysis results were used in the Mendelian randomization, in order to increase statistical power for detecting potential causal relationships.

### Candidate gene characterization

A locus was defined as a window of 2MB (± 1,000,000 bases) that contains at least one variant associated with LSS at P<5×10^−8^. Using literature and databases (Genbank^39^, UniProt^40^, GTEx-Portal^41^), and identified a potential candidate gene with a relevant biological function for the loci that had not been reported in association with LSS in previous studies.

### Conditional analyses

With help of GCTA software (1.93.0 beta Linux)^42^, we performed conditional analyzes for the loci to identify possible secondary signals. We used lead variants observed from the loci as a covariate and utilized FinnGen as the reference sample to estimate linkage disequilibrium (LD) corrections. The associations were first conditioned on the most significant variant at each locus using the --cojo-cond option with the default settings. For those loci where secondary signals were detected, the analysis was repeated using the first conditional analysis results. Conditioning was continued until no variant reached genome-wide significance (p < 5×10-8).

### Downstream analyses Heritability

The LD score regression-derived SNP-based heritability estimate was computed using LDSC software (version 1.0.1)^8^. Heritability estimation was performed using the liability scale, with a sample prevalence of 0.039 and a population prevalence of 0.11 as estimated by Jensen et al. (2020)^4^. For heritability estimation we used the HapMap3, a European-only reference panel provided by Alkes Price’s group (https://alkesgroup.broadinstitute.org/LDSCORE/). Summary statistics included 1202708 SNPs, and after merging with the LD reference panel 1177076 SNPs remained. FinnGen data showed a sample prevalence of 0.065. The reference panel remained unchanged; 1191433 SNPs were included in the FinnGen summary statistics, and after merging with the LD reference panel, 1168512 SNPs remained.

### Functional annotations

FUMA^27^ was used for the functional annotations that were performed based on the meta-analysis summary statistics. We selected that the analysis would use functional information for the mapping. Positional mapping was also carried out, with SNP markers chosen from regions of exons or introns that affect post-transcriptional modifications and are involved in gene regulation. Gene expression data were also used for mapping, as well as eQTL mapping. We used whole blood, cultured fibroblasts, tibial nerve, and cervical spinal cord cervical (GTEx v8) tissues in the analysis, and the focused on the protein-coding genes only. MAGMA analysis^26^, a functional association test, was also performed, it focuses on gene-level information, unlike GWAS, where associations are reported at the variant level. MAGMA uses curated gene sets and GO annotations from MSigDB^28^ in the analyses. We used 10kb gene window and selected GTEx v8 tissue variants in the analysis. We also used fastBAT from the GCTA software package (1.93.0 beta Linux)^42^ for gene prioritization. re calculates the association p-value for a set of SNPs from an approximated distribution of the sum of χ2-statistics over the SNPs at gene regions using LD correlations between SNPs from a reference sample with individual-level genotypes and summary data from GWAS^43^. We utilized FinnGen as a LD reference and used 2MB window for the loci. For LD pruning 0.9 r2 threshold was used alongside default methods. We considered results with pfastBAT<5×10^−8^ and pGWAS<5×10^−8^ significant^43^. Lead variants that located in the HLA region were left out from the annotation analyses.

### eQTL Colocalizations

We investigated eQTL colocalizations between LSS association signals and gene expression using the ‘coloc.abf’ function from the R-library ‘coloc’ (version 5.2.3)^44^. The ‘biomaRt’ (version 2.58.2) R-library was used to extract genes from Emsembl archives^29^ within a 1 MB (± 500000) window surrounding each lead variant. Colocalizations were investigated for 754 genes that were available in GTEx. We chose tissues relevant to LSS for analysis, including the tibial nerve, cervical spinal cord, and whole blood. For the selected tissues, data on variant-gene expression associations (GTEx v8) were downloaded from GTExportal^41^. Colocalizations with a posterior probability of ≥ 0.8 for the variant were deemed significant^30^.

### Survival analysis

Our aim was to observe how the LSS diagnoses accumulate for different variants according to age and to evaluate whether there are early onset variants, by exploring the cumulative events We determined the diagnosis age in years when the curve of the effect allele (EA) homozygotes statistically differed from the curve of other allele (OA) homozygotes of the same variant, of the variants where significant differences in the accumulation of diagnoses were observed. The calculation was performed with the two-tailed test by using the survival rates and survival rate standard errors obtained with the ‘survfit’ function, which is part of the ‘survival’ (https://github.com/therneau/survival, version 3.2-7) R library. The equation used was: *diff*=(*‘survival rate EA/EA’-’survival rate OA/OA’)/sqrt(‘se_survival_rate_EA/EA2’+’se_survival_rate_OA/OA2’*), p=2*(1-*diff*). A significant difference was defined as a P-value < 0.05. Furthermore, we calculated cumulative morbidity for each variant to determine whether some variants result in more diagnoses.

### Genetic correlations

Genetic correlations were calculated between meta-analysed LSS summary statistics and 517 other phenotypes extracted from the GWAS database provided by the MRC Integrative Epidemiology Unit (IEU) (https://gwas.mrcieu.ac.uk/). These calculations were performed using the LDSC software (version 1.0.1)^8,31^. We set the threshold for significant correlations at a false discovery rate (FDR)-corrected p-value (pFDR) < 0.05. Genetic correlations were performed using HapMap3 European-only reference panel provided by Alkes Price’s group (https://alkesgroup.broadinstitute.org/LDSCORE/). For genetic correlations, we used our LSS summary statistics and the GWAS summary statistics that were similar to those that have been previously available in LDhub (https://ldsc.broadinstitute.org).

### Mendelian randomization

In order to investigate the causal relationships between LSS and its associated risk factors, we used the Two-Sample MR (version 0.5.8) R library to perform a bi-directional Mendelian randomization. Risk factors related to body composition, pain, and inflammation were included in the analysis (Table S11). We were able to assess if LSS is causal for risk factors and, conversely, if risk factors are causal for LSS due to a bi-directional study approach. To ensure that there was no overlap between the study populations. We obtained the LSS instruments from the FinnGen+EstBB meta-analysis results since many of the GWAS data provided by the MRC-IEU are UKBB-based. Variants that were correlated were removed from the data, leaving only independent variants for analysis. We used the default clumping settings in the analysis (clumping window 10 000kb, r^2^ 0.001). The principal analysis method we employed was the Inverse Variance Weighted (IVW) model. In the sensitivity analyses, we obtained MR Egger estimates and performed Cochran’s Q-test and the MR Egger intercept test to evaluate the heterogeneity and pleiotropy of the instruments. In order to determine whether a particular SNP is responsible for a potentially detectable causal relationship, a leave-one-out analysis was also carried out.

## Code availability

The citations and URLs listed in the Methods section contain the code for every tool used in the analyses. All tools used in our study were open source.

## Acknowledgements

E.S. was funded by Academy of Finland (grant number: 338229) and Orion Research Foundation sr. J.K. was funded by Sigrid Juselius foundation. The authors wish to acknowledge CSC – IT Center for Science, Finland, for computational resources. We want to acknowledge the participants and investigators of FinnGen study. The FinnGen project is funded by two grants from Business Finland (HUS 4685/31/2016 and UH 4386/31/2016) and the following industry partners: AbbVie Inc., AstraZeneca UK Ltd, Biogen MA Inc., Bristol Myers Squibb (and Celgene Corporation & Celgene International II Sàrl), Genentech Inc., Merck Sharp & Dohme LCC, Pfizer Inc., GlaxoSmithKline Intellectual Property Development Ltd., Sanofi US Services Inc., Maze Therapeutics Inc., Janssen Biotech Inc, Novartis Pharma AG, and Boehringer Ingelheim International GmbH. Following biobanks are acknowledged for delivering biobank samples to FinnGen: Auria Biobank (www.auria.fi/biopankki), THL Biobank (www.thl.fi/biobank), Helsinki Biobank (www.helsinginbiopankki.fi), Biobank Borealis of Northern Finland (https://www.ppshp.fi/Tutkimus-ja-opetus/Biopankki/Pages/Biobank-Borealis-briefly-in-English.aspx), Finnish Clinical Biobank Tampere (www.tays.fi/en-US/Research_and_development/Finnish_Clinical_Biobank_Tampere), Biobank of Eastern Finland (www.ita-suomenbiopankki.fi/en), Central Finland Biobank (www.ksshp.fi/fi-FI/Potilaalle/Biopankki), Finnish Red Cross Blood Service Biobank (www.veripalvelu.fi/verenluovutus/biopankkitoiminta), Terveystalo Biobank (www.terveystalo.com/fi/Yritystietoa/Terveystalo-Biopankki/Biopankki/) and Arctic Biobank (https://www.oulu.fi/en/university/faculties-and-units/faculty-medicine/northern-finland-birth-cohorts-and-arctic-biobank). All Finnish Biobanks are members of BBMRI.fi infrastructure (www.bbmri.fi). Finnish Biobank Cooperative -FINBB (https://finbb.fi/) is the coordinator of BBMRI-ERIC operations in Finland. The Finnish biobank data can be accessed through the Fingenious^®^ services (https://site.fingenious.fi/en/) managed by FINBB.

This study was funded by European Union through the European Regional Development Fund Project No. 2014-2020.4.01.15-0012 GENTRANSMED, the Estonian Research Council Grant PUTs (PRG1911, PRG1291) and by Estonian Ministry of Education and Research Funding (TK214). Data analysis was carried out in part in the High-Performance Computing Center of University of Tartu. The activities of the EstBB are regulated by the Human Genes Research Act, which was adopted in 2000 specifically for the operations of the EstBB. Individual level data analysis in the EstBB was carried out under ethical approval [1.1-12/624] from the Estonian Committee on Bioethics and Human Research (Estonian Ministry of Social Affairs), using data according to release application [N04] from the Estonian Biobank.

## Consortia

### FinnGen

Aarno Palotie^11,12,13^, Mark Daly^11,12,13^, Bridget Riley-Gills^14^, Howard Jacob^14^, Dirk Paul^15^, Slavé Petrovski^15^, Heiko Runz^16^, Sally John^16^, George Okafo^17^, Nathan Lawless^17^, Heli Salminen-Mankonen^17^, Robert Plenge^18^, Joseph Maranville^18^, Mark McCarthy^19^, Margaret G. Ehm^20^, Kirsi Auro^21^, Simonne Longerich^22^, Anders Mälarstig^23^, Katherine Klinger^24^, Clement Chatelain^24^, Matthias Gossel^24^, Karol Estrada^25^, Robert Graham^25^, Robert Yang^26^, Chris O’Donnell^27^, Tomi P. Mäkelä^11^, Jaakko Kaprio^11^, Petri Virolainen^28^, Antti Hakanen^28^, Terhi Kilpi^29^, Markus Perola^29^, Jukka Partanen^30^, Anne Pitkäranta^31^, Taneli Raivio^31^, Jani Tikkanen^32^, Raisa Serpi^32^, Tarja Laitinen^33^, Veli-Matti Kosma^34^, Jari Laukkanen^35,36^, Marco Hautalahti^37^, Outi Tuovila^38^, Raimo Pakkanen^38^, Jeffrey Waring^14^, Fedik Rahimov^14^, Ioanna Tachmazidou^15^, Chia-Yen Chen^16^, Zhihao Ding^17^, Marc Jung^17^, Shameek Biswas^18^, Rion Pendergrass^19^, David Pulford^39^, Neha Raghavan^22^, Adriana Huertas-Vazquez^22^, Jae-Hoon Sul^22^, Xinli Hu^23^, Åsa Hedman^23^, Manuel Rivas^25,40^, Dawn Waterworth^41^, Nicole Renaud^27^, Ma’en Obeidat^27^, Samuli Ripatti^11^, Johanna Schleutker^28^, Mikko Arvas^30^, Olli Carpén^31^, Reetta Hinttala^32^, Arto Mannermaa^34^, Katriina Aalto-Setälä^42^, Mika Kähönen^33^, Johanna Mäkelä^37^, Reetta Kälviäinen^43^, Valtteri Julkunen^43^, Hilkka Soininen^43^, Anne Remes^44^, Mikko Hiltunen^45^, Jukka Peltola^46^, Minna Raivio^47^, Pentti Tienari^47^, Juha Rinne^48^, Roosa Kallionpää^48^, Juulia Partanen^11^, Ali Abbasi^14^, Adam Ziemann^14^, Nizar Smaoui^14^, Anne Lehtonen^14^, Susan Eaton^16^, Sanni Lahdenperä^16^, Natalie Bowers^19^, Edmond Teng^19^, Fanli Xu^49^, Laura Addis^49^, John Eicher^49^, Qingqin S Li^50^, Karen He^41^, Ekaterina Khramtsova^41^, Martti Färkkilä^47^, Jukka Koskela^47^, Sampsa Pikkarainen^47^, Airi Jussila^46^, Katri Kaukinen^46^, Timo Blomster^44^, Mikko Kiviniemi^43^, Markku Voutilainen^48^, Tim Lu^19^, Linda McCarthy^49^, Amy Hart^41^, Meijian Guan^41^, Jason Miller^22^, Kirsi Kalpala^23^, Melissa Miller^23^, Kari Eklund^47^, Antti Palomäki^48^, Pia Isomäki^46^, Laura Pirilä^48^, Oili Kaipiainen-Seppänen^43^, Johanna Huhtakangas^44^, Nina Mars^11^, Apinya Lertratanakul^14^, Coralie Viollet^15^, Marla Hochfeld^18^, Jorge Esparza Gordillo^49^, Fabiana Farias^22^, Nan Bing^23^, Margit Pelkonen^43^, Paula Kauppi^47^, Hannu Kankaanranta^42,51,52^, Terttu Harju^44^, Riitta Lahesmaa^48^, Hubert Chen^19^, Joanna Betts^49^, Rajashree Mishra^49^, Majd Mouded^53^, Debby Ngo^53^, Teemu Niiranen^54^, Felix Vaura^54^, Veikko Salomaa^54^, Kaj Metsärinne^48^, Jenni Aittokallio^48^, Jussi Hernesniemi^46^, Daniel Gordin^47^, Juha Sinisalo^47^, Marja-Riitta Taskinen^47^, Tiinamaija Tuomi^47^, Timo Hiltunen^47^, Amanda Elliott^11,13,55^, Mary Pat Reeve^11^, Sanni Ruotsalainen^11^, Audrey Chu^49^, Dermot Reilly^56^, Mike Mendelson^57^, Jaakko Parkkinen^23^, Tuomo Meretoja^47^, Heikki Joensuu^47^, Johanna Mattson^47^, Eveliina Salminen^47^, Annika Auranen^46^, Peeter Karihtala^44^, Päivi Auvinen^43^, Klaus Elenius^48^, Esa Pitkänen^11^, Relja Popovic^14^, Margarete Fabre^15^, Jennifer Schutzman^19^, Diptee Kulkarni^49^, Alessandro Porello^41^, Andrey Loboda^22^, Heli Lehtonen^23^, Stefan McDonough^23^, Sauli Vuoti^58^, Kai Kaarniranta^43,59^, Joni A. Turunen^60,61^, Terhi Ollila^47^, Hannu Uusitalo^46^, Juha Karjalainen^11^, Mengzhen Liu^14^,

Stephanie Loomis^16^, Erich Strauss^19^, Hao Chen^19^, Kaisa Tasanen^44^, Laura Huilaja^44^, Katariina Hannula-Jouppi^47^, Teea Salmi^46^, Sirkku Peltonen^48^, Leena Koulu^48^, David Choy^19^, Ying Wu^23^, Pirkko Pussinen^47^, Aino Salminen^47^, Tuula Salo^47^, David Rice^47^, Pekka Nieminen^47^, Ulla Palotie^47^, Maria Siponen^43^, Liisa Suominen^43^, Päivi Mäntylä^43^, Ulvi Gursoy^48^, Vuokko Anttonen^44^, Kirsi Sipilä^62,63^, Hannele Laivuori^11^, Venla Kurra^46^, Laura Kotaniemi-Talonen^46^, Oskari Heikinheimo^47^, Ilkka Kalliala^47^, Lauri Aaltonen^47^, Varpu Jokimaa^48^, Marja Vääräsmäki^44^, Outi Uimari^44^, Laure Morin-Papunen^44^, Maarit Niinimäki^44^, Terhi Piltonen^44^, Katja Kivinen^11^, Elisabeth Widen^11^, Taru Tukiainen^11^, Niko Välimäki^64^, Eija Laakkonen^65^, Jaakko Tyrmi^42,62^, Heidi Silven^62^, Riikka Arffman^62^, Susanna Savukoski^62^, Triin Laisk^6^, Natalia Pujol^6^, Janet Kumar^20^, Iiris Hovatta^64^, Erkki Isometsä^47^, Hanna Ollila^11^, Jaana Suvisaari^54^, Thomas Damm Als^66^, Antti Mäkitie^67^, Argyro Bizaki-Vallaskangas^46^, Sanna Toppila-Salmi^68^, Tytti Willberg^48^, Elmo Saarentaus^11^, Antti Aarnisalo^47^, Elisa Rahikkala^44^, Kristiina Aittomäki^69^, Fredrik Åberg^70^, Mitja Kurki^11,55^, Aki Havulinna^11,54^, Juha Mehtonen^11^, Shabbeer Hassan^11^, Pietro Della Briotta Parolo^11^, Wei Zhou^55^, Mutaamba Maasha^55^, Susanna Lemmelä^11^, Aoxing Liu^11^, Arto Lehisto^11^, Andrea Ganna^11^, Vincent Llorens^11^, Henrike Heyne^11^, Joel Rämö^11^, Rodos Rodosthenous^11^, Satu Strausz^11^, Tuula Palotie^47,64^, Kimmo Palin^64^, Javier Gracia-Tabuenca^42^, Harri Siirtola^42^, Tuomo Kiiskinen^11^, Jiwoo Lee^11,55^, Kristin Tsuo^11,55^, Kati Kristiansson^29^, Kati Hyvärinen^71^, Jarmo Ritari^71^, Katri Pylkäs^62^, Minna Karjalainen^62^, Tuomo Mantere^32^, Eeva Kangasniemi^33^, Sami Heikkinen^45^, Nina Pitkänen^28^, Samuel Lessard^24^, Lila Kallio^28^, Tiina Wahlfors^29^, Eero Punkka^31^, Sanna Siltanen^33^, Teijo Kuopio^35,36^, Anu Jalanko^11^, Huei-Yi Shen^11^, Risto Kajanne^11^, Mervi Aavikko^11^, Helen Cooper^11^, Denise Öller^11^, Rasko Leinonen^11,72^, Henna Palin^33^, Malla-Maria Linna^31^, Masahiro Kanai^55^, Zhili Zheng^55^, L. Elisa Lahtela^11^, Mari Kaunisto^11^, Elina Kilpeläinen^11^, Timo P. Sipilä^11^, Oluwaseun Alexander Dada^11^, Awaisa Ghazal^11^, Anastasia Kytölä^11^, Rigbe Weldatsadik^11^, Kati Donner^11^, Anu Loukola^31^, Päivi Laiho^29^, Tuuli Sistonen^29^, Essi Kaiharju^29^, Markku Laukkanen^29^, Elina Järvensivu^29^, Sini Lähteenmäki^29^, Lotta Männikkö^29^, Regis Wong^29^, Auli Toivola^29^, Minna Brunfeldt^29^, Hannele Mattsson^29^, Sami Koskelainen^29^, Tero Hiekkalinna^29^, Teemu Paajanen^29^, Kalle Pärn^11^, Mart Kals^11^, Shuang Luo^11^, Shanmukha Sampath Padmanabhuni^11^, Marianna Niemi^42^, Mika Helminen^42^, Tiina Luukkaala^42^, Iida Vähätalo^42^, Jyrki Tammerluoto^11^, Sarah Smith^37^, Tom Southerington^37^, Petri Lehto^37^

^11^ Institute for Molecular Medicine Finland (FIMM), HiLIFE, University of Helsinki, Helsinki, Finland, ^12^ Broad Institute of MIT and Harvard, Cambridge, MA, United States, ^13^ Massachusetts General Hospital, Boston, MA, United States, ^14^ Abbvie, Chicago, IL, United States, ^15^ Astra Zeneca, Cambridge, United Kingdom, ^16^ Biogen, Cambridge, MA, United States, ^17^ Boehringer Ingelheim, Ingelheim am Rhein, Germany, ^18^ Bristol Myers Squibb, New York, NY, United States, ^19^ Genentech, San Francisco, CA, United States, ^20^ GlaxoSmithKline, Collegeville, PA, United States, ^21^ GlaxoSmithKline, Espoo, Finland, ^22^ Merck, Kenilworth, NJ, United States, ^23^ Pfizer, New York, NY, United States, ^24^ Translational Sciences, Sanofi R&D, Framingham, MA, USA, ^25^ Maze Therapeutics, San Francisco, CA, United States, ^26^ Janssen Biotech, Beerse, Belgium, ^27^ Novartis Institutes for BioMedical Research, Cambridge, MA, United States, ^28^ Auria Biobank, University of Turku, Hospital District of Southwest Finland, Turku, Finland, ^29^ THL Biobank, Finnish Institute for Health and Welfare (THL), Helsinki, Finland, ^30^ Finnish Red Cross Blood Service, Finnish Hematology Registry and Clinical Biobank, Helsinki, Finland, ^31^ Helsinki Biobank, Helsinki University and Hospital District of Helsinki and Uusimaa, Helsinki, ^32^ Northern Finland Biobank Borealis, University of Oulu, Northern Ostrobothnia Hospital District, Oulu, Finland, ^33^ Finnish Clinical Biobank Tampere, University of Tampere, Pirkanmaa Hospital District, Tampere, Finland, ^34^ Biobank of Eastern Finland, University of Eastern Finland, Northern Savo Hospital District, Kuopio, Finland, ^35^ Central Finland Biobank, University of Jyväskylä, Central Finland Health Care District, Jyväskylä, Finland, ^36^ Central Finland Health Care District, Jyväskylä, Finland,^37^ FINBB - Finnish biobank cooperative, ^38^ Business Finland, Helsinki, Finland, ^39^ GlaxoSmithKline, Stevenage, United Kingdom, ^40^ University of Stanford, Stanford, CA, United States, ^41^ Janssen Research & Development, LLC, Spring House, PA, United States, ^42^ University of Tampere, Tampere, Finland, ^43^ Northern Savo Hospital District, Kuopio, Finland, ^44^ Northern Ostrobothnia Hospital District, Oulu, Finland, ^45^ University of Eastern Finland, Kuopio, Finland, ^46^ Pirkanmaa Hospital District, Tampere, Finland, ^47^ Hospital District of Helsinki and Uusimaa, Helsinki, Finland, ^48^ Hospital District of Southwest Finland, Turku, Finland, ^49^ GlaxoSmithKline, Brentford, United Kingdom, ^50^ Janssen Research & Development, LLC, Titusville, NJ 08560, United States, ^51^ University of Gothenburg, Gothenburg, Sweden, ^52^ Seinäjoki Central Hospital, Seinäjoki, Finland, ^53^ Novartis, Basel, Switzerland, ^54^ Finnish Institute for Health and Welfare (THL), Helsinki, Finland, ^55^ Broad Institute, Cambridge, MA, United States, ^56^ Janssen Research & Development, LLC, Boston, MA, United States, ^57^ Novartis, Boston, MA, United States, ^58^ Janssen-Cilag Oy, Espoo, Finland, ^59^ Department of Molecular Genetics, University of Lodz, Lodz, Poland, ^60^ Helsinki University Hospital and University of Helsinki, Helsinki, Finland, ^61^ Eye Genetics Group, Folkhälsan Research Center, Helsinki, Finland, ^62^ University of Oulu, Oulu, Finland, ^63^ Medical Research Center, Oulu, Oulu University Hospital and University of Oulu, Oulu, Finland, ^64^ University of Helsinki, Helsinki, Finland, ^65^ University of Jyväskylä, Jyväskylä, Finland, ^66^ Aarhus University, Denmark, ^67^ Department of Otorhinolaryngology - Head and Neck Surgery, University of Helsinki and Helsinki University Hospital, Helsinki, Finland, ^68^ University of Eastern Finland and Kuopio University Hospital, Department of Otorhinolaryngology, Kuopio, Finland and Department of Allergy, Helsinki University Hospital and University of Helsinki, Finland, ^69^ Department of Medical Genetics, Helsinki University Central Hospital, Helsinki, Finland, ^70^ Transplantation and Liver Surgery Clinic, Helsinki University Hospital, Helsinki University, Helsinki, Finland, ^71^ Finnish Red Cross Blood Service, Helsinki, Finland, ^72^ European Molecular Biology Laboratory, European Bioinformatics Institute, Cambridge, UK

### Estonian Biobank Research Team

Andres Metspalu^6^, Mari Nelis^6^, Lili Milani^6^, Georgi Hudjashov^6^

## References

1. Katz, J. N., Zimmerman, Z. E., Mass, H. & Makhni, M. C. Diagnosis and Management of Lumbar Spinal Stenosis. JAMA 327, 1688 (2022).

2. Lurie, J. & Tomkins-Lane, C. Management of lumbar spinal stenosis. BMJ h6234 (2016) doi:10.1136/bmj.h6234.

3. Lai, M. K. L., Cheung, P. W. H. & Cheung, J. P. Y. A systematic review of developmental lumbar spinal stenosis. European Spine Journal 29, 2173–2187 (2020).

4. Jensen, R. K., Jensen, T. S., Koes, B. & Hartvigsen, J. Prevalence of lumbar spinal stenosis in general and clinical populations: a systematic review and meta-analysis. European Spine Journal vol. 29 2143–2163 Preprint at 10.1007/s00586-020-06339-1 (2020).

5. Sakaue, S. et al. A cross-population atlas of genetic associations for 220 human phenotypes. Nat Genet 53, 1415–1424 (2021).

6. Suri, P. et al. Genome-wide association studies of low back pain and lumbar spinal disorders using electronic health record data identify a locus associated with lumbar spinal stenosis. Pain 162, 2263–2272 (2021).

7. Bovonratwet, P. et al. Identification of Novel Genetic Markers for the Risk of Spinal Pathologies: A Genome-Wide Association Study of 2 Biobanks. J Bone Joint Surg Am (2023) doi:10.2106/JBJS.22.00872.

8. Bulik-Sullivan, B. K. et al. LD Score regression distinguishes confounding from polygenicity in genome-wide association studies. Nat Genet 47, 291–295 (2015).

9. Salo, V. et al. Genome-wide meta-analysis conducted in three large biobanks expands the genetic landscape of lumbar disc herniations. doi:10.1101/2023.10.15.23296916.

10. Bjornsdottir, G. et al. Rare SLC13A1 variants associate with intervertebral disc disorder highlighting role of sulfate in disc pathology. Nat Commun 13, 634 (2022).

11. Johnston, K. J. A. et al. Genome-wide association study of multisite chronic pain in UK biobank. PLoS Genet 15, (2019).

12. Zhang, Y. et al. Genome-wide Association Analysis Across 16,956 Patients Identifies a Novel Genetic Association Between BMP6, NIPAL1, CNGA1 and Spondylosis. Spine (Phila Pa 1976) 46, E625–E631 (2021).

13. Koike, Y. et al. Genetic insights into ossification of the posterior longitudinal ligament of the spine. Elife 12, (2023).

14. Nakajima, M. et al. A genome-wide association study identifies susceptibility loci for ossification of the posterior longitudinal ligament of the spine. Nat Genet 46, 1012–1016 (2014).

15. Suri, P. et al. Genome-wide meta-analysis of 158,000 individuals of European ancestry identifies three loci associated with chronic back pain. PLoS Genet 14, e1007601 (2018).

16. Freidin, M. B. et al. Insight into the genetic architecture of back pain and its risk factors from a study of 509,000 individuals. Pain 160, 1361–1373 (2019).

17. Katz, J. N., Zimmerman, Z. E., Mass, H. & Makhni, M. C. Diagnosis and Management of Lumbar Spinal Stenosis: A Review. JAMA vol. 327 1688–1699 Preprint at 10.1001/jama.2022.5921 (2022).

18. McDonald, M.-L. N. et al. Novel genetic loci associated with osteoarthritis in multi-ancestry analyses in the Million Veteran Program and UK Biobank. Nat Genet 54, 1816–1826 (2022).

19. Ebisawa, T. et al. Characterization of bone morphogenetic protein-6 signaling pathways in osteoblast differentiation. J Cell Sci 112 (Pt 20), 3519–27 (1999).

20. Matsunaga, S. et al. Pathogenesis of myelopathy in patients with ossification of the posterior longitudinal ligament. J Neurosurg 96, 168–72 (2002).

21. Abiola, R., Rubery, P. & Mesfin, A. Ossification of the Posterior Longitudinal Ligament: Etiology, Diagnosis, and Outcomes of Nonoperative and Operative Management. Global Spine J 6, 195–204 (2016).

22. Itoh, Y. et al. A common SNP risk variant MT1-MMP causative for Dupuytren’s disease has a specific defect in collagenolytic activity. Matrix Biology 97, 20–39 (2021).

23. Kosaka, H. et al. Pathomechanism of Loss of Elasticity and Hypertrophy of Lumbar Ligamentum Flavum in Elderly Patients With Lumbar Spinal Canal Stenosis. Spine (Phila Pa 1976) 32, 2805–2811 (2007).

24. Ng, M. et al. A Genome-wide Association Study of Dupuytren Disease Reveals 17 Additional Variants Implicated in Fibrosis. Am J Hum Genet 101, 417–427 (2017).

25. Green, H. D. et al. A genome-wide association study identifies 5 loci associated with frozen shoulder and implicates diabetes as a causal risk factor. PLoS Genet 17, (2021).

26. de Leeuw, C. A., Mooij, J. M., Heskes, T. & Posthuma, D. MAGMA: Generalized Gene-Set Analysis of GWAS Data. PLoS Comput Biol 11, e1004219 (2015).

27. Watanabe, K., Taskesen, E., van Bochoven, A. & Posthuma, D. Functional mapping and annotation of genetic associations with FUMA. Nat Commun 8, 1826 (2017).

28. Liberzon, A. et al. Molecular signatures database (MSigDB) 3.0. Bioinformatics 27, 1739–40 (2011).

29. Martin, F. J. et al. Ensembl 2023. Nucleic Acids Res 51, D933–D941 (2023).

30. Sliz, E. et al. Evidence of a causal effect of genetic tendency to gain muscle mass on uterine leiomyomata. Nat Commun 14, 542 (2023).

31. Bulik-Sullivan, B. et al. An atlas of genetic correlations across human diseases and traits. Nat Genet 47, 1236–1241 (2015).

32. Salmenkivi, J., Sund, R., Paavola, M., Ruuth, I. & Malmivaara, A. Mortality Caused by Surgery for Degenerative Lumbar Spine. Spine (Phila Pa 1976) 42, 1080–1087 (2017).

33. Leitsalu, L., Alavere, H., Tammesoo, M.-L., Leego, E. & Metspalu, A. Linking a population biobank with national health registries-the estonian experience. J Pers Med 5, 96–106 (2015).

34. Bycroft, C. et al. The UK Biobank resource with deep phenotyping and genomic data. Nature 562, 203–209 (2018).

35. Mitt, M. et al. Improved imputation accuracy of rare and low-frequency variants using population-specific high-coverage WGS-based imputation reference panel. Eur J Hum Genet 25, 869–876 (2017).

36. Howie, B. N., Donnelly, P. & Marchini, J. A flexible and accurate genotype imputation method for the next generation of genome-wide association studies. PLoS Genet 5, e1000529 (2009).

37. Mbatchou, J. et al. Computationally efficient whole-genome regression for quantitative and binary traits. Nat Genet 53, 1097–1103 (2021).

38. Willer, C. J., Li, Y. & Abecasis, G. R. METAL: fast and efficient meta-analysis of genomewide association scans. Bioinformatics 26, 2190–2191 (2010).

39. Clark, K., Karsch-Mizrachi, I., Lipman, D. J., Ostell, J. & Sayers, E. W. GenBank. Nucleic Acids Res 44, D67–D72 (2016).

40. UniProt: a worldwide hub of protein knowledge. Nucleic Acids Res 47, D506–D515 (2019).

41. Stanfill, A. G. & Cao, X. Enhancing Research Through the Use of the Genotype-Tissue Expression (GTEx) Database. Biol Res Nurs 23, 533–540 (2021).

42. Yang, J., Lee, S. H., Goddard, M. E. & Visscher, P. M. GCTA: a tool for genome-wide complex trait analysis. Am J Hum Genet 88, 76–82 (2011).

43. Bakshi, A. et al. Fast set-based association analysis using summary data from GWAS identifies novel gene loci for human complex traits. Sci Rep 6, (2016).

44. Giambartolomei, C. et al. Bayesian test for colocalisation between pairs of genetic association studies using summary statistics. PLoS Genet 10, e1004383 (2014).

