## Supplementary material for "Genetic architecture of lumbar spinal stenosis": Regional association plots

[1] Research Unit of Health Sciences and Technology, Faculty of Medicine, University of Oulu, Oulu, Finland
[2] Biocenter Oulu, Oulu, Finland
[3] Medical Research Center Oulu, Oulu University Hospital and University of Oulu, Oulu, Finland
[4] Research unit of Population Health, Faculty of Medicine, and Biocenter Oulu, University of Oulu, Oulu, Finland
[5] A list of FinnGen authors and their affiliations available in the supplements
[6] Estonian Genome Centre, Institute of Genomics, University of Tartu, Tartu, Estonia
[7] A list of Estonian Biobank Research Team authors and their affiliations available in the supplements
[8] Institute for Molecular Medicine Finland (FIMM), HiLIFE, University of Helsinki
[9] Neurosurgery, Institute of Clinical Medicine, School of Medicine, Faculty of Health Sciences, University of Eastern Finland and Department of Neurosurgery, Kuopio University Hospital, Kuopio, Finland
[10] Rehabilitation Services of Wellbeing Services County of South Karelia, Lappeenranta, Finland
* These authors contributed equally to this work

**Supplementary Note.** FinnGen DF9 Ethics statement
**Supplementary Data 1** Regional association plot on chr1 near *RUNX3*
**Supplementary Data 2** Regional association plot on chr1 near *NGF*
**Supplementary Data 3** Regional association plot on chr1 near *OTUD7B*
**Supplementary Data 4** Regional association plot on chr1 near *ZNF687*
**Supplementary Data 5** Regional association plot on chr1 near *PTPRC*
**Supplementary Data 6** Regional association plot on chr2 near *USP37*
**Supplementary Data 7** Regional association plot on chr3 near *DAG1*
**Supplementary Data 8** Regional association plot on chr4 near *RAB28*
**Supplementary Data 9** Regional association plot on chr5 near *DAB2*
**Supplementary Data 10** Regional association plot on chr5 near *ITGA2*
**Supplementary Data 11** Regional association plot on chr5 near *EBF1*
**Supplementary Data 12** Regional association plot on chr6 near *BMP6*
**Supplementary Data 13** Regional association plot on chr6 near *TRIM38*
**Supplementary Data 14** Regional association plot on chr6 near *HLA*
**Supplementary Data 15** Regional association plot on chr6 near *HLA*
**Supplementary Data 16** Regional association plot on chr6 near *HLA*
**Supplementary Data 17** Regional association plot on chr6 near *CDC5L*
**Supplementary Data 18** Regional association plot on chr6 near *LIN28B*
**Supplementary Data 19** Regional association plot on chr6 near *EYA4*
**Supplementary Data 20** Regional association plot on chr7 near *FOXK1*
**Supplementary Data 21** Regional association plot on chr7 near *TWIST1*
**Supplementary Data 22** Regional association plot on chr7 near *JAZF1*
**Supplementary Data 23** Regional association plot on chr7 near *Empty*
**Supplementary Data 24** Regional association plot on chr10 near *MKX*
**Supplementary Data 25** Regional association plot on chr11 near *MPPED2*
**Supplementary Data 26** Regional association plot on chr12 near *SOX5*
**Supplementary Data 27** Regional association plot on chr12 near *CCDC91*
**Supplementary Data 28** Regional association plot on chr12 near *GLI1*
**Supplementary Data 29** Regional association plot on chr12 near *USP15*
**Supplementary Data 30** Regional association plot on chr12 near *ZCCHC8*
**Supplementary Data 31** Regional association plot on chr13 near *TNFSF11*
**Supplementary Data 32** Regional association plot on chr14 near *MMP14*
**Supplementary Data 33** Regional association plot on chr14 near *GPR132*
**Supplementary Data 34** Regional association plot on chr15 near *ALDH1A2*
**Supplementary Data 35** Regional association plot on chr15 near *PPIB*
**Supplementary Data 36** Regional association plot on chr15 near *PDE8A*
**Supplementary Data 37** Regional association plot on chr16 near *GAN*
**Supplementary Data 38** Regional association plot on chr17 near *SOCS7*
**Supplementary Data 39** Regional association plot on chr17 near *AXIN2*
**Supplementary Data 40** Regional association plot on chr17 near Empty
**Supplementary Data 41** Regional association plot on chr18 near *DCC*
**Supplementary Data 42** Regional association plot on chr19 near *CACTIN*
**Supplementary Data 43** Regional association plot on chr19 near *CRLF1*
**Supplementary Data 44** Regional association plot on chr20 near *DDRGK*
**Supplementary Data 45** Regional association plot on chr21 near *ETS2*
**Supplementary Data 46** Regional association plot on chr22 near *OSM*
**Supplementary Data 47** Regional association plot on chrX near *CHRDL1*

**Supplementary Note.** FinnGen R9 Ethics statement

Patients and control subjects in FinnGen provided informed consent for biobank research, based on the Finnish Biobank Act. Alternatively, separate research cohorts, collected prior the Finnish Biobank Act came into effect (in September 2013) and start of FinnGen (August 2017), were collected based on study-specific consents and later transferred to the Finnish biobanks after approval by Fimea (Finnish Medicines Agency), the National Supervisory Authority for Welfare and Health. Recruitment protocols followed the biobank protocols approved by Fimea. The Coordinating Ethics Committee of the Hospital District of Helsinki and Uusimaa (HUS) statement number for the FinnGen study is Nr HUS/990/2017.

The FinnGen study is approved by Finnish Institute for Health and Welfare (permit numbers: THL/2031/6.02.00/2017, THL/1101/5.05.00/2017, THL/341/6.02.00/2018, THL/2222/6.02.00/2018, THL/283/6.02.00/2019, THL/1721/5.05.00/2019 and THL/1524/5.05.00/2020), Digital and population data service agency (permit numbers: VRK43431/2017-3, VRK/6909/2018-3, VRK/4415/2019-3), the Social Insurance Institution (permit numbers: KELA 58/522/2017, KELA 131/522/2018, KELA 70/522/2019, KELA 98/522/2019, KELA 134/522/2019, KELA 138/522/2019, KELA 2/522/2020, KELA 16/522/2020), Findata permit numbers THL/2364/14.02/2020, THL/4055/14.06.00/2020,,THL/3433/14.06.00/2020, THL/4432/14.06/2020, THL/5189/14.06/2020, THL/5894/14.06.00/2020, THL/6619/14.06.00/2020, THL/209/14.06.00/2021, THL/688/14.06.00/2021, THL/1284/14.06.00/2021, THL/1965/14.06.00/2021, THL/5546/14.02.00/2020, THL/2658/14.06.00/2021, THL/4235/14.06.00/202, Statistics Finland (permit numbers: TK-53-1041-17 and TK/143/07.03.00/2020 (earlier TK-53-90-20) TK/1735/07.03.00/2021, TK/3112/07.03.00/2021) and Finnish Registry for Kidney Diseases permission/extract from the meeting minutes on 4$^{th}$ July 2019.

The Biobank Access Decisions for FinnGen samples and data utilized in FinnGen Data Freeze 9 include: THL Biobank BB2017_55, BB2017_111, BB2018_19, BB_2018_34, BB_2018_67, BB2018_71, BB2019_7, BB2019_8, BB2019_26, BB2020_1, Finnish Red Cross Blood Service Biobank 7.12.2017, Helsinki Biobank HUS/359/2017, HUS/248/2020, Auria Biobank AB17-5154 and amendment #1 (August 17 2020), AB20-5926 and amendment #1 (April 23 2020) and it´s modification (Sep 22 2021), Biobank Borealis of Northern Finland_2017_1013, Biobank of Eastern Finland 1186/2018 and amendment 22 § /2020, Finnish Clinical Biobank Tampere MH0004 and amendments (21.02.2020 & 06.10.2020), Central Finland Biobank 1-2017, and Terveystalo Biobank STB 2018001 and amendment 25$^{th}$ Aug 2020.

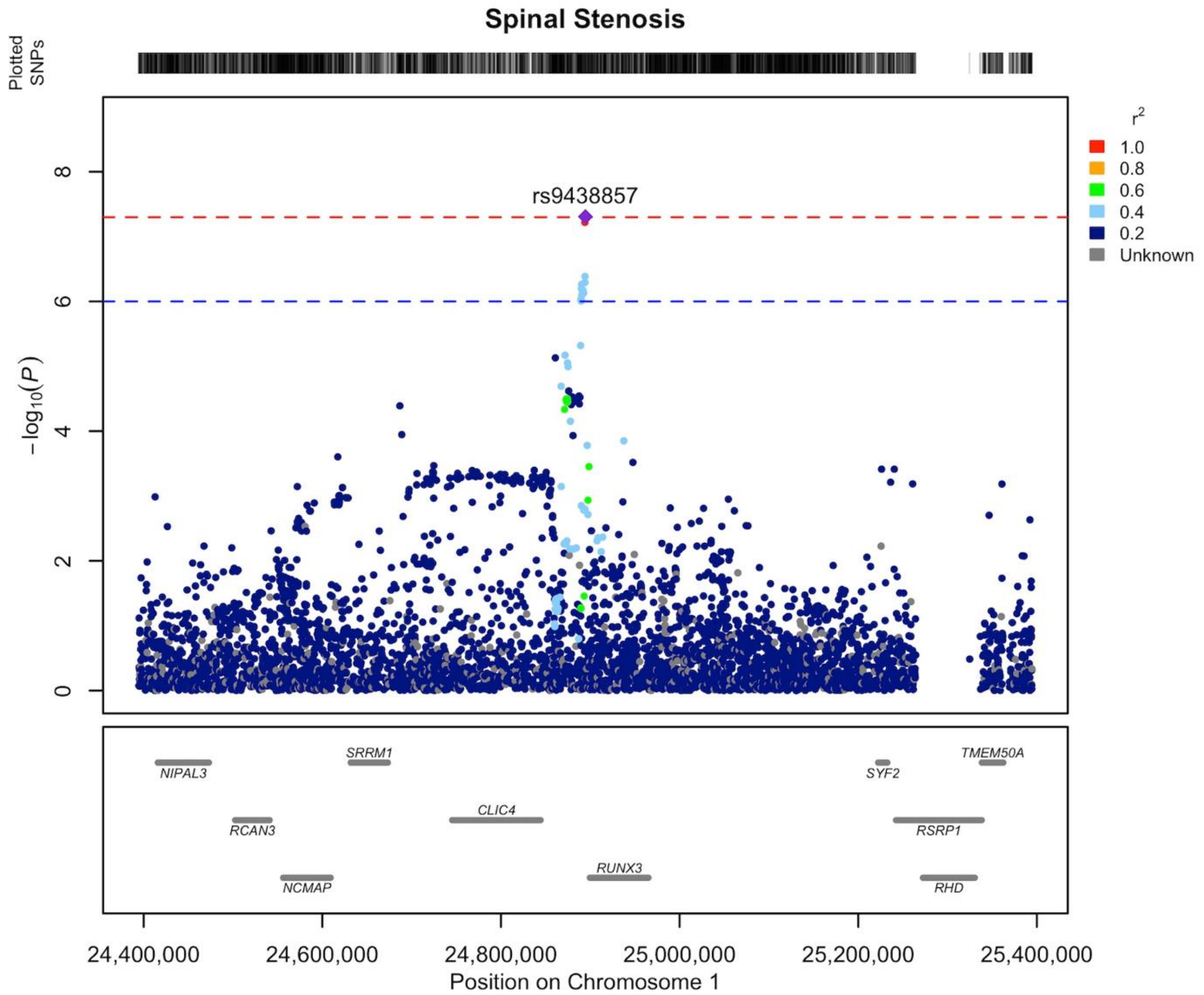


**Supplementary Data 1** Regional association plot of novel LSS association on chromosome 1 area of 24.4-25.4 MB. Our findings indicate that the gene responsible for the locus's association is most likely *RUNX3 (RUNX family transcription factor 3).* The LD structure of the plot does not fully represent the meta-analysis as it was calculated using FinnGen. The Locuszooms (https://github.com/Geeketics/LocusZooms) R package was used to generate the plot, and the Ensembl archive (https://jul2023.archive.ensembl.org) was used for the gene list.

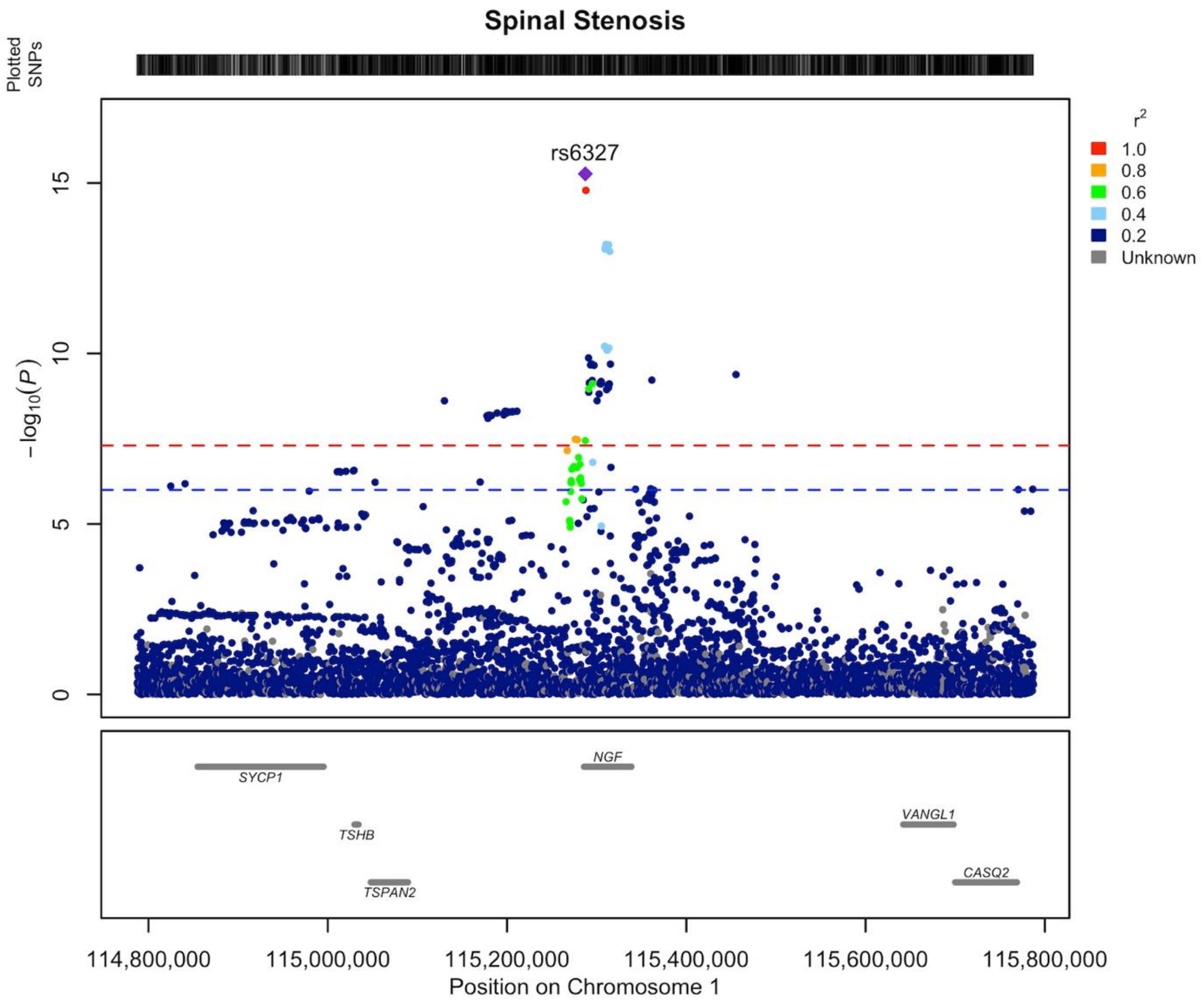


**Supplementary Data 2** Regional association plot of novel LSS association on chromosome 1 area of 114.8-115.8 MB. Our findings indicate that the gene responsible for the locus's association is most likely *NGF (nerve growth factor)*. The LD structure of the plot does not fully represent the meta-analysis as it was calculated using FinnGen. The Locuszooms (https://github.com/Geeketics/LocusZooms) R package was used to generate the plot, and the Ensembl archive (https://jul2023.archive.ensembl.org) was used for the gene list.

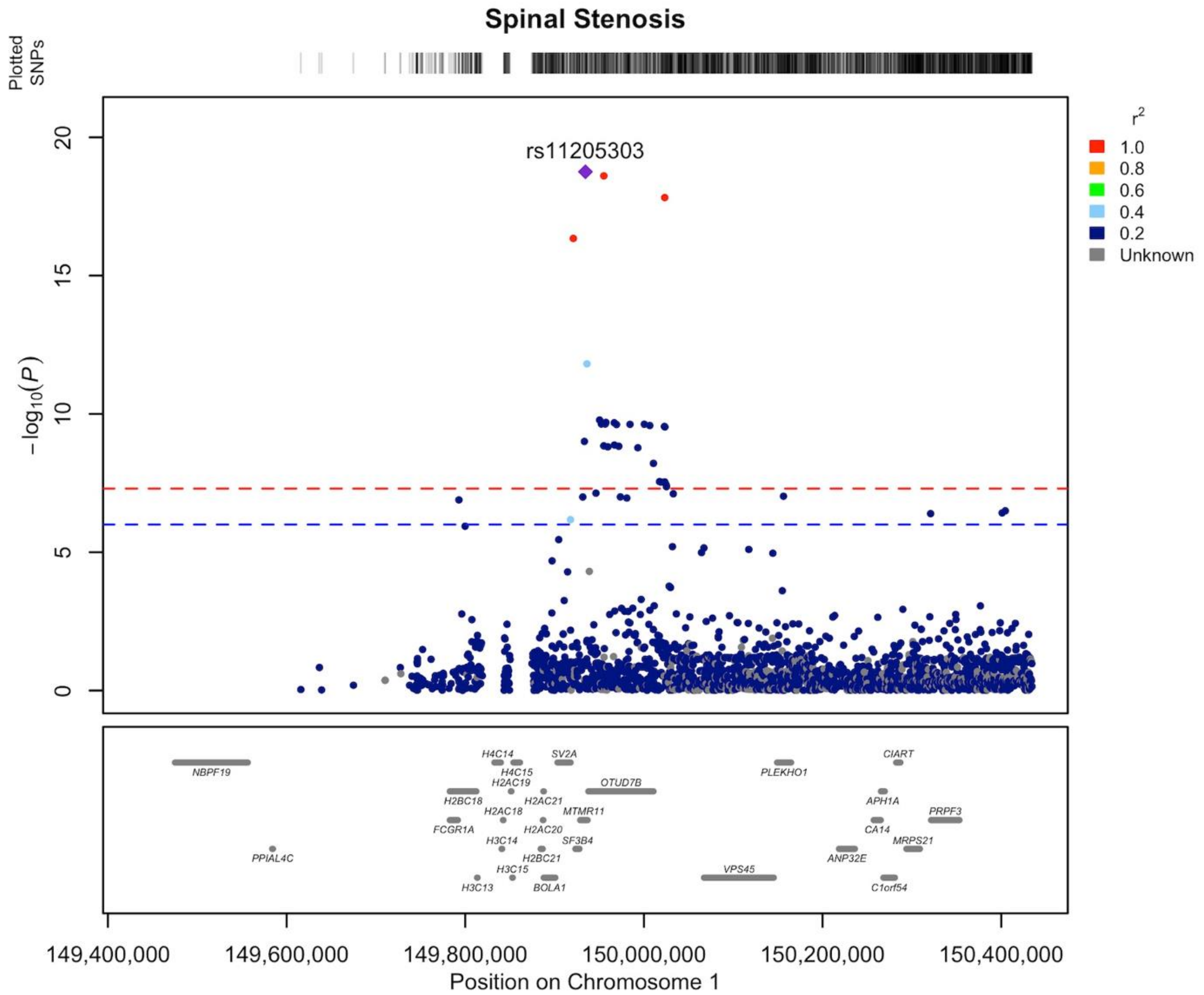


**Supplementary Data 3** Regional association plot of novel LSS association on chromosome 1 area of 149.4-150.4 MB. Our findings indicate that the gene responsible for the locus's association is most likely *OTUD7B (OTU deubiquitinase 7B)*. The LD structure of the plot does not fully represent the meta-analysis as it was calculated using FinnGen. The Locuszooms (https://github.com/Geeketics/LocusZooms) R package was used to generate the plot, and the Ensembl archive (https://jul2023.archive.ensembl.org) was used for the gene list.

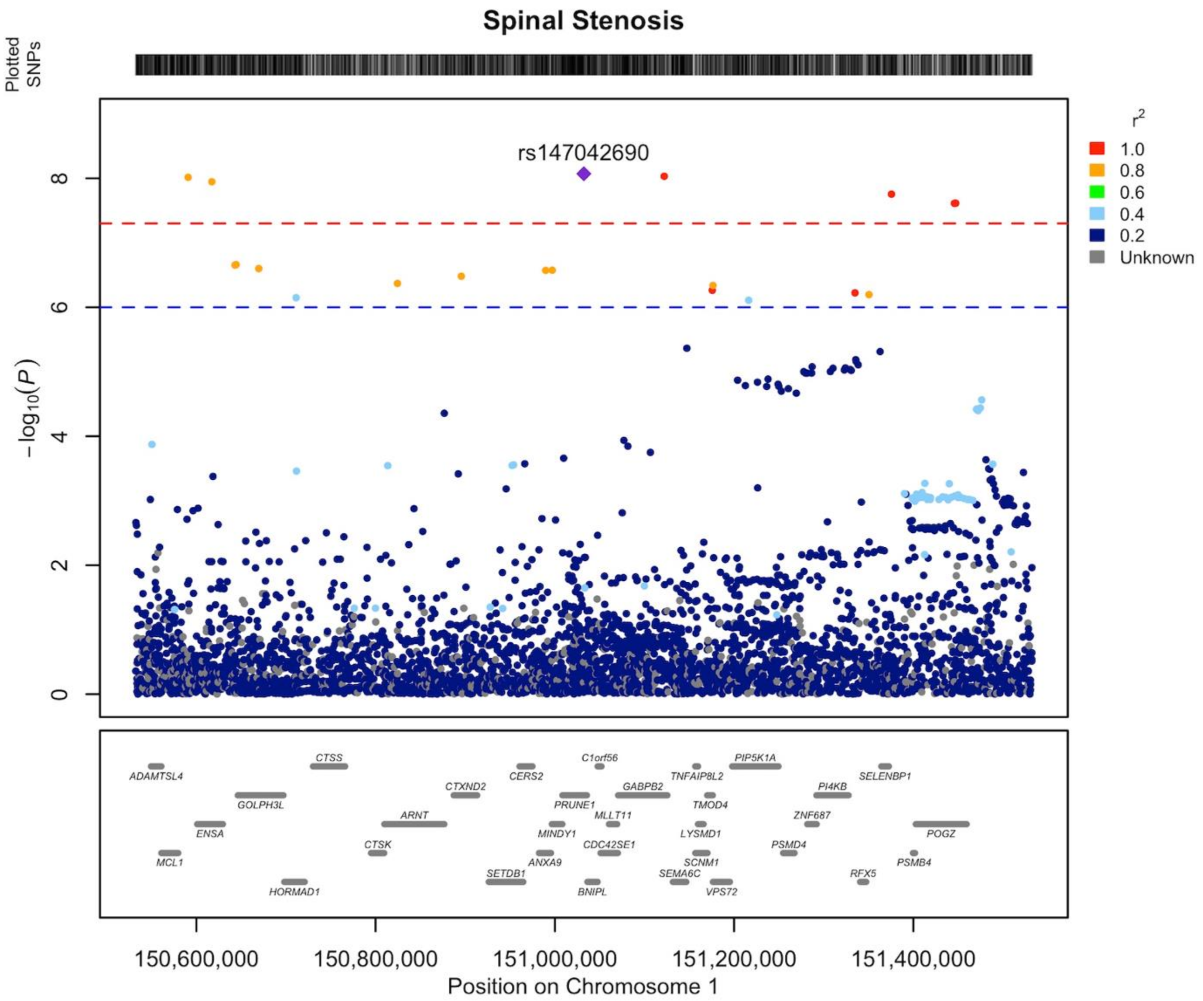


**Supplementary Data 4** Regional association plot of novel LSS association on chromosome 1 area of 150.5-151.5 MB. Our findings indicate that the gene responsible for the locus's association is most likely *ZNF687 (zinc finger protein 687).* The LD structure of the plot does not fully represent the meta-analysis as it was calculated using FinnGen. The Locuszooms (https://github.com/Geeketics/LocusZooms) R package was used to generate the plot, and the Ensembl archive (https://jul2023.archive.ensembl.org) was used for the gene list.

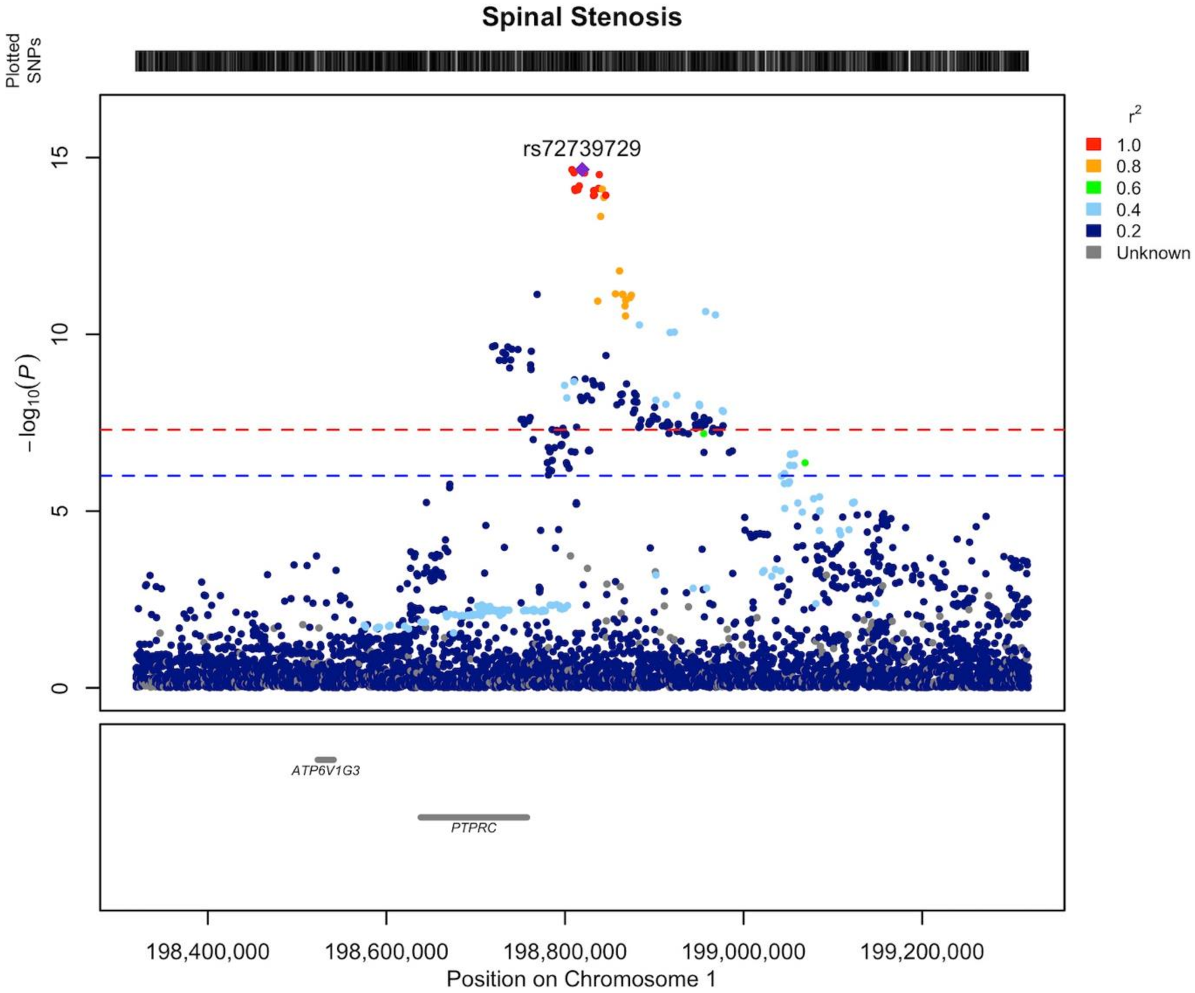


**Supplementary Data 5** Regional association plot of novel spinal stenosis association on chromosome 1 area of 198.3-199.3 MB. Our findings indicate that the gene responsible for the locus's association is most likely *PTPRC (protein tyrosine phosphatase receptor type C).* The LD structure of the plot does not fully represent the meta-analysis as it was calculated using FinnGen. The Locuszooms (https://github.com/Geeketics/LocusZooms) R package was used to generate the plot, and the Ensembl archive (https://jul2023.archive.ensembl.org) was used for the gene list.

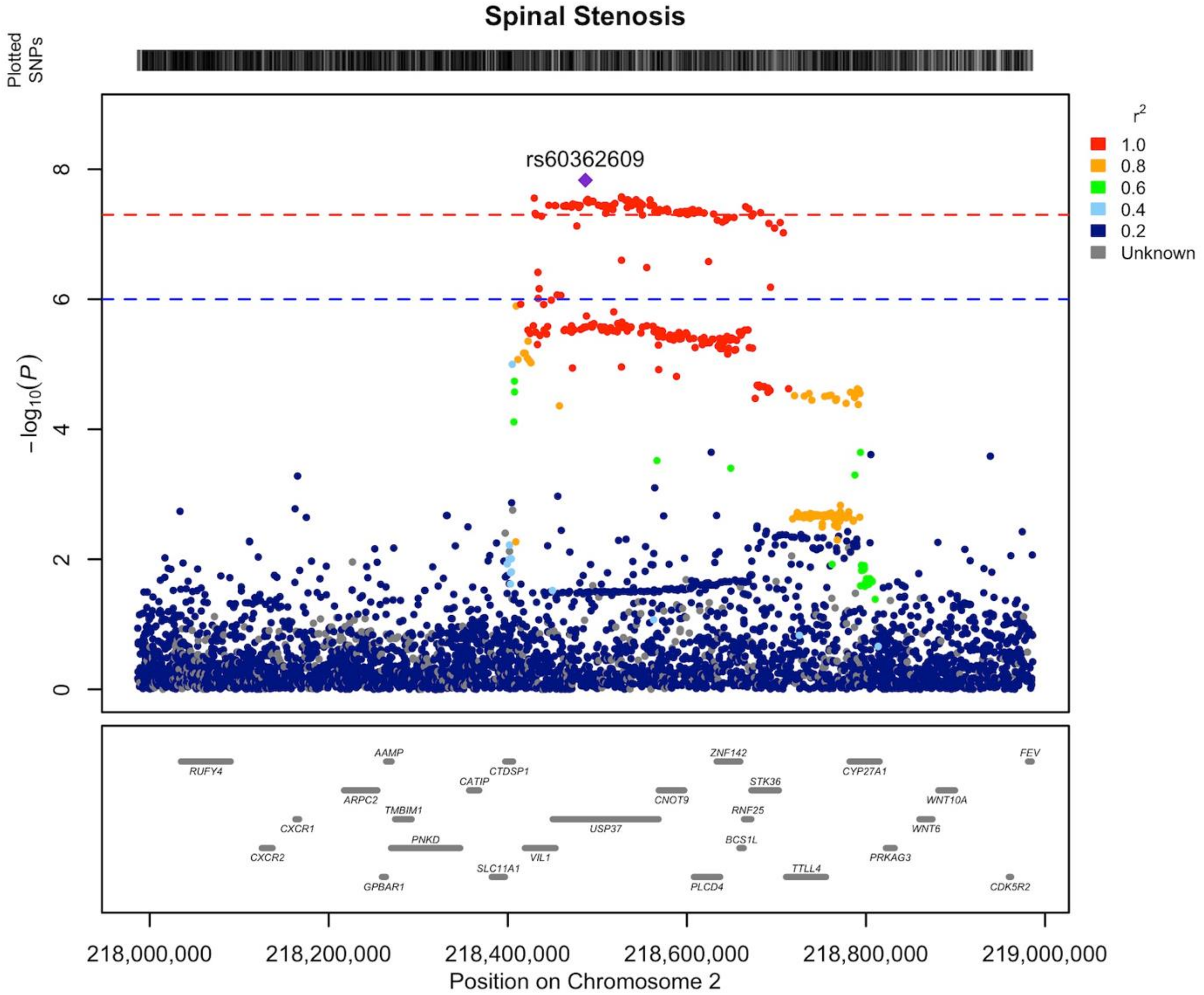


**Supplementary Data 6** Regional association plot of novel LSS association on chromosome 2 area of 217.9-218.9 MB. Our findings indicate that the gene responsible for the locus's association is most likely *USP37 (ubiquitin specific peptidase 37).* The LD structure of the plot does not fully represent the meta-analysis as it was calculated using FinnGen. The Locuszooms (https://github.com/Geeketics/LocusZooms) R package was used to generate the plot, and the Ensembl archive (https://jul2023.archive.ensembl.org) was used for the gene list.

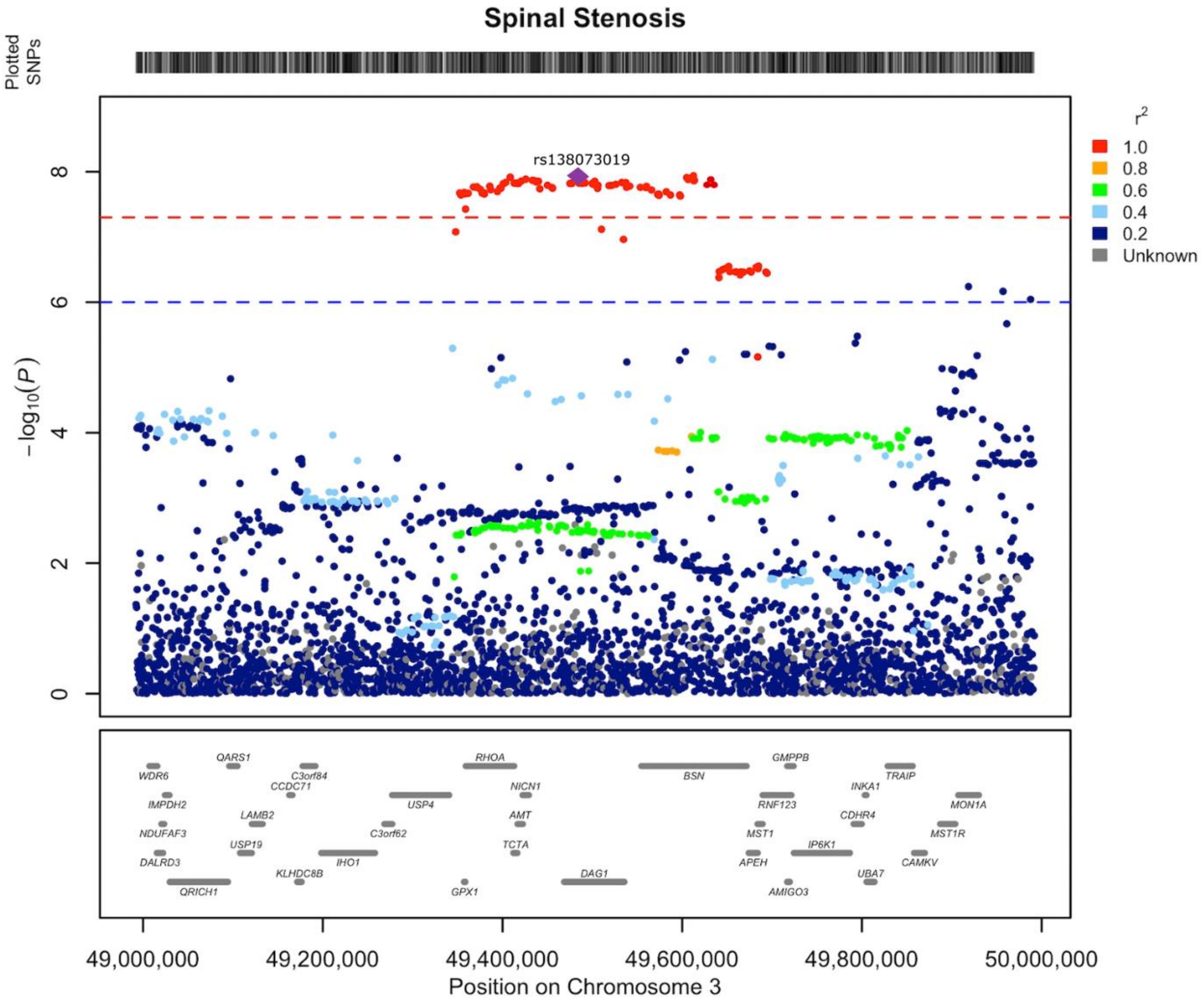


**Supplementary Data 7** Regional association plot of novel LSS association on chromosome 3 area of 49.0-50.0 MB. Our findings indicate that the gene responsible for the locus's association is most likely *DAG1 (dystroglycan 1)*. The LD structure of the plot does not fully represent the meta-analysis as it was calculated using FinnGen. The Locuszooms (https://github.com/Geeketics/LocusZooms) R package was used to generate the plot, and the Ensembl archive (https://jul2023.archive.ensembl.org) was used for the gene list.

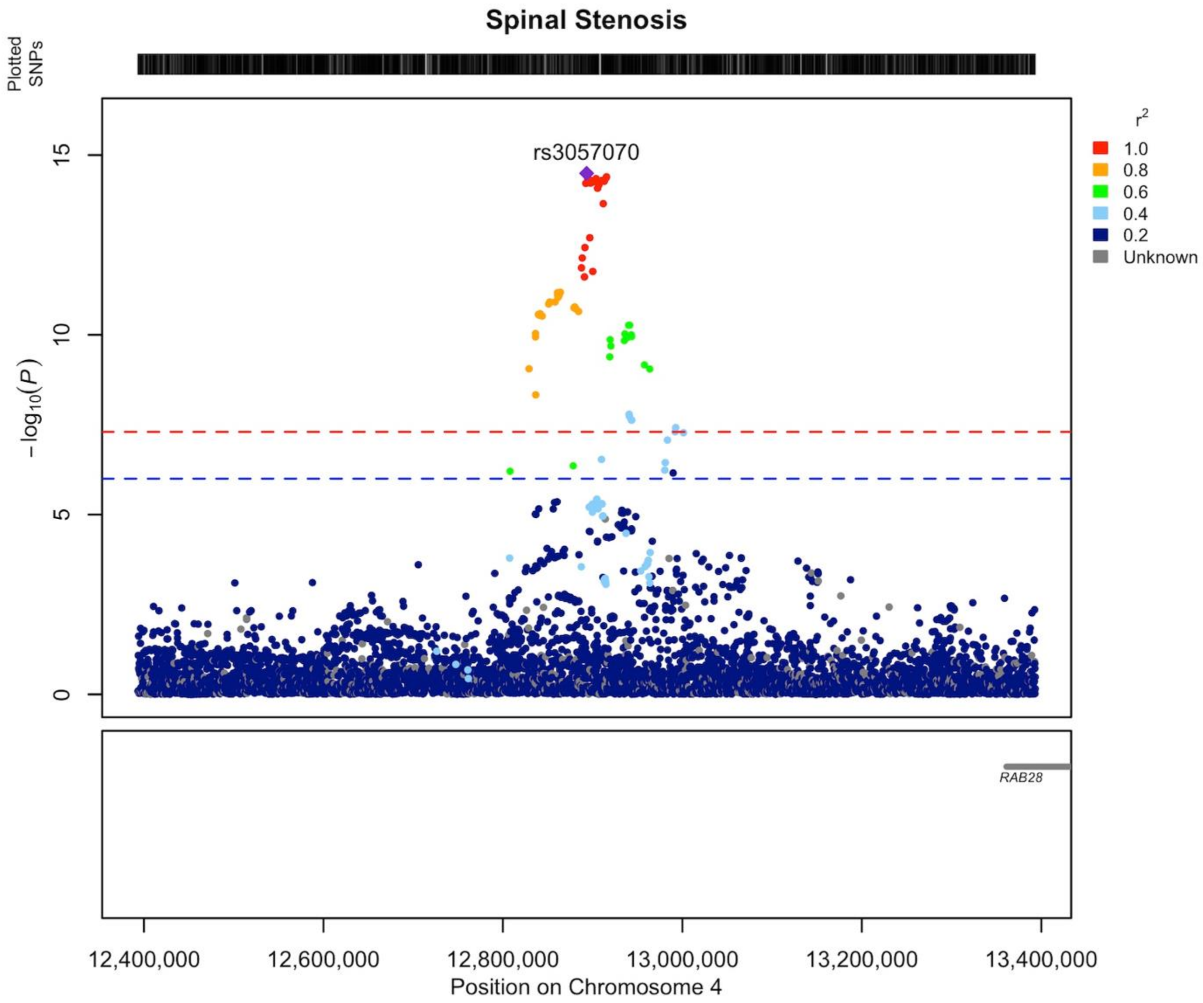


**Supplementary Data 8** Regional association plot of novel LSS association on chromosome 4 area of 12.4-13.4 MB. Our findings indicate that the gene responsible for the locus's association is most likely *RAB28 (RAB28, member RAS oncogene family).* The LD structure of the plot does not fully represent the meta-analysis as it was calculated using FinnGen. The Locuszooms (https://github.com/Geeketics/LocusZooms) R package was used to generate the plot, and the Ensembl archive (https://jul2023.archive.ensembl.org) was used for the gene list.

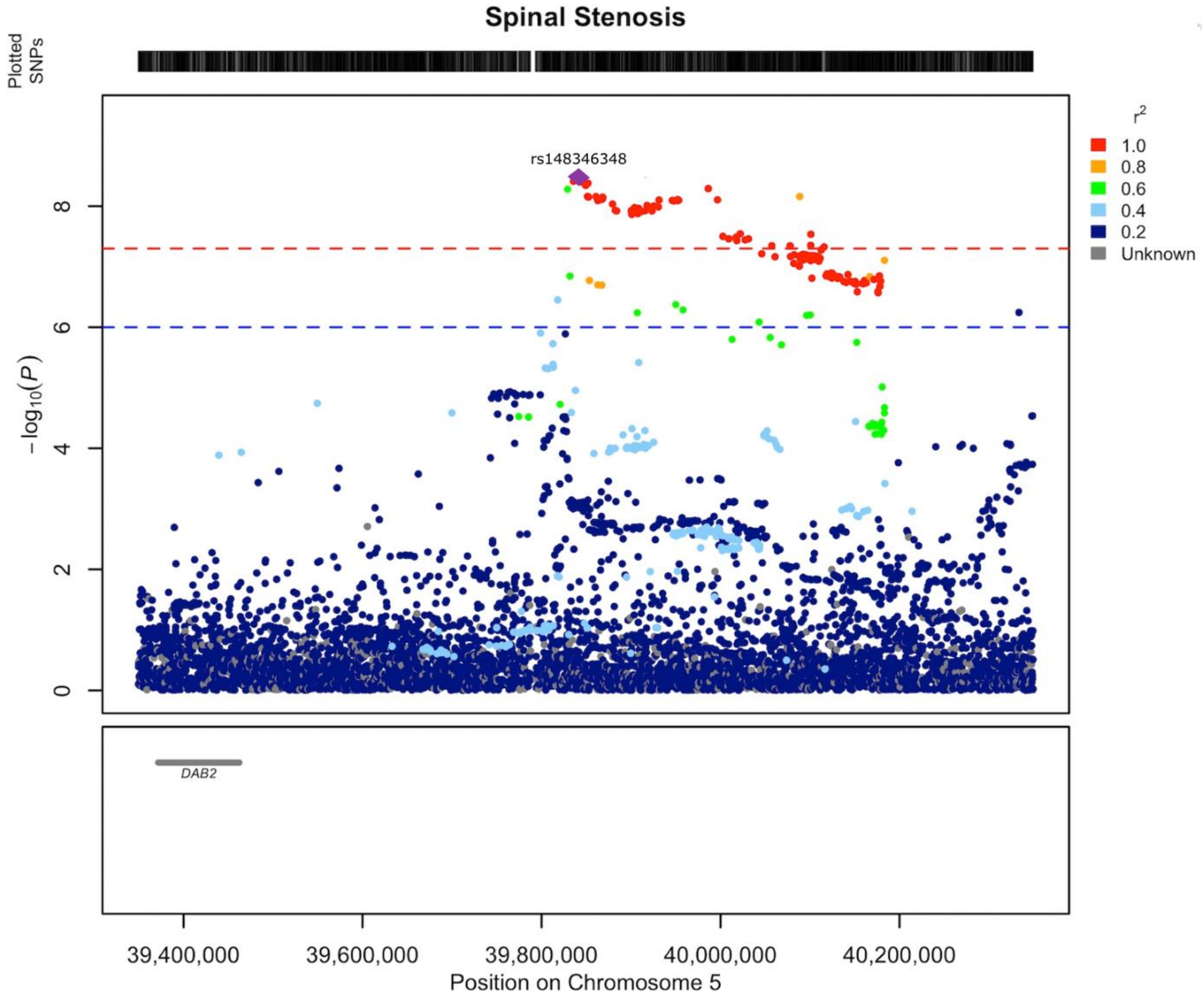


**Supplementary Data 9** Regional association plot of novel LSS association on chromosome 5 area of 39.3-40.3 MB. Our findings indicate that the gene responsible for the locus's association is most likely *DAB2 (DAB adaptor protein 2).* The LD structure of the plot does not fully represent the meta-analysis as it was calculated using FinnGen. The Locuszooms (https://github.com/Geeketics/LocusZooms) R package was used to generate the plot, and the Ensembl archive (https://jul2023.archive.ensembl.org) was used for the gene list.

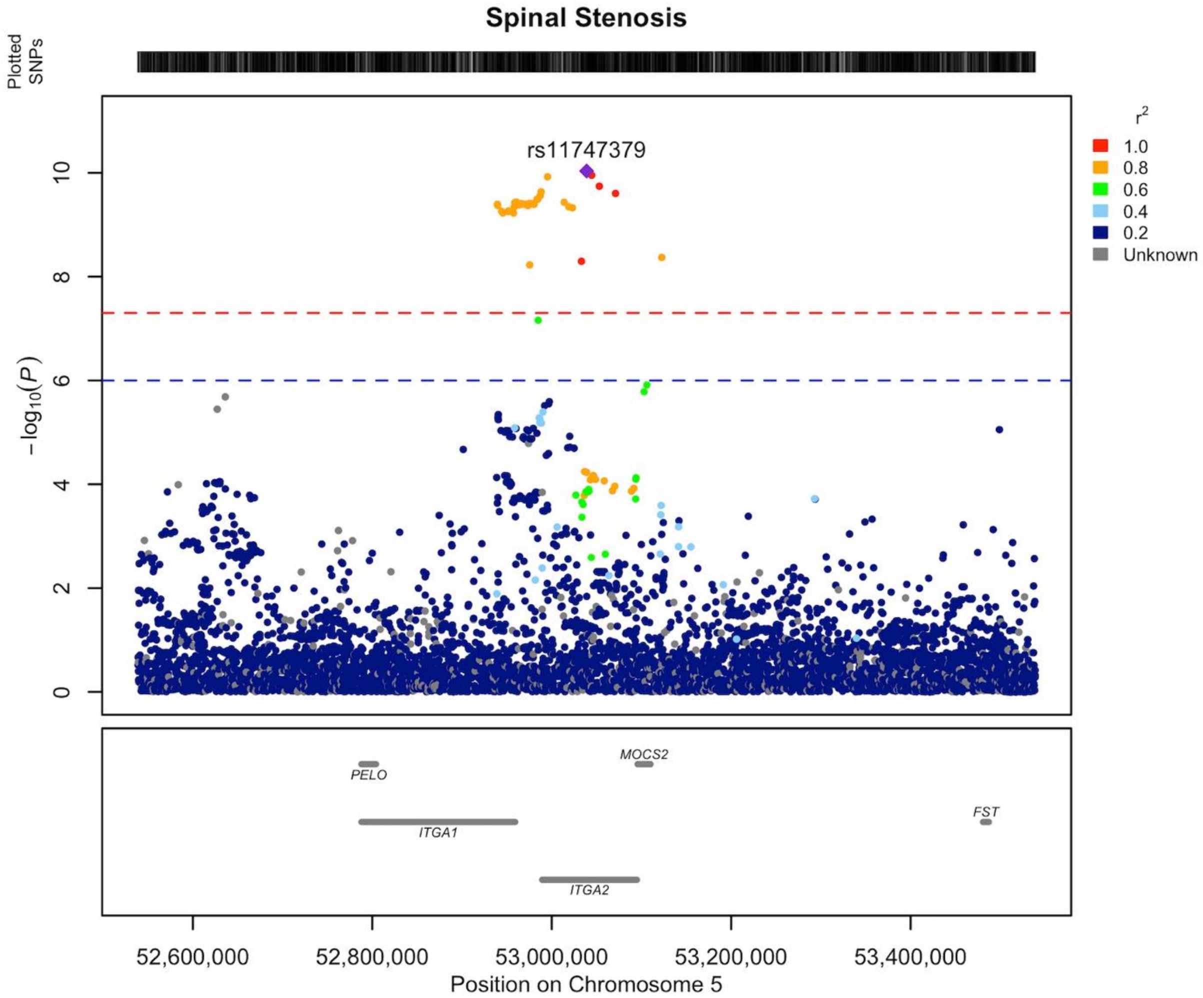


**Supplementary Data 10** Regional association plot of novel LSS association on chromosome 5 area of 52.5-53.5 MB. Our findings indicate that the gene responsible for the locus's association is most likely *ITGA2 (integrin subunit alpha 2).* The LD structure of the plot does not fully represent the meta-analysis as it was calculated using FinnGen. The Locuszooms (https://github.com/Geeketics/LocusZooms) R package was used to generate the plot, and the Ensembl archive (https://jul2023.archive.ensembl.org) was used for the gene list.

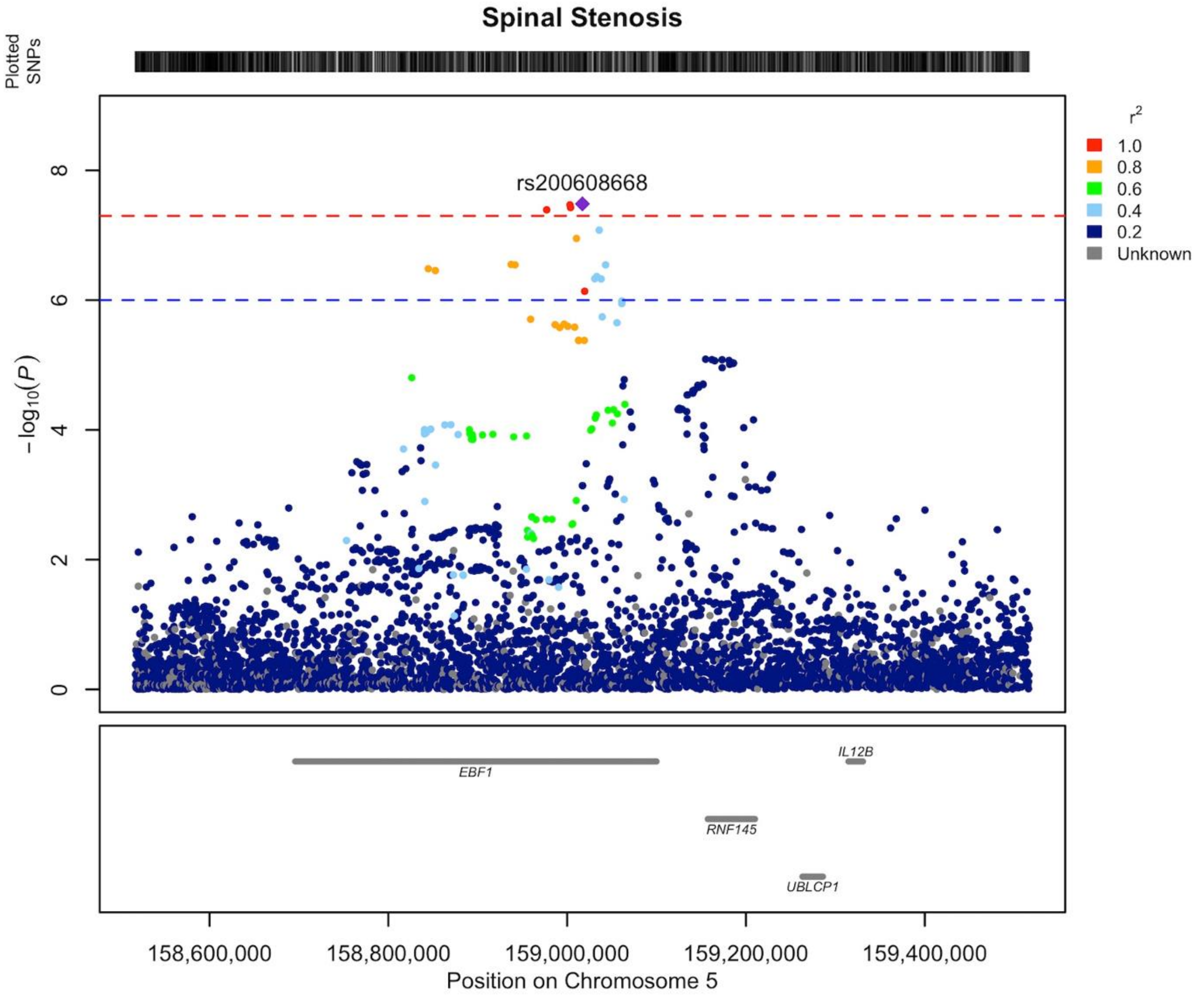


**Supplementary Data 11** Regional association plot of novel LSS association on chromosome 5 area of 158.5-159.5 MB. Our findings indicate that the gene responsible for the locus's association is most likely *EBF1 (EBF transcription factor 1).* The LD structure of the plot does not fully represent the meta-analysis as it was calculated using FinnGen. The Locuszooms (https://github.com/Geeketics/LocusZooms) R package was used to generate the plot, and the Ensembl archive (https://jul2023.archive.ensembl.org) was used for the gene list.

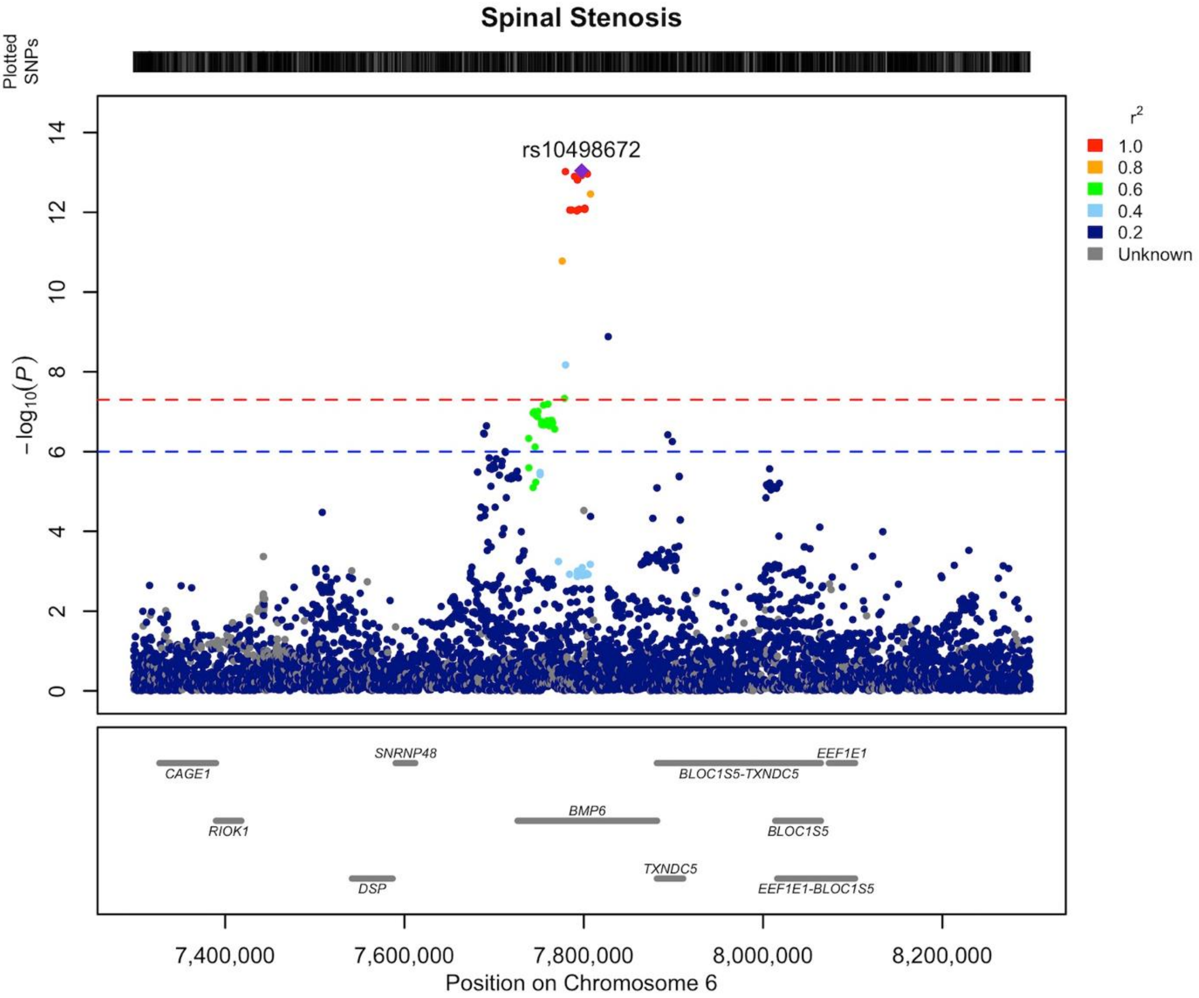


**Supplementary Data 12** Regional association plot of novel LSS association on chromosome 6 area of 7.3-8.3 MB. Our findings indicate that the gene responsible for the locus's association is most likely *BMP6 (bone morphogenetic protein 6).* The LD structure of the plot does not fully represent the meta-analysis as it was calculated using FinnGen. The Locuszooms (https://github.com/Geeketics/LocusZooms) R package was used to generate the plot, and the Ensembl archive (https://jul2023.archive.ensembl.org) was used for the gene list.

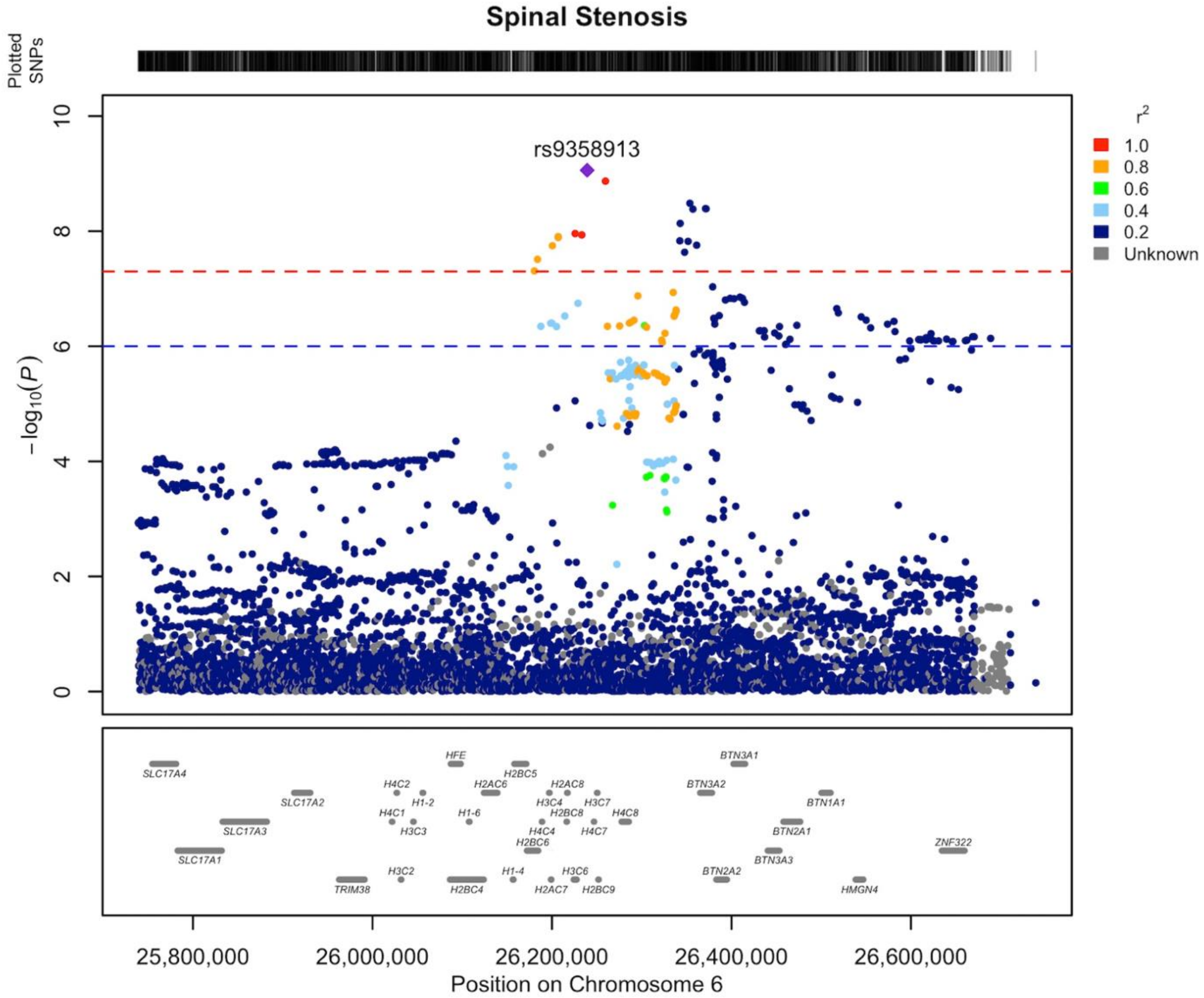


**Supplementary Data 13** Regional association plot of novel LSS association on chromosome 6 area of 25.7-26.7 MB. Our findings indicate that the gene responsible for the locus's association is most likely *TRIM38 (tripartite motif containing 38).* The LD structure of the plot does not fully represent the meta-analysis as it was calculated using FinnGen. The Locuszooms (https://github.com/Geeketics/LocusZooms) R package was used to generate the plot, and the Ensembl archive (https://jul2023.archive.ensembl.org) was used for the gene list.

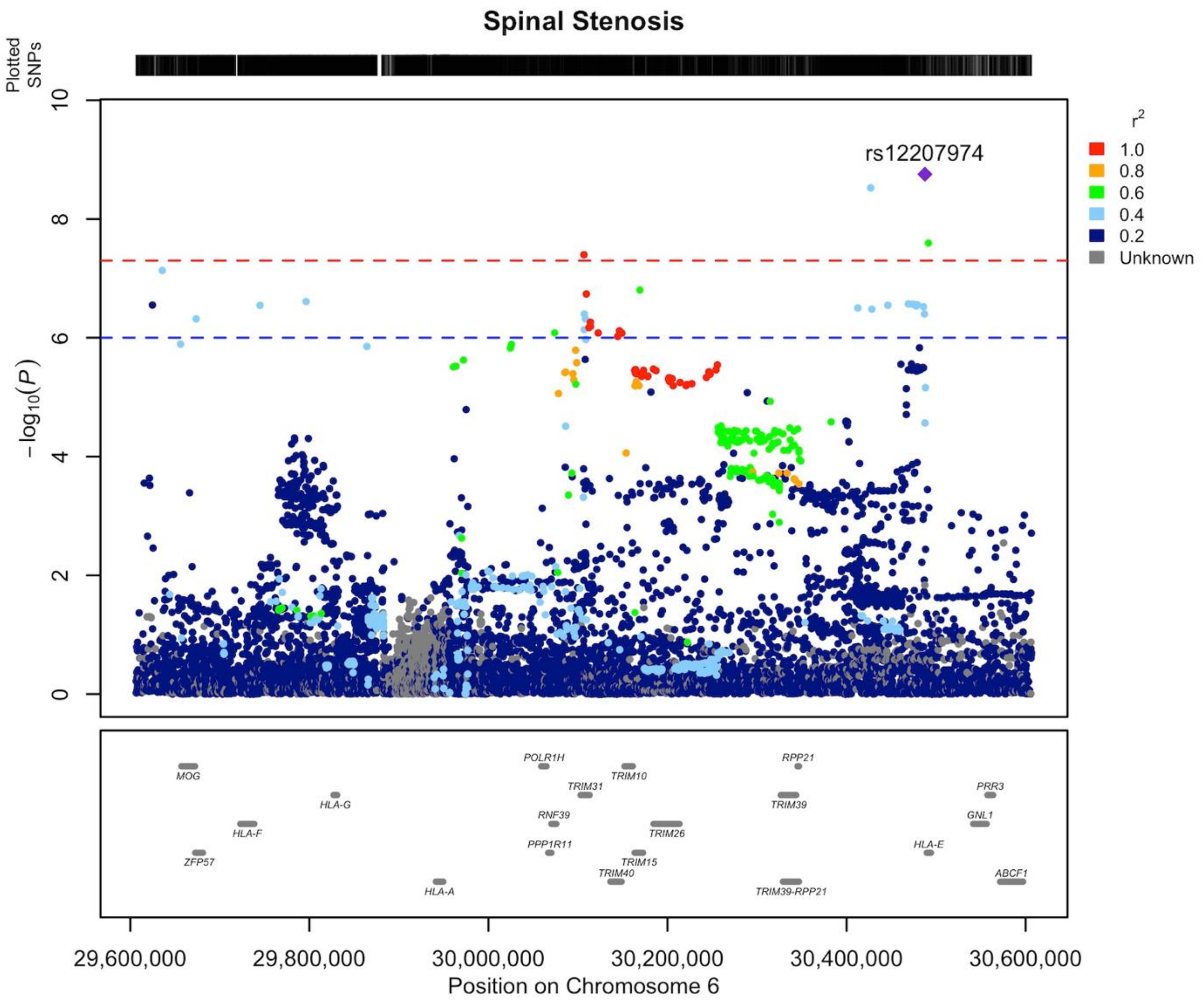


**Supplementary Data 14** Regional association plot of novel LSS association on chromosome 6 area of 29.6-30.6 MB. Our findings indicate that the gene responsible for the locus's association is most likely *HLA*. The LD structure of the plot does not fully represent the meta-analysis as it was calculated using FinnGen. The Locuszooms (https://github.com/Geeketics/LocusZooms) R package was used to generate the plot, and the Ensembl archive (https://jul2023.archive.ensembl.org) was used for the gene list.

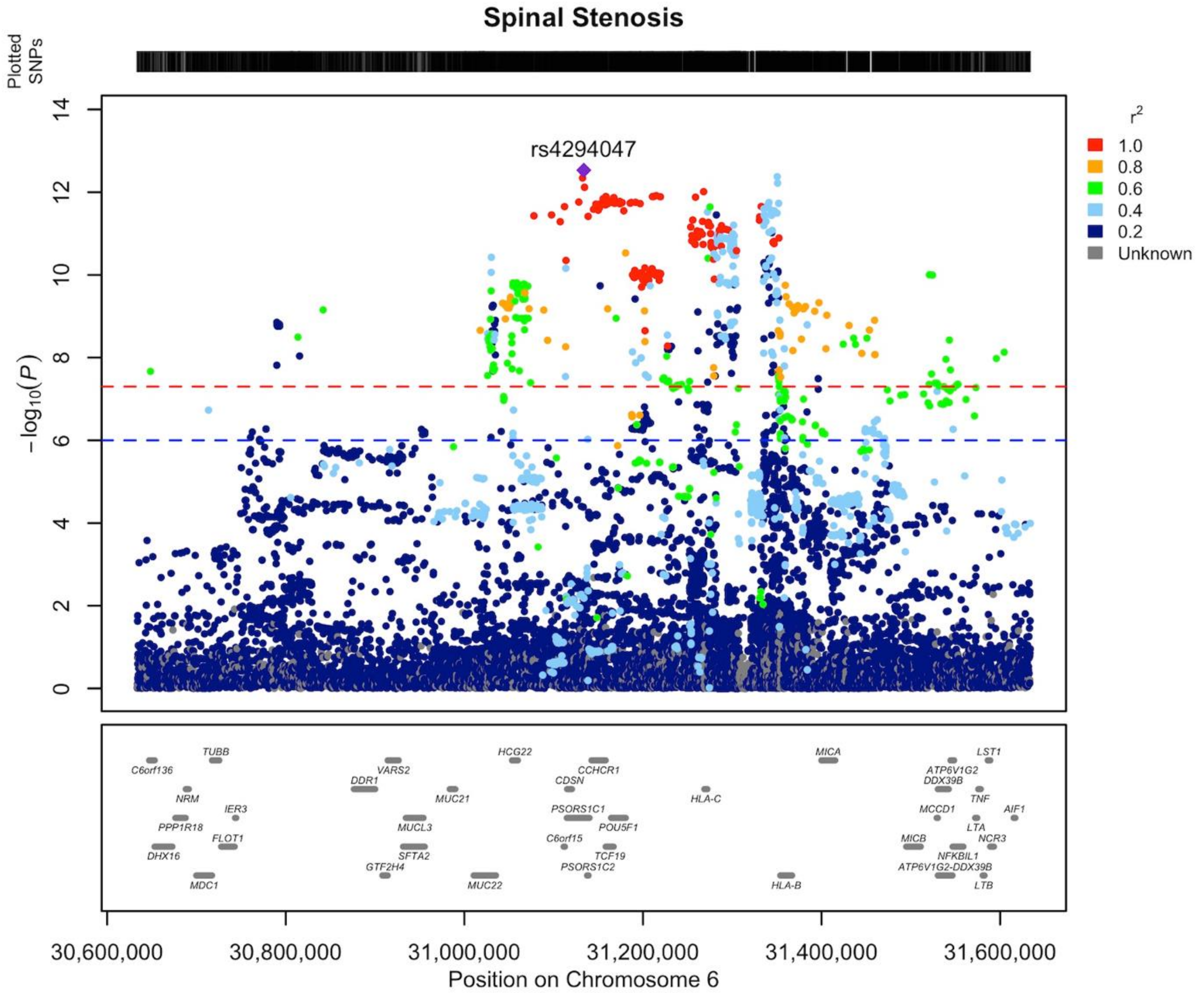


**Supplementary Data 15** Regional association plot of novel LSS association on chromosome 6 area of 30.6-31.6 MB. Our findings indicate that the gene responsible for the locus's association is most likely *HLA*. The LD structure of the plot does not fully represent the meta-analysis as it was calculated using FinnGen. The Locuszooms (https://github.com/Geeketics/LocusZooms) R package was used to generate the plot, and the Ensembl archive (https://jul2023.archive.ensembl.org) was used for the gene list.

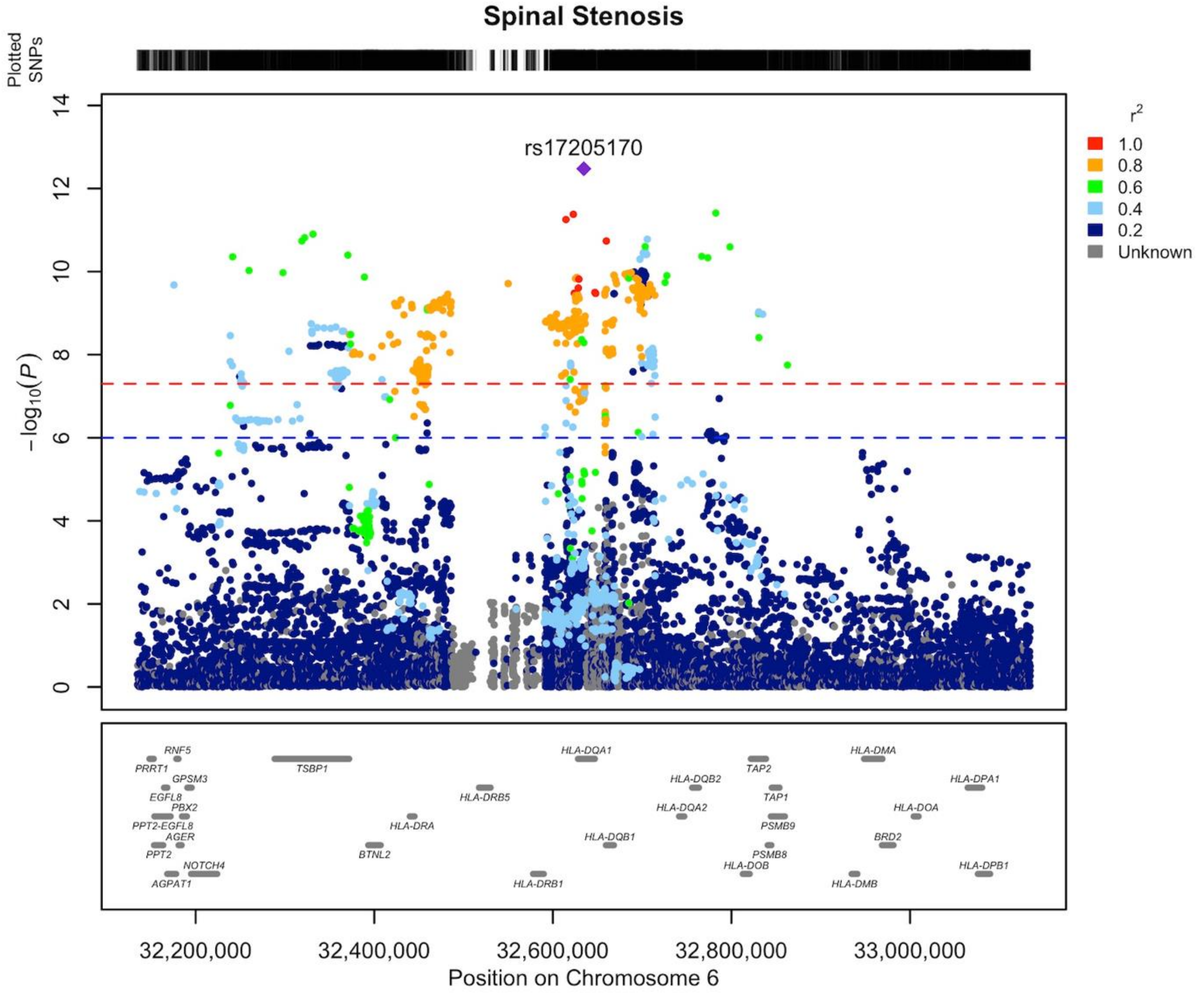


**Supplementary Data 16** Regional association plot of novel LSS association on chromosome 6 area of 32.1-33.1 MB. Our findings indicate that the gene responsible for the locus's association is most likely *HLA*. The LD structure of the plot does not fully represent the meta-analysis as it was calculated using FinnGen. The Locuszooms (https://github.com/Geeketics/LocusZooms) R package was used to generate the plot, and the Ensembl archive (https://jul2023.archive.ensembl.org) was used for the gene list.

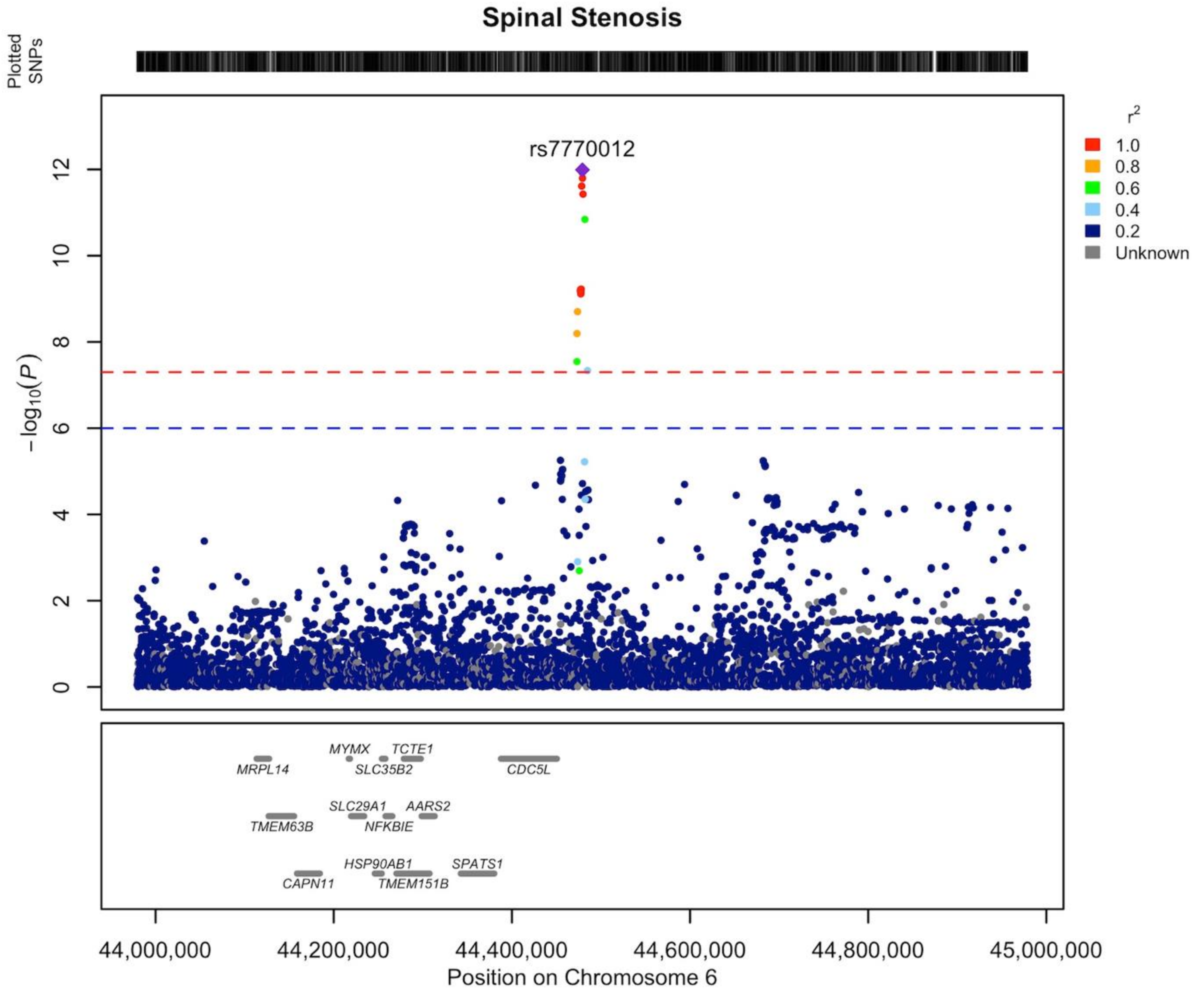


**Supplementary Data 17** Regional association plot of novel LSS association on chromosome 6 area of 43.9-44.9 MB. Our findings indicate that the gene responsible for the locus's association is most likely *CDC5L (cell division cycle 5 like).* The LD structure of the plot does not fully represent the meta-analysis as it was calculated using FinnGen. The Locuszooms (https://github.com/Geeketics/LocusZooms) R package was used to generate the plot, and the Ensembl archive (https://jul2023.archive.ensembl.org) was used for the gene list.

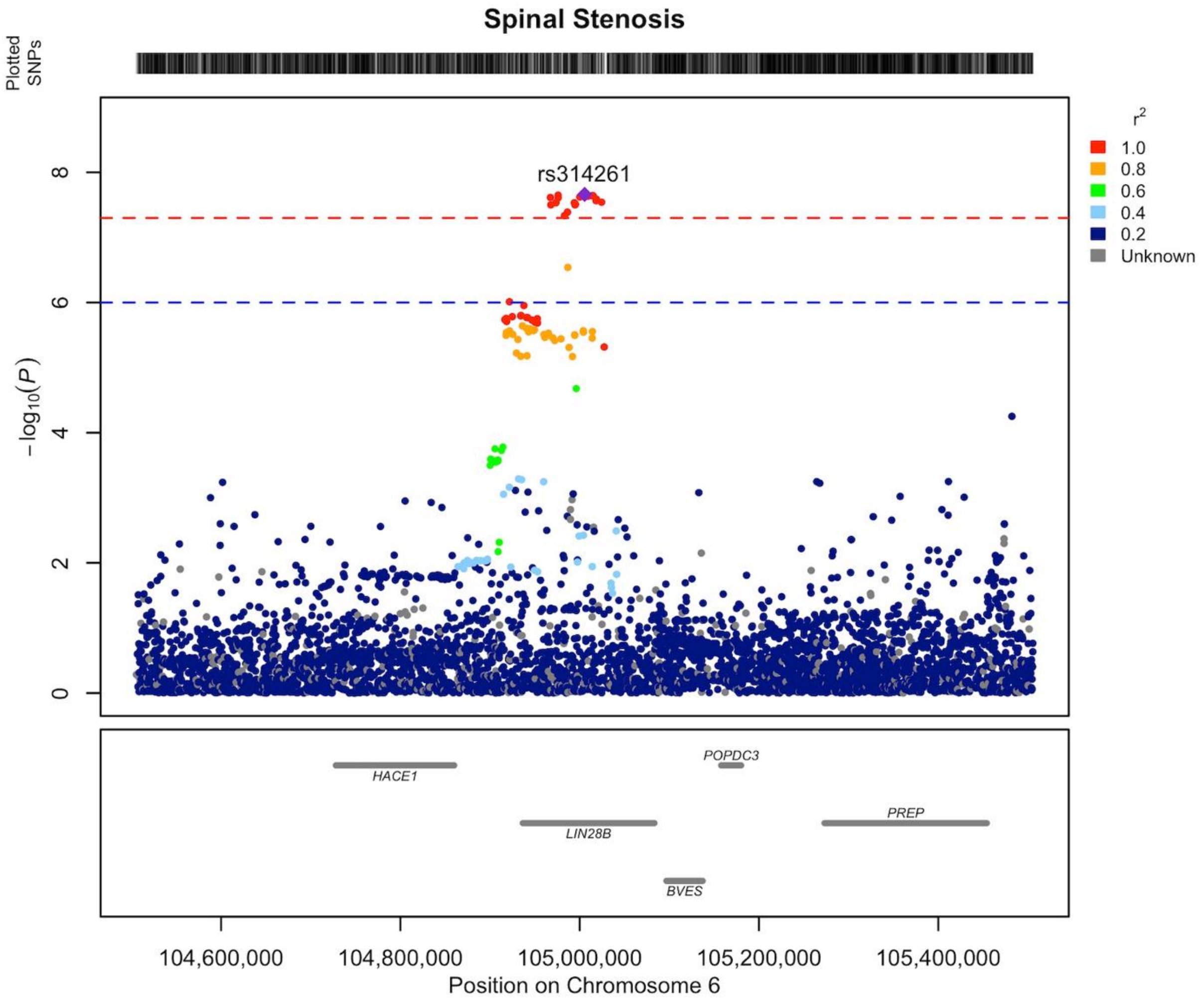


**Supplementary Data 18** Regional association plot of novel LSS association on chromosome 6 area of 104.5-105.5 MB. Our findings indicate that the gene responsible for the locus's association is most likely *LIN28B (lin-28 homolog B)*. The LD structure of the plot does not fully represent the meta-analysis as it was calculated using FinnGen. The Locuszooms (https://github.com/Geeketics/LocusZooms) R package was used to generate the plot, and the Ensembl archive (https://jul2023.archive.ensembl.org) was used for the gene list.

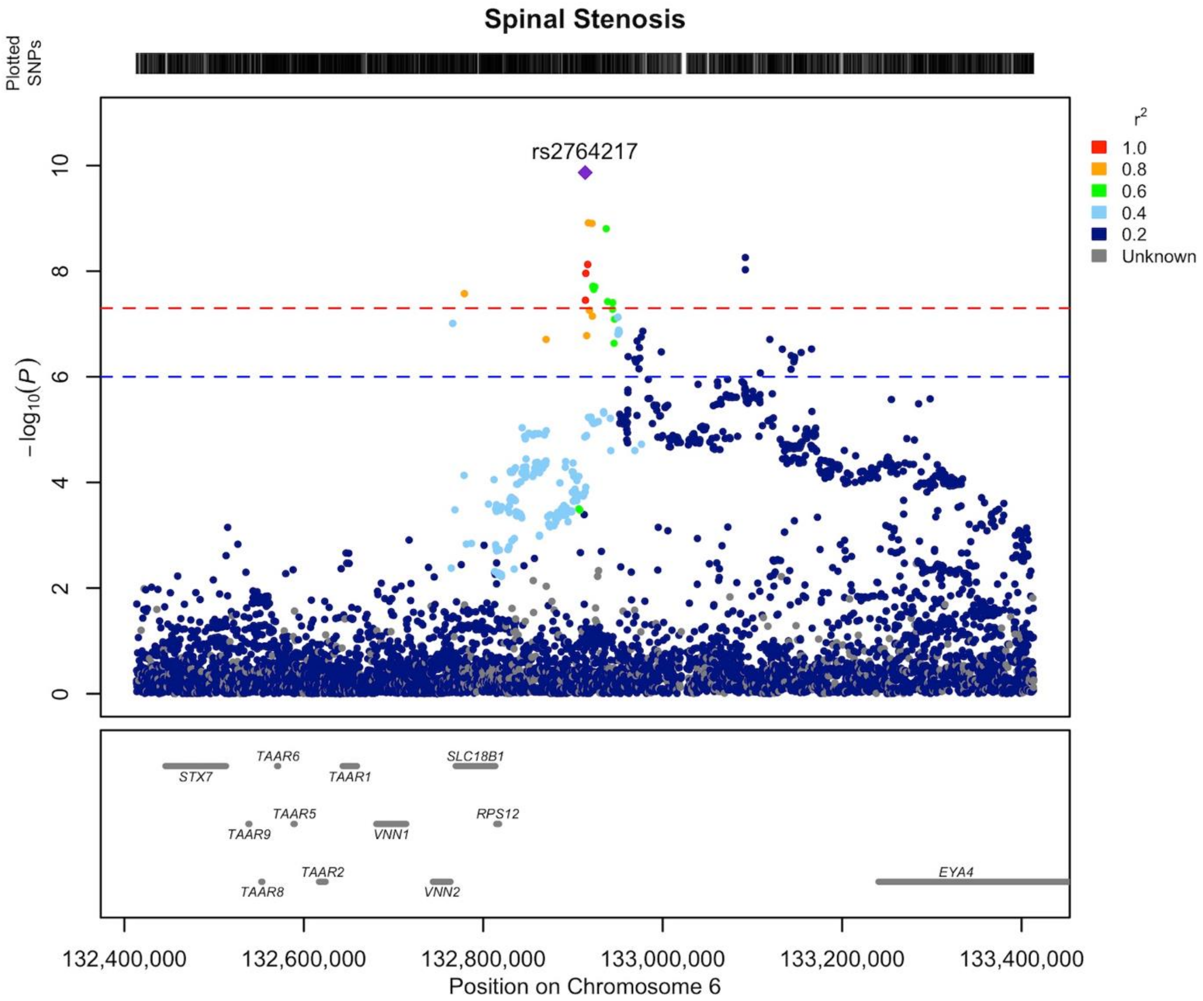


**Supplementary Data 19** Regional association plot of novel LSS association on chromosome 6 area of 132.4-133.4 MB. Our findings indicate that the gene responsible for the locus's association is most likely *EYA4 (EYA transcriptional coactivator and phosphatase 4).* The LD structure of the plot does not fully represent the meta-analysis as it was calculated using FinnGen. The Locuszooms (https://github.com/Geeketics/LocusZooms) R package was used to generate the plot, and the Ensembl archive (https://jul2023.archive.ensembl.org) was used for the gene list.

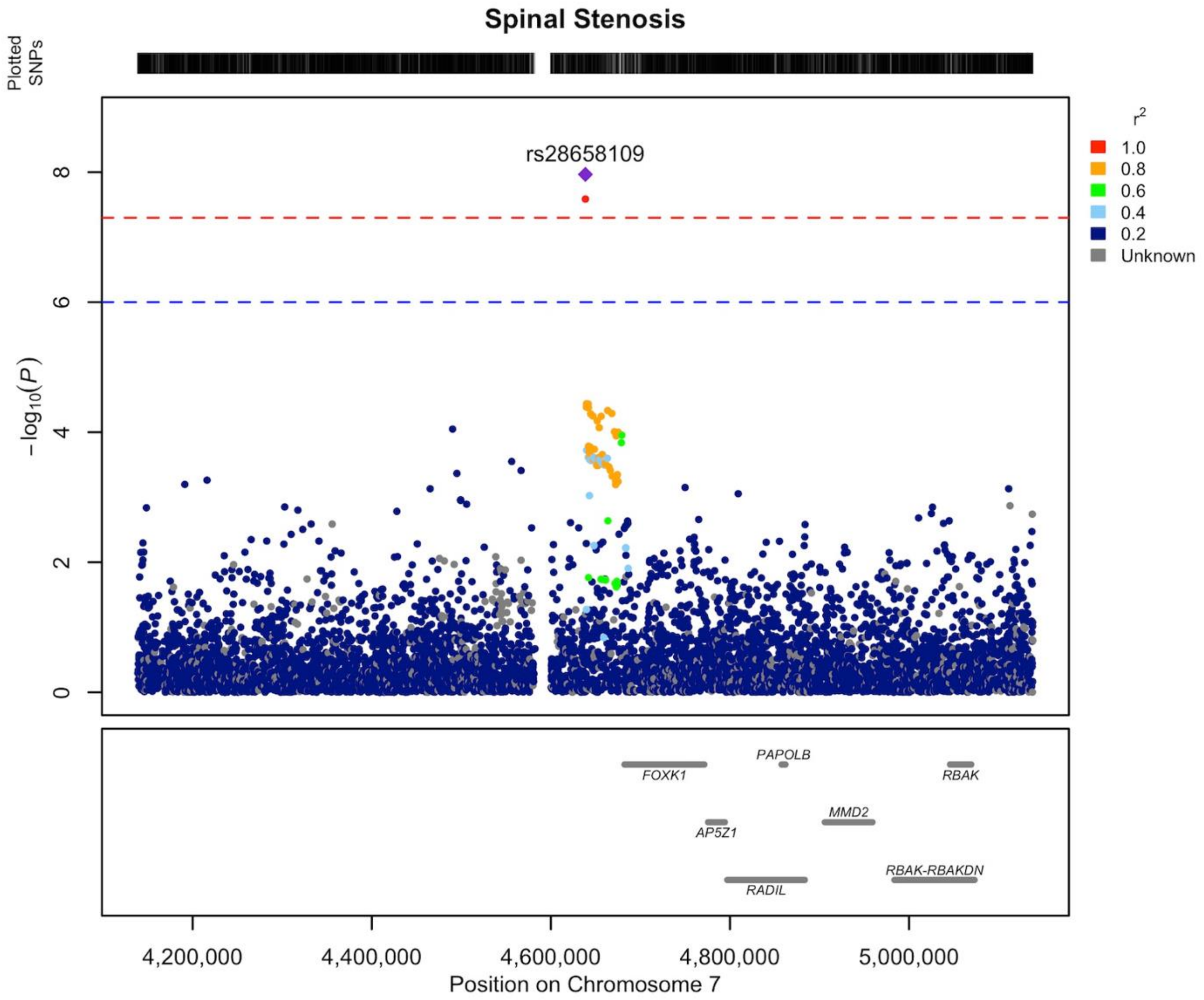


**Supplementary Data 20** Regional association plot of novel LSS association on chromosome 7 area of 4.1-5.1 MB. Our findings indicate that the gene responsible for the locus's association is most likely *FOXK1 (forkhead box K1)*. The LD structure of the plot does not fully represent the meta-analysis as it was calculated using FinnGen. The Locuszooms (https://github.com/Geeketics/LocusZooms) R package was used to generate the plot, and the Ensembl archive (https://jul2023.archive.ensembl.org) was used for the gene list.

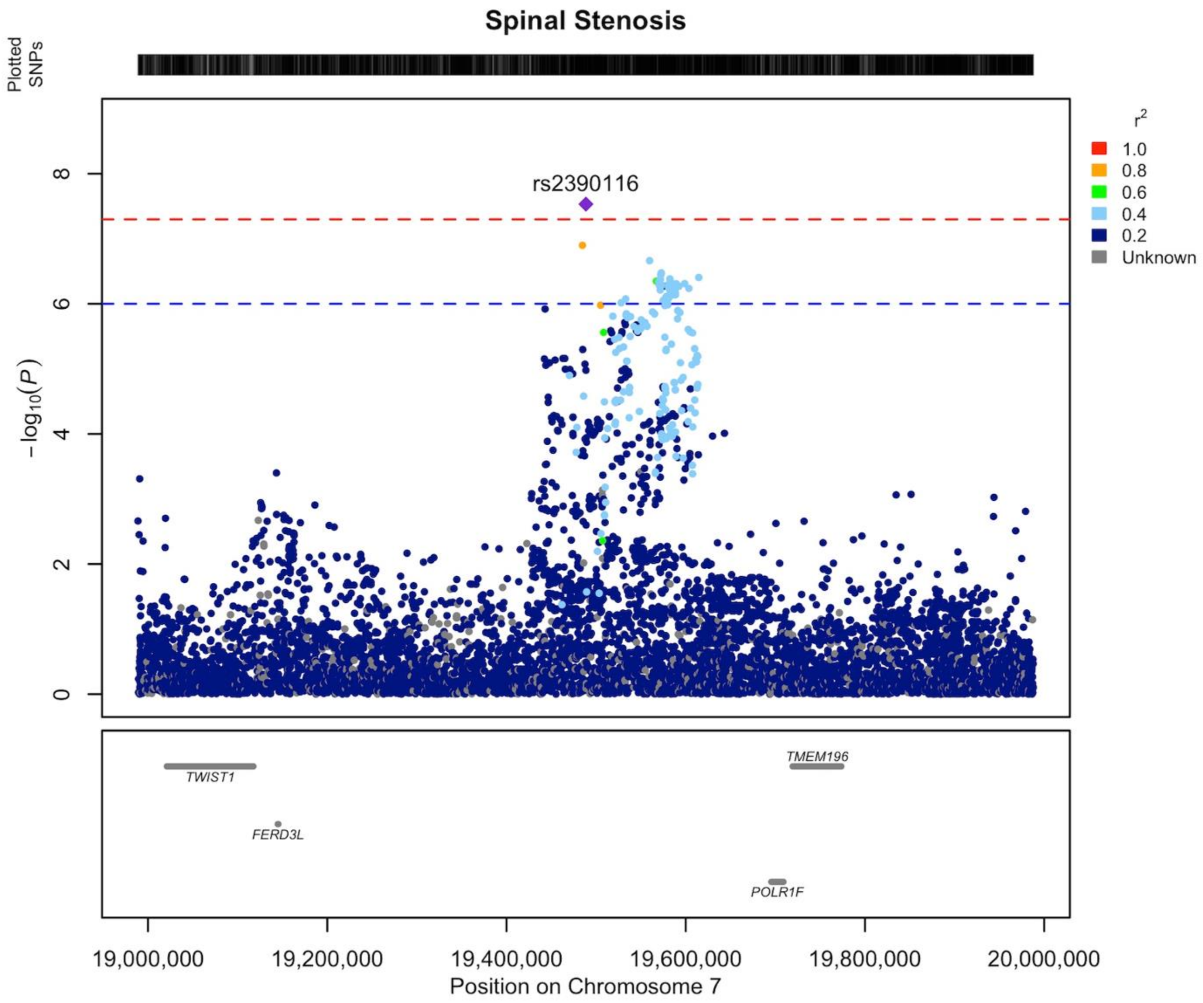


**Supplementary Data 21** Regional association plot of novel LSS association on chromosome 7 area of 18.9-19.9 MB. Our findings indicate that the gene responsible for the locus's association is most likely *TWIST1 (twist family bHLH transcription factor 1).* The LD structure of the plot does not fully represent the meta-analysis as it was calculated using FinnGen. The Locuszooms (https://github.com/Geeketics/LocusZooms) R package was used to generate the plot, and the Ensembl archive (https://jul2023.archive.ensembl.org) was used for the gene list.

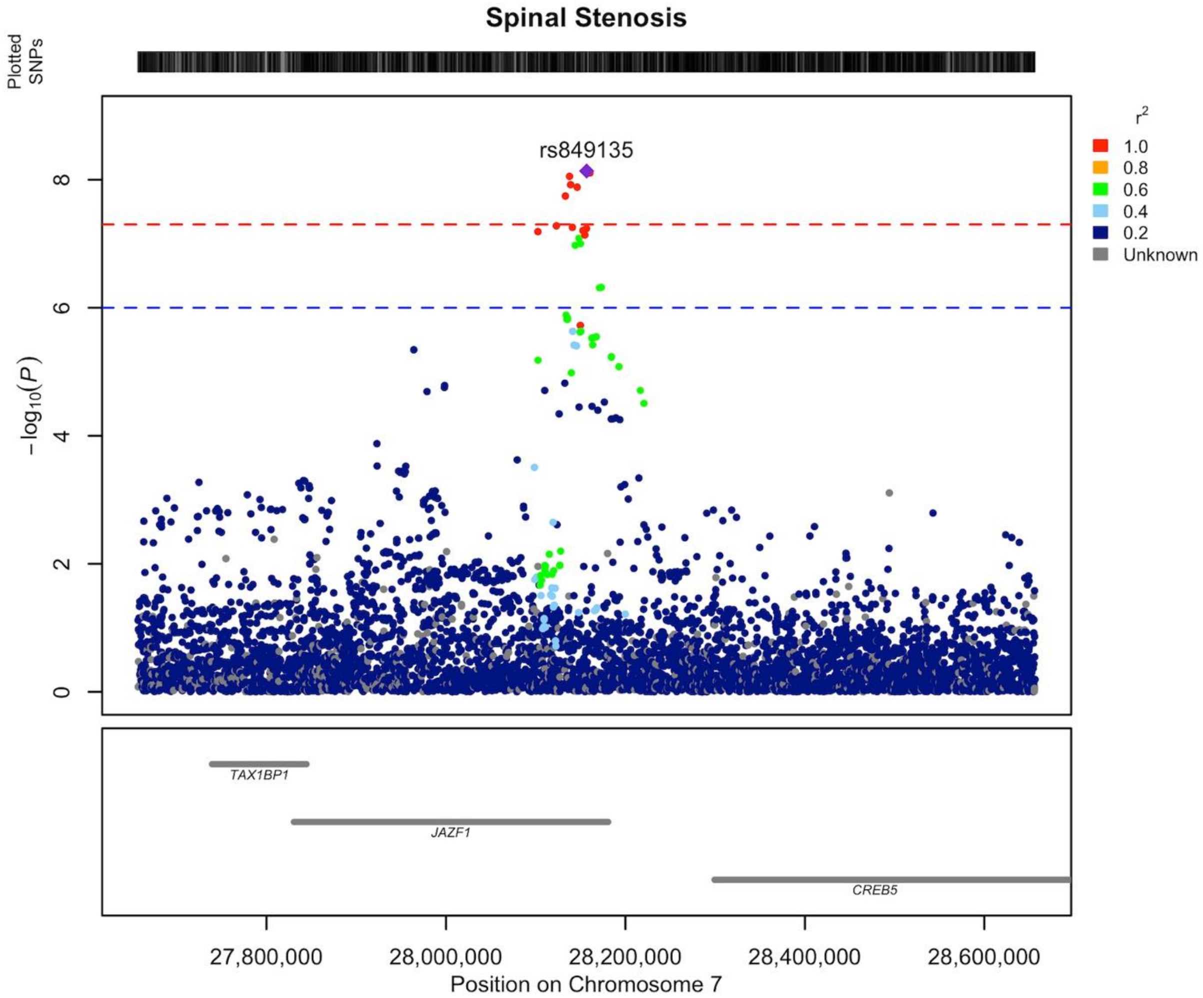


**Supplementary Data 22** Regional association plot of novel LSS association on chromosome 7 area of 27.7-28.7 MB. Our findings indicate that the gene responsible for the locus's association is most likely *JAZF1 (JAZF1 zinc finger 1)*. The LD structure of the plot does not fully represent the meta-analysis as it was calculated using FinnGen. The Locuszooms (https://github.com/Geeketics/LocusZooms) R package was used to generate the plot, and the Ensembl archive (https://jul2023.archive.ensembl.org) was used for the gene list.

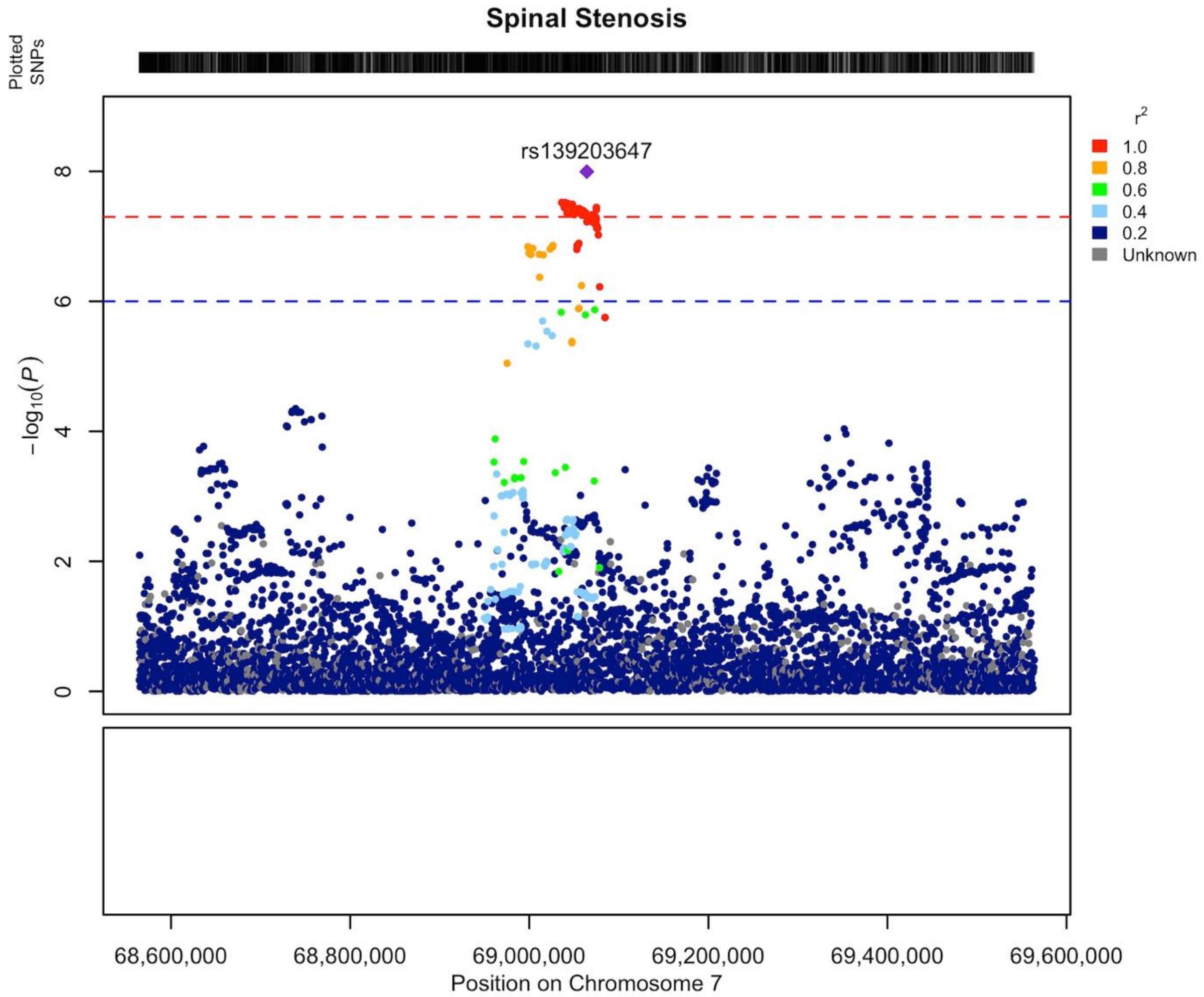


**Supplementary Data 23** Regional association plot of novel LSS association on chromosome 7 area of 68.6-69.6 MB. There was not any protein coding gene in this locus, so it was classified as *Empty*. The LD structure of the plot does not fully represent the meta-analysis as it was calculated using FinnGen. The Locuszooms (https://github.com/Geeketics/LocusZooms) R package was used to generate the plot, and the Ensembl archive (https://jul2023.archive.ensembl.org) was used for the gene list.

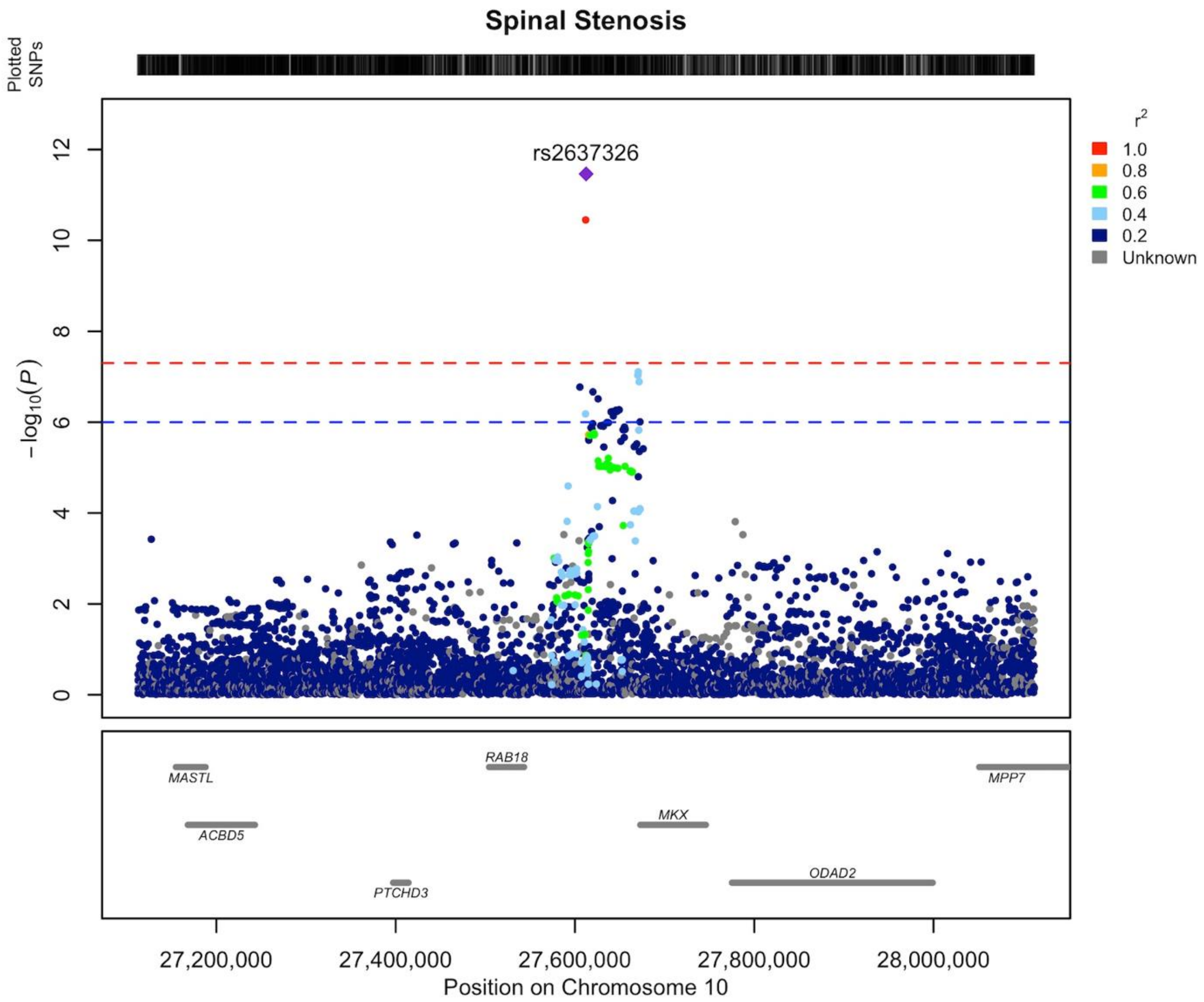


**Supplementary Data 24** Regional association plot of novel LSS association on chromosome 10 area of 27.1-28.1 MB. Our findings indicate that the gene responsible for the locus's association is most likely *MKX (mohawk homeobox)*. The LD structure of the plot does not fully represent the meta-analysis as it was calculated using FinnGen. The Locuszooms (https://github.com/Geeketics/LocusZooms) R package was used to generate the plot, and the Ensembl archive (https://jul2023.archive.ensembl.org) was used for the gene list.

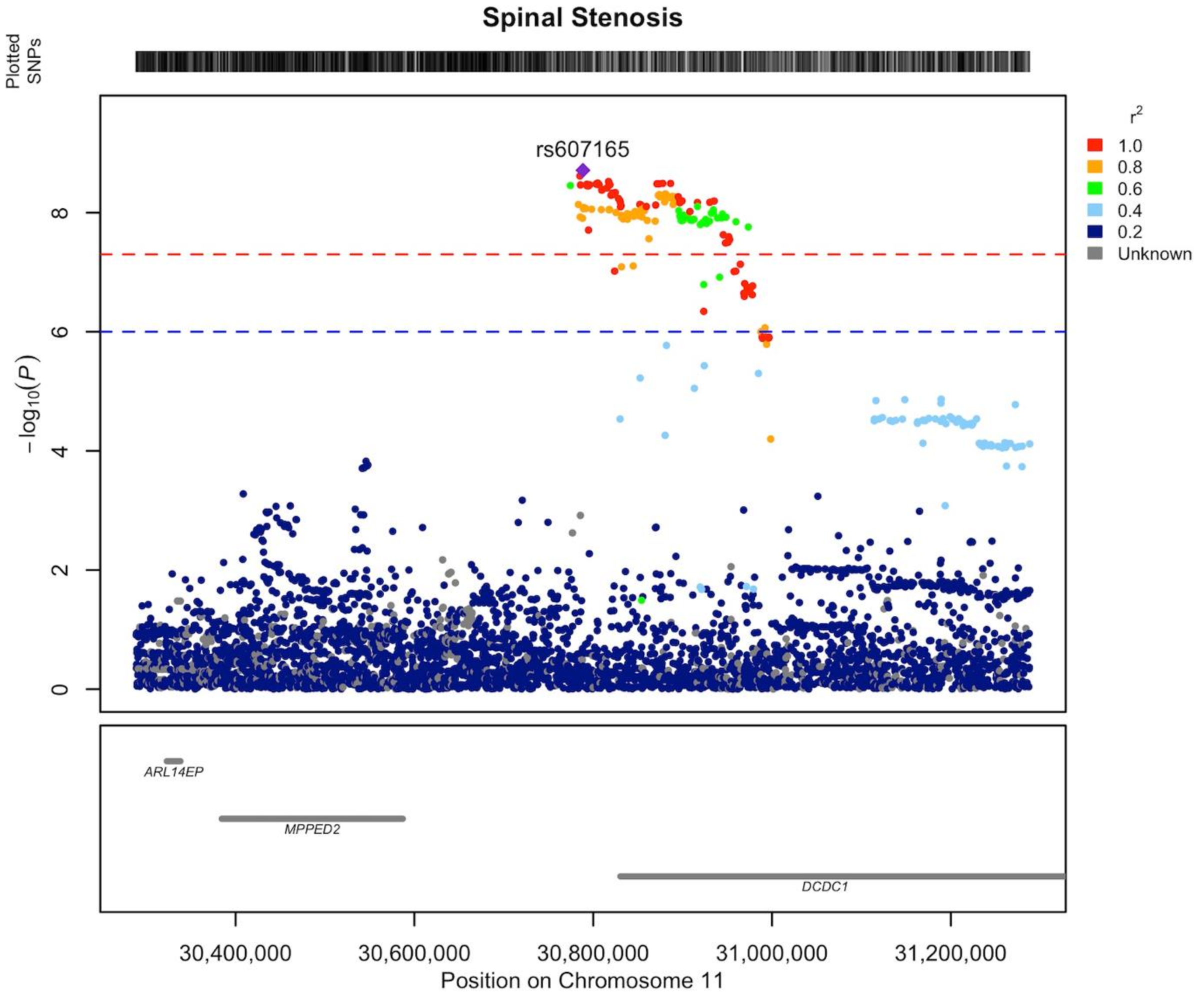


**Supplementary Data 25** Regional association plot of novel LSS association on chromosome 11 area of 30.3-31.3 MB. Our findings indicate that the gene responsible for the locus's association is most likely *MPPED2 (metallophosphosterase domain containing 2).* The LD structure of the plot does not fully represent the meta-analysis as it was calculated using FinnGen. The Locuszooms (https://github.com/Geeketics/LocusZooms) R package was used to generate the plot, and the Ensembl archive (https://jul2023.archive.ensembl.org) was used for the gene list.

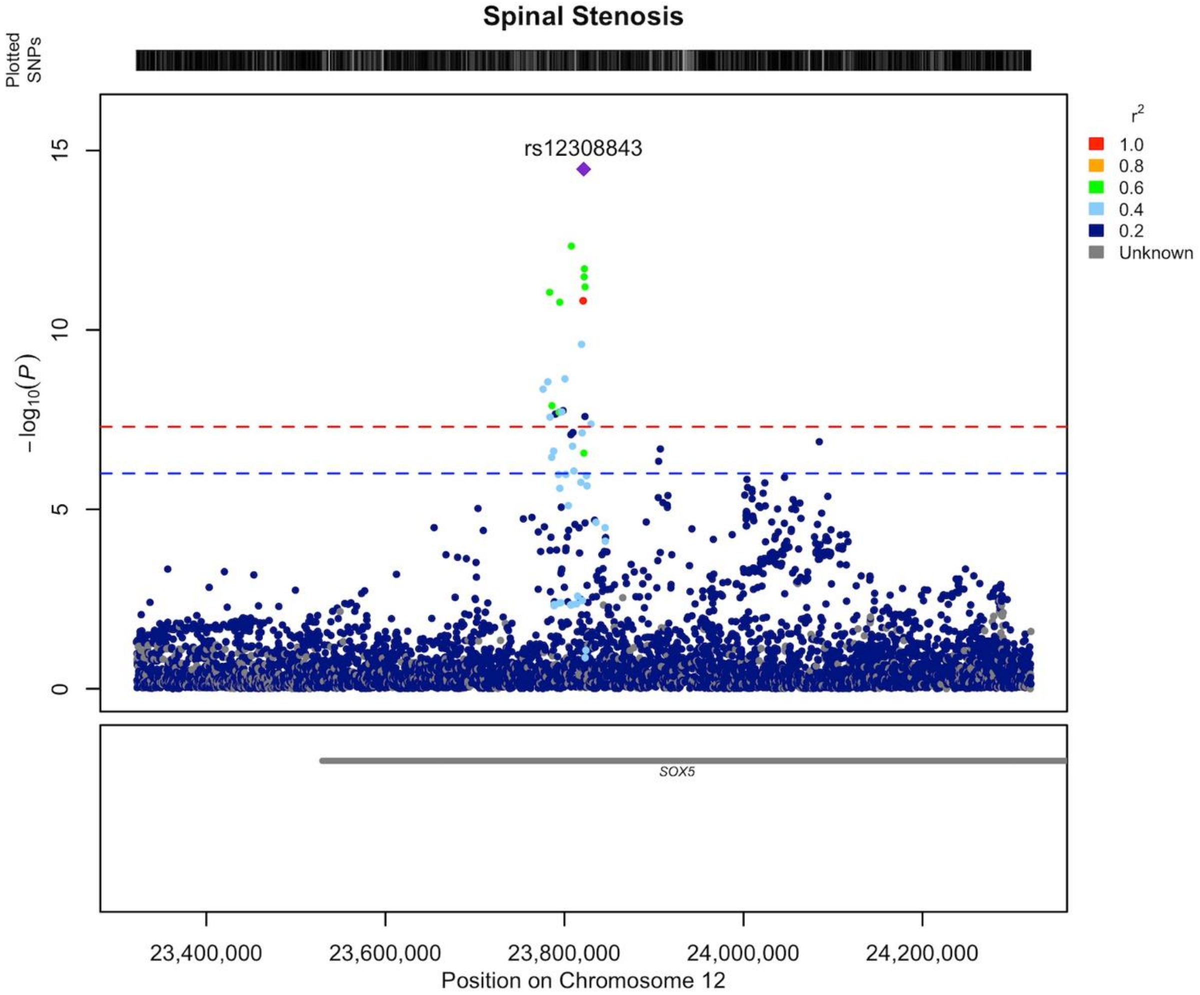


**Supplementary Data 26** Regional association plot of novel LSS association on chromosome 12 area of 23.3-24.3 MB. Our findings indicate that the gene responsible for the locus's association is most likely *SOX5 (SRY-box transcription factor 5).* The LD structure of the plot does not fully represent the meta-analysis as it was calculated using FinnGen. The Locuszooms (https://github.com/Geeketics/LocusZooms) R package was used to generate the plot, and the Ensembl archive (https://jul2023.archive.ensembl.org) was used for the gene list.

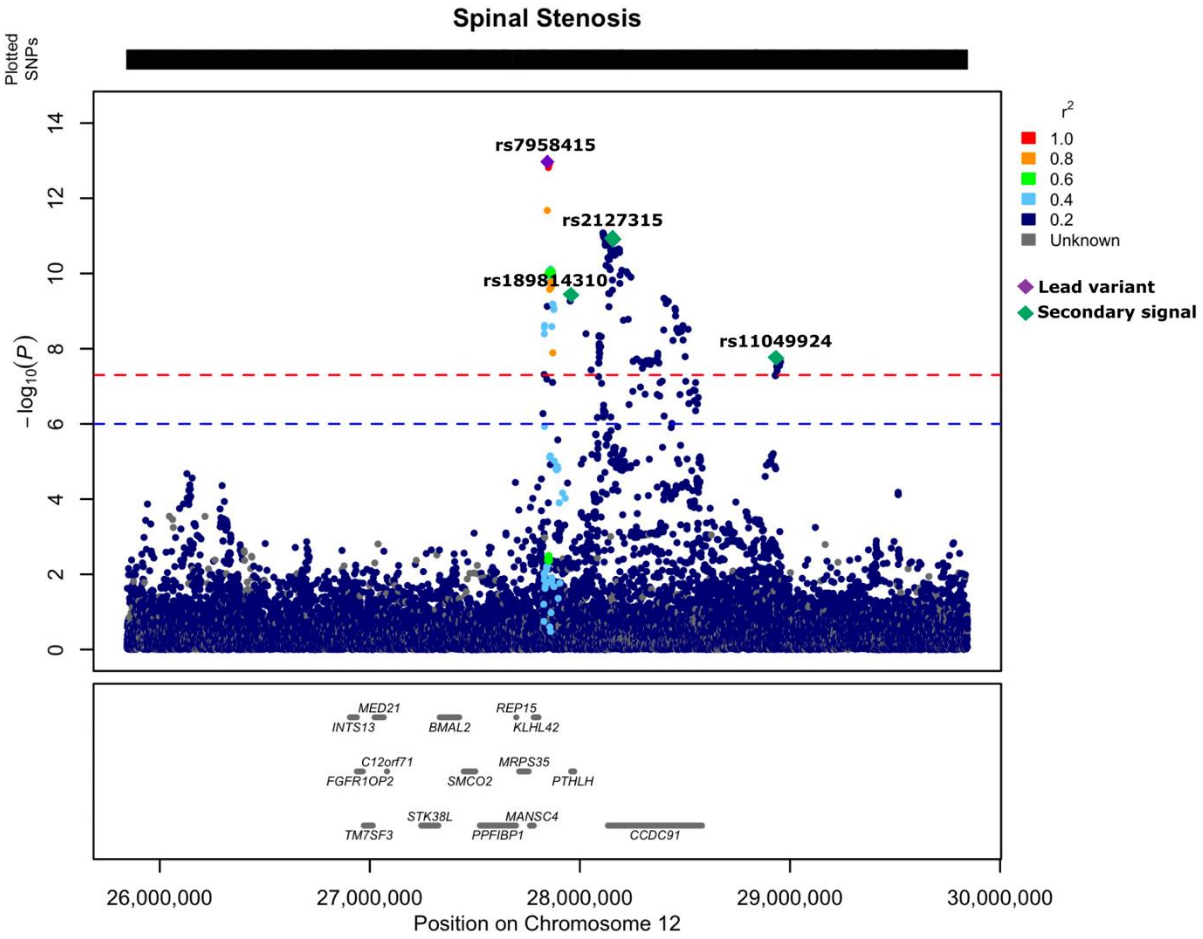


**Supplementary Data 27** Regional association plot of novel LSS associations on chromosome 12 area of 25.8-29.8 MB. Our findings indicate that the gene responsible for the locus's association is most likely *CCDC91* (*coiled-coil domain containing 91*). Lead variant is highlighted in purple and secondary signals observed in the conditional analysis are marked in green. More information about secondary signals in Table S2. The LD structure of the plot does not fully represent the meta-analysis as it was calculated using FinnGen. The Locuszooms (https://github.com/Geeketics/LocusZooms) R package was used to generate the plot, and the Ensembl archive (https://jul2023.archive.ensembl.org) was used for the gene list.

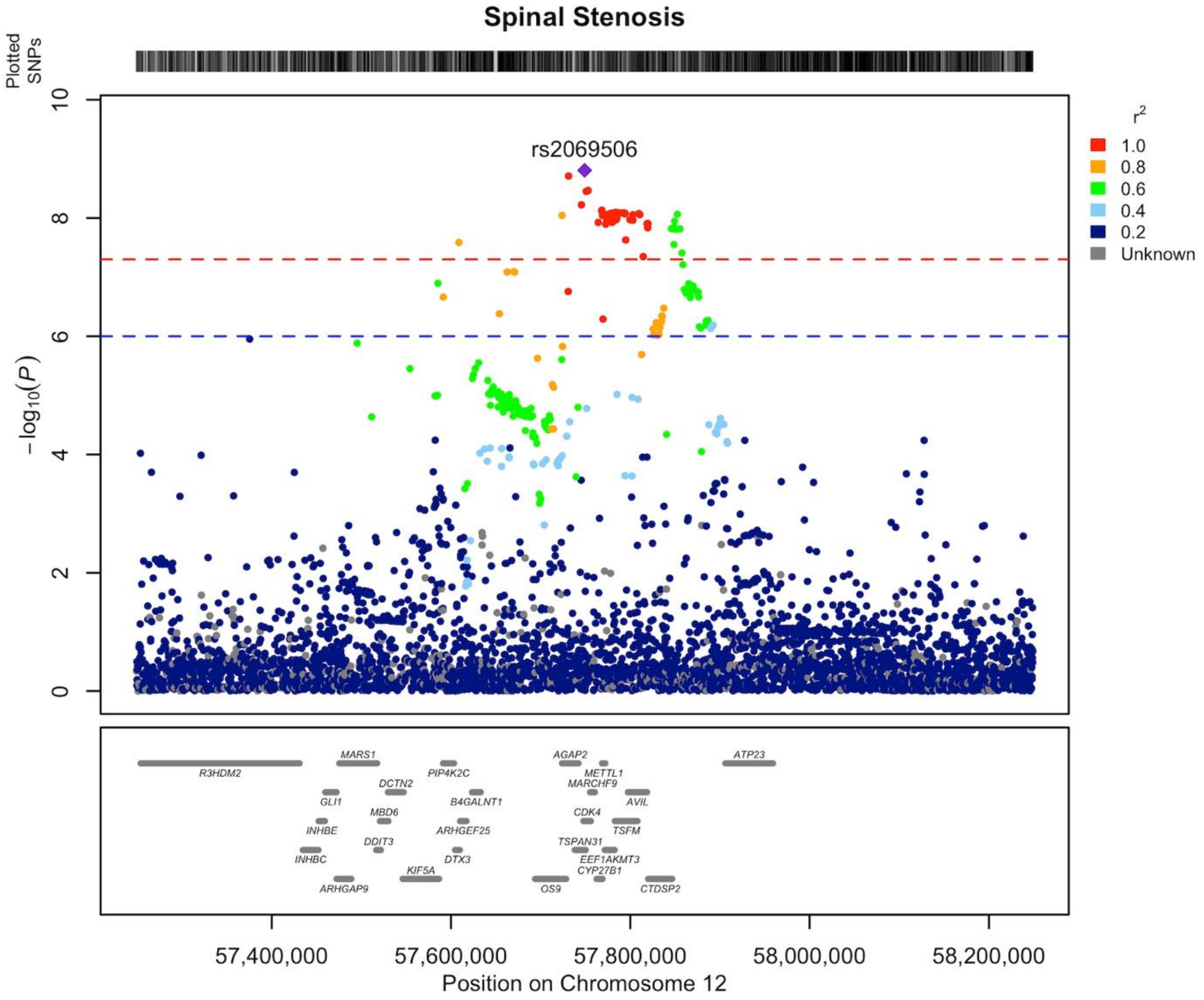


**Supplementary Data 28** Regional association plot of novel LSS association on chromosome 12 area of 57.2-58.2 MB. Our findings indicate that the gene responsible for the locus's association is most likely *GLI1 (GLI family zinc finger 1).* The LD structure of the plot does not fully represent the meta-analysis as it was calculated using FinnGen. The Locuszooms (https://github.com/Geeketics/LocusZooms) R package was used to generate the plot, and the Ensembl archive (https://jul2023.archive.ensembl.org) was used for the gene list.

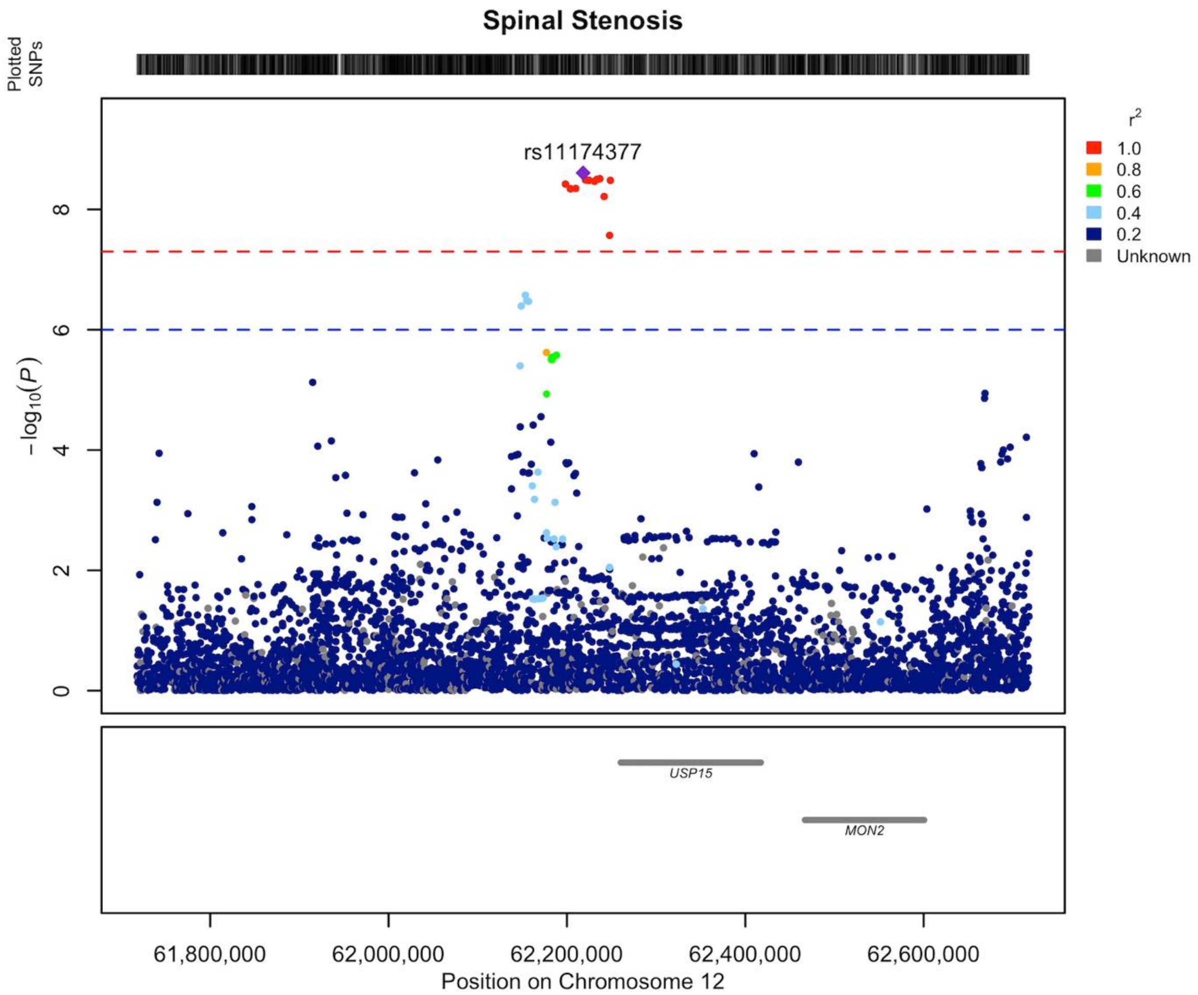


**Supplementary Data 29** Regional association plot of novel LSS association on chromosome 12 area of 61.7-62.7 MB. Our findings indicate that the gene responsible for the locus's association is most likely *USP15 (ubiquitin specific peptidase 15).* The LD structure of the plot does not fully represent the meta-analysis as it was calculated using FinnGen. The Locuszooms (https://github.com/Geeketics/LocusZooms) R package was used to generate the plot, and the Ensembl archive (https://jul2023.archive.ensembl.org) was used for the gene list.

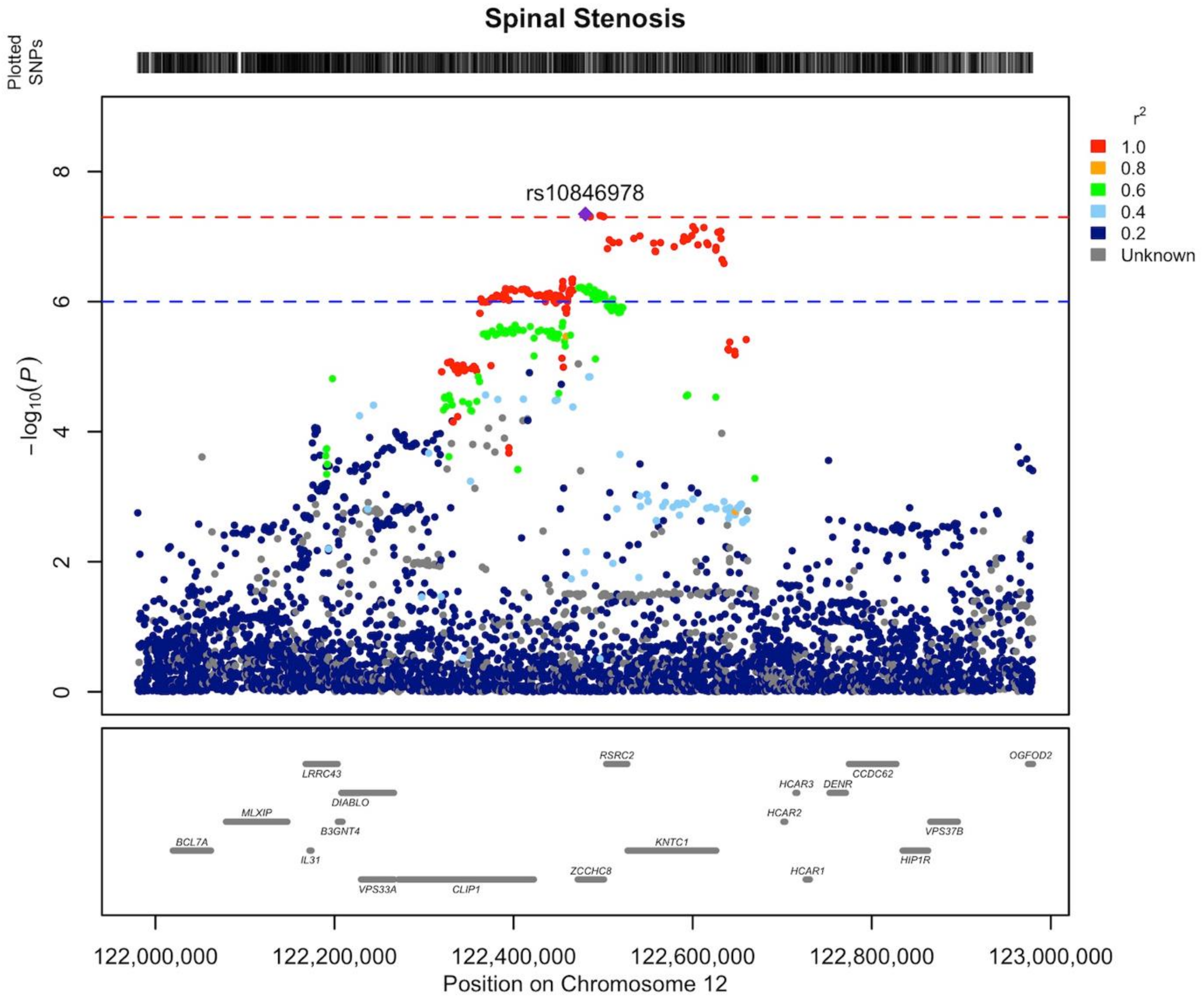


**Supplementary Data 30** Regional association plot of novel LSS association on chromosome 12 area of 121.9-122.9 MB. Our findings indicate that the gene responsible for the locus's association is most likely *ZCCHC8 (zinc finger CCHC-type containing 8).* The LD structure of the plot does not fully represent the meta-analysis as it was calculated using FinnGen. The Locuszooms (https://github.com/Geeketics/LocusZooms) R package was used to generate the plot, and the Ensembl archive (https://jul2023.archive.ensembl.org) was used for the gene list.

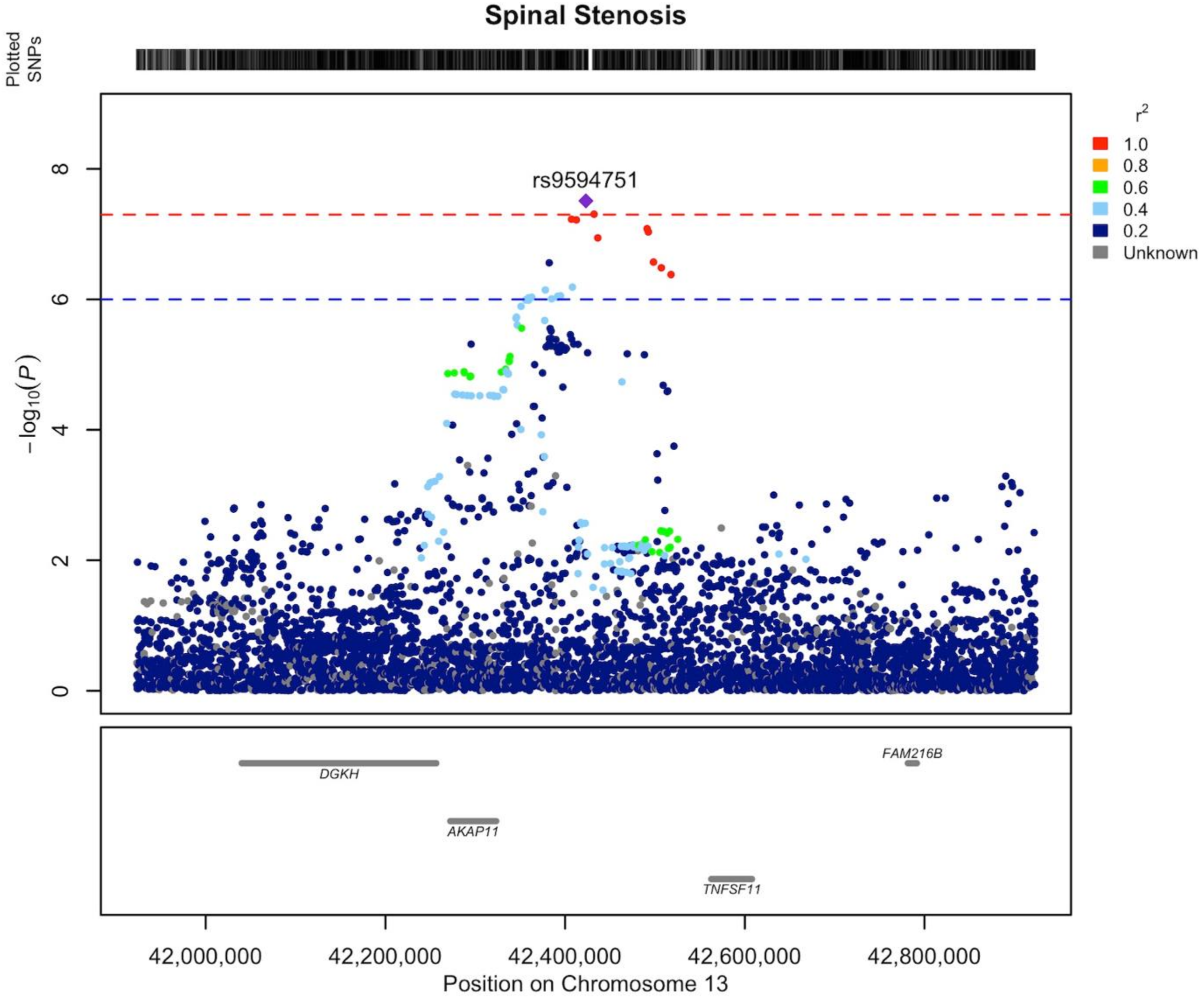


**Supplementary Data 31** Regional association plot of novel LSS association on chromosome 13 area of 41.9-42.9 MB. Our findings indicate that the gene responsible for the locus's association is most likely *TNFSF11 (TNF superfamily member 11)*. The LD structure of the plot does not fully represent the meta-analysis as it was calculated using FinnGen. The Locuszooms (https://github.com/Geeketics/LocusZooms) R package was used to generate the plot, and the Ensembl archive (https://jul2023.archive.ensembl.org) was used for the gene list.

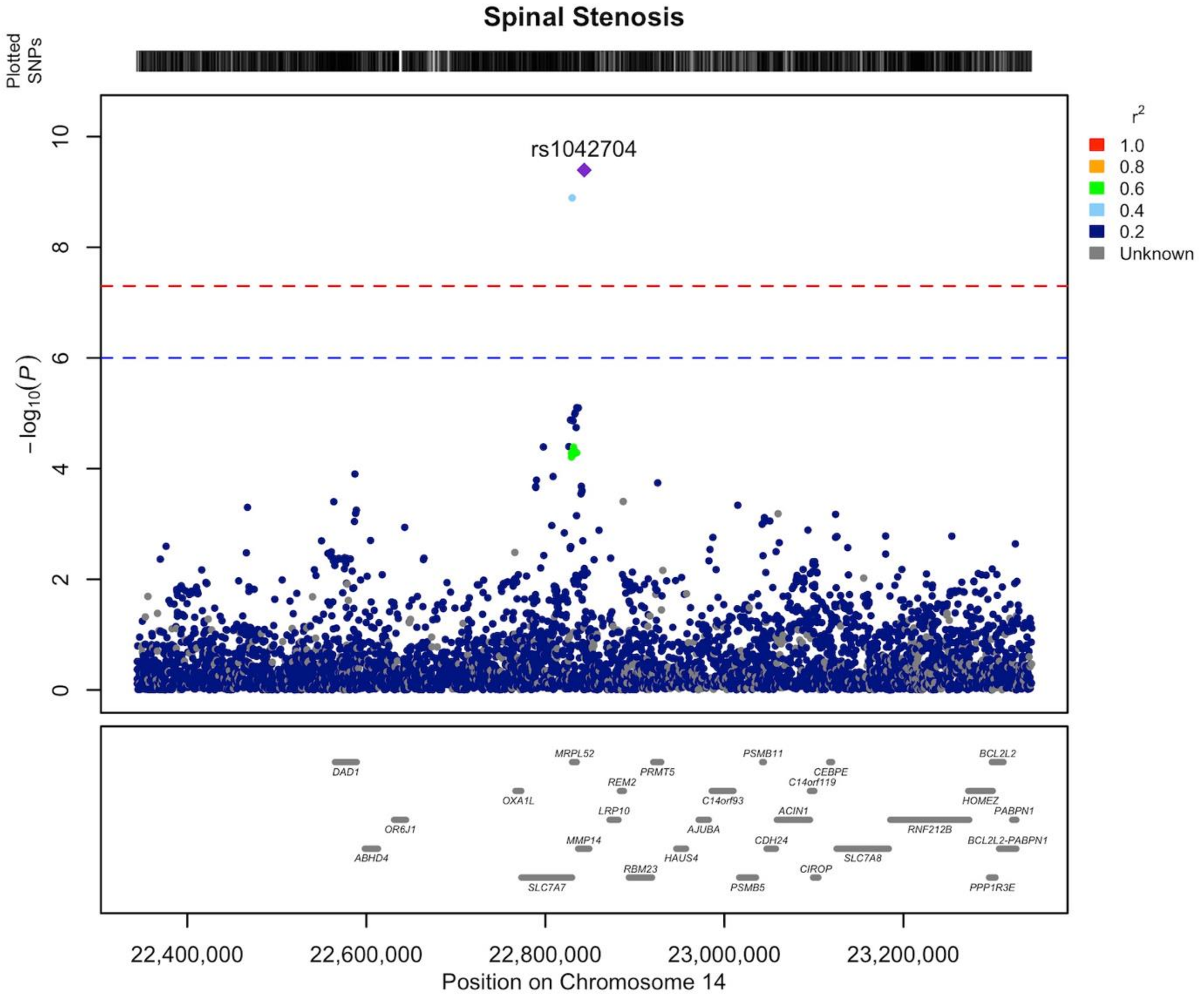


**Supplementary Data 32** Regional association plot of novel LSS association on chromosome 14 area of 22.3-23.3 MB. Our findings indicate that the gene responsible for the locus's association is most likely *MMP14 (matrix metallopeptidase 14)*. The LD structure of the plot does not fully represent the meta-analysis as it was calculated using FinnGen. The Locuszooms (https://github.com/Geeketics/LocusZooms) R package was used to generate the plot, and the Ensembl archive (https://jul2023.archive.ensembl.org) was used for the gene list.

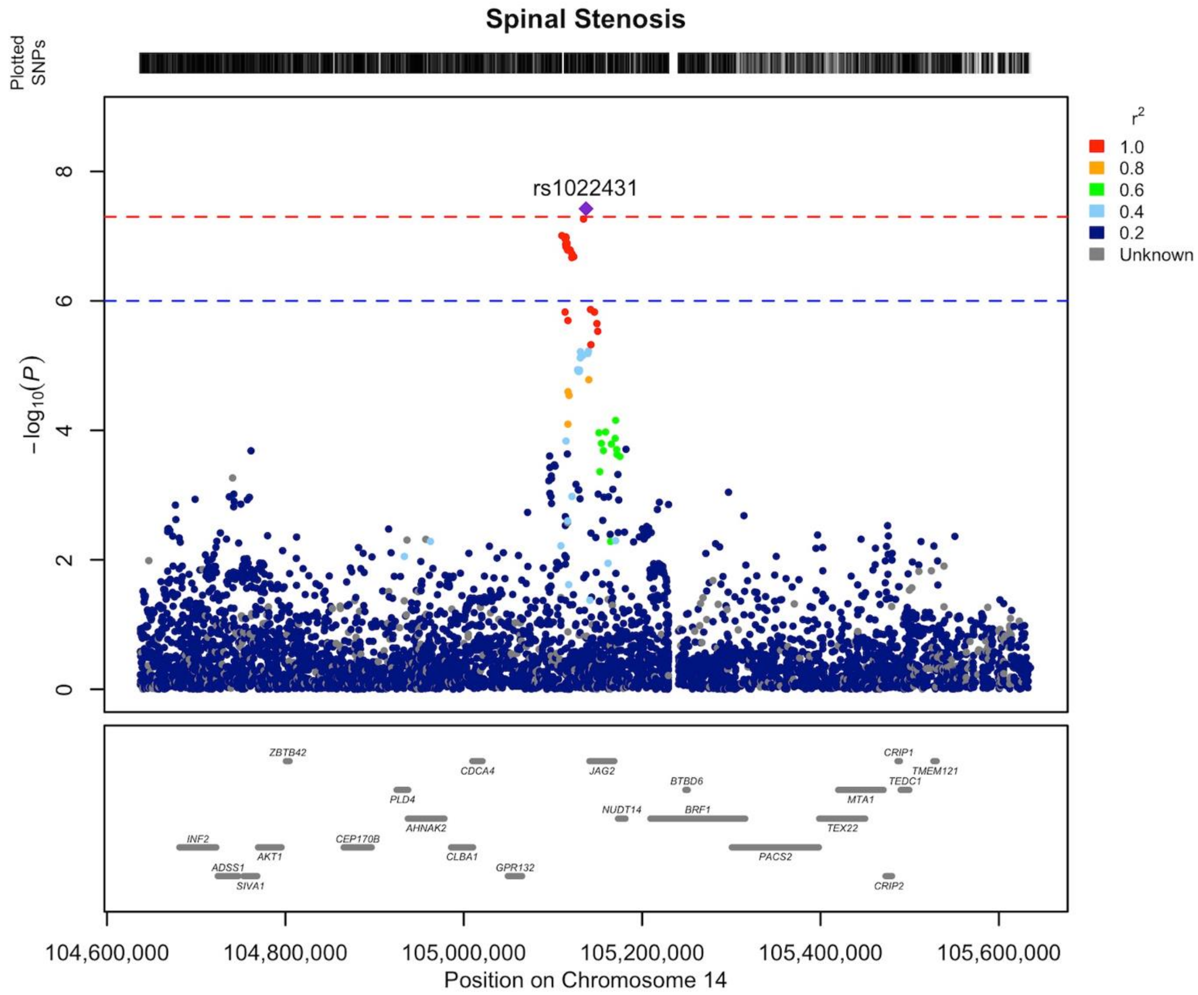


**Supplementary Data 33** Regional association plot of novel LSS association on chromosome 14 area of 104.6-105.6 MB. Our findings indicate that the gene responsible for the locus's association is most likely *GPR132 (G protein-coupled receptor 132).* The LD structure of the plot does not fully represent the meta-analysis as it was calculated using FinnGen. The Locuszooms (https://github.com/Geeketics/LocusZooms) R package was used to generate the plot, and the Ensembl archive (https://jul2023.archive.ensembl.org) was used for the gene list.

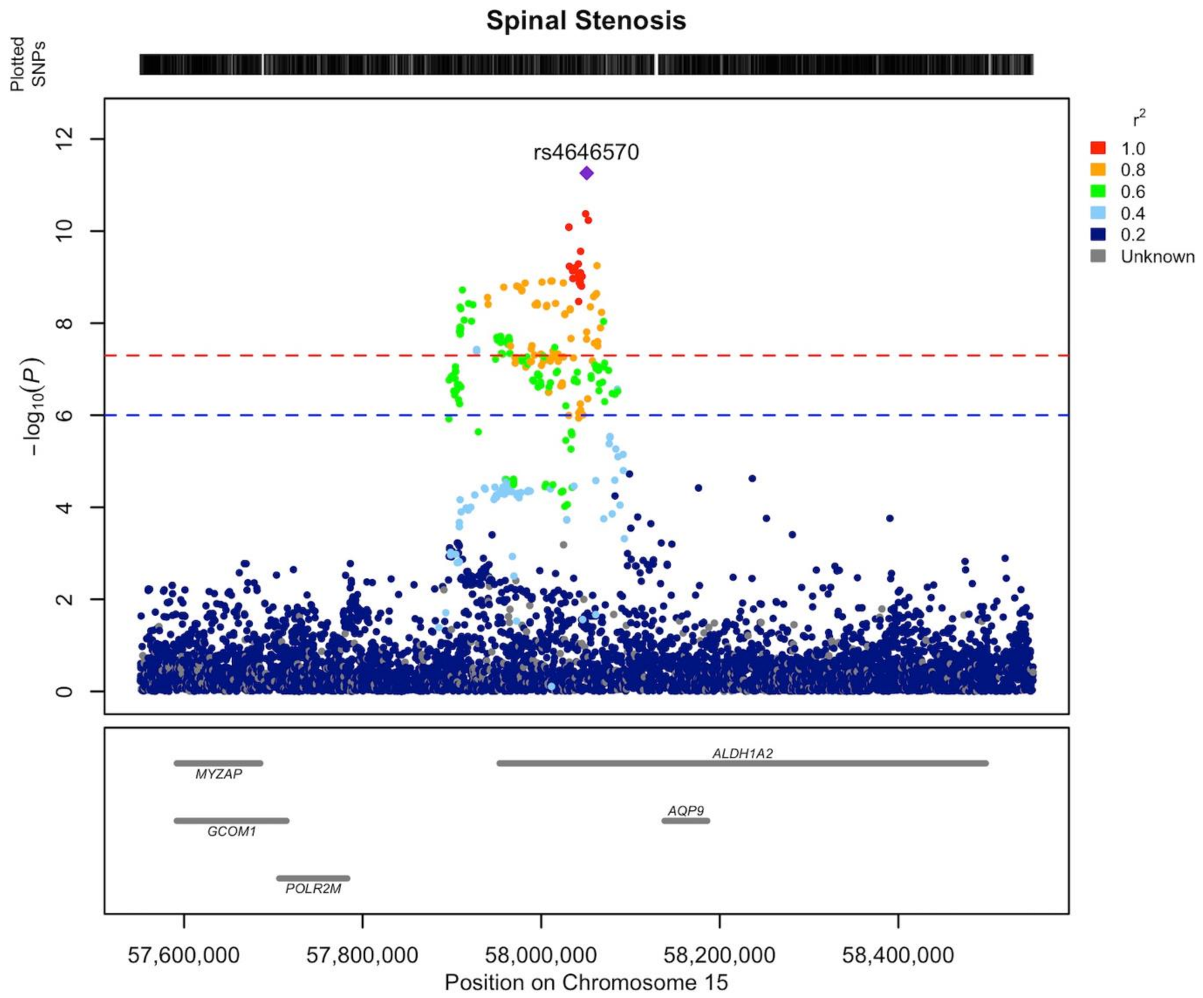


**Supplementary Data 34** Regional association plot of novel LSS association on chromosome 15 area of 57.6-58.6 MB. Our findings indicate that the gene responsible for the locus's association is most likely *ALDH1A2 (aldehyde dehydrogenase 1 family member A2).* The LD structure of the plot does not fully represent the meta-analysis as it was calculated using FinnGen. The Locuszooms (https://github.com/Geeketics/LocusZooms) R package was used to generate the plot, and the Ensembl archive (https://jul2023.archive.ensembl.org) was used for the gene list.

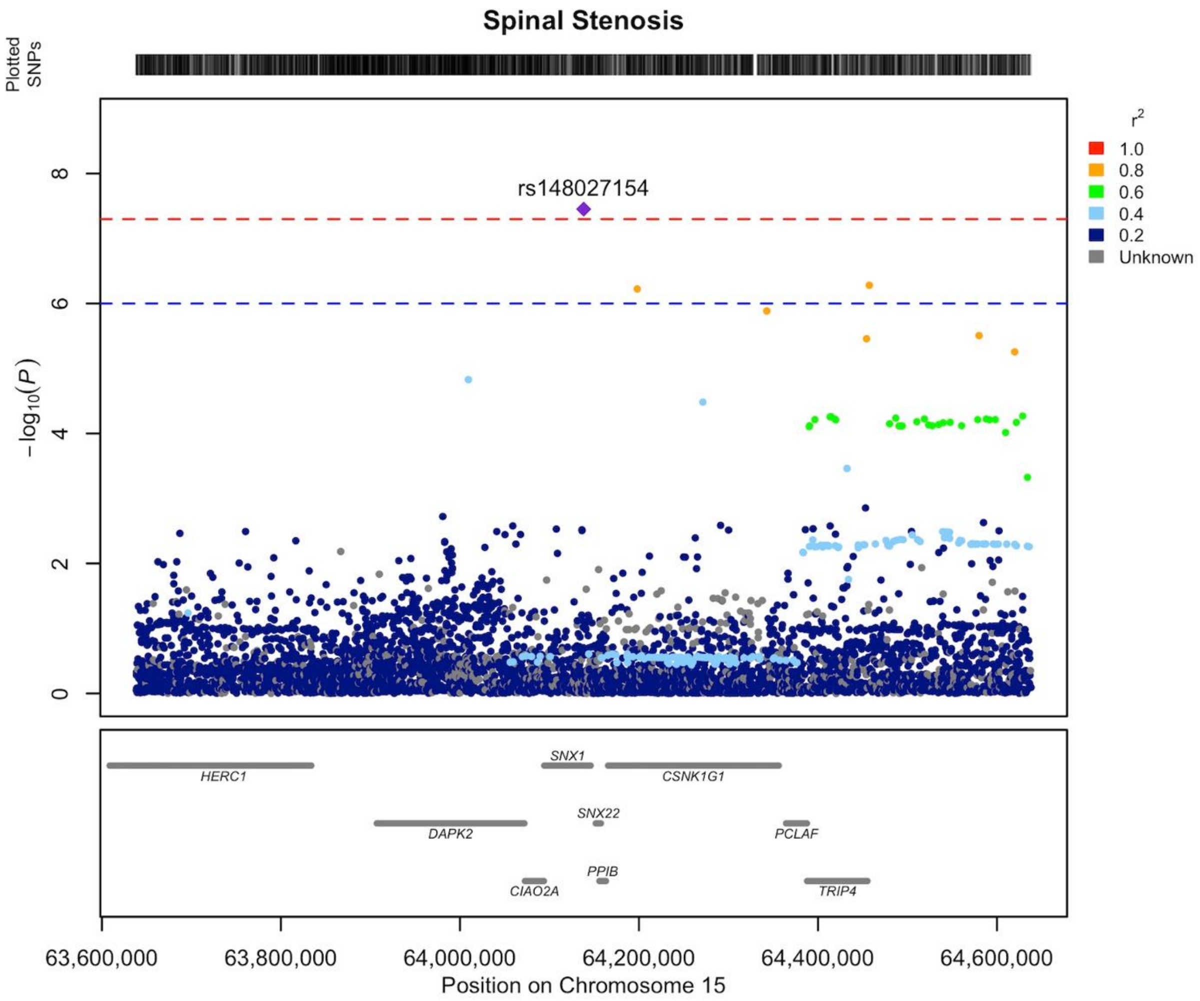


**Supplementary Data 35** Regional association plot of novel LSS association on chromosome 15 area of 63.6-64.6 MB. Our findings indicate that the gene responsible for the locus's association is most likely *PPIB (peptidylprolyl isomerase B).* The LD structure of the plot does not fully represent the meta-analysis as it was calculated using FinnGen. The Locuszooms (https://github.com/Geeketics/LocusZooms) R package was used to generate the plot, and the Ensembl archive (https://jul2023.archive.ensembl.org) was used for the gene list.

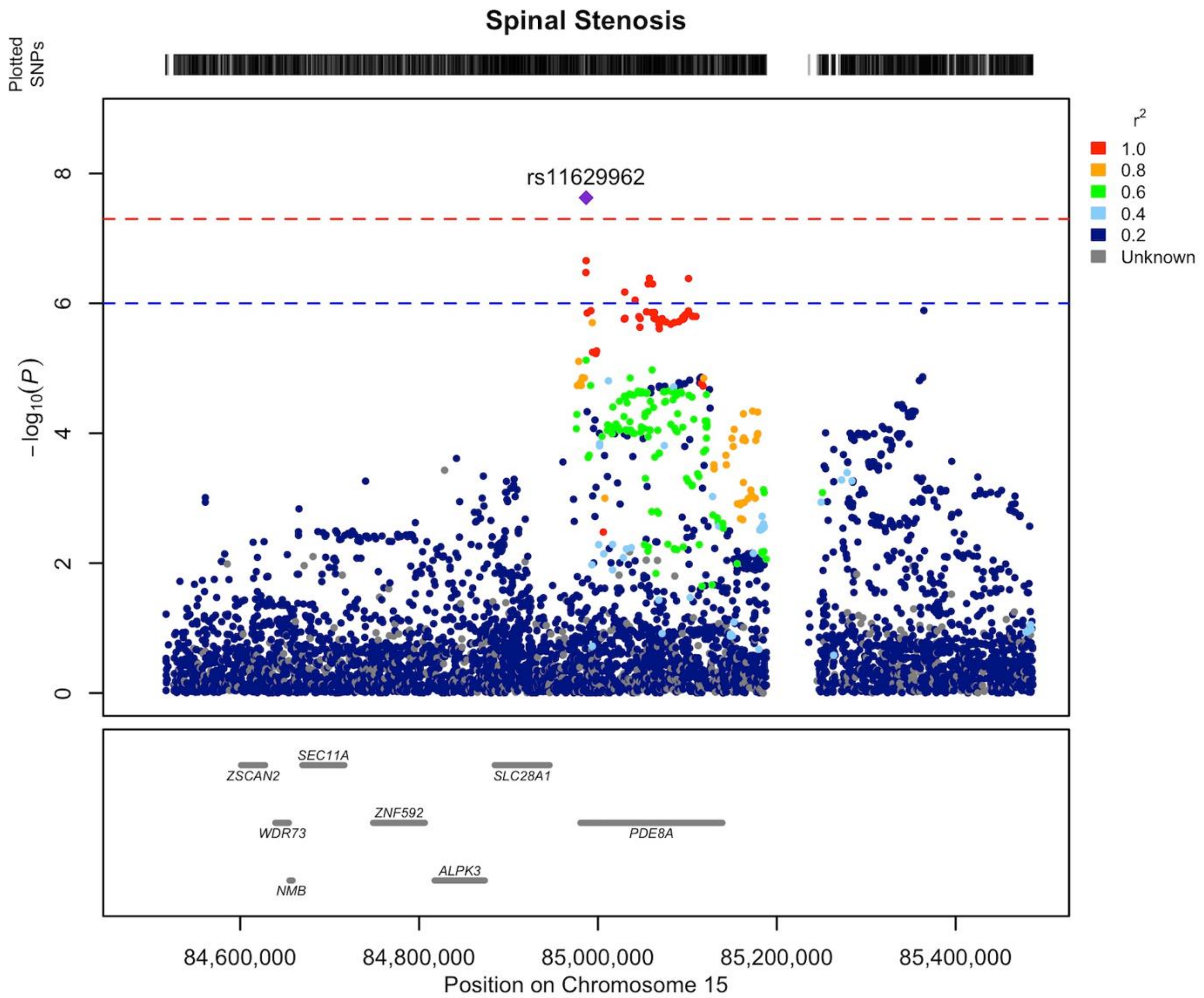


**Supplementary Data 36** Regional association plot of novel LSS association on chromosome 15 area of 84.5-85.5 MB. Our findings indicate that the gene responsible for the locus's association is most likely *PDE8A (phosphodiesterase 8A)*. The LD structure of the plot does not fully represent the meta-analysis as it was calculated using FinnGen. The Locuszooms (https://github.com/Geeketics/LocusZooms) R package was used to generate the plot, and the Ensembl archive (https://jul2023.archive.ensembl.org) was used for the gene list.

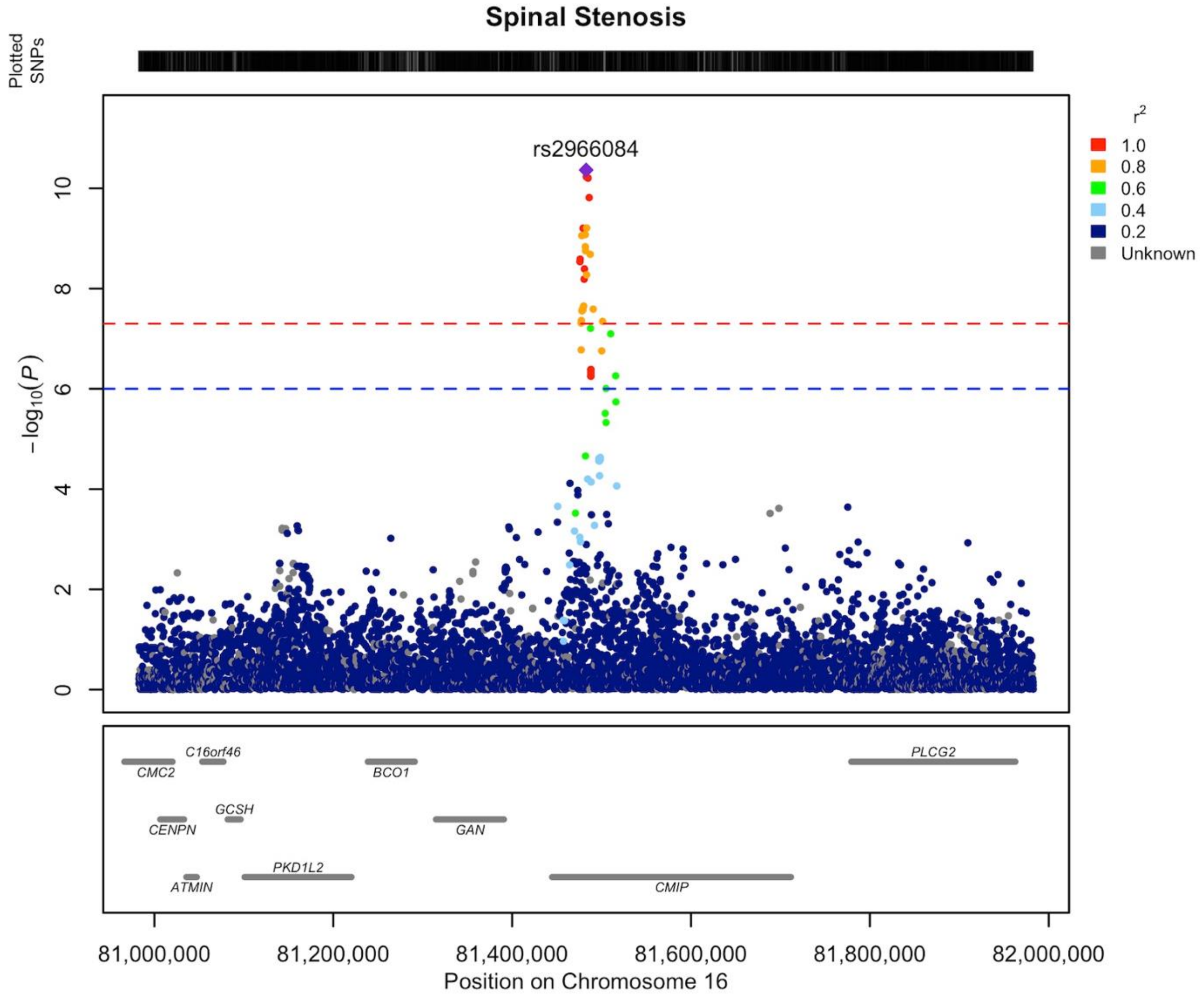


**Supplementary Data 37** Regional association plot of novel LSS association on chromosome 16 area of 80.9-81.9 MB. Our findings indicate that the gene responsible for the locus's association is most likely *GAN (gigaxonin).* The LD structure of the plot does not fully represent the meta-analysis as it was calculated using FinnGen. The Locuszooms (https://github.com/Geeketics/LocusZooms) R package was used to generate the plot, and the Ensembl archive (https://jul2023.archive.ensembl.org) was used for the gene list.

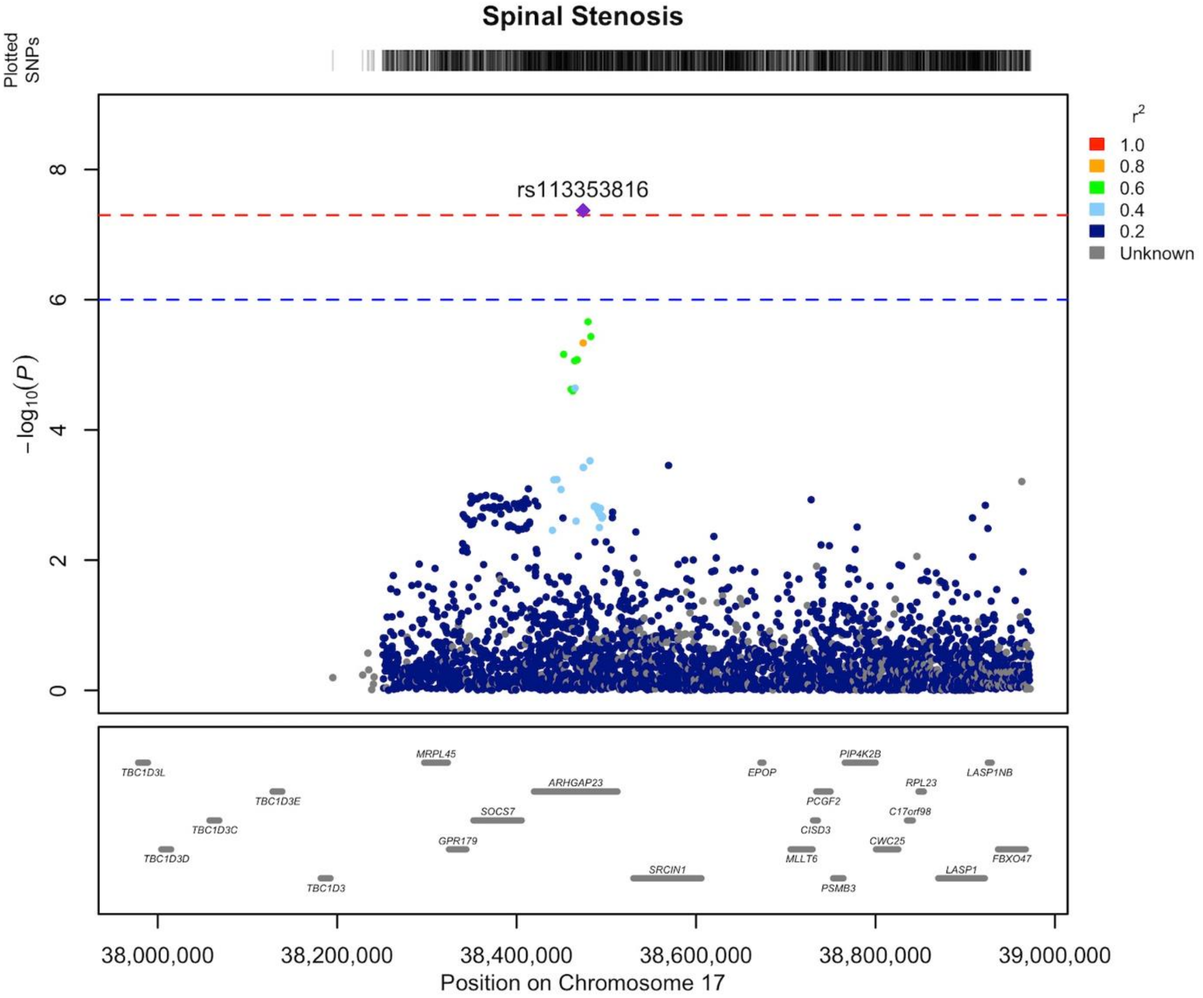


**Supplementary Data 38** Regional association plot of novel LSS association on chromosome 17 area of 37.9-38.9 MB. Our findings indicate that the gene responsible for the locus's association is most likely *SOCS7 (suppressor of cytokine signaling 7)*. The LD structure of the plot does not fully represent the meta-analysis as it was calculated using FinnGen. The Locuszooms (https://github.com/Geeketics/LocusZooms) R package was used to generate the plot, and the Ensembl archive (https://jul2023.archive.ensembl.org) was used for the gene list.

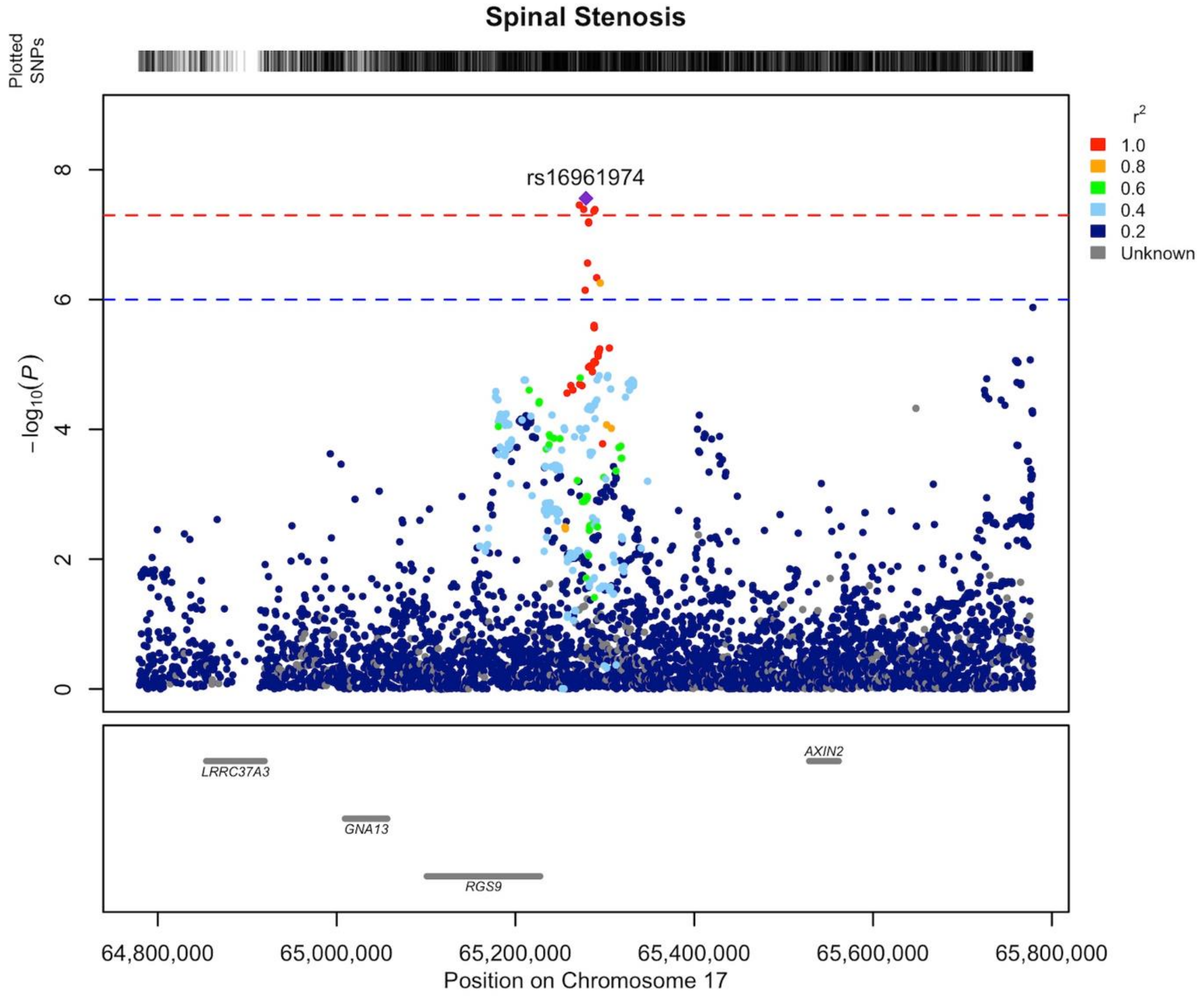


**Supplementary Data 39** Regional association plot of novel LSS association on chromosome 17 area of 64.8-65.8 MB. Our findings indicate that the gene responsible for the locus's association is most likely *AXIN2 (axin 2).* The LD structure of the plot does not fully represent the meta-analysis as it was calculated using FinnGen. The Locuszooms (https://github.com/Geeketics/LocusZooms) R package was used to generate the plot, and the Ensembl archive (https://jul2023.archive.ensembl.org) was used for the gene list.

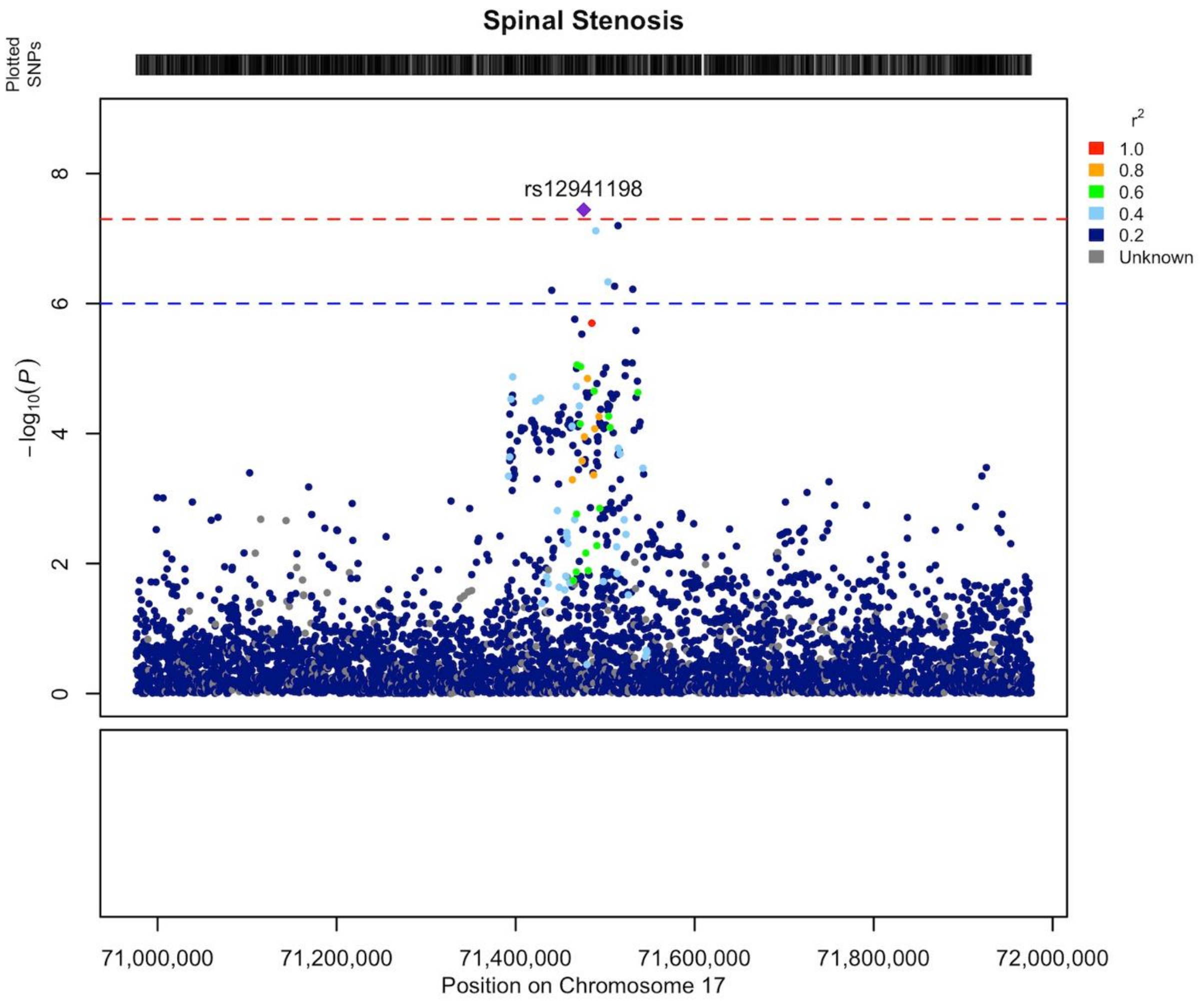


**Supplementary Data 40** Regional association plot of novel LSS association on chromosome 17 area of 70.9-71.9 MB. There was not any protein coding gene in this locus, so it was classified as Empty. The LD structure of the plot does not fully represent the meta-analysis as it was calculated using FinnGen. The Locuszooms (https://github.com/Geeketics/LocusZooms) R package was used to generate the plot, and the Ensembl archive (https://jul2023.archive.ensembl.org) was used for the gene list.

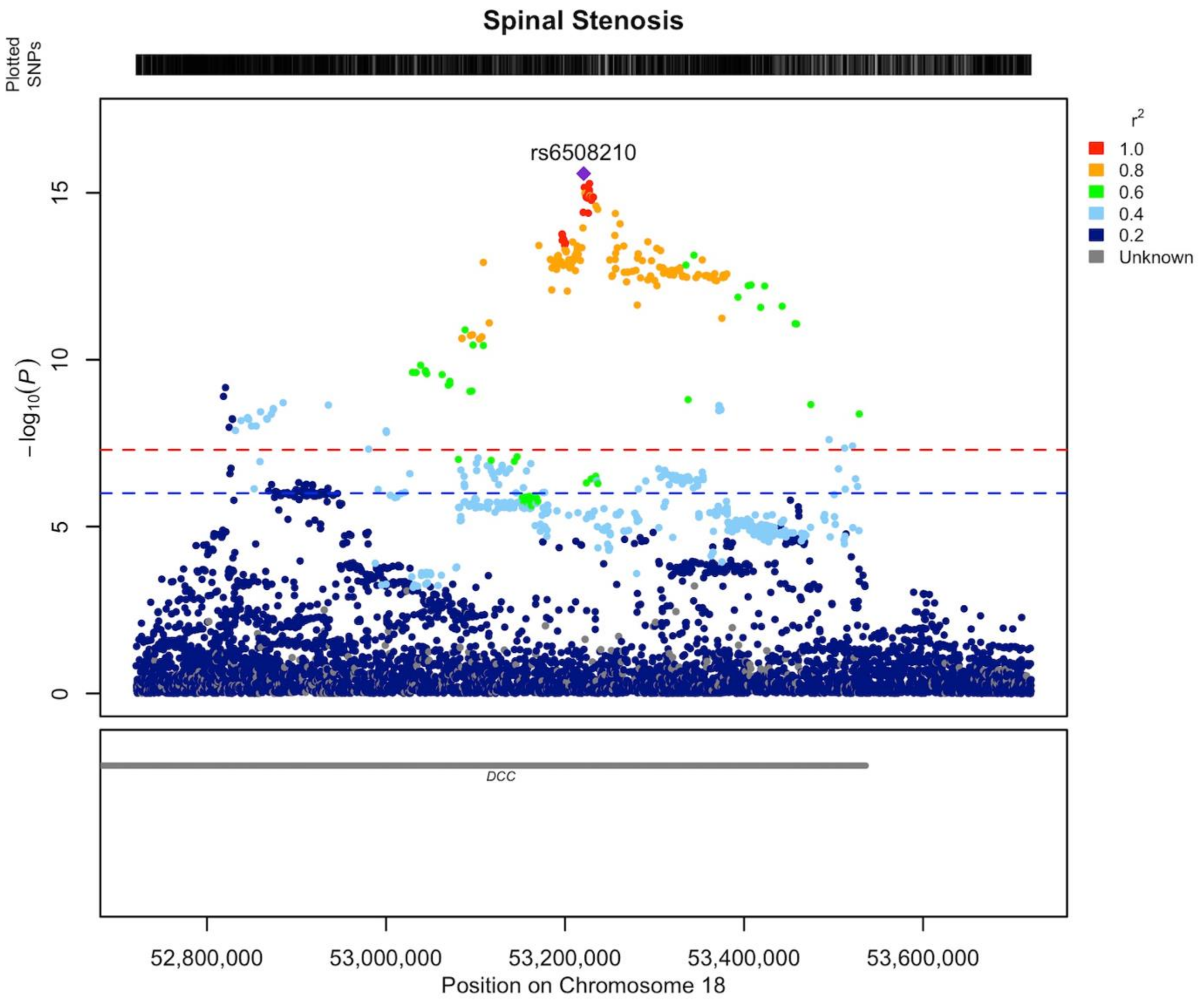


**Supplementary Data 41** Regional association plot of novel LSS association on chromosome 18 area of 52.7-53.7 MB. Our findings indicate that the gene responsible for the locus's association is most likely *DCC (DCC netrin 1 receptor)*. The LD structure of the plot does not fully represent the meta-analysis as it was calculated using FinnGen. The Locuszooms (https://github.com/Geeketics/LocusZooms) R package was used to generate the plot, and the Ensembl archive (https://jul2023.archive.ensembl.org) was used for the gene list.

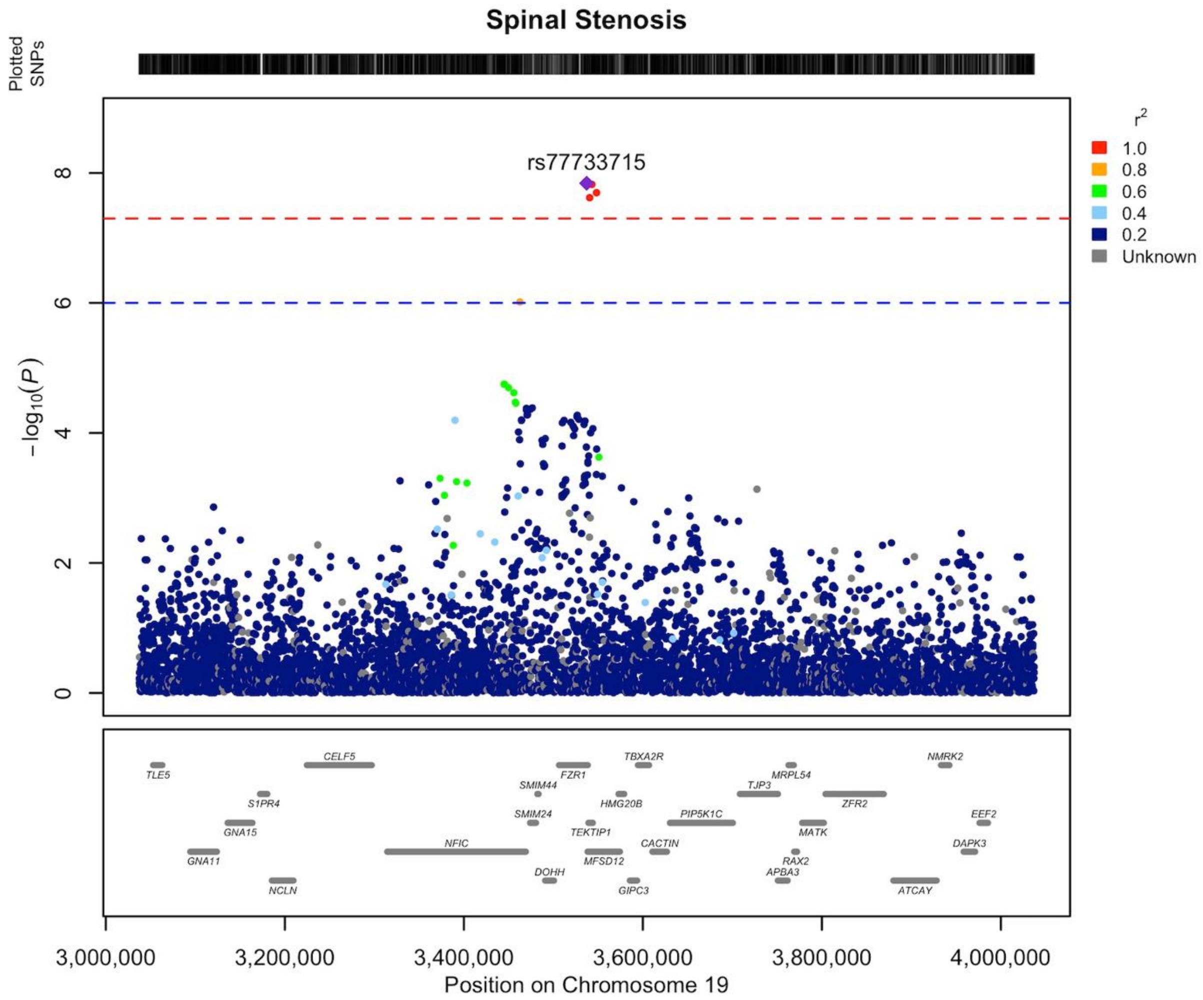


**Supplementary Data 42** Regional association plot of novel LSS association on chromosome 19 area of 3.0-4.0 MB. Our findings indicate that the gene responsible for the locus's association is most likely *CACTIN (cactin, spliceosome C complex subunit).* The LD structure of the plot does not fully represent the meta-analysis as it was calculated using FinnGen. The Locuszooms (https://github.com/Geeketics/LocusZooms) R package was used to generate the plot, and the Ensembl archive (https://jul2023.archive.ensembl.org) was used for the gene list.

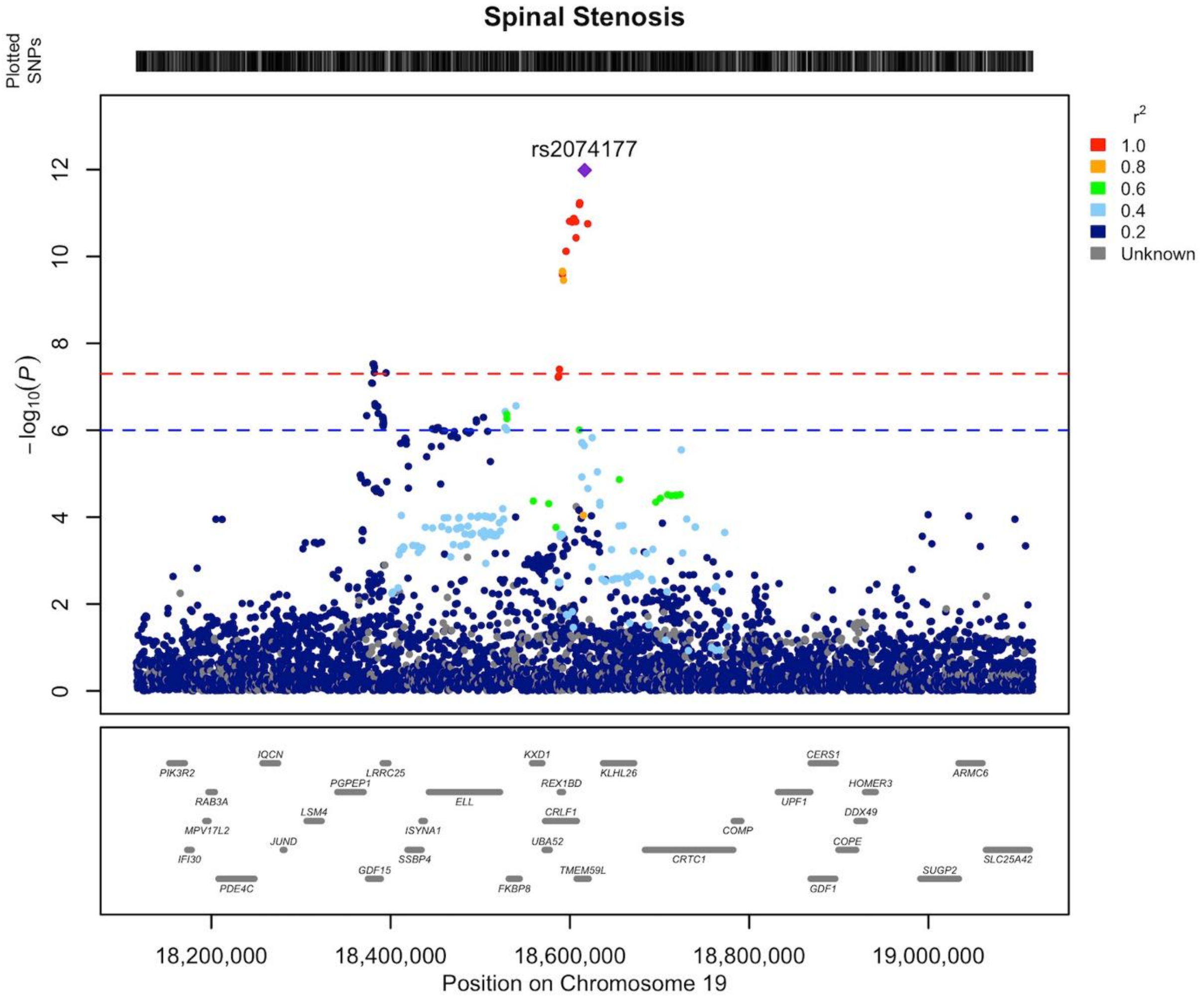


**Supplementary Data 43** Regional association plot of novel LSS association on chromosome 19 area of 18.1-19.1 MB. Our findings indicate that the gene responsible for the locus's association is most likely *CRLF1 (cytokine receptor like factor 1)*. The LD structure of the plot does not fully represent the meta-analysis as it was calculated using FinnGen. The Locuszooms (https://github.com/Geeketics/LocusZooms) R package was used to generate the plot, and the Ensembl archive (https://jul2023.archive.ensembl.org) was used for the gene list.

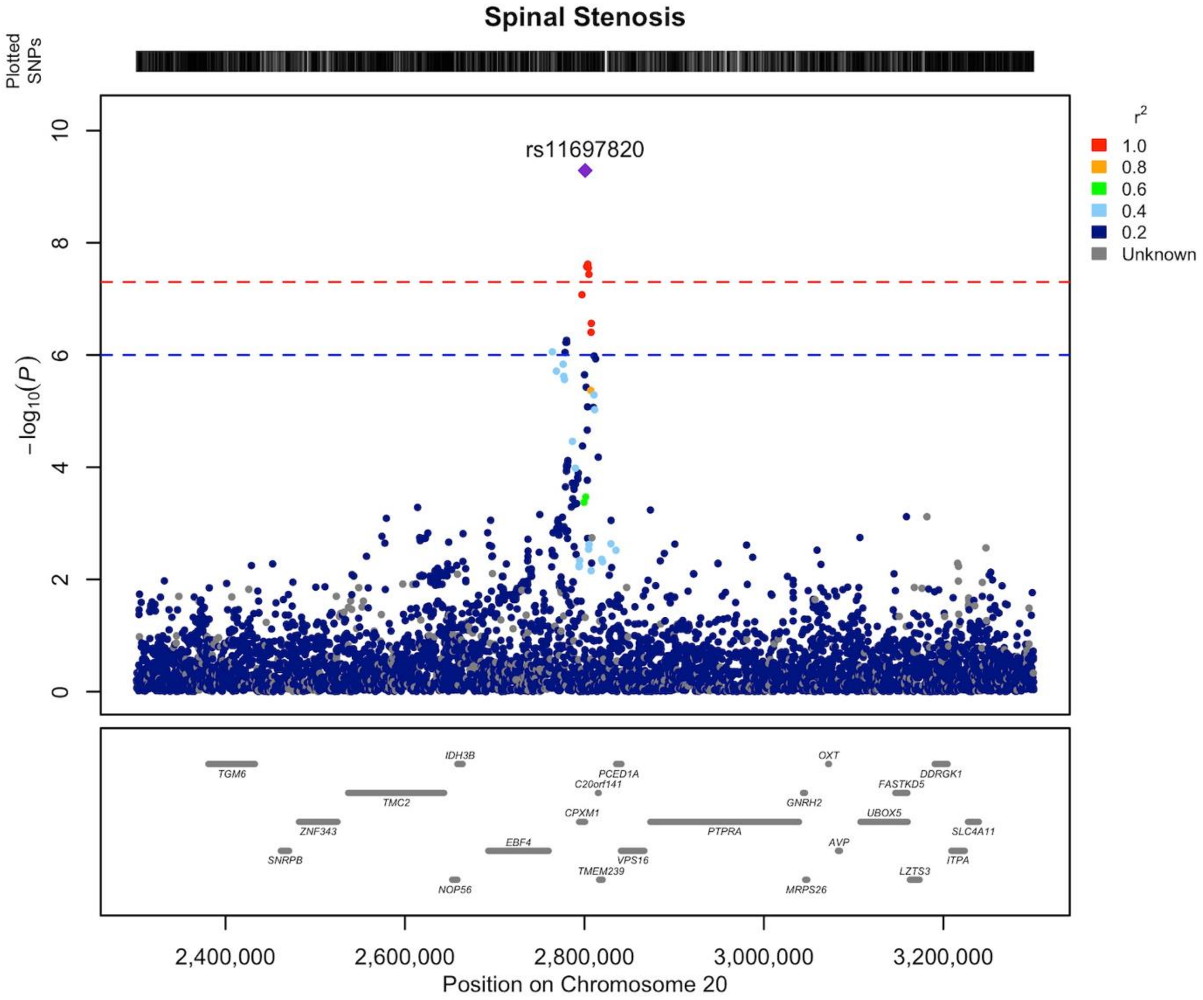


**Supplementary Data 44** Regional association plot of novel LSS association on chromosome 20 area of 2.3-3.3 MB. Our findings indicate that the gene responsible for the locus's association is most likely *DDRGK1 (DDRGK domain containing 1).* The LD structure of the plot does not fully represent the meta-analysis as it was calculated using FinnGen. The Locuszooms (https://github.com/Geeketics/LocusZooms) R package was used to generate the plot, and the Ensembl archive (https://jul2023.archive.ensembl.org) was used for the gene list.

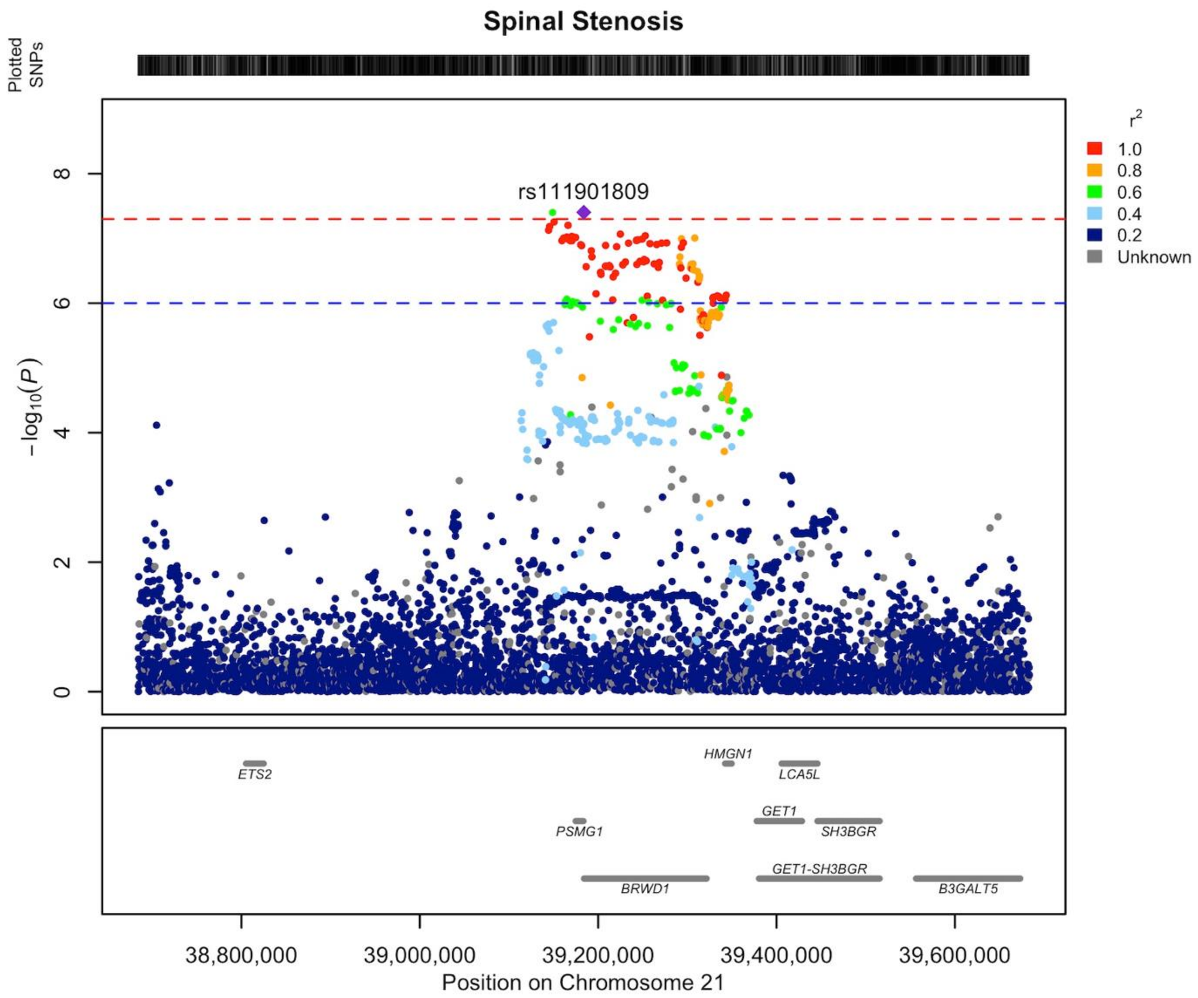


**Supplementary Data 45** Regional association plot of novel LSS association on chromosome 21 area of 38.7-39.7 MB. Our findings indicate that the gene responsible for the locus's association is most likely *ETS2 (ETS proto-oncogene 2, transcription factor).* The LD structure of the plot does not fully represent the meta-analysis as it was calculated using FinnGen. The Locuszooms (https://github.com/Geeketics/LocusZooms) R package was used to generate the plot, and the Ensembl archive (https://jul2023.archive.ensembl.org) was used for the gene list.

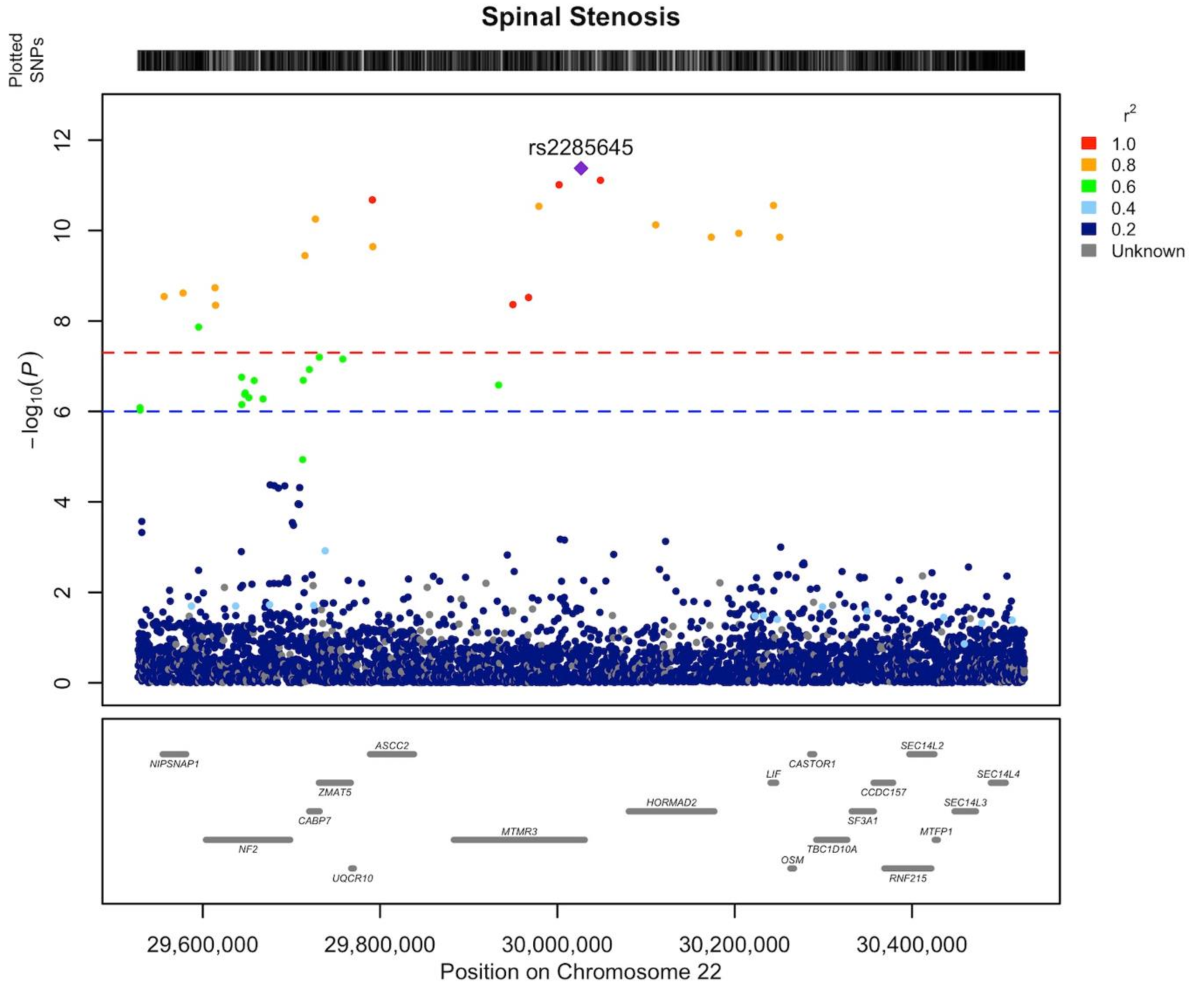


**Supplementary Data 46** Regional association plot of novel LSS association on chromosome 21 area of 29.5-30.5 MB. Our findings indicate that the gene responsible for the locus's association is most likely *OSM (oncostatin M)*. The LD structure of the plot does not fully represent the meta-analysis as it was calculated using FinnGen. The Locuszooms (https://github.com/Geeketics/LocusZooms) R package was used to generate the plot, and the Ensembl archive (https://jul2023.archive.ensembl.org) was used for the gene list.

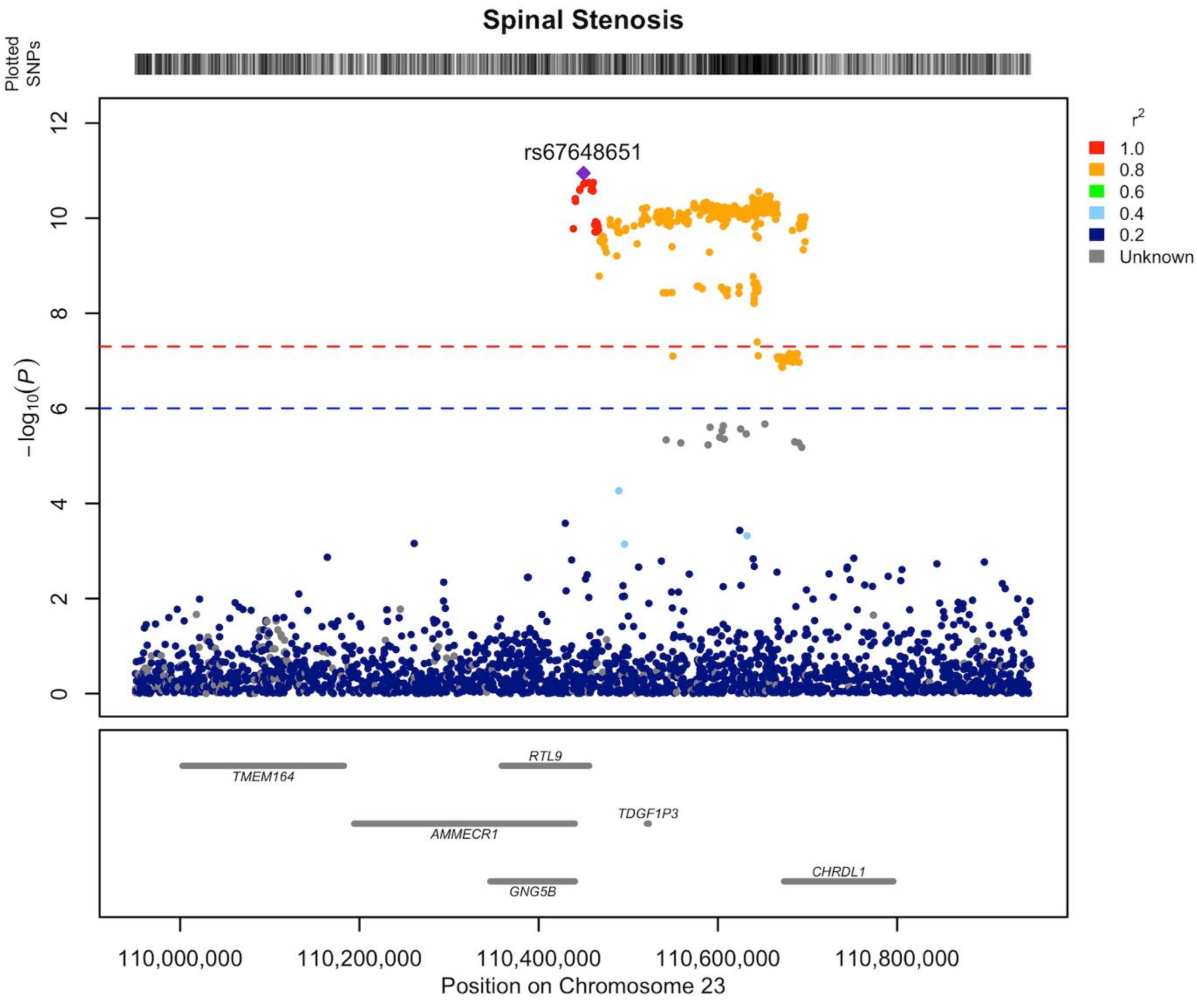


**Supplementary Data 47** Regional association plot of novel LSS association on chromosome X area of 109.9-110.9 MB. Our findings indicate that the gene responsible for the locus's association is most likely *CHRDL1 (chordin like 1).* The LD structure of the plot does not fully represent the meta-analysis as it was calculated using FinnGen. The Locuszooms (https://github.com/Geeketics/LocusZooms) R package was used to generate the plot, and the Ensembl archive (https://jul2023.archive.ensembl.org) was used for the gene list. Both sexes were included in the LD calculation.
