## Supplementary materials for "Genetic architecture of lumbar spinal stenosis"

[1] Research Unit of Health Sciences and Technology, Faculty of Medicine, University of Oulu, Oulu, Finland
[2] Biocenter Oulu, Oulu, Finland
[3] Medical Research Center Oulu, Oulu University Hospital and University of Oulu, Oulu, Finland
[4] Research unit of Population Health, Faculty of Medicine, and Biocenter Oulu, University of Oulu, Oulu, Finland
[5] A list of FinnGen authors and their affiliations available in the supplements
[6] Estonian Genome Centre, Institute of Genomics, University of Tartu, Tartu, Estonia
[7] A list of Estonian Biobank Research Team authors and their affiliations available in the supplements
[8] Institute for Molecular Medicine Finland (FIMM), HiLIFE, University of Helsinki
[9] Neurosurgery, Institute of Clinical Medicine, School of Medicine, Faculty of Health Sciences, University of Eastern Finland and Department of Neurosurgery, Kuopio University Hospital, Kuopio, Finland
[10] Rehabilitation Services of Wellbeing Services County of South Karelia, Lappeenranta, Finland
* These authors contributed equally to this work

**Supplementary Note.** FinnGen DF9 Ethics statement
**Fig. S1** Lead variant effect estimates in every dataset
**Fig. S2** Lead variant effect estimates in meta-analysis and sensitivity analysis
**Fig. S3** MAGMA tissue expression analysis results
**Fig. S4** Kaplan Meier plots for potential early-onset variants
**Fig. S5.1** Scatterplot for whole body fat-free mass that could potentially be causal for LSS
**Fig. S5.2** Scatterplot for body mass index (BMI) that could potentially be causal for LSS
**Fig. S5.3** Scatterplot for hip circumference that could potentially be causal for LSS
**Fig. S5.4** Scatterplot for whole body fat mass that could potentially be causal for LSS
**Fig. S6** Scatterplot for back pain for which LSS is potentially causal
**Fig. S7.1** Leave-one-out: Whole body fat-free mass -> LSS
**Fig. S7.2** Leave-one-out: Body mass index (BMI) -> LSS
**Fig. S7.3** Leave-one-out: Hip circumference -> LSS
**Fig. S7.4** Leave-one-out: Whole body fat mass -> LSS
**Fig. S8** Leave-one-out: LSS -> Back pain

**Table S1** Study populations and phenotype characterization
**Table S2** The lead variants at the 56 LSS-associated loci observed in the meta-analysis
**Table S3** Structural information about lead variants and potential candidate genes
**Table S4** Cohort level odds ratios and effect allele frequencies
**Table S5** Effect estimate differences between meta-analysis and surgical GWAS
**Table S6** FastBAT results
**Table S7** eQTL colocalization results
**Table S8** Cumulative morbidities and age differences between lead variant homozygotes
**Table S9** Genetic correlations between LSS and 517 other traits
**Table S10** Mendelian randomization results
**Table S11** Bi-directional Mendelian randomization risk factors

**References**

**Supplementary Note.** FinnGen R9 Ethics statement

Patients and control subjects in FinnGen provided informed consent for biobank research, based on the Finnish Biobank Act. Alternatively, separate research cohorts, collected prior the Finnish Biobank Act came into effect (in September 2013) and start of FinnGen (August 2017), were collected based on study-specific consents and later transferred to the Finnish biobanks after approval by Fimea (Finnish Medicines Agency), the National Supervisory Authority for Welfare and Health. Recruitment protocols followed the biobank protocols approved by Fimea. The Coordinating Ethics Committee of the Hospital District of Helsinki and Uusimaa (HUS) statement number for the FinnGen study is Nr HUS/990/2017.

The FinnGen study is approved by Finnish Institute for Health and Welfare (permit numbers: THL/2031/6.02.00/2017, THL/1101/5.05.00/2017, THL/341/6.02.00/2018, THL/2222/6.02.00/2018, THL/283/6.02.00/2019, THL/1721/5.05.00/2019 and THL/1524/5.05.00/2020), Digital and population data service agency (permit numbers: VRK43431/2017-3, VRK/6909/2018-3, VRK/4415/2019-3), the Social Insurance Institution (permit numbers: KELA 58/522/2017, KELA 131/522/2018, KELA 70/522/2019, KELA 98/522/2019, KELA 134/522/2019, KELA 138/522/2019, KELA 2/522/2020, KELA 16/522/2020), Findata permit numbers THL/2364/14.02/2020, THL/4055/14.06.00/2020,,THL/3433/14.06.00/2020, THL/4432/14.06/2020, THL/5189/14.06/2020, THL/5894/14.06.00/2020, THL/6619/14.06.00/2020, THL/209/14.06.00/2021, THL/688/14.06.00/2021, THL/1284/14.06.00/2021, THL/1965/14.06.00/2021, THL/5546/14.02.00/2020, THL/2658/14.06.00/2021, THL/4235/14.06.00/202, Statistics Finland (permit numbers: TK-53-1041-17 and TK/143/07.03.00/2020 (earlier TK-53-90-20) TK/1735/07.03.00/2021, TK/3112/07.03.00/2021) and Finnish Registry for Kidney Diseases permission/extract from the meeting minutes on 4th July 2019.

The Biobank Access Decisions for FinnGen samples and data utilized in FinnGen Data Freeze 9 include: THL Biobank BB2017_55, BB2017_111, BB2018_19, BB_2018_34, BB_2018_67, BB2018_71, BB2019_7, BB2019_8, BB2019_26, BB2020_1, Finnish Red Cross Blood Service Biobank 7.12.2017, Helsinki Biobank HUS/359/2017, HUS/248/2020, Auria Biobank AB17-5154 and amendment #1 (August 17 2020), AB20-5926 and amendment #1 (April 23 2020) and it´s modification (Sep 22 2021), Biobank Borealis of Northern Finland_2017_1013, Biobank of Eastern Finland 1186/2018 and amendment 22 § /2020, Finnish Clinical Biobank Tampere MH0004 and amendments (21.02.2020 & 06.10.2020), Central Finland Biobank 1-2017, and Terveystalo Biobank STB 2018001 and amendment 25th Aug 2020.

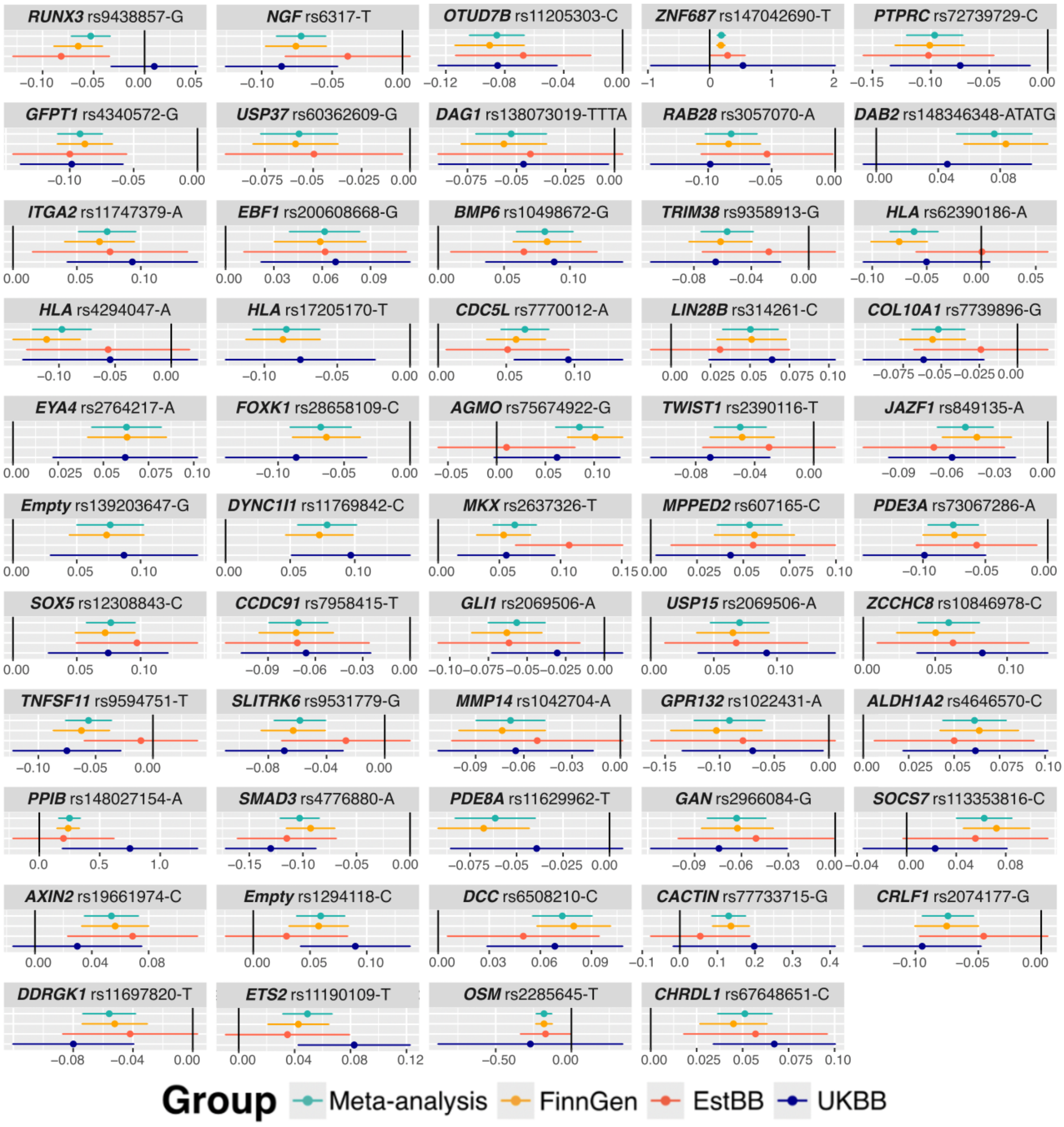


**Fig. S1.** The forest plot displays the effect estimates and 95% confidence intervals for LSS-associated lead variants in the meta-analysis [green], as well as each dataset separately (FinnGen [orange], Estonian Biobank [red], and UK Biobank [blue]). Candidate gene, rsid and effect allele were used as variant identifiers. Table S4 displays the cohort level odds ratios and effect allele frequencies.

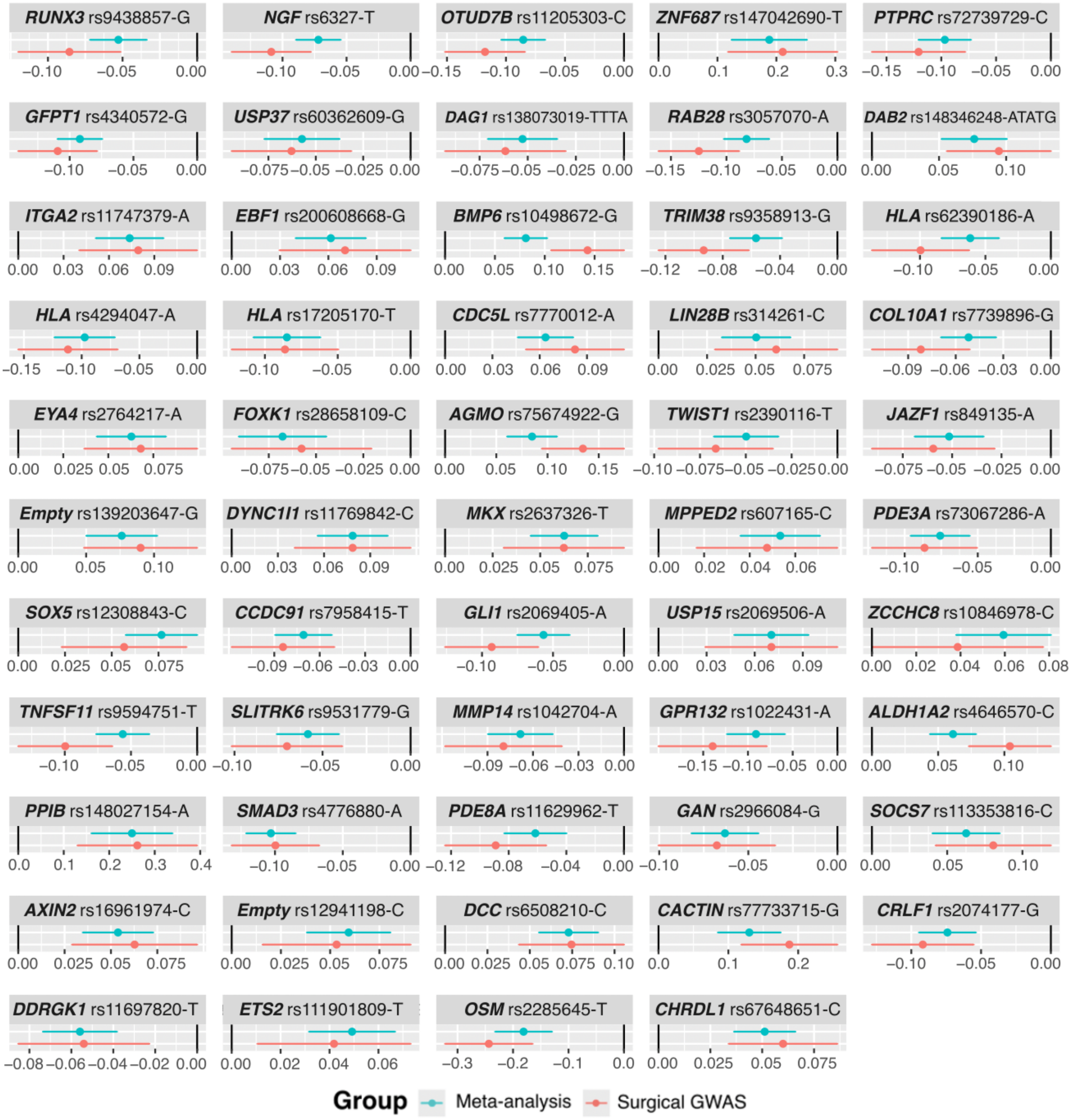


**Fig. S2.** A comparison of the effect sizes of the lead variants discovered in the original meta-analysis (Meta-analysis, ICD-10:M48.0 [green]) and in sensitivity analysis with LSS patients requiring surgery (Surgical GWAS, *Nomesco v1.15*: ABC36, ABC56, ABC66, ABC99, NAG61, NAG62, NAG63, NAG66, NAG67, and NAG99 [red]). Between meta-analysis and surgical GWAS we observed statistically significant effect differences for *NGF*, *RAB28*, *BMP6*, *TRIM38*, *AGMO*, *TNFSF11* and *ALDH1A2* variants (Fig. 1B). Results of the effect differences and p-values for difference can also be seen from Table S5.

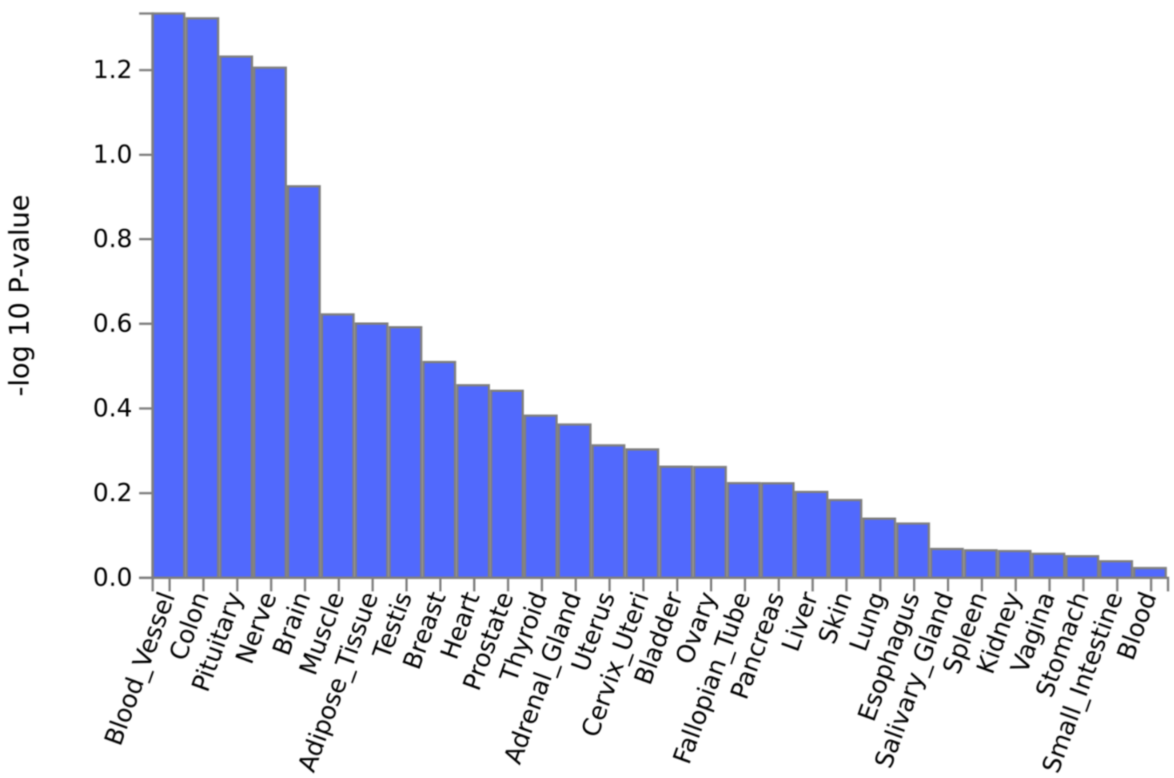


**Fig S3.** MAGMA tissue expression analysis[1] results, no genome-wide significant results were observed in tissue expression analysis. y-axis, -log 10 P-value; x-axis, tissues (GTEx Output-General tissues). Analysis was performed by using FUMA[2].

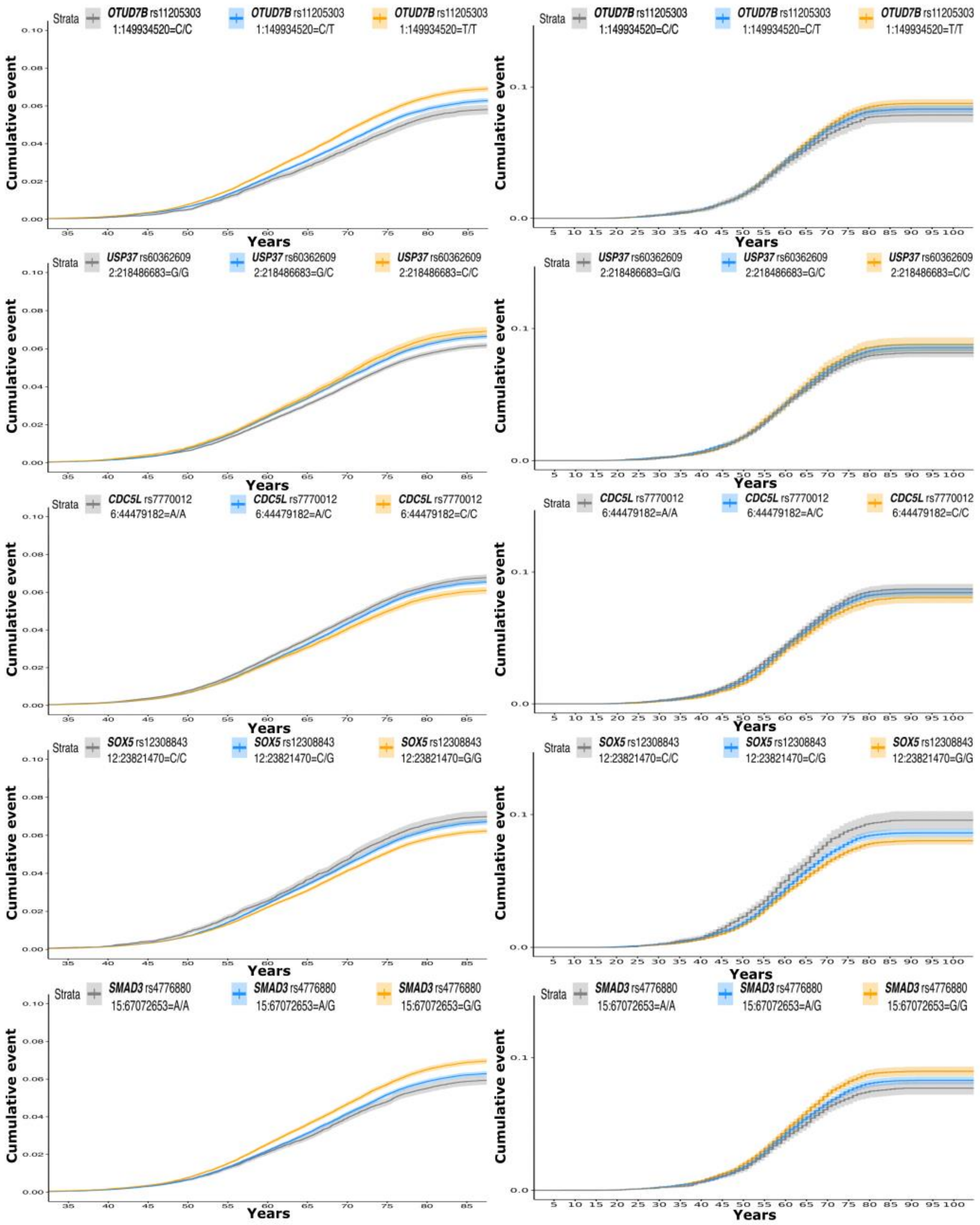


**Fig. S4** Kaplan-Meier plots for variants located near the *OTUD7B*, *USP37*, *CDC5L*, *SOX5* and *SMAD3* genes. For these variants, we observed that the cumulative incidence of LSS differed between homozygotes already at the age of 40. The graphs on the left are based on FinnGen results, while those on the right are based on Estonian Biobank replication. Age in years is on the x-axis, and the corresponding fraction of the cases is on the y-axis. The grey line depicts homozygotes for the effect allele, the orange homozygotes for the other allele, and the blue line corresponds to heterozygotes having one copy of each allele. The heterozygote differences were not tested and are just shown for illustration.

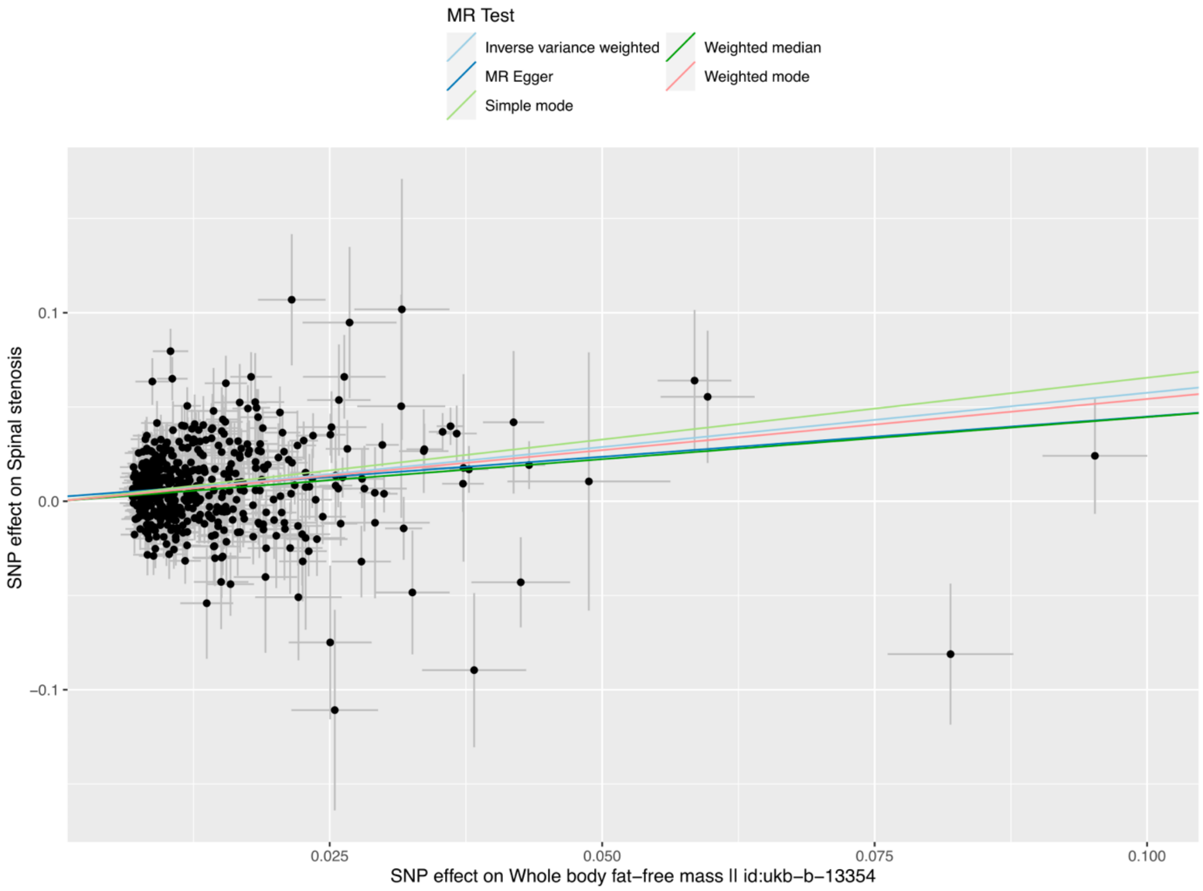


**Fig. S5.1** Scatterplot for whole body fat-free mass that could potentially be causal for LSS. Genetic instruments for LSS were extracted from the meta-analysis conducted in FinnGen and Estonian Biobank. For "Whole body fat-free mass || id:ukb-b-13354" from the GWAS database provided by the MRC-IEU, which is integrated in the TwoSampleMR R library. European population references was utilised in LD pruning, with thresholds of r2=0.001 and clumping window of 10kb. The error bars indicate the 95% confidence intervals of the SNP effects.

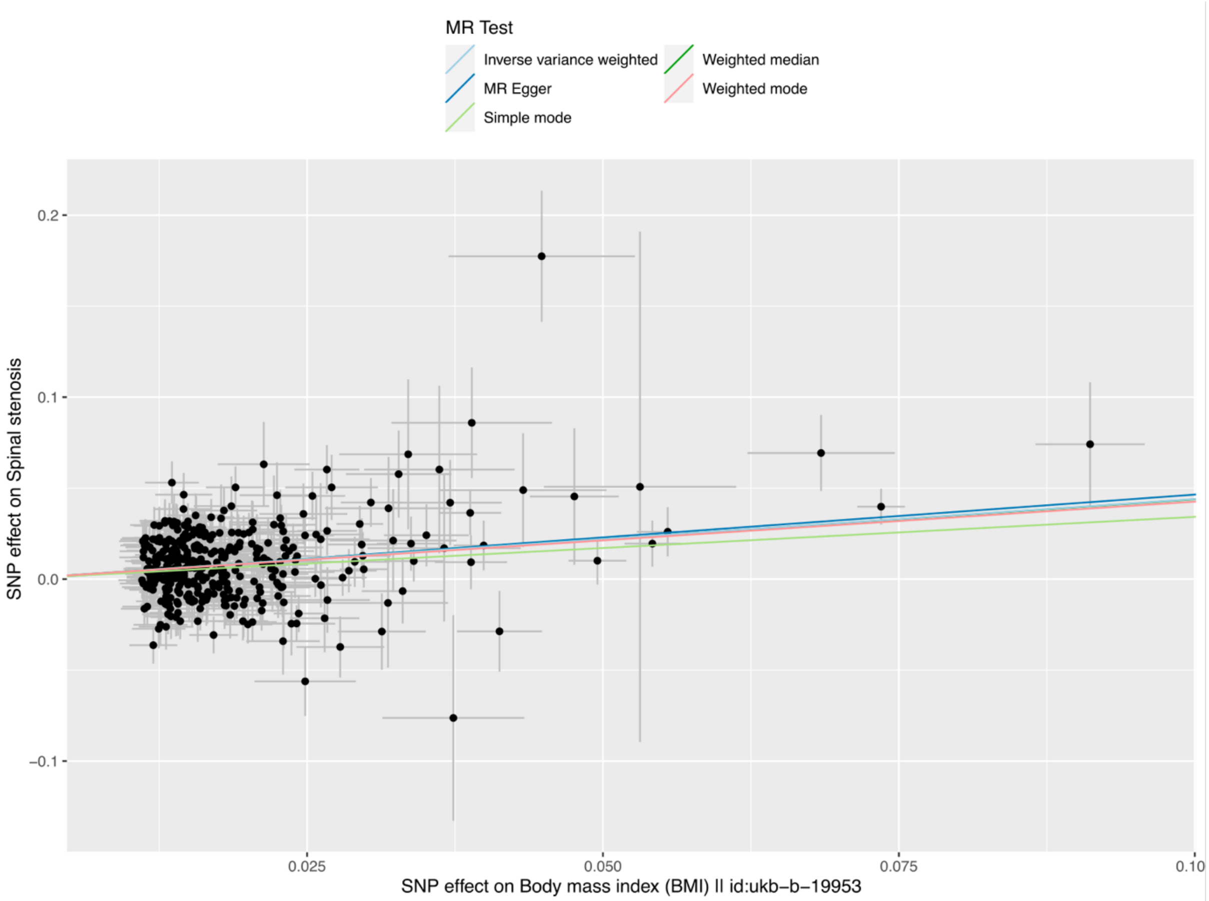


**Fig. S5.2** Scatterplot for body mass index (BMI) that could potentially be causal for LSS. Genetic instruments for LSS were extracted from the meta-analysis conducted in FinnGen and Estonian Biobank. For "Body mass index (BMI) || id:ukb-b-19953" from the GWAS database provided by the MRC-IEU, which is integrated in the TwoSampleMR R library. European population references was utilised in LD pruning, with thresholds of r2=0.001 and clumping window of 10kb. The error bars indicate the 95% confidence intervals of the SNP effects.

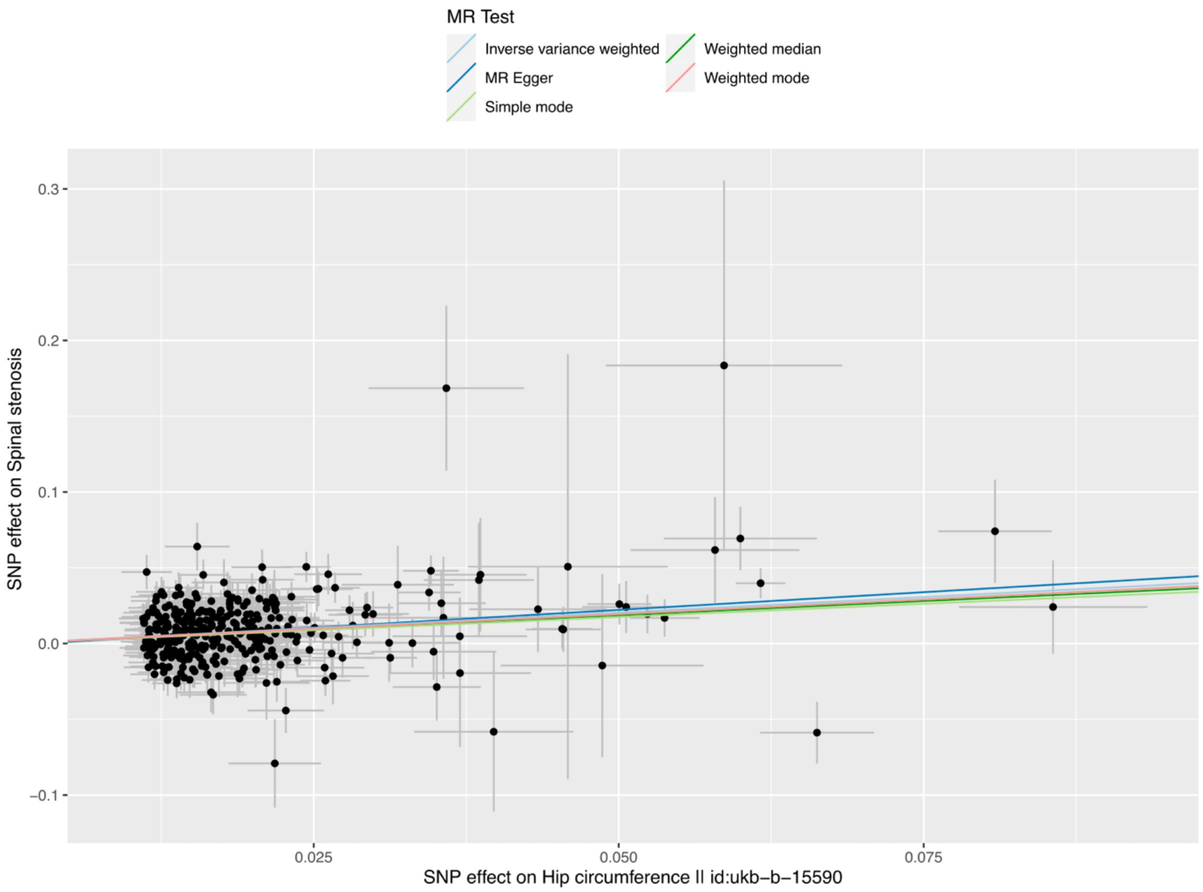


**Fig. S5.3** Scatterplot for hip circumference that could potentially be causal for LSS. Genetic instruments for LSS were extracted from the meta-analysis conducted in FinnGen and Estonian Biobank. For "Hip circumference || id:ukb-b-15590" from the GWAS database provided by the MRC-IEU, which is integrated in the TwoSampleMR R library. European population references was utilised in LD pruning, with thresholds of r2=0.001 and clumping window of 10kb. The error bars indicate the 95% confidence intervals of the SNP effects.

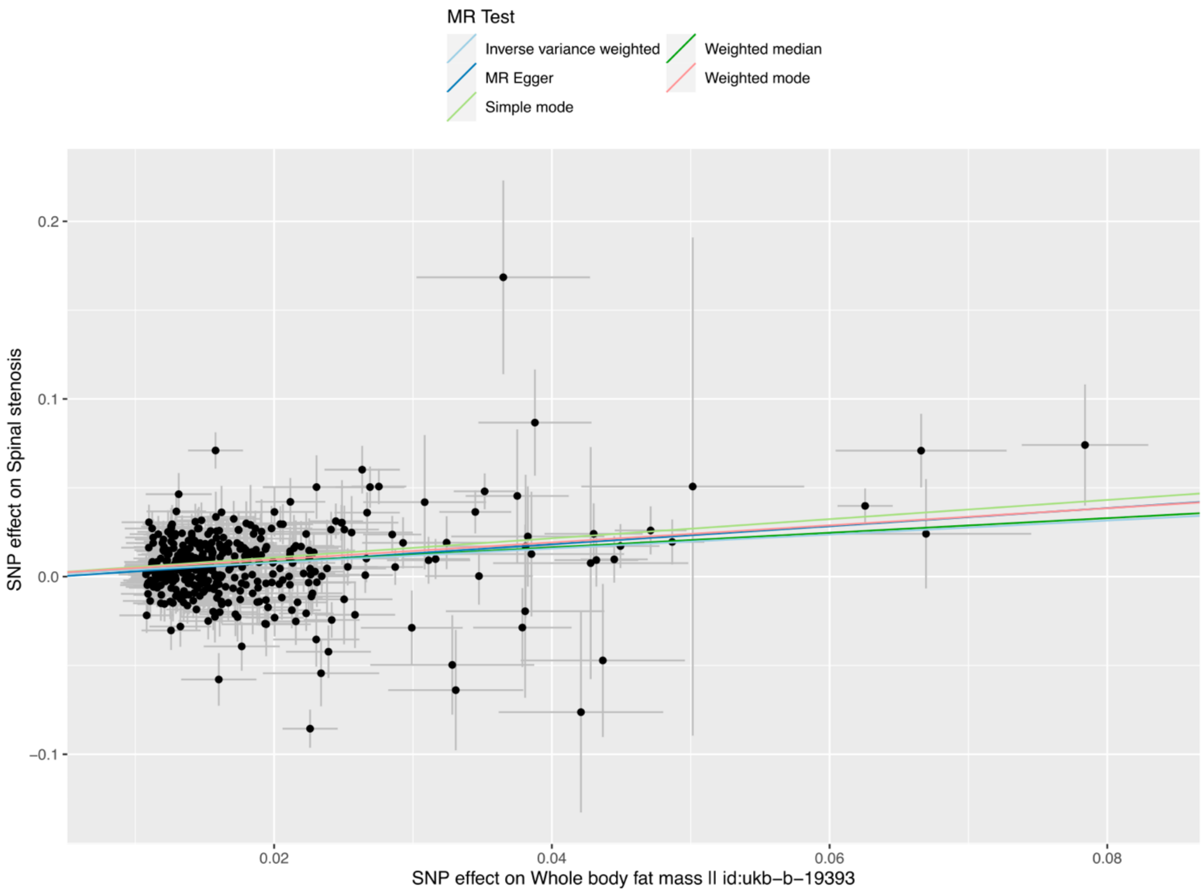


**Fig. S5.4** Scatterplot for whole body fat mass that could potentially be causal for LSS. Genetic instruments for LSS were extracted from the meta-analysis conducted in FinnGen and Estonian Biobank. For "Whole body fat mass || id:ukb-b-19393" from the GWAS database provided by the MRC-IEU, which is integrated in the TwoSampleMR R library. European population references was utilised in LD pruning, with thresholds of r2=0.001 and clumping window of 10kb. The error bars indicate the 95% confidence intervals of the SNP effects.

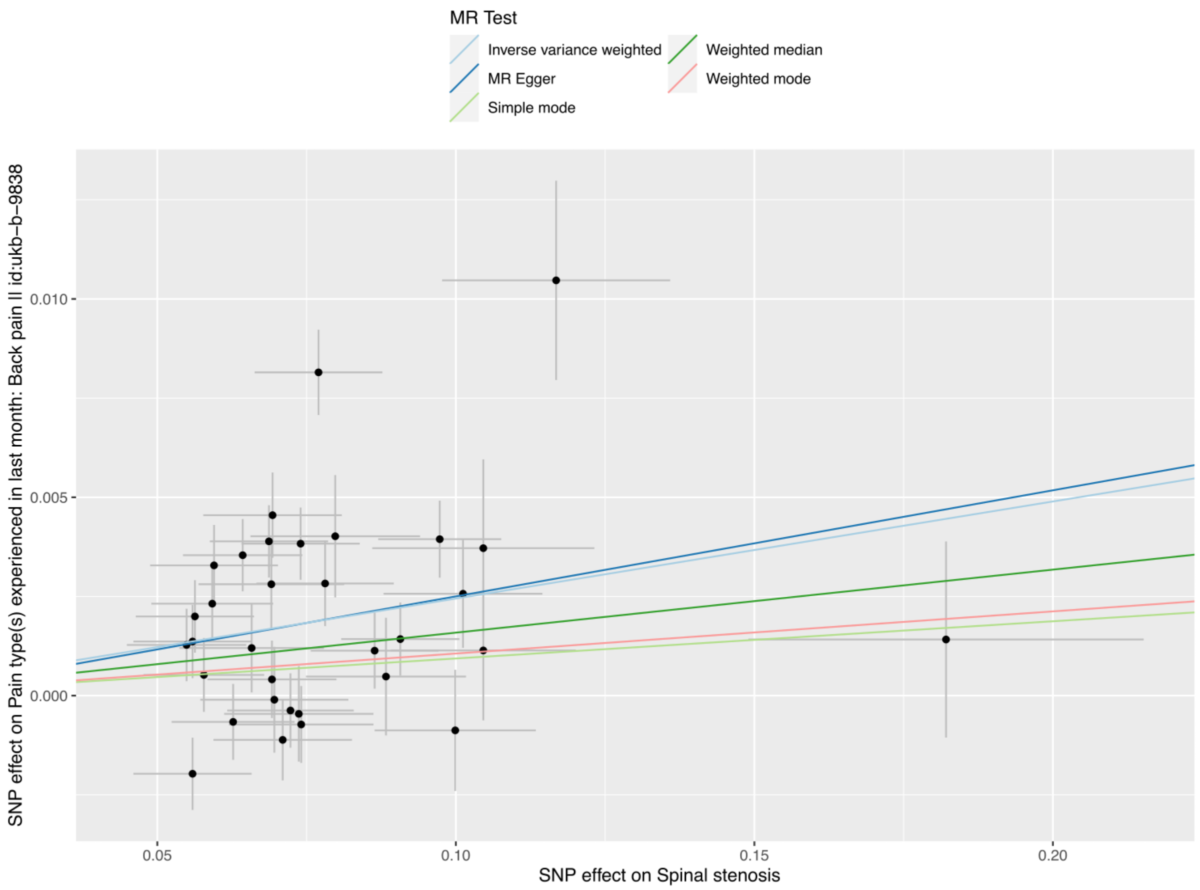


**Fig. S6** Scatterplot for back pain for which LSS is potentially causal. Genetic instruments for LSS were extracted from the meta-analysis conducted in FinnGen and Estonian Biobank. For " Pain type(s) experienced in last month: Back pain || id:ukb-b-9838" from the GWAS database provided by the MRC-IEU, which is integrated in the TwoSampleMR R library. European population references was utilised in LD pruning, with thresholds of r2=0.001 and clumping window of 10kb. The error bars indicate the 95% confidence intervals of the SNP effects.

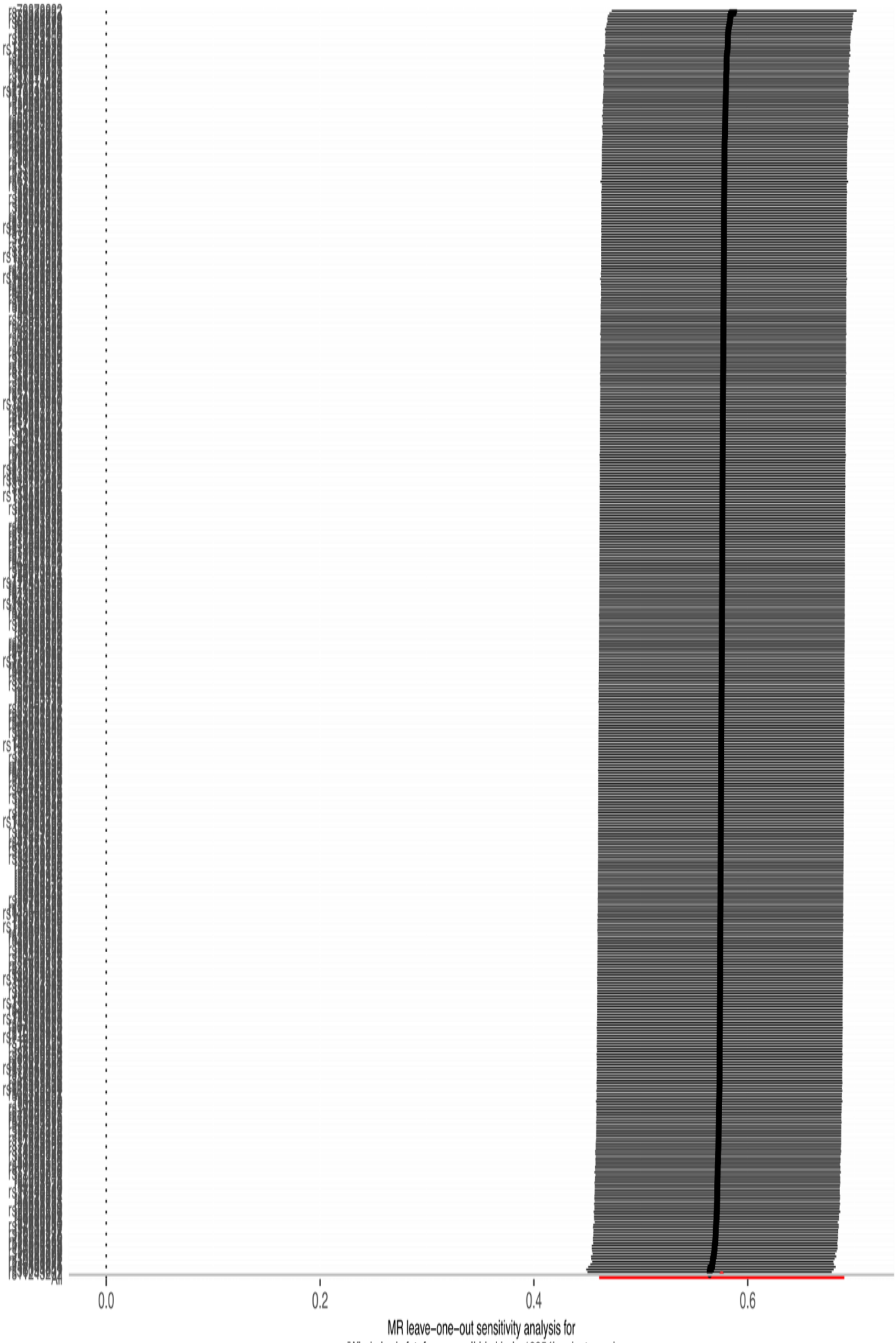


**Fig. S7.1** Leave-one-out: Whole body fat-free mass -> LSS

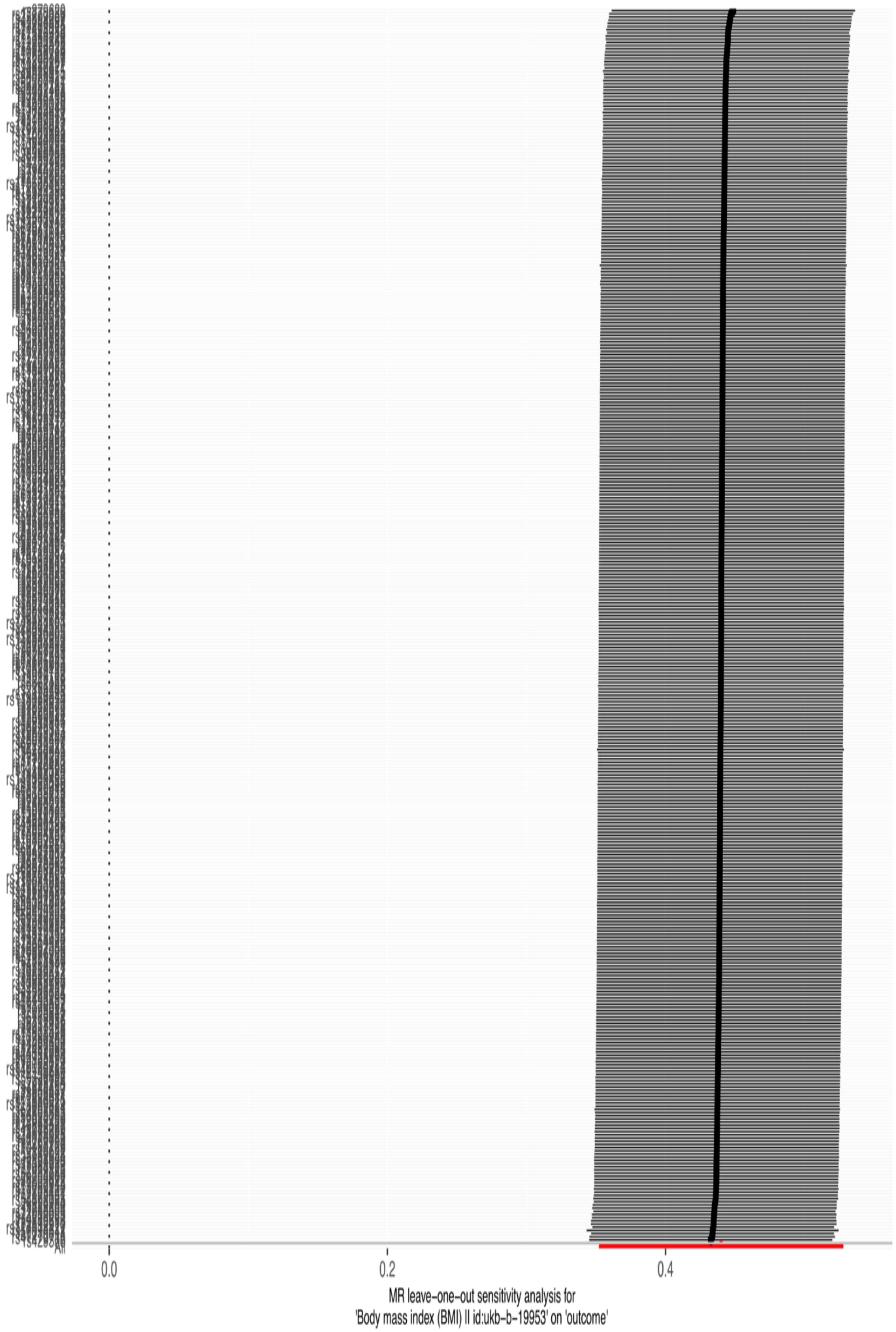


**Fig. S7.2** Leave-one-out: Body mass index (BMI) -> LSS

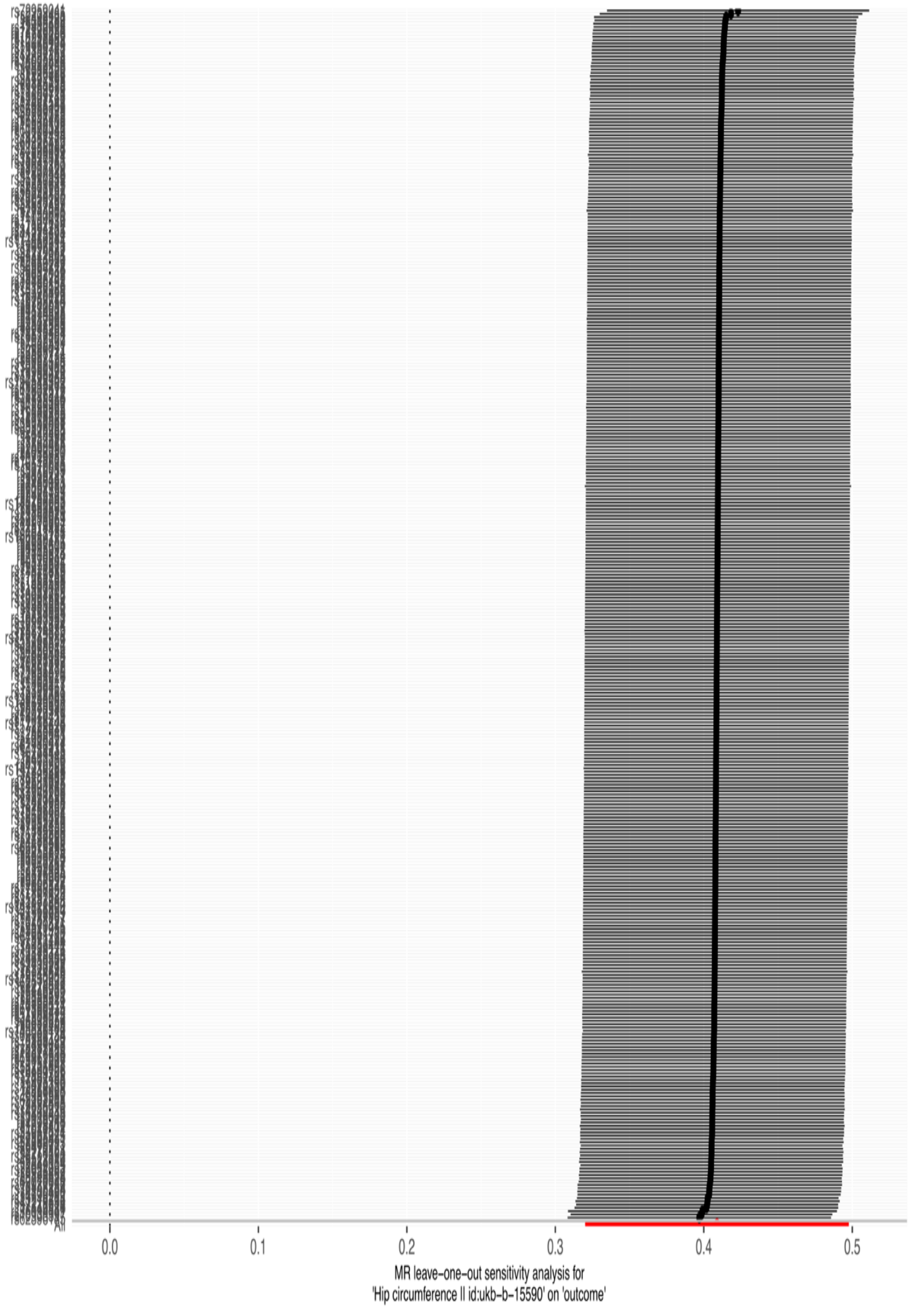


**Fig. S7.3** Leave-one-out: Hip circumference -> LSS

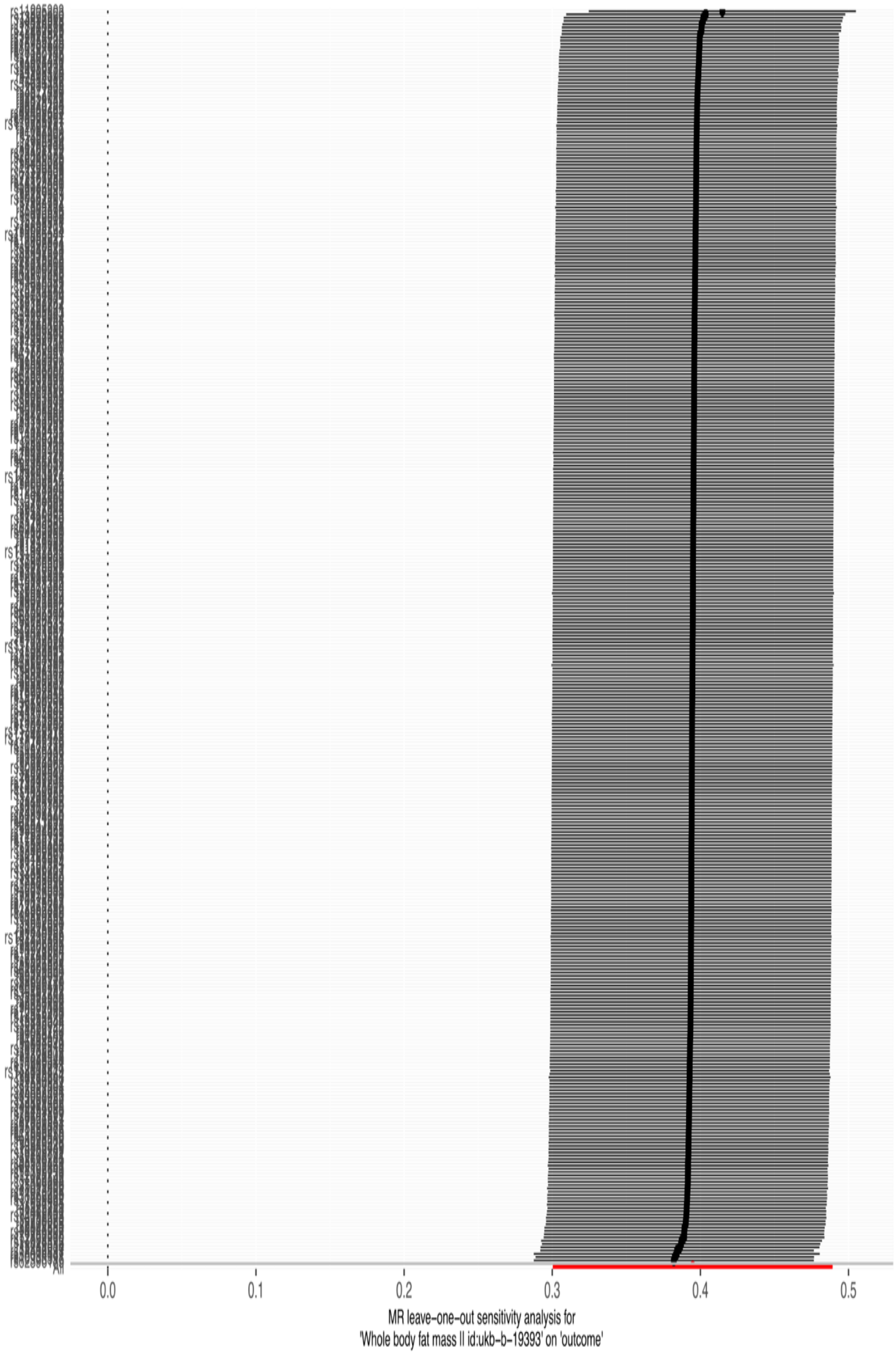


**Fig. S7.4** Leave-one-out: Whole body fat mass -> LSS

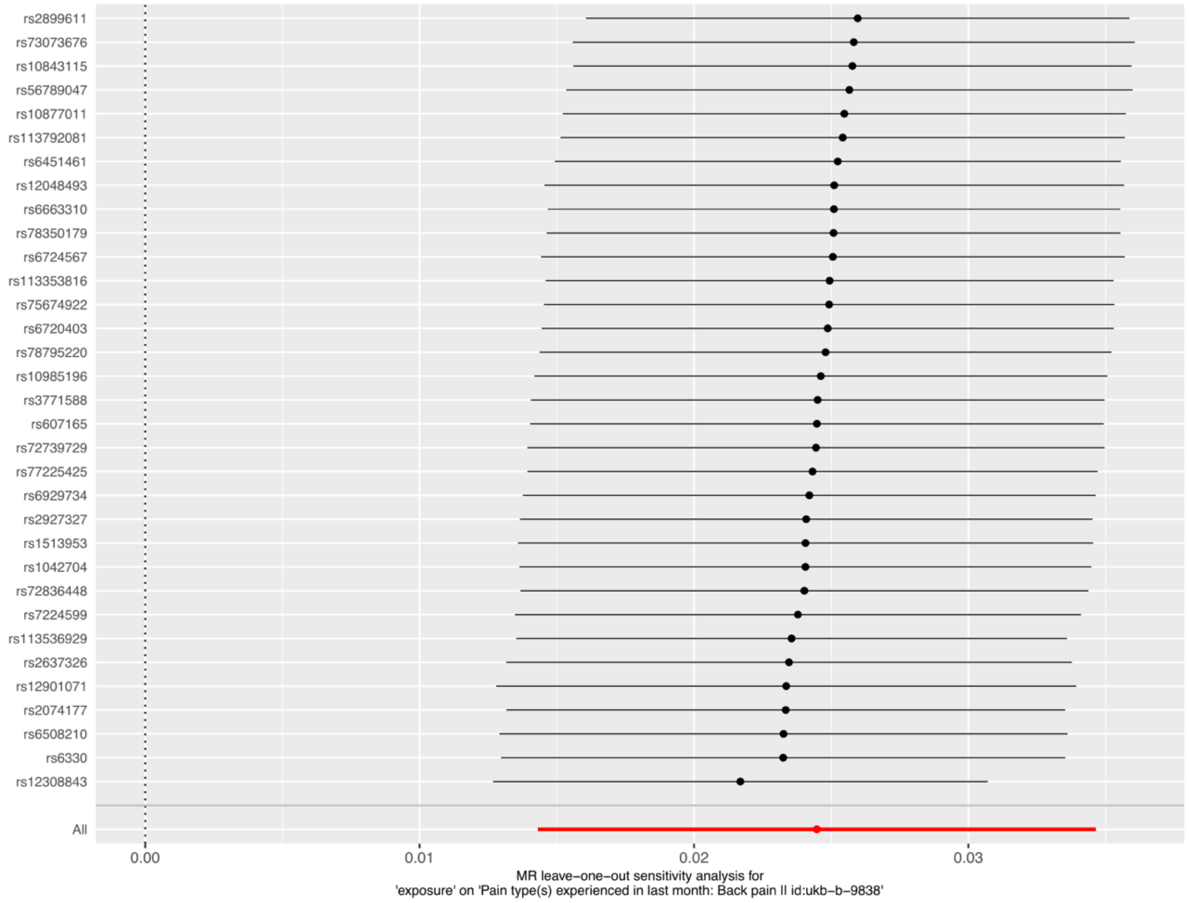


**Fig. S8.1** Leave-one-out: LSS -> Back pain

**Table S1** Case and controls in all study populations and phenotype characterization.

| Study population | Cases | Controls | Sample prevalence | Total |
|---|---|---|---|---|
| FinnGen | 18 989 | 270 964 | 6.5% | 289 953 |
| Estonian Biobank | 6146 | 66 533 | 8.4% | 73 279 |
| UK Biobank | 5134 | 401 917 | 1.3% | 407 051 |
| **Meta-analysis** | **30 269** | **739 414** | **3.9%** | **769 683** |
| Sensitivity analysis (FinnGen) | | | | |
| Surgical GWAS | 8627 | 270 773 | 3.1% | 279 400 |

| Study | Revision / Codes | Codes |
|---|---|---|
| **Meta-analysis** | | |
| EstBB | ICD-10 | M48.0 |
| UKBB | ICD-10 | M48.0 |
| FinnGen | ICD-10 | M48.0 |
| | ICD-9 | 7230A, 7240A, 7240B |
| FinnGen control exclusions | ICD-10 | M40 (M40.0, M40.1, M40.2, M40.3, M40.4, M40.5), M41 (M41.0, M41.1, M41.2, M41.3, M41.4, M41.5, M41.6, M41.8, M41.9), M42 (M42.0, M42.1, M42.9), M43 (M43.0, M43.1, M43.2, M43.3, M43.4, M43.4, M43.5, M43.6, M43.8, M43.9), M45, M46 (M46.0, M46.1, M46.2, M46.3, M46.4, M46.5, M46.8, M46.9), M47 (M47.0, M47.1, M47.2,M47.8, M47.9), M48.1, M48.2, M48.3, M48.4, M48.5, M48.8, M48.9, M49 (M49.0, M49.1, M49.2, M49.3, M49.4, M49.5, M49.8), M50 (M50.0, M50.1, M50.2, M50.3, M50.8, M50.9), M51 (M51.1-M51.9), M53 (M53.0, M53.1, M53.2, M53.3, M53.8, M53.9), M54 (M54.0, M54.1, M54.2, M54.3, M54.4, M54.5, M54.6, M54.8, M54.9), M76 (M76.0, M76.1, M76.2, M76.3, M76.4, M76.5, M76.6, M76.7, M76.8, M76.9) |
| | ICD-9 | 737.0A, 727.1A, 737.2A, 738.4A, 723.5A, 732.8A, 732.8B, 732.8X, 732.9X, 737.9X, 720.0A, 720.2A, 720.8A, 720.9X, 721.0A, 721.1A, 721.2A, 721.3A, 721.4A, 724.0B, 721.6A, 722.7A, 722.7C, 722.4A, 722.0A, 722.7B, 722.4B, 722.8A, 723.2A, 723.3A, 724.6A, 724.8X, 724.9X, 724.4A, 723.4A, 723.1A, 724.2A, 724.1A, 724.5A, 726.5B, 726.6G, 726.6C, 726.6B, 726.6D, 726.7A, 726.6B, 726.6X, 726.7B, 726.7X, 722.1A, 722.1C, 722.3A, 722.5A, 722.5B, 722.6X, 722.8C, 722.9X |
| | ICD-8 | 71720, 73599, 73500, 71240, 71310, 71311, 71312, 71319, 72699, 71320, 72500, 72501, 72509, 72810, 72820, 72828, 78880, 78830, 78888, 72899, 72898, 72890, 72870, 71700, 72850, 72800, 72898, 72899, 72510, 72519, 72520, 72551, 72559, 72588, 72599 |
| **Sensitivity analysis (FinnGen)** | | |
| **<u>Surgical GWAS</u>** | | |
| Step 1) Characterization of LSS patients | | |
| | ICD-10 | M48.0 |
| | ICD-9 | 7230A, 7240A, 7240B |
| Step 2) Patients who have undergone surgery were selected from the characterized LSS patient group. | | |
| | NOMESCO, version 1.15 | ABC36, ABC56, ABC66, ABC99, NAG61, NAG62, NAG63, NAG66, NAG67, NAG99 |
| Step 3) LSS cases that had not had surgery were omitted from the analysis. Patients who had undergone surgery but had not been diagnosed with LSS were also excluded. Otherwise, the control group was chosen in a similar manner as in the meta-analysis. | | |

**Table S2.** The lead variants at 54 LSS-associated (p<$5x10^{-8}$) loci in the meta-analysis. The meta-analysis consisted total of 30 269 cases and 739 414 controls from FinnGen, the Estonian Biobank, and the UK Biobank. Secondary signals were observed from four loci and conditional variants have been marked with *-symbol.

| Locus | Candidate gene | CHR: POS | rsid | EA | OA | OR (95% CI) | p-value | EAF | Fin Enric. | Ref. |
|---|---|---|---|---|---|---|---|---|---|---|
| **Meta-analysis (FinnGen, EstBB, UKBB)** | | | | | | | | | | |
| 1p36.11 | RUNX3 | 1:24894533 | rs9438857 | G | A | 0.90 (0.88-0.92) | 4.92e-08 | 0.31 | 0.86 | Novel |
| 1p13.2 | NGF | 1:115287365 | rs6327 | T | C | 0.93 (0.91-0.95) | 5.4e-16 | 0.47 | 1.06 | Novel |
| " | NGF* | 1:115170045 | rs1286225 | G | C | 0.94 (0.92-0.96) | 9.68e-11 | 0.68 | 1.01 | |
| 1q21.2 | OTUD7B | 1:149934520 | rs11205303 | C | T | 0.92 (0.90-0.94) | 1.76e-19 | 0.38 | 0.75 | Novel |
| 1q21.3 | ZNF687 | 1:151032308 | rs147042690 | T | C | 1.21 (1.14-1.27) | 8.5e-09 | 0.01 | 34.27 | Novel |
| 1q32.1 | PTPRC | 1:198819354 | rs72739729 | C | G | 0.91 (0.88-0.93) | 2.18e-15 | 0.15 | 1.19 | Novel |
| 2p13.3 | GFPT1 | 2:69341550 | rs4340572 | G | A | 0.91 (0.89-0.93) | 5.64e-25 | 0.57 | 0.95 | [3,4] |
| " | GFPT1* | 2:70422594 | rs17500661 | A | G | 0.95 (0.93-0.97) | 1.23e-08 | 0.29 | 1.06 | |
| 2q35 | USP37 | 2:218486683 | rs60362609 | G | C | 0.94 (0.92-0.96) | 1.47e-08 | 0.61 | 0.98 | Novel |
| 3p21.31 | DAG1 | 3:49498696 | rs138073019 | TTTA | T | 0.95 (0.93-0.97) | 9.31e-09 | 0.35 | 1.48 | Novel |
| 4p15.33 | RAB28 | 4:12893286 | rs3057070 | A | AGAG | 0.92 (0.90-0.94) | 3.22e-15 | 0.24 | 1.08 | Novel |
| 5p13.1 | DAB2 | 5:39844572 | rs148346348 | ATATG | A | 1.08 (1.05-1.10) | 7.56e-10 | 0.17 | 1.29 | Novel |
| 5q11.2 | ITGA2 | 5:53038840 | rs11747379 | A | C | 1.08 (1.05-1.10) | 9.21e-11 | 0.18 | 1.07 | Novel |
| 5q33.3 | EBF1 | 5:159017122 | rs200608668 | G | GC | 1.06 (1.04-1.09) | 3.28e-08 | 0.22 | 0.66 | Novel |
| 6p24.3 | BMP6 | 6:7797607 | rs10498672 | G | C | 1.08 (1.06-1.11) | 9.09e-14 | 0.20 | 1.24 | Novel |
| 6p22.2 | TRIM38 | 6:26239176 | rs9358913 | G | A | 0.94 (0.93-0.96) | 8.72e-10 | 0.32 | 1.62 | Novel |
| 6p22.1 | HLA | 6:30106665 | rs62390186 | A | T | 0.94 (0.92-0.96) | 3.99e-08 | 0.18 | 1.83 | Novel |
| 6p21.33 | HLA | 6:31133806 | rs4294047 | A | G | 0.91 (0.88-0.93) | 2.93e-13 | 0.11 | 1.97 | Novel |
| 6p21.32 | HLA | 6:32634706 | rs17205170 | T | G | 0.92 (0.90-0.94) | 3.33e-13 | 0.20 | 1.34 | Novel |
| 6p21.1 | CDC5L | 6:44479182 | rs7770012 | A | C | 1.07 (1.05-1.08) | 1.02e-12 | 0.53 | 0.99 | Novel |
| 6q21 | LIN28B | 6:105005542 | rs314261 | C | A | 1.05 (1.03-1.07) | 2.18e-08 | 0.57 | 1.03 | Novel |
| 6q22.1 | COL10A1 | 6:116167743 | rs7739896 | G | A | 0.95 (0.93-0.97) | 4.57e-09 | 0.51 | 0.92 | [3] |
| 6q23.2 | EYA4 | 6:132913577 | rs2764217 | A | G | 1.06 (1.05-1.08) | 1.36e-10 | 0.43 | 0.97 | Novel |
| 7p22.1 | FOXK1 | 7:4638590 | rs28658109 | C | T | 0.93 (0.91-0.96) | 1.08e-08 | 0.19 | 1.35 | Novel |
| 7p21.2 | AGMO | 7:15344063 | rs75674922 | G | A | 1.09 (1.06-1.11) | 6.72e-12 | 0.13 | 1.83 | [3] |
| 7p21.1 | TWIST1 | 7:19488615 | rs2390116 | T | C | 0.95 (0.93-0.97) | 2.93e-08 | 0.49 | 0.97 | Novel |
| 7p15.1 | JAZF1 | 7:28156794 | rs849135 | A | G | 0.95 (0.93-0.97) | 7.32e-09 | 0.49 | 0.90 | Novel |
| 7q11.22 | Empty | 7:69064281 | rs139203647 | G | GT | 1.08 (1.05-1.11) | 1.01e-08 | 0.15 | 1.22 | Novel |
| 7q21.3 | DYNC1I1 | 7:95941237 | rs11769842 | C | T | 1.08 (1.06-1.10) | 1.18e-11 | 0.75 | 1.06 | [3] |
| 10p12.1 | MKX | 10:27612430 | rs2637326 | T | G | 1.06 (1.05-1.08) | 3.44e-12 | 0.47 | 0.73 | Novel |
| 11p14.1 | MPPED2 | 11:30788512 | rs607165 | C | T | 1.05 (1.04-1.07) | 1.94e-09 | 0.44 | 1.19 | Novel |
| 12p12.2 | PDE3A | 12:20427791 | rs73067286 | A | T | 0.93 (0.91-0.95) | 2.1e-13 | 0.23 | 1.22 | [3] |
| " | PDE3A* | 12:20215335 | rs60691990 | C | T | 0.93 (0.91-0.95) | 1.86e-11 | 0.29 | 0.65 | |
| 12p12.1 | SOX5 | 12:23821470 | rs12308843 | C | G | 1.08 (1.06-1.10) | 3.3e-15 | 0.27 | 1.28 | Novel |
| 12p11.22 | CCDC91 | 12:27844985 | rs7958415 | T | C | 0.93 (0.91-0.95) | 1.07e-13 | 0.35 | 0.72 | Novel |
| " | CCDC91* | 12:27958482 | rs189814310 | C | A | 0.65 (0.51-0.80) | 3.20e-09 | 0.005 | 3.97 | |
| " | CCDC91* | 12:28156247 | rs2127315 | T | A | 0.94 (0.92-0.96) | 1.45e-08 | 0.25 | 0.85 | |
| " | CCDC91* | 12:28933226 | rs11049924 | A | C | 0.93 (0.91-0.96) | 2.90e-08 | 0.15 | 0.92 | |
| 12q14.1 | GLI1 | 12:57749071 | rs2069506 | A | C | 0.94 (0.93-0.96) | 1.56e-09 | 0.33 | 1.19 | Novel |
| 12q14.1 | USP15 | 12:62218131 | rs11174377 | A | G | 1.07 (1.05-1.10) | 2.47e-09 | 0.17 | 0.97 | Novel |
| 12q24.31 | ZCCHC8 | 12:122480161 | rs10846978 | C | T | 1.06 (1.04-1.09) | 4.49e-08 | 0.77 | 1.07 | Novel |
| 13q14.11 | TNFSF11 | 13:42423131 | rs9594751 | T | C | 0.95 (0.93-0.97) | 3.09e-08 | 0.25 | 1.13 | Novel |
| 13q31.1 | SLITRK6 | 13:85356178 | rs9531779 | G | C | 0.94 (0.93-0.96) | 9.15e-11 | 0.59 | 1.03 | [3] |
| 14q11.2 | MMP14 | 14:22843385 | rs1042704 | A | G | 0.93 (0.91-0.96) | 4.02e-10 | 0.22 | 1.00 | Novel |
| 14q32.33 | GPR132 | 14:105137078 | rs1022431 | A | C | 0.91 (0.88-0.95) | 3.76e-08 | 0.09 | 0.78 | Novel |
| 15q21.3 | ALDH1A2 | 15:58050948 | rs4646570 | C | G | 1.06 (1.05-1.08) | 5.47e-12 | 0.49 | 1.07 | Novel |
| 15q22.31 | PPIB | 15:64138226 | rs148027154 | A | T | 1.28 (1.20-1.37) | 3.52e-08 | 0.006 | 5.12 | Novel |
| 15q22.33 | SMAD3 | 15:67072653 | rs4776880 | A | G | 0.90 (0.88-0.92) | 1.05e-28 | 0.35 | 1.10 | [3] |
| 15q25.3 | PDE8A | 15:84986797 | rs11629962 | T | C | 0.94 (0.92-0.96) | 2.36e-08 | 0.25 | 1.14 | Novel |
| 16q23.2 | GAN | 16:81482778 | rs2966084 | G | A | 0.94 (0.92-0.96) | 4.3e-11 | 0.69 | 0.95 | Novel |
| 17q12 | SOCS7 | 17:38474030 | rs113353816 | C | T | 1.06 (1.04-1.09) | 4.25e-08 | 0.83 | 0.90 | Novel |
| 17q24.1 | AXIN2 | 17:65278900 | rs16961974 | C | T | 1.06 (1.04-1.07) | 2.75e-08 | 0.28 | 1.05 | Novel |
| 17q24.3 | Empty | 17:71476022 | rs12941198 | C | A | 1.06 (1.04-1.08) | 3.6e-08 | 0.78 | 1.02 | Novel |
| 18q21.2 | DCC | 18:53220821 | rs6508210 | C | A | 1.08 (1.06-1.09) | 2.64e-16 | 0.52 | 0.85 | Novel |
| 19p13.3 | CACTIN | 19:3537186 | rs77733715 | G | A | 1.14 (1.09-1.18) | 1.44e-08 | 0.03 | 5.24 | Novel |
| 19p13.11 | CRLF1 | 19:18616479 | rs2074177 | G | A | 0.93 (0.91-0.95) | 1.03e-12 | 0.25 | 1.05 | Novel |
| 20p13 | DDRGK1 | 20:2800795 | rs11697820 | T | C | 0.95 (0.93-0.96) | 5.15e-10 | 0.58 | 0.98 | Novel |
| 21q22.2 | ETS2 | 21:39183865 | rs111901809 | T | TAGA GGGG CAGG GGGC G | 1.05 (1.03-1.07) | 3.94e-08 | 0.49 | 1.32 | Novel |
| 22q12.2 | OSM | 22:30026719 | rs2285645 | T | C | 0.83 (0.78-0.89) | 4.19e-12 | 0.02 | 14.95 | Novel |
| Xq23 | CHRDL1 | X:110450046 | rs67648651 | C | T | 1.05 (1.04-1.07) | 1.13e-11 | 0.40 | 0.87 | Novel |

Candidate gene, a gene at a new locus the biological function of which is likely to explain the LSS association; CHR: POS, chromosome and position (genome build hg38); rsid; SNP markers identification number; EA, effect allele; OA, other allele; OR, odds ratio; 95% Cl, odds ratio 95% confidence interval; EAF, effect allele frequency; Fin Enric., enrichment in Finns (calculated FIN AF/NFEE AF in the Genome Aggregation Database [gnomAD, v.3.1.2], FIN AF is the allele frequency in Finns and NFEE AF is the allele frequency in Europeans (does not include Finns or Estonians)); REF, reference to literature reporting LSS association in the vicinity (± 1MB) of the lead variant. *, for this variant p-value and OR have been adjusted for the effect of original lead variant at the locus through conditional analysis, the original lead variant is always above the conditional variant in the table

**Table S3** Structural information about the LSS-associated and Surgical LSS variants. Also, biological functions for novel potential candidate genes that we determined with the help of literature and databases (Genbank[5], UniProt[6], GTEX-Portal[7]). HLA region variants were excluded from the PheWAS.

| Candidate gene | rsid | Variant type | PheWAS ⇑ | PheWAS ⇓ | A possible function related to LSS pathogenesis |
|---|---|---|---|---|---|
| ***Meta-analysis (FinnGen, Estonian Biobank, UK Biobank)*** | | | | | |
| ***RUNX3*** | rs9438857 | Downstream gene variant | - | - | *RUNX3* lowers CD8+ T cell counts, so it has a role in the regulation of immune defence[8]. |
| ***NGF*** | rs6327 | Intron | Adhesive capsulitis | Carpal tunnel syndrome, arthrosis, spondylosis, spondylolisthesis, hallux valgus | *NGF* stimulates axonal regeneration in the central nervous system as well as collateral sprouting of the peripheral sensory nerves. *NGF* production can lead to nociceptive nerve ingrowth in the spine, and it is also involved in neuronal apoptosis[9,10]. |
| ***OTUD7B*** | rs11205303 | Missense | Angina pectoris, cardiac arrhythmias, major coronary heart disease event, coronary atherosclerosis, coronary revascularization | Hallux valgus, lumbar disc herniation, carpal tunnel syndrome | *OTUD7B* controls non-canonical NF-κB activation through deubiquitination[11], its involvement in LSS pathogenesis has already been speculated in previous clinical studies concerning LSS[12]. |
| ***ZNF687*** | rs147042690 | Intron | - | - | *ZNF687* has been linked to Paget disease in earlier studies[13,14]. There is evidence that *ZNF687* may influence bone development and differentiation[15]. |
| ***PTPRC*** | rs72739729 | Intron | - | Low back pain, Spondylosis, lumbar disc herniation | Regulates T cell antigen receptor signaling[16,17]. |
| ***GFPT1*** | rs4340572 | Intron | Dupuytren contracture | Lumbar disc herniation, spondylosis, spondylolisthesis, cervical disc disorders | - |
| ***USP37*** | rs60362609 | Intron | - | - | The hedgehog pathway, which is regulated by *USP37*, has previously been connected to ligamentum flavum hypertrophy along with *GLI1*[18–20]. |
| ***DAG1*** | rs138073019 | Intron | Ulcerative proctitis, inflammatory bowel disease, ulcerative colitis | Pain, spondylosis, shoulder lesions, lumbar disc herniation, hypertension, carpal tunnel syndrome | *DAG1* plays a role in neural development and has a known role in axon guidance[21,22]. It has also been linked to muscle dystrophies[23]. |
| ***RAB28*** | rs5856174 | Intergenic | - | - | *RAB28* has been linked to kyphosis (curvature of the spine) in a previous study[24], the genetic mechanism of action is potentially related to membrane trafficking or regulation of NF-κB nuclear transportation [25,26]. |
| ***DAB2*** | rs148346348 | Intergenic | Spondylolisthesis | - | *DAB2* has multifaceted roles in signaling pathways involved in cellular differentiation, proliferation, migration, and it may contribute to immunological tolerance and suppression of inflammatory responses. |

| | | | | | |
|---|---|---|---|---|---|
| | | | | | In addition, it has been linked to TGF-β signaling[27,28]. |
| ***ITGA2*** | rs11747379 | Intron | Lumbar disc herniation, spondylolisthesis | | A transmembrane receptor for collagens and other related proteins, the alpha subunit is encoded by *ITGA2*, also referred to as integrin α2. *ITGA2* mediates the adhesion of various cell types to the extracellular matrix (ECM) by forming a heterodimer with an integrin beta subunit[29]. |
| ***EBF1*** | rs200608668 | Intron | Obesity, hypertension | Varicose veins | *EBF1* regulates early B-cell differentiation by poising or activating lineage-specific genes and repressing genes associated with alternative cell fates[30,31]. |
| ***BMP6*** | rs10498672 | Intron | Arthrosis, coxarthrosis | | *BMP6* has an important role in bone and cartilage formation and repair[32,33]. It also potentially plays a role in inducing angiogenesis and neurogenesis[34,35]. |
| ***TRIM38*** | rs9358913 | Upstream gene variant | Subacute thyroiditis | Hypothyroidism, atherosclerosis, hypertension, varicose veins, coeliac disease, arthrosis, gonarthrosis, autoimmune disease | *TRIM38* is an inflammatory modulator[36]. By suppressing NF-κB signalling, *TRIM38* can lessen apoptosis and degeneration of chondrocytes caused by IL-1β, it has been suggested that *TRIM38* may be involved in osteoarthritis[37]. |
| ***HLA*** | rs62390186 | Intron | - | - | - |
| ***HLA*** | rs4294047 | Intron | - | - | - |
| ***HLA*** | rs17205170 | Intron | - | - | - |
| ***CDC5L*** | rs7770012 | Intergenic | Bell's palsy, lumbar disc herniation, coxarthrosis, arthrosis | - | *CDC5L* has previously been associated with ossification of the posterior longitudinal ligament (OPLL)[38,39]. It has been discovered that *CDC5L* stimulates early chondrogenesis and cartilage growth and may contribute to the pathophysiology of OPLL by initiating the endochondral ossification process[40]. |
| ***LIN28B*** | rs314261 | Intron | Carpal tunnel syndrome, lumbar disc herniation, spondylosis | | *LIN28B* has been associated with bone mineral density (BMD)[41]. |
| ***COL10A1*** | rs7739896 | Intron | Myopia | Gonarthrosis, spondylolisthesis | - |
| ***EYA4*** | rs2764217 | Intergenic | - | - | Through bone remodelling, *EYA4* has been found to have an effect on BMD[42]. |
| ***FOKX1*** | rs28658109 | Intergenic | - | - | It has been discovered that *FOKX1* acts via the TGF-β pathway and has been associated with BMD[43]. Degenerative diseases of the spine have also been linked to the TGF-β pathway[44,45]. |
| ***AGMO*** | rs75674922 | Intron | - | - | - |
| ***TWIST1*** | rs2390116 | Intron | - | Lumbar disc herniation | *TWIST1* has been found to be an inflammatory regulator[46], and has also been associated with lumbar disc herniation (LDH)[47,48]. |

| | | | | | |
|---|---|---|---|---|---|
| ***JAZF1*** | rs849135 | Intron | Actinic keratosis, basal cell carcinoma | Type II diabetes, asthma, statin medication, coxarthrosis | *JAZF1* is a transcriptional repressor, which has been associated to height and spine length in addition to lowering IGF-1 serum concentrations[49,50]. |
| ***Empty*** | rs139203647 | Intergenic | - | - | - |
| ***DYNC1I1*** | rs11769842 | Intron | - | - | - |
| ***MKX*** | rs2637326 | Intergenic | Low back pain, spondylosis, lumbar disc herniation | Nonsuppurative otitis media | In addition to its well-known function in the development of tendons and ligaments, *MKX* has been discovered to be a crucial transcription factor in the formation of the intervertebral disc's annulus fibrosus[51,52]. *MKX* has also previously been associated with LDH[47,48]. |
| ***MPPED2*** | rs607165 | Intergenic | Hypertension, cervical disc disorders, arthrosis, spondylosis, depression medications | - | *MPPED2* regulates many cellular functions, such as differentiation, proliferation, and apoptosis, it also possibly regulates systemic inflammation[53,54]. |
| ***PDE3A*** | rs73067286 | Intron | Umbilical hernia | Type II diabetes | - |
| ***SOX5*** | rs12308843 | Intron | Arthrosis, lumbar disc herniation, spondylosis, gonarthrosis, low back pain | | Transcription factor involved in chondrocytes differentiation and cartilage formation[55–57]. |
| ***CCDC91*** | rs7958415 | Regulatory region variant | - | - | *CCDC91* has been previously associated with OPLL and may have a role in the regulation of osteogenesis[39,58]. |
| ***GLI1*** | rs2069506 | 3 prime UTR variant | - | Lumbar disc herniation | *GLI1* is an activator and is highly expressed in proliferating chondrocytes and the perichondrium adjacent to the prehypertrophic and hypertrophic zones[59]. *GLI1* expression has been found to be increased in hypertrophic ligamentum flavum (LF) and is required for the fibrogenesis of fibroblasts. Hypertrophy of the LF is a significant contributor to LSS[19]. |
| ***USP15*** | rs11174377 | Intron | - | - | *USP15* is an important regulator of TGF-β and BMP-signalling pathways; it modulates an alternative β-catenin pathway that stimulates bone formation, it is also a positive modulator of both TNF-α- and IL-1β-stimulated NF-κB activation[18,60]. |
| ***ZCCHC8*** | rs10846978 | Intron | Spondylosis, obesity | - | *ZCCHC8* is part of the NEXT complex, which regulates mRNA splicing[61]. *ZCCHC8* has also been found to play a role in the development of the central nervous system[62,63]. |
| ***TNFSF11*** | rs9594751 | Intron | - | - | Through bone remodelling, *TNFSF11* has been found to have an effect on BMD[42]. |
| ***SLITRK6*** | rs9531779 | Intergenic | - | - | - |
| ***MMP14*** | rs1042704 | Missense | Adhesive capsulitis, dupuytren | Lumbar disc herniation | *MMP14* missense variant is known to reduce collagen catabolism[64] |

| | | | | | |
|---|---|---|---|---|---|
| | | | contracture, shoulder lesions, fibroblastic disorders | | and through that it can be related to spinal degeneration[65]. |
| ***GPR132*** | rs1022431 | Downstream gene variant | - | - | *GPR132* reduces inflammatory responses by decreasing the number of immune cells and the release of proinflammatory cytokines and growth factors at the site of nerve injury[66]. |
| ***ALDH1A2*** | rs4646570 | Intron | Coxarthrosis, polyarthrosis | - | The enzyme retinaldehyde dehydrogenase 2, which is encoded by *ALDH1A2,* catalyses the conversion of retinaldehyde to retinoic acid, which has a role in maintaining bone and cartilage tissues. *ALDH1A2* has also been associated with osteoarthritis[67–69]. |
| ***PPIB*** | rs148027154 | 3 prime UTR variant | - | - | *PPIB* is involved in procollagen export and secretion[70]. Certain *PPIB* variants have been found to be associated with osteogenesis imperfecta[71]. |
| ***SMAD3*** | rs4776880 | Intron | - | Sciatica, hallux valgus, lumbar disc herniation, spondylosis, cervical disc disorders, coxarthrosis, spondylolisthesis | - |
| ***PDE8A*** | rs11629962 | Intron | - | - | *PDE8A* has a role in T-cell activation[72], and through the cAMP pathway, it participates in the regulation of many biological pathways[73]. |
| ***GAN*** | rs2966084 | Intron | - | Hypertension, type II diabetes | Mutations in the *GAN* gene cause giant axonal neuropathy, a severe form of sensory and motor neuropathy. Axonal degeneration is accompanied by the accumulation and aggregation of cytoskeletal components. According to these findings, *GAN* plays a critical role in axonal structure and neuron maintenance[74]. |
| ***SOCS7*** | rs113353816 | Intron | - | - | *SOCS7* regulates a variety of signalling pathways[75]. |
| ***AXIN2*** | rs16961974 | Regulatory region variant | Osteoporosis | - | *AXIN2* is most likely engaged in controlling β-catenin within the Wnt-signaling pathway[76]. The Wnt-signaling pathway is linked to autoimmunity, bone and cartilage regeneration, and repair[77]. |
| ***Empty*** | rs12941198 | Intergenic | - | - | - |
| ***DCC*** | rs6508210 | Intron | Sciatica, obesity, metabolic disorders, depression, any mental disorder, pain, low back pain, spondylosis, depression medications, lumbar disc herniation | - | *DCC* might have a role in neurovascular ingrowth/axonal guidance[78,79]. |

| | | | | | |
|---|---|---|---|---|---|
| ***CACTIN*** | rs77733715 | 3 prime UTR variant | Malignant melanoma of skin | - | *CACTIN* has been associated with longitudinal pediatric bone accrual, it has been annotated to the toll-like receptor pathway which has a role in osteoblast function, including mediating bone inflammatory responses and regulating cell viability, proliferation, and osteoblast-mediated osteoclastogenesis[80]. |
| ***CRLF1*** | rs2074177 | Intron | - | - | One frequent cause of LSS is ligamentum flavum hypertrophy[19]. Through the ERK signalling pathway, *CRLF1* increases LF fibrosis and plays a part in mediating the effects of TGF-β1[81]. |
| ***DDRGK1*** | rs11697820 | Upstream gene variant | - | - | Potentially acting as an NF-κB regulator[82], *DDRGK1* can also bind directly to *SOX9*, preventing its ubiquitination and proteasomal degradation, which can affect the chondrogenesis[83]. |
| ***ETS2*** | rs111901809 | Upstream gene variant | - | Atrial fibrillation and flutter, cardiac arrhythmias | Through osteoblast differentiation and bone formation, *ETS2* potentially plays a role in skeletal development[84]. |
| ***OSM*** | rs2285645 | 3 prime UTR variant | - | - | *OSM* is a growth and cytokine production regulator that has been found to have the ability to stimulate bone accrual[85,86]. |
| ***CHRDL1*** | rs67648651 | Intron | Carpal tunnel syndrome, lumbar disc herniation | Coronary revascularization, hernia, hypercholesterolemia, statin medication, metabolic disorders, | Through its potential to affect the activity of the BMP- and SMAD-signalling pathways, *CHRDL1* may play a significant role in osteogenic differentiation, bone remodelling and neuronal differentation[87,88]. |
| ***Surgical GWAS (FinnGen)*** | | | | | |
| ***FOXO1*** | rs12872985 | Intron | Leiomyoma of uterus, lumbar disc herniation | - | In addition to being required for *SOX9* expression, *FOXO1* also plays a crucial role in cell-cycle arrest during chondrogenic differentiation through TGFβ1 signalling[89]. It also plays a role in B-cell differentiation[30]. |

Candidate gene, a gene at a novel locus which biological function is likely to explain the LSS or LSS surgical GWAS association; rsid, SNP markers identification number, Variant type, genomic location of associated variant, PheWAS results for the LSS-associated variants, with an arrow showing the direction of the association. Only associations that reached genome-wide significance ($p<5x10^{-8}$) are reported.

**Table S4.** The lead variants at 54 LSS-associated ($p<5x10^{-8}$) loci in the meta-analysis of 30 269 LSS cases and 739 414 controls from FinnGen, the Estonian Biobank, and the UK Biobank. Odds ratios and their 95% confidence intervals for each LSS-associated variant both in the meta-analysis and in each dataset separately. EAF, effect allele frequency.

| Candidate gene | CHR:POS | EA | OA | Cohort | OR (95 % CI) | EAF |
|---|---|---|---|---|---|---|
| ***RUNX3*** | **1:24894533** | **G** | **A** | **Meta-analysis** | **0.90 (0.88-0.92)** | **0.31** |
| | | | | FinnGen | 0.94 (0.91-0.96) | 0.30 |
| | | | | UK Biobank | 1.01 (0.97-1.05) | 0.32 |
| | | | | Estonian Biobank | 0.92 (0.87-0.97) | 0.32 |
| ***NGF*** | **1:115287365** | **T** | **C** | **Meta-analysis** | **0.93 (0.91-0.95)** | **0.47** |
| | | | | FinnGen | 0.93 (0.91–0.96) | 0.47 |
| | | | | UK Biobank | 0.92 (0.88-0.96) | 0.46 |
| | | | | Estonian Biobank | 0.96 (0.92-1.01) | 0.47 |
| ***OTUD7B*** | **1:149934520** | **C** | **T** | **Meta-analysis** | **0.92 (0.90-0.94)** | **0.38** |
| | | | | FinnGen | 0.91 (0.89-0.94) | 0.33 |
| | | | | UK Biobank | 0.92 (0.88-0.96) | 0.41 |
| | | | | Estonian Biobank | 0.93 (0.89-0.98) | 0.36 |
| ***ZNF687*** | **1:151032308** | **T** | **C** | **Meta-analysis** | **1.21 (1.14-1.27)** | **0.01** |
| | | | | FinnGen | 1.20 (1.13-1.26) | 0.03 |
| | | | | UK Biobank | 1.71 (0.22-3.21) | 0.0002 |
| | | | | Estonian Biobank | 1.34 (1.05-1.62) | 0.006 |
| ***PTPRC*** | **1:198819354** | **C** | **G** | **Meta-analysis** | **0.91 (0.88-0.93)** | **0.15** |
| | | | | FinnGen | 0.90 (0.87-0.93) | 0.16 |
| | | | | UK Biobank | 0.93 (0.87-0.99) | 0.13 |
| | | | | Estonian Biobank | 0.90 (0.85-0.96) | 0.20 |
| ***GFPT1*** | **2:69341550** | **G** | **A** | **Meta-analysis** | **0.91 (0.89-0.93)** | **0.57** |
| | | | | FinnGen | 0.92 (0.89-0.94) | 0.55 |
| | | | | UK Biobank | 0.91 (0.87-0.95) | 0.58 |
| | | | | Estonian Biobank | 0.90 (0.86-0.95) | 0.58 |
| ***USP37*** | **2:218486683** | **G** | **C** | **Meta-analysis** | **0.94 (0.92-0.96)** | **0.61** |
| | | | | FinnGen | 0.94 (0.92-0.96) | 0.61 |
| | | | | UK Biobank | - | - |
| | | | | Estonian Biobank | 0.95 (0.91-1.00) | 0.62 |
| ***DAG1*** | **3:49498696** | **TTTA** | **T** | **Meta-analysis** | **0.95 (0.93-0.97)** | **0.35** |
| | | | | FinnGen | 0.95 (0.92-0.97) | 0.44 |
| | | | | UK Biobank | 0.95 (0.91-1.00) | 0.32 |
| | | | | Estonian Biobank | 0.96 (0.91-1.01) | 0.30 |
| ***RAB28*** | **4:12893286** | **A** | **AGAG** | **Meta-analysis** | **0.92 (0.90-0.94)** | **0.24** |
| | | | | FinnGen | 0.92 (0.89-0.94) | 0.25 |
| | | | | UK Biobank | 0.91 (0.86-0.95) | 0.24 |
| | | | | Estonian Biobank | 0.95 (0.90-1.00) | 0.23 |
| ***DAB2*** | **5:39844572** | **ATATG** | **A** | **Meta-analysis** | **1.08 (1.05-1.10)** | **0.17** |
| | | | | FinnGen | 1.09 (1.06-1.11) | 0.19 |
| | | | | UK Biobank | 1.05 (0.99-1.10) | 0.16 |
| | | | | Estonian Biobank | - | - |
| ***ITGA2*** | **5:53038840** | **A** | **C** | **Meta-analysis** | **1.08 (1.05-1.10)** | **0.18** |
| | | | | FinnGen | 1.07 (1.04-1.10) | 0.19 |
| | | | | UK Biobank | 1.10 (1.05-1.15) | 0.18 |
| | | | | Estonian Biobank | 1.08 (1.02-1.14) | 0.16 |
| ***EBF1*** | **5:159017122** | **G** | **GC** | **Meta-analysis** | **1.06 (1.04-1.09)** | **0.22** |
| | | | | FinnGen | 1.06 (1.03-1.09) | 0.17 |
| | | | | UK Biobank | 1.07 (1.02-1.12) | 0.24 |
| | | | | Estonian Biobank | 1.06 (1.01-1.11) | 0.27 |
| ***BMP6*** | **6:7797607** | **G** | **C** | **Meta-analysis** | **1.08 (1.06-1.11)** | **0.20** |
| | | | | FinnGen | 1.09 (1.06-1.11) | 0.22 |
| | | | | UK Biobank | 1.09 (1.04-1.14) | 0.18 |
| | | | | Estonian Biobank | 1.07 (1.01-1.12) | 0.19 |
| ***TRIM38*** | **6:26239176** | **G** | **A** | **Meta-analysis** | **0.94 (0.93-0.96)** | **0.32** |
| | | | | FinnGen | 0.94 (0.92-0.96) | 0.41 |
| | | | | UK Biobank | 0.94 (0.89-0.98) | 0.26 |
| | | | | Estonian Biobank | 0.97 (0.93-1.02) | 0.34 |
| ***HLA*** | **6:30106665** | **A** | **T** | **Meta-analysis** | **0.94 (0.92-0.96)** | **0.18** |
| | | | | FinnGen | 0.93 (0.90-0.95) | 0.23 |
| | | | | UK Biobank | 0.95 (0.89-1.01) | 0.14 |
| | | | | Estonian Biobank | 1.00 (0.94-1.06) | 0.16 |
| ***HLA*** | **6:31133806** | **A** | **G** | **Meta-analysis** | **0.91 (0.88-0.93)** | **0.11** |
| | | | | FinnGen | 0.89 (0.86-0.92) | 0.16 |
| | | | | UK Biobank | 0.95 (0.87-1.02) | 0.07 |
| | | | | Estonian Biobank | 0.95 (0.87-1.02) | 0.10 |
| ***HLA*** | **6:32634706** | **T** | **G** | **Meta-analysis** | **0.92 (0.90-0.94)** | **0.20** |
| | | | | FinnGen | 0.92 (0.89-0.94) | 0.24 |
| | | | | UK Biobank | 0.93 (0.88-0.98) | 0.18 |
| | | | | Estonian Biobank | - | - |
| ***CDC5L*** | **6:44479182** | **A** | **C** | **Meta-analysis** | **1.07 (1.05-1.08)** | **0.53** |
| | | | | FinnGen | 1.06 (1.04-1.08) | 0.53 |
| | | | | UK Biobank | 1.10 (1.06-1.14) | 0.53 |
| | | | | Estonian Biobank | 1.05 (1.01-1.10) | 0.53 |
| ***LIN28B*** | **6:105005542** | **C** | **A** | **Meta-analysis** | **1.05 (1.03-1.07)** | **0.57** |
| | | | | FinnGen | 1.05 (1.03-1.07) | 0.59 |
| | | | | UK Biobank | 1.07 (1.03-1.11) | 0.56 |
| | | | | Estonian Biobank | 1.03 (0.99-1.08) | 0.55 |

| | | | | | | |
|---|---|---|---|---|---|---|
| ***COL10A1*** | **6:116167743** | **G** | **A** | **Meta-analysis** | **0.95 (0.93-0.97)** | **0.51** |
| | | | | FinnGen | 0.95 (0.92-0.97) | 0.50 |
| | | | | UK Biobank | 0.94 (0.90-0.98) | 0.53 |
| | | | | Estonian Biobank | 0.98 (0.93-1.02) | 0.47 |
| ***EYA4*** | **6:132913577** | **A** | **G** | **Meta-analysis** | **1.06 (1.05-1.08)** | **0.43** |
| | | | | FinnGen | 1.07 (1.04-1.09) | 0.41 |
| | | | | UK Biobank | 1.06 (1.02-1.10) | 0.44 |
| | | | | Estonian Biobank | - | - |
| ***FOXK1*** | **7:4638590** | **C** | **T** | **Meta-analysis** | **0.93 (0.91-0.96)** | **0.19** |
| | | | | FinnGen | 0.94 (0.91-0.96) | 0.23 |
| | | | | UK Biobank | 0.92 (0.86-0.97) | 0.17 |
| | | | | Estonian Biobank | - | - |
| ***AGMO*** | **7:15344063** | **G** | **A** | **Meta-analysis** | **1.09 (1.06-1.11)** | **0.13** |
| | | | | FinnGen | 1.11 (1.08-1.13) | 0.17 |
| | | | | UK Biobank | 1.06 (1.00-1.13) | 0.11 |
| | | | | Estonian Biobank | 1.01 (0.94-1.08) | 0.11 |
| ***TWIST1*** | **7:19488615** | **T** | **C** | **Meta-analysis** | **0.95 (0.93-0.97)** | **0.49** |
| | | | | FinnGen | 0.95 (0.93-0.97) | 0.48 |
| | | | | UK Biobank | 0.93 (0.89-0.97) | 0.50 |
| | | | | Estonian Biobank | 0.97 (0.93-1.02) | 0.49 |
| ***JAZF1*** | **7:28156794** | **A** | **G** | **Meta-analysis** | **0.95 (0.93-0.97)** | **0.49** |
| | | | | FinnGen | 0.96 (0.94-0.98) | 0.47 |
| | | | | UK Biobank | 0.94 (.90-0.98) | 0.50 |
| | | | | Estonian Biobank | 0.93 (0.89-0.98) | 0.50 |
| ***Empty*** | **7:69064281** | **G** | **GT** | **Meta-analysis** | **1.08 (1.05-1.11)** | **0.15** |
| | | | | FinnGen | 1.08 (1.05-1.11) | 0.16 |
| | | | | UK Biobank | 1.09 (1.03-1.15) | 0.14 |
| | | | | Estonian Biobank | - | - |
| ***DYNC1I1*** | **7:95941237** | **C** | **T** | **Meta-analysis** | **1.08 (1.06-1.10)** | **0.75** |
| | | | | FinnGen | 1.08 (1.05-1.10) | 0.77 |
| | | | | UK Biobank | 1.10 (1.06-1.15) | 0.74 |
| | | | | Estonian Biobank | - | - |
| ***MKX*** | **10:27612430** | **T** | **G** | **Meta-analysis** | **1.06 (1.05-1.08)** | **0.47** |
| | | | | FinnGen | 1.05 (1.03-1.08) | 0.40 |
| | | | | UK Biobank | 1.06 (1.02-1.10) | 0.51 |
| | | | | Estonian Biobank | 1.11 (1.07-1.16) | 0.49 |
| ***MPPED2*** | **11:30788512** | **C** | **T** | **Meta-analysis** | **1.05 (1.04-1.07)** | **0.44** |
| | | | | FinnGen | 1.06 (1.04-1.08) | 0.48 |
| | | | | UK Biobank | 1.04 (1.00-1.08) | 0.42 |
| | | | | Estonian Biobank | 1.06 (1.10-1.10) | 0.43 |
| ***PDE3A*** | **12:20427791** | **A** | **T** | **Meta-analysis** | **0.93 (0.91-0.95)** | **0.23** |
| | | | | FinnGen | 0.93 (0.90-0.95) | 0.26 |
| | | | | UK Biobank | 0.91 (0.86-0.95) | 0.21 |
| | | | | Estonian Biobank | 0.94 (0.90-0.99) | 0.31 |
| ***SOX5*** | **12:23821470** | **C** | **G** | **Meta-analysis** | **1.08 (1.06-1.10)** | **0.27** |
| | | | | FinnGen | 1.07 (1.05-1.10) | 0.31 |
| | | | | UK Biobank | 1.08 (1.03-1.12) | 0.23 |
| | | | | Estonian Biobank | 1.10 (1.05-1.15) | 0.32 |
| ***CCDC91*** | **12:27844985** | **T** | **C** | **Meta-analysis** | **0.93 (0.91-0.95)** | **0.35** |
| | | | | FinnGen | 0.93 (0.91-0.95) | 0.31 |
| | | | | UK Biobank | 0.94 (0.90-0.98) | 0.38 |
| | | | | Estonian Biobank | 0.93 (0.89-0.98) | 0.39 |
| ***GLI1*** | **12:57749071** | **A** | **C** | **Meta-analysis** | **0.94 (0.93-0.96)** | **0.33** |
| | | | | FinnGen | 0.92 (0.88-0.95) | 0.34 |
| | | | | UK Biobank | 0.97 (0.91-1.02) | 0.32 |
| | | | | Estonian Biobank | 0.94 (0.88-1.01) | 0.36 |
| ***USP15*** | **12:62218131** | **A** | **G** | **Meta-analysis** | **1.07 (1.05-1.10)** | **0.17** |
| | | | | FinnGen | 1.07 (1.04-1.10) | 0.17 |
| | | | | UK Biobank | 1.10 (1.04-1.15) | 0.16 |
| | | | | Estonian Biobank | 1.07 (1.01-1.13) | 0.18 |
| ***ZCCHC8*** | **12:122480161** | **C** | **T** | **Meta-analysis** | **1.06 (1.04-1.09)** | **0.77** |
| | | | | FinnGen | 1.05 (1.02-1.08) | 0.80 |
| | | | | UK Biobank | 1.09 (1.04-1.13) | 0.74 |
| | | | | Estonian Biobank | 1.06 (1.01-1.12) | 0.76 |
| ***TNFSF11*** | **13:42423131** | **T** | **C** | **Meta-analysis** | **0.95 (0.93-0.97)** | **0.25** |
| | | | | FinnGen | 0.94 (0.92-0.96) | 0.27 |
| | | | | UK Biobank | 0.93 (0.88-0.97) | 0.23 |
| | | | | Estonian Biobank | 0.99 (0.94-1.04) | 0.27 |
| ***SLITRK6*** | **13:85356178** | **G** | **C** | **Meta-analysis** | **0.94 (0.93-0.96)** | **0.59** |
| | | | | FinnGen | 0.94 (0.92-0.96) | 0.59 |
| | | | | UK Biobank | 0.93 (0.89-0.97) | 0.60 |
| | | | | Estonian Biobank | 0.97 (0.93-1.02) | 0.54 |
| ***MMP14*** | **14:22843385** | **A** | **G** | **Meta-analysis** | **0.93 (0.91-0.96)** | **0.21** |
| | | | | FinnGen | 0.93 (0.90-0.96) | 0.21 |
| | | | | UK Biobank | 0.94 (0.89-0.99) | 0.21 |
| | | | | Estonian Biobank | 0.95 (0.90-1.00) | 0.21 |
| ***GPR132*** | **14:105137078** | **A** | **C** | **Meta-analysis** | **0.91 (0.88-0.95)** | **0.09** |
| | | | | FinnGen | 0.90 (0.86-0.94) | 0.07 |
| | | | | UK Biobank | 0.93 (0.87-1.00) | 0.11 |
| | | | | Estonian Biobank | 0.92 (0.84-1.01) | 0.09 |
| ***ALDH1A2*** | **15:58050948** | **C** | **G** | **Meta-analysis** | **1.06 (1.05-1.08)** | **0.49** |

| | | | | | | |
|---|---|---|---|---|---|---|
| | | | | FinnGen | 1.07 (1.04-1.09) | 0.49 |
| | | | | UK Biobank | 1.06 (1.02-1.10) | 0.49 |
| | | | | Estonian Biobank | 1.05 (1.01-1.10) | 0.47 |
| ***PPIB*** | **15:64138226** | **A** | **T** | **Meta-analysis** | **1.28 (1.20-1.37)** | **0.006** |
| | | | | FinnGen | 1.27 (1.18-1.36) | 0.01 |
| | | | | UK Biobank | 2.12 (1.02-1.10) | 0.002 |
| | | | | Estonian Biobank | 1.22 (0.80-1.64) | 0.003 |
| ***SMAD3*** | **15:67072653** | **A** | **G** | **Meta-analysis** | **0.90 (0.88-0.92)** | **0.35** |
| | | | | FinnGen | 0.91 (0.89-0.93) | 0.37 |
| | | | | UK Biobank | 0.88 (0.84-0.92) | 0.33 |
| | | | | Estonian Biobank | 0.89 (0.85-0.94) | 0.41 |
| ***PDE8A*** | **15:84986797** | **T** | **C** | **Meta-analysis** | **0.94 (0.92-0.96)** | **0.25** |
| | | | | FinnGen | 0.93 (0.91-0.96) | 0.27 |
| | | | | UK Biobank | 0.96 (0.92-1.01) | 0.24 |
| | | | | Estonian Biobank | - | - |
| ***GAN*** | **16:81482778** | **G** | **A** | **Meta-analysis** | **0.94 (0.92-0.96)** | **0.69** |
| | | | | FinnGen | 0.94 (0.92-0.96) | 0.67 |
| | | | | UK Biobank | 0.93 (0.88-0.97) | 0.71 |
| | | | | Estonian Biobank | 0.95 (0.90-1.00) | 0.72 |
| ***SOCS7*** | **17:38474030** | **C** | **T** | **Meta-analysis** | **1.06 (1.04-1.09)** | **0.83** |
| | | | | FinnGen | 1.08 (1.05-1.10) | 0.79 |
| | | | | UK Biobank | 1.02 (0.97-1.08) | 0.86 |
| | | | | Estonian Biobank | 1.06 (1.00-1.12) | 0.81 |
| ***AXIN2*** | **17:65278900** | **C** | **T** | **Meta-analysis** | **1.06 (1.04-1.07)** | **0.28** |
| | | | | FinnGen | 1.06 (1.03-1.08) | 0.29 |
| | | | | UK Biobank | 1.03 (0.98-1.08) | 0.26 |
| | | | | Estonian Biobank | 1.07 (1.03-1.12) | 0.36 |
| ***Empty*** | **17:71476022** | **C** | **A** | **Meta-analysis** | **1.06 (1.04-1.08)** | **0.78** |
| | | | | FinnGen | 1.06 (1.03-1.09) | 0.78 |
| | | | | UK Biobank | 1.09 (1.05-1.14) | 0.78 |
| | | | | Estonian Biobank | 1.03 (0.98-1.08) | 0.78 |
| ***DCC*** | **18:53220821** | **C** | **A** | **Meta-analysis** | **1.08 (1.06-1.09)** | **0.52** |
| | | | | FinnGen | 1.08 (1.06-1.10) | 0.49 |
| | | | | UK Biobank | 1.07 (1.03-1.11) | 0.53 |
| | | | | Estonian Biobank | 1.05 (1.01-1.10) | 0.56 |
| ***CACTIN*** | **19:3537186** | **G** | **A** | **Meta-analysis** | **1.14 (1.09-1.18)** | **0.03** |
| | | | | FinnGen | 1.15 (1.10-1.19) | 0.05 |
| | | | | UK Biobank | 1.22 (1.00-1.43) | 0.009 |
| | | | | Estonian Biobank | 1.06 (0.92-1.19) | 0.03 |
| ***CRLF1*** | **19:18616479** | **G** | **A** | **Meta-analysis** | **0.93 (0.91-0.95)** | **0.25** |
| | | | | FinnGen | 0.93 (0.90-0.95) | 0.25 |
| | | | | UK Biobank | 0.91 (0.86-0.96) | 0.24 |
| | | | | Estonian Biobank | 0.96 (0.90-1.01) | 0.26 |
| ***DDRGK1*** | **20:2800795** | **T** | **C** | **Meta-analysis** | **0.95 (0.93-0.96)** | **0.58** |
| | | | | FinnGen | 0.95 (0.93-0.97) | 0.58 |
| | | | | UK Biobank | 0.92 (0.88-0.96) | 0.59 |
| | | | | Estonian Biobank | 0.96 (0.91-1.00) | 0.56 |
| ***ETS2*** | **21:39183865** | **T** | **TAGAGGGGCA GGGGGCG** | **Meta-analysis** | **1.05 (1.03-1.07)** | **0.49** |
| | | | | FinnGen | 1.04 (1.02-1.07) | 0.56 |
| | | | | UK Biobank | 1.09 (1.05-1.13) | 0.44 |
| | | | | Estonian Biobank | 1.04 (0.99-1.08) | 0.53 |
| ***OSM*** | **22:30026719** | **T** | **C** | **Meta-analysis** | **0.83 (0.78-0.89)** | **0.02** |
| | | | | FinnGen | 0.83 (0.78-0.89) | 0.05 |
| | | | | UK Biobank | 0.76 (0.15-1.37) | 0.001 |
| | | | | Estonian Biobank | 0.84 (0.68-1.01) | 0.02 |
| ***CHRDL1*** | **X:110450046** | **C** | **T** | **Meta-analysis** | **1.05 (1.04-1.07)** | **0.40** |
| | | | | FinnGen | 1.05 (1.03-1.06) | 0.38 |
| | | | | UK Biobank | 1.07 (1.04-1.10) | 0.41 |
| | | | | Estonian Biobank | 1.06 (1.02-1.10) | 0.39 |

**Table S5.** Meta-analysis and surgical GWAS effect differences for every LSS-associated variant. Two-tailed test was used for statistical testing, for that we used following formula: *((Effect_Meta-Effect_SURG)/sqrt(standarderror_Meta$^2$+standarderror_SURG$^2$)), P_diff=2*(1-dif)*. $P_{diff}$ <0.05 was considered as a threshold for statistical significance.

| Candidate gene | rsid | Effect Meta | Effect SURG | Standard error Meta | Standard error SURG | P_diff |
|---|---|---|---|---|---|---|
| *RUNX3* | rs9438857 | -0,053 | -0,086 | 0,010 | 0,017 | 0,101 |
| ***NGF*** | **rs6327** | **-0,072** | **-0,109** | **0,009** | **0,016** | **0,042** |
| *OTUD7B* | rs11205303 | -0,085 | -0,118 | 0,009 | 0,017 | 0,100 |
| *ZNF687* | rs147042690 | 0,188 | 0,211 | 0,033 | 0,047 | 0,691 |
| *PTPRC* | rs72739729 | -0,097 | -0,121 | 0,012 | 0,022 | 0,337 |
| *GFPT1* | rs4340572 | -0,092 | -0,109 | 0,009 | 0,016 | 0,337 |
| *USP37* | rs60362609 | -0,057 | -0,063 | 0,010 | 0,016 | 0,771 |
| *DAG1* | rs138073019 | -0,053 | -0,062 | 0,009 | 0,016 | 0,629 |
| ***RAB28*** | **rs5856174** | **-0,082** | **-0,125** | **0,010** | **0,019** | **0,043** |
| *ITGA2* | rs148346348 | 0,076 | 0,095 | 0,012 | 0,020 | 0,431 |
| *DAB2* | rs11747379 | 0,074 | 0,079 | 0,011 | 0,020 | 0,805 |
| *EBF1* | rs200608668 | 0,062 | 0,070 | 0,011 | 0,021 | 0,707 |
| ***BMP6*** | **rs10498672** | **0,081** | **0,143** | **0,011** | **0,019** | **0,004** |
| ***TRIM38*** | **rs9358913** | **-0,057** | **-0,093** | **0,009** | **0,016** | **0,049** |
| *HLA* | rs62390186 | -0,062 | -0,100 | 0,011 | 0,019 | 0,085 |
| *HLA* | rs4294047 | -0,097 | -0,112 | 0,013 | 0,022 | 0,573 |
| *HLA* | rs17205170 | -0,085 | -0,086 | 0,012 | 0,019 | 0,953 |
| *CDC5L* | rs7770012 | 0,064 | 0,082 | 0,009 | 0,016 | 0,305 |
| *LIN28B* | rs314261 | 0,050 | 0,061 | 0,009 | 0,016 | 0,576 |
| *COL10A1* | rs7739896 | -0,052 | -0,082 | 0,009 | 0,016 | 0,095 |
| *EYA4* | rs2764217 | 0,063 | 0,068 | 0,010 | 0,016 | 0,781 |
| *FOKX1* | rs28658109 | -0,068 | -0,058 | 0,012 | 0,019 | 0,651 |
| ***AGMO*** | **rs75674922** | **0,085** | **0,135** | **0,012** | **0,020** | **0,038** |
| *TWIST1* | rs2390116 | -0,050 | -0,066 | 0,009 | 0,016 | 0,364 |
| *JAZF1* | rs849135 | -0,052 | -0,060 | 0,009 | 0,016 | 0,658 |
| *Empty* | rs139203647 | 0,076 | 0,090 | 0,013 | 0,021 | 0,579 |
| *DYNC1I1* | rs11769842 | 0,079 | 0,079 | 0,012 | 0,019 | 0,999 |
| *MKX* | rs2637326 | 0,063 | 0,062 | 0,009 | 0,016 | 0,992 |
| *MPPED2* | rs607165 | 0,053 | 0,048 | 0,009 | 0,016 | 0,748 |
| *PDE3A* | rs73067286 | -0,076 | -0,086 | 0,010 | 0,018 | 0,609 |
| *SOX5* | rs12308843 | 0,077 | 0,057 | 0,010 | 0,017 | 0,308 |
| *CCDC91* | rs7958415 | -0,071 | -0,084 | 0,010 | 0,017 | 0,493 |
| *GLI1* | rs2069506 | -0,057 | -0,093 | 0,009 | 0,017 | 0,057 |
| *USP15* | rs11174377 | 0,070 | 0,070 | 0,012 | 0,021 | 1,000 |
| *ZCCHC8* | rs10846978 | 0,059 | 0,039 | 0,011 | 0,020 | 0,358 |
| ***TNFSF11*** | **rs9594751** | **-0,056** | **-0,100** | **0,010** | **0,018** | **0,037** |
| *LINC00351* | rs9531779 | -0,058 | -0,070 | 0,009 | 0,016 | 0,518 |
| *MMP14* | rs1042704 | -0,068 | -0,079 | 0,011 | 0,020 | 0,619 |
| *GPR132* | rs1022431 | -0,091 | -0,139 | 0,017 | 0,031 | 0,166 |
| ***ALDH1A2*** | **rs4646570** | **0,061** | **0,104** | **0,009** | **0,016** | **0,018** |
| *PPIB* | rs148027154 | 0,250 | 0,262 | 0,045 | 0,067 | 0,884 |
| *SMAD3* | rs4776880 | -0,103 | -0,100 | 0,009 | 0,016 | 0,860 |
| *PDE8A* | rs11629962 | -0,062 | -0,089 | 0,011 | 0,018 | 0,190 |
| *GAN* | rs2966084 | -0,063 | -0,068 | 0,010 | 0,017 | 0,813 |
| *SOCS7* | rs113353816 | 0,063 | 0,081 | 0,011 | 0,020 | 0,425 |
| *AXIN2* | rs16961974 | 0,054 | 0,063 | 0,010 | 0,017 | 0,651 |
| *Empty* | rs12941198 | 0,059 | 0,053 | 0,011 | 0,019 | 0,780 |
| *DCC* | rs6508210 | 0,073 | 0,075 | 0,009 | 0,016 | 0,926 |
| *CACTIN* | rs77733715 | 0,130 | 0,187 | 0,023 | 0,035 | 0,171 |
| *CRLF1* | rs2074177 | -0,074 | -0,092 | 0,010 | 0,019 | 0,407 |
| *DDRGK1* | rs11697820 | -0,056 | -0,054 | 0,009 | 0,016 | 0,920 |
| *ETS2* | rs111901809 | 0,049 | 0,042 | 0,009 | 0,016 | 0,687 |
| *OSM* | rs2285645 | -0,181 | -0,244 | 0,026 | 0,040 | 0,192 |
| *CHRDL1* | rs67648651 | 0,051 | 0,060 | 0,008 | 0,013 | 0,564 |

**Table S6.** FastBAT gene prioritisation results, that we obtained from analysis performed with GCTA software[90]. For loci we used 2MB window and for LD reference we used FinnGen. We considered results with $p_{FASTBAT}<5x10^{-8}$ and $p_{GWAS}<5x10^{-8}$ significant[91].

| Chr | Gene | pFASTBAT | pGWAS | TopSNP |
|---|---|---|---|---|
| 1 | FAM72C | 1.80416e-08 | 1.76e-19 | chr1_149934520_T_C |
| 1 | MTMR11 | 4.79029e-11 | 4.54e-17 | chr1_149920979_A_G |
| 1 | OTUD7B | 7.38031e-17 | 1.76e-19 | chr1_149934520_T_C |
| 1 | SF3B4 | 1.83939e-11 | 4.54e-17 | chr1_149920979_A_G |
| 1 | SRGAP2D | 1.76634e-08 | 1.76e-19 | chr1_149934520_T_C |
| 1 | VPS45 | 8.59157e-12 | 1.51e-18 | chr1_150023307_G_A |
| 1 | FAM72C | 4.04694e-11 | 2.18e-15 | chr1_198819354_G_C |
| 1 | SRGAP2D | 4.04694e-11 | 2.18e-15 | chr1_198819354_G_C |
| 2 | ANTXR1 | 1.09116e-15 | 3.45e-21 | chr2_69418633_C_A |
| 2 | ARHGAP25 | 5.51267e-14 | 7.86e-25 | chr2_69322904_T_C |
| 2 | BMP10 | 1.80052e-21 | 5.64e-25 | chr2_69341550_A_G |
| 2 | GKN1 | 7.54269e-18 | 1.5e-21 | chr2_69406431_G_GA |
| 2 | GKN2 | 2.23381e-20 | 6.31e-25 | chr2_69352523_C_G |
| 2 | ANTXR1 | 6.32123e-15 | 1.99e-19 | chr2_69433082_G_A |
| 2 | GKN1 | 2.75363e-17 | 1.99e-19 | chr2_69433082_G_A |
| 2 | GKN2 | 1.58446e-15 | 1.99e-19 | chr2_69433082_G_A |
| 3 | AMT | 1.5124e-08 | 9.31e-09 | chr3_49498696_T_TTTA |
| 3 | BSN | 4.84111e-09 | 1.08e-08 | chr3_49629531_C_A |
| 3 | DAG1 | 9.3841e-09 | 9.31e-09 | chr3_49498696_T_TTTA |
| 3 | GPX1 | 4.34351e-09 | 1.21e-08 | chr3_49408664_G_GTATA |
| 3 | NICN1 | 7.8823e-09 | 9.31e-09 | chr3_49498696_T_TTTA |
| 3 | RHOA | 3.89366e-09 | 9.31e-09 | chr3_49498696_T_TTTA |
| 3 | TCTA | 5.76385e-09 | 9.31e-09 | chr3_49498696_T_TTTA |
| 6 | MOXD1 | 2.50626e-09 | 1.36e-10 | chr6_132913577_G_A |
| 6 | BTN3A2 | 4.16556e-08 | 3.27e-09 | chr6_26353699_A_G |
| 6 | BMP6 | 8.56593e-11 | 9.09e-14 | chr6_7797607_C_G |
| 7 | AGMO | 1.50745e-11 | 6.72e-12 | chr7_15344063_A_G |
| 7 | JAZF1 | 4.21504e-08 | 7.32e-09 | chr7_28156794_G_A |
| 7 | SHFM1 | 2.66193e-09 | 1.18e-11 | chr7_95941237_T_C |
| 11 | DCDC5 | 4.8859e-08 | 3.23e-09 | chr11_30877762_C_T |
| 12 | KDM2B | 4.68808e-08 | 4.49e-08 | chr12_122480161_T_C |
| 12 | SOX5 | 3.0539e-10 | 3.3e-15 | chr12_23821470_G_C |
| 12 | KLHL42 | 1.78641e-09 | 8.41e-12 | chr12_28109762_G_GT |
| 12 | MANSC4 | 1.97722e-09 | 8.41e-12 | chr12_28109762_G_GT |
| 12 | MRPS35 | 7.05575e-09 | 8.41e-12 | chr12_28109762_G_GT |
| 12 | PPFIBP1 | 3.18281e-11 | 1.07e-13 | chr12_27844985_C_T |
| 12 | SMCO2 | 3.30074e-08 | 1.07e-13 | chr12_27844985_C_T |
| 12 | KLHL42 | 1.78641e-09 | 8.41e-12 | chr12_28109762_G_GT |
| 12 | MANSC4 | 1.97722e-09 | 8.41e-12 | chr12_28109762_G_GT |
| 12 | MRPS35 | 7.05575e-09 | 8.41e-12 | chr12_28109762_G_GT |
| 12 | HSD17B6 | 2.42438e-08 | 2.59e-08 | chr12_57608816_C_T |
| 12 | MYO1A | 5.15097e-09 | 7.37e-09 | chr12_57768302_G_T |
| 12 | RDH16 | 4.13935e-08 | 1.56e-09 | chr12_57749071_C_A |
| 12 | SDR9C7 | 2.48397e-08 | 1.56e-09 | chr12_57749071_C_A |
| 12 | TAC3 | 8.61341e-09 | 1.56e-09 | chr12_57749071_C_A |
| 12 | ZBTB39 | 2.4375e-08 | 1.56e-09 | chr12_57749071_C_A |
| 15 | CGNL1 | 3.63258e-09 | 5.47e-12 | chr15_58050948_G_C |
| 15 | LCTL | 7.2707e-10 | 4.6e-13 | chr15_67085695_A_G |
| 15 | MAP2K1 | 1.50604e-23 | 1.05e-28 | chr15_67072653_G_A |
| 15 | RPL4 | 1.98261e-30 | 1.05e-28 | chr15_67072653_G_A |
| 15 | SCARNA14 | 2.38228e-08 | 2.38e-10 | chr15_66975565_CG_C |
| 15 | SNAPC5 | 2.68647e-29 | 1.05e-28 | chr15_67072653_G_A |
| 15 | TIPIN | 1.89533e-08 | 6.13e-11 | chr15_66991209_C_A |
| 15 | ZWILCH | 5.50651e-22 | 1.05e-28 | chr15_67072653_G_A |
| 17 | CASKIN2 | 1.82154e-09 | 3.6e-08 | chr17_71476022_A_C |
| 17 | LLGL2 | 1.12465e-09 | 3.6e-08 | chr17_71476022_A_C |
| 17 | TSEN54 | 2.68638e-09 | 3.6e-08 | chr17_71476022_A_C |
| 18 | ATP8B1 | 6.87157e-10 | 3.76e-14 | chr18_53170750_C_T |
| 18 | NEDD4L | 4.54904e-10 | 7.36e-14 | chr18_53344021_T_A |
| 19 | IFI30 | 4.87103e-08 | 2.98e-08 | chr19_18381183_G_A |
| 19 | LSM4 | 1.28778e-09 | 2.19e-10 | chr19_18591691_G_A |
| 19 | MPV17L2 | 1.31597e-08 | 2.98e-08 | chr19_18381183_G_A |
| 19 | PDE4C | 8.52371e-09 | 2.98e-08 | chr19_18381183_G_A |
| 19 | PGPEP1 | 5.8441e-09 | 1.03e-12 | chr19_18616479_A_G |
| 19 | RAB3A | 8.4324e-09 | 2.98e-08 | chr19_18381183_G_A |
| 20 | C20orf141 | 1.37456e-08 | 5.15e-10 | chr20_2800795_C_T |
| 20 | CPXM1 | 1.38587e-08 | 5.15e-10 | chr20_2800795_C_T |
| 20 | PCED1A | 1.32913e-08 | 5.15e-10 | chr20_2800795_C_T |
| 20 | TMEM239 | 1.24452e-08 | 5.15e-10 | chr20_2800795_C_T |
| 20 | VPS16 | 1.23202e-08 | 5.15e-10 | chr20_2800795_C_T |

**Table S7.** We investigated eQTL colocalizations between LSS association signals and gene expression using the 'coloc.abf' function from the R-library coloc[92]. The 'biomaRt' R-library was used to extract genes from Ensembl archives[93] within a 1 MB (± 500000) window surrounding each lead variant. Colocalizations were investigated for 754 genes that were available in GTEx. We chose tissues relevant to LSS for analysis, including the tibial nerve, cervical spinal cord, and whole blood. For the selected tissues, data on variant-gene expression associations (GTEx v8) were downloaded from GTExportal[7]. Colocalizations with a posterior probability of ≥ 0.8 for the shared variant (PP4) were deemed significant[94].

| Tissue | Gene_id | Gene | PP0 | PP1 | PP2 | PP3 | PP4 |
|---|---|---|---|---|---|---|---|
| Whole_Blood | ENSG00000198380.12 | *GFPT1* | 2.49e-36 | 1.05e-20 | 3.17e-17 | 0.133 | 0.867 |
| Whole_Blood | ENSG00000135913.10 | *USP37* | 3.47e-09 | 6.07e-05 | 6.15e-06 | 0.107 | 0.893 |
| Whole_Blood | ENSG00000144580.13 | *CNOT9* | 8.89e-42 | 4.66e-05 | 1.58e-38 | 0.082 | 0.918 |
| Whole_Blood | ENSG00000178467.17 | *P4HTM* | 3.9e-05 | 0.049 | 9.93e-05 | 0.123 | 0.829 |
| Whole_Blood | ENSG00000164062.12 | *APEH* | 3.73e-05 | 3.58e-05 | 0.085 | 0.08 | 0.835 |
| Whole_Blood | ENSG00000173531.15 | *MST1* | 0.001 | 0.022 | 0.006 | 0.095 | 0.875 |
| Whole_Blood | ENSG00000197409.7 | *HIST1H3D* | 0.000363 | 0.000388 | 0.008 | 0.008 | 0.984 |
| Whole_Blood | ENSG00000276966.2 | *HIST1H4E* | 7.21e-05 | 2.55e-05 | 0.013 | 0.004 | 0.983 |
| Whole_Blood | ENSG00000124549.14 | *BTN2A3P* | 0.005 | 0.01 | 0.032 | 0.06 | 0.891 |
| Whole_Blood | ENSG00000204610.12 | *TRIM15* | 1.02e-06 | 0.004 | 4.58e-05 | 0.19 | 0.806 |
| Whole_Blood | ENSG00000228022.5 | *HCG20* | 1.43e-08 | 3.63e-06 | 3.73e-05 | 0.008 | 0.991 |
| Whole_Blood | ENSG00000137310.11 | *TCF19* | 4.87e-25 | 2.55e-09 | 2.21e-17 | 0.115 | 0.885 |
| Whole_Blood | ENSG00000204472.12 | *AIF1* | 3.03e-11 | 0.016 | 3.61e-11 | 0.018 | 0.967 |
| Whole_Blood | ENSG00000247934.4 | *RP11-967K21.1* | 2.08e-06 | 0.004 | 4.78e-05 | 0.08 | 0.917 |
| Whole Blood | ENSG00000123427.16 | *METTL21B* | 2.81e-81 | 1.71e-05 | 1.12e-77 | 0.068 | 0.932 |
| Whole_Blood | ENSG00000123297.17 | *TSFM* | 5.98e-06 | 1.48e-05 | 0.018 | 0.044 | 0.939 |
| Whole_Blood | ENSG00000033030.13 | *ZCCHC8* | 0.037 | 0.018 | 0.03 | 0.014 | 0.9 |
| Whole_Blood | ENSG00000111011.17 | *RSRC2* | 2.04e-07 | 0.000427 | 6.97e-05 | 0.145 | 0.855 |
| Whole_Blood | ENSG00000150967.17 | *ABCB9* | 7.04e-05 | 0.015 | 0.00061 | 0.128 | 0.856 |
| Whole_Blood | ENSG00000129562.10 | *DAD1* | 2.09e-13 | 0.179 | 4.71e-15 | 0.003 | 0.818 |
| Whole_Blood | ENSG00000172590.18 | *MRPL52* | 2.35e-10 | 2.37e-06 | 5.07e-07 | 0.004 | 0.996 |
| Whole_Blood | ENSG00000108370.16 | *RGS9* | 0.076 | 0.072 | 0.017 | 0.016 | 0.818 |
| Whole_Blood | ENSG00000183397.5 | *TEKTIP1* | 2.06e-18 | 0.127 | 4.21e-19 | 0.025 | 0.848 |
| Whole_Blood | ENSG00000130517.13 | *PGPEP1* | 1.13e-12 | 0.000281 | 1.76e-10 | 0.043 | 0.957 |
| Whole_Blood | ENSG00000105655.18 | *ISYNA1* | 0.001 | 0.025 | 0.008 | 0.136 | 0.83 |
| Whole_Blood | ENSG00000132670.20 | *PTPRA* | 0.000581 | 0.004 | 0.007 | 0.048 | 0.941 |
| Whole_Blood | ENSG00000255568.3 | *BRWD1-AS2* | 3.61e-08 | 0.000268 | 1.22e-05 | 0.09 | 0.91 |
| Whole_Blood | ENSG00000205581.10 | *HMGN1* | 0.00033 | 0.028 | 0.002 | 0.16 | 0.81 |
| Cells_Cultured_fibroblasts | ENSG00000270882.2 | *HIST2H4A* | 6.39e-05 | 2.08e-06 | 0.11 | 0.003 | 0.887 |
| Cells_Cultured_fibroblasts | ENSG00000261716.1 | *RP11-196G18.22* | 8.45e-16 | 4.4e-14 | 0.003 | 0.176 | 0.821 |
| Cells_Cultured_fibroblasts | ENSG00000014914.20 | *MTMR11* | 1.7e-11 | 4.18e-09 | 5.98e-06 | 0.000471 | 1 |
| Cells_Cultured_fibroblasts | ENSG00000135913.10 | *USP37* | 5.09e-06 | 6.28e-05 | 0.009 | 0.108 | 0.883 |
| Cells_Cultured_fibroblasts | ENSG00000115556.13 | *PLCD4* | 1.31e-25 | 6.42e-05 | 2.32e-22 | 0.113 | 0.887 |
| Cells_Cultured_fibroblasts | ENSG00000115568.15 | *ZNF142* | 2.82e-06 | 7.9e-05 | 0.005 | 0.137 | 0.858 |
| Cells_Cultured_fibroblasts | ENSG00000178467.17 | *P4HTM* | 1.31e-26 | 0.046 | 3.78e-26 | 0.132 | 0.822 |
| Cells_Cultured_fibroblasts | ENSG00000198218.10 | *QRICH1* | 0.026 | 0.062 | 0.027 | 0.063 | 0.823 |
| Cells_Cultured_fibroblasts | ENSG00000235908.1 | *RHOA-IT1* | 6.28e-06 | 3.23e-05 | 0.02 | 0.101 | 0.879 |
| Cells_Cultured_fibroblasts | ENSG00000164330.16 | *EBF1* | 6.73e-05 | 0.003 | 0.001 | 0.047 | 0.949 |
| Cells_Cultured_fibroblasts | ENSG00000145860.11 | *RNF145* | 0.037 | 0.019 | 0.047 | 0.023 | 0.874 |
| Cells_Cultured_fibroblasts | ENSG00000228022.5 | *HCG20* | 1.04e-21 | 3.18e-06 | 3.55e-18 | 0.01 | 0.99 |
| Cells_Cultured_fibroblasts | ENSG00000204314.10 | *PRRT1* | 2.49e-05 | 0.01 | 8.27e-05 | 0.032 | 0.957 |
| Cells_Cultured_fibroblasts | ENSG00000221988.12 | *PPT2* | 4.09e-05 | 0.024 | 0.000144 | 0.083 | 0.894 |
| Cells_Cultured_fibroblasts | ENSG00000204308.7 | *RNF5* | 2.02e-07 | 0.004 | 9.91e-06 | 0.195 | 0.801 |
| Cells_Cultured_fibroblasts | ENSG00000204305.13 | *AGER* | 1.27e-11 | 0.007 | 9.08e-11 | 0.047 | 0.947 |
| Cells_Cultured_fibroblasts | ENSG00000204301.6 | *NOTCH4* | 2.25e-08 | 2.39e-08 | 0.081 | 0.085 | 0.833 |
| Cells_Cultured_fibroblasts | ENSG00000204287.13 | *HLA-DRA* | 0.003 | 0.000956 | 0.041 | 0.011 | 0.944 |
| Cells_Cultured_fibroblasts | ENSG00000179912.20 | *R3HDM2* | 0.015 | 0.071 | 0.016 | 0.074 | 0.824 |
| Cells_Cultured_fibroblasts | ENSG00000111012.9 | *CYP27B1* | 7.03e-08 | 1.36e-05 | 0.000277 | 0.052 | 0.947 |
| Cells_Cultured_fibroblasts | ENSG00000123427.16 | *METTL21B* | 2.54e-152 | 1.62e-05 | 1.02e-148 | 0.064 | 0.936 |
| Cells_Cultured_fibroblasts | ENSG00000123297.17 | *TSFM* | 8.3e-208 | 1.39e-05 | 3.32e-204 | 0.055 | 0.945 |
| Cells_Cultured_fibroblasts | ENSG00000245651.2 | *RP11-620J15.2* | 0.001 | 0.000443 | 0.055 | 0.017 | 0.926 |
| Cells_Cultured_fibroblasts | ENSG00000273805.1 | *RP11-620J15.4* | 0.038 | 0.025 | 0.043 | 0.028 | 0.866 |
| Cells_Cultured_fibroblasts | ENSG00000023516.8 | *AKAP11* | 0.00029 | 0.004 | 0.002 | 0.023 | 0.971 |
| Cells_Cultured_fibroblasts | ENSG00000129562.10 | *DAD1* | 0.013 | 0.09 | 0.000211 | 0.000525 | 0.896 |
| Cells_Cultured_fibroblasts | ENSG00000157227.12 | *MMP14* | 0.000106 | 0.01 | 8.56e-05 | 0.007 | 0.982 |
| Cells_Cultured_fibroblasts | ENSG00000137834.14 | *SMAD6* | 0.022 | 0.012 | 0.036 | 0.019 | 0.911 |
| Cells_Cultured_fibroblasts | ENSG00000276170.4 | *AC124789.1* | 0.000557 | 0.000869 | 0.011 | 0.016 | 0.971 |
| Cells_Cultured_fibroblasts | ENSG00000183397.5 | *TEKTIP1* | 8.19e-05 | 0.099 | 5.73e-06 | 0.006 | 0.895 |

| | | | | | | | |
|---|---|---|---|---|---|---|---|
| Cells_Cultured_fibroblasts | ENSG00000130513.6 | *GDF15* | 0.02 | 0.018 | 0.01 | 0.008 | 0.945 |
| Cells_Cultured_fibroblasts | ENSG00000088882.7 | *CPXM1* | 3.56e-08 | 1.99e-05 | 4.03e-05 | 0.022 | 0.978 |
| Cells_Cultured_fibroblasts | ENSG00000235701.1 | *PCBP2P1* | 4.08e-06 | 0.000411 | 0.002 | 0.158 | 0.84 |
| Cells_Cultured_fibroblasts | ENSG00000255568.3 | *BRWD1-AS2* | 5.96e-08 | 0.000261 | 1.76e-05 | 0.076 | 0.924 |
| Nerve_Tibial | ENSG00000014914.20 | *MTMR11* | 7.07e-09 | 2.44e-08 | 0.003 | 0.008 | 0.99 |
| Nerve_Tibial | ENSG00000081237.18 | *PTPRC* | 6.97e-07 | 1.34e-06 | 0.046 | 0.087 | 0.867 |
| Nerve_Tibial | ENSG00000169604.19 | *ANTXR1* | 1.35e-09 | 5.23e-09 | 0.023 | 0.088 | 0.889 |
| Nerve_Tibial | ENSG00000169599.12 | *NFU1* | 6.43e-23 | 8.79e-21 | 0.000817 | 0.111 | 0.888 |
| Nerve_Tibial | ENSG00000135913.10 | *USP37* | 6.18e-13 | 0.000103 | 1.1e-09 | 0.181 | 0.819 |
| Nerve_Tibial | ENSG00000115556.13 | *PLCD4* | 1.74e-07 | 9.02e-05 | 0.000302 | 0.156 | 0.844 |
| Nerve_Tibial | ENSG00000115568.15 | *ZNF142* | 1.13e-05 | 0.005 | 0.000303 | 0.143 | 0.852 |
| Nerve_Tibial | ENSG00000270441.1 | *RP11-694I15.7* | 0.002 | 0.055 | 0.003 | 0.067 | 0.873 |
| Nerve_Tibial | ENSG00000164062.12 | *APEH* | 5.48e-10 | 5.6e-05 | 1.77e-06 | 0.18 | 0.82 |
| Nerve_Tibial | ENSG00000164171.10 | *ITGA2* | 5.77e-06 | 0.009 | 1.34e-05 | 0.021 | 0.97 |
| Nerve_Tibial | ENSG00000239264.8 | *TXNDC5* | 1.76e-10 | 0.021 | 1.69e-10 | 0.019 | 0.96 |
| Nerve_Tibial | ENSG00000228022.5 | *HCG20* | 2.56e-16 | 3.15e-06 | 6.7e-13 | 0.007 | 0.993 |
| Nerve_Tibial | ENSG00000168631.12 | *DPCR1* | 8.79e-24 | 0.001 | 9.55e-22 | 0.117 | 0.882 |
| Nerve_Tibial | ENSG00000204498.10 | *NFKBIL1* | 2.49e-05 | 2.33e-05 | 0.006 | 0.005 | 0.989 |
| Nerve_Tibial | ENSG00000204472.12 | *AIF1* | 0.075 | 0.036 | 0.019 | 0.008 | 0.861 |
| Nerve_Tibial | ENSG00000204256.12 | *BRD2* | 1.83e-05 | 0.005 | 0.000553 | 0.138 | 0.857 |
| Nerve_Tibial | ENSG00000111816.7 | *FRK* | 0.001 | 0.004 | 0.012 | 0.047 | 0.936 |
| Nerve_Tibial | ENSG00000200534.1 | *SNORA33* | 0.009 | 0.051 | 0.015 | 0.079 | 0.846 |
| Nerve_Tibial | ENSG00000187546.13 | *AGMO* | 0.007 | 0.001 | 0.091 | 0.012 | 0.889 |
| Nerve_Tibial | ENSG00000153814.11 | *JAZF1* | 1.05e-10 | 5.59e-05 | 5.79e-08 | 0.03 | 0.97 |
| Nerve_Tibial | ENSG00000066382.16 | *MPPED2* | 2.24e-06 | 2.48e-05 | 0.012 | 0.132 | 0.856 |
| Nerve_Tibial | ENSG00000166908.17 | *PIP4K2C* | 0.000103 | 0.000483 | 0.014 | 0.063 | 0.923 |
| Nerve_Tibial | ENSG00000135506.15 | *OS9* | 0.000573 | 0.021 | 0.004 | 0.142 | 0.832 |
| Nerve_Tibial | ENSG00000135452.9 | *TSPAN31* | 1.04e-05 | 1.65e-05 | 0.03 | 0.047 | 0.923 |
| Nerve_Tibial | ENSG00000135446.16 | *CDK4* | 1.61e-05 | 0.000158 | 0.013 | 0.128 | 0.858 |
| Nerve_Tibial | ENSG00000111012.9 | *CYP27B1* | 2.31e-06 | 1.51e-05 | 0.008 | 0.051 | 0.941 |
| Nerve_Tibial | ENSG00000123427.16 | *METTL21B* | 9.51e-57 | 1.64e-05 | 3.81e-53 | 0.065 | 0.935 |
| Nerve_Tibial | ENSG00000123297.17 | *TSFM* | 6.28e-21 | 1.25e-05 | 2.52e-17 | 0.049 | 0.951 |
| Nerve_Tibial | ENSG00000033030.13 | *ZCCHC8* | 0.013 | 0.002 | 0.056 | 0.009 | 0.92 |
| Nerve_Tibial | ENSG00000184445.11 | *KNTC1* | 2.37e-06 | 0.091 | 5.68e-07 | 0.021 | 0.888 |
| Nerve_Tibial | ENSG00000103657.13 | *HERC1* | 0.043 | 0.099 | 0.004 | 0.009 | 0.845 |
| Nerve_Tibial | ENSG00000259347.6 | *RP11-798K3.2* | 2.77e-12 | 5.79e-13 | 0.043 | 0.008 | 0.949 |
| Nerve_Tibial | ENSG00000105325.13 | *FZR1* | 2.73e-18 | 0.097 | 6.55e-19 | 0.022 | 0.88 |
| Nerve_Tibial | ENSG00000183397.5 | *TEKTIP1* | 1.33e-27 | 0.14 | 3.81e-28 | 0.039 | 0.821 |
| Nerve_Tibial | ENSG00000255568.3 | *BRWD1-AS2* | 1.56e-07 | 0.00034 | 4.6e-05 | 0.099 | 0.9 |
| Brain_Spinal_cord_cervical_c-1 | ENSG00000178467.17 | *P4HTM* | 5.38e-07 | 0.051 | 1.5e-06 | 0.141 | 0.808 |
| Brain_Spinal_cord_cervical_c-1 | ENSG00000178252.17 | *WDR6* | 2.72e-05 | 0.055 | 6.69e-05 | 0.133 | 0.812 |
| Brain_Spinal_cord_cervical_c-1 | ENSG00000243753.5 | *HLA-L* | 0.016 | 0.063 | 0.015 | 0.059 | 0.846 |
| Brain_Spinal_cord_cervical_c-1 | ENSG00000228022.5 | *HCG20* | 2.64e-08 | 3.28e-06 | 0.000102 | 0.012 | 0.988 |
| Brain_Spinal_cord_cervical_c-1 | ENSG00000168631.12 | *DPCR1* | 0.002 | 0.002 | 0.054 | 0.07 | 0.872 |
| Brain_Spinal_cord_cervical_c-1 | ENSG00000196260.4 | *SFTA2* | 4.26e-11 | 0.002 | 1.34e-09 | 0.059 | 0.939 |
| Brain_Spinal_cord_cervical_c-1 | ENSG00000137310.11 | *TCF19* | 1.99e-09 | 3.82e-09 | 0.058 | 0.11 | 0.832 |
| Brain_Spinal_cord_cervical_c-1 | ENSG00000206344.7 | *HCG27* | 2.45e-07 | 3.37e-07 | 0.056 | 0.076 | 0.868 |
| Brain_Spinal_cord_cervical_c-1 | ENSG00000204525.16 | *HLA-C* | 1.61e-18 | 1.28e-06 | 1.29e-13 | 0.102 | 0.898 |
| Brain_Spinal_cord_cervical_c-1 | ENSG00000204308.7 | *RNF5* | 0.004 | 0.003 | 0.052 | 0.043 | 0.898 |
| Brain_Spinal_cord_cervical_c-1 | ENSG00000111816.7 | *FRK* | 0.024 | 0.085 | 0.005 | 0.017 | 0.868 |
| Brain_Spinal_cord_cervical_c-1 | ENSG00000231023.6 | *LINC00326* | 0.007 | 0.003 | 0.06 | 0.021 | 0.909 |
| Brain_Spinal_cord_cervical_c-1 | ENSG00000247934.4 | *RP11-967K21.1* | 0.000392 | 0.013 | 0.004 | 0.117 | 0.866 |
| Brain_Spinal_cord_cervical_c-1 | ENSG00000123427.16 | *METTL21B* | 2.7e-14 | 1.61e-05 | 1.06e-10 | 0.062 | 0.938 |
| Brain_Spinal_cord_cervical_c-1 | ENSG00000270039.1 | *RP11-571M6.17* | 0.041 | 0.016 | 0.058 | 0.022 | 0.862 |
| Brain_Spinal_cord_cervical_c-1 | ENSG00000073417.14 | *PDE8A* | 0.001 | 0.003 | 0.032 | 0.081 | 0.882 |
| Brain_Spinal_cord_cervical_c-1 | ENSG00000183527.11 | *PSMG1* | 2.85e-10 | 0.000245 | 1.1e-07 | 0.094 | 0.905 |
| Brain_Spinal_cord_cervical_c-1 | ENSG00000255568.3 | *BRWD1-AS2* | 0.000527 | 0.000327 | 0.023 | 0.013 | 0.963 |

**Table S8**. Calculated cumulative morbidities and surgery rates for every LSS-associated variant. We also calculated age when the curve of the effect allele (EA) homozygotes statistically differed from the curve of other allele (OA) homozygotes of the same variant (AGE (pval)). Following formula was utilised: *diff*=(*'survival rate EA/EA'-'survival rate OA/OA')/sqrt('se_survival_rate_EA/EA2'+'se_survival_rate_OA/OA2'*), p=2*(1-*diff*). P < 0.05 was used as the threshold for a significant difference. -, no significant difference between the homozygotes.

| Candidate Gene | rsid | EA | OA | AGE (pval) | Cum. Morb. EA | Cum. Morb. OA | Cum. Surg. EA | Cum. Surg. OA |
|---|---|---|---|---|---|---|---|---|
| ***RUNX3*** | rs9438857 | G | A | 54 (*p=0.006*) | 6.1% | 6.6% | 2.7% | 3.1% |
| ***NGF*** | rs6327 | T | C | 54 (*p=0.005*) | 6.1% | 6.7% | 2.8% | 3.2% |
| ***OTUD7B*** | rs11205303 | C | T | 40 (*p=1.82e-06*) | 5.8% | 6.6% | 2.6% | 3.2% |
| ***ZNF687*** | rs147042690 | T | C | - | 11.6% | 6.5% | 5.4% | 3.1% |
| ***PTPRC*** | rs72739729 | C | G | 57 (*p=0.03*) | 5.7% | 6.6% | 2.6% | 3.1% |
| ***GFPT1*** | rs4340572 | G | A | 44 (*p=0.0004*) | 6.1% | 6.7% | 2.8% | 3.2% |
| ***USP37*** | rs60362609 | G | C | 40 (*p=0.001*) | 6.2% | 6.8% | 2.9% | 3.2% |
| ***DAG1*** | rs138073019 | TTTA | T | 72 (p=0.004) | 6.4% | 6.6% | 3.0% | 3.1% |
| ***RAB28*** | rs3057070 | A | AGAG | 61 (p=0.01) | 6.0% | 6.6% | 2.7% | 3.1% |
| ***DAB2*** | rs148346348 | ATATG | A | 48 (*p=0.01*) | 7.8% | 6.5% | 4.0% | 3.1% |
| ***ITGA2*** | rs11747379 | A | C | 52 (*p=0.049*) | 7.1% | 6.5% | 3.4% | 3.1% |
| ***EBF1*** | rs200608668 | G | GC | - | 6.9% | 6.5% | 3.1% | 3.1% |
| ***BMP6*** | rs10498672 | G | C | 55 (*p=0.0004*) | 7.3% | 6.5% | 3.8% | 3.1% |
| ***TRIM38*** | rs9358913 | G | A | 61 (*p=0.02*) | 6.3% | 6.6% | 2.9% | 3.1% |
| ***HLA*** | rs62390186 | A | T | - | 6.1% | 6.6% | 2.9% | 3.1% |
| ***HLA*** | rs4294047 | A | G | - | 6.1% | 6.6% | 2.9% | 3.1% |
| ***HLA*** | rs17205170 | T | G | 54 (*p=0.02*) | 6.0% | 6.6% | 2.7% | 3.1% |
| ***CDC5L*** | rs7770012 | A | C | 40 (*p=1.52e-06*) | 6.8% | 6.4% | 3.3% | 3.0% |
| ***LIN28B*** | rs314261 | C | A | 50 (*p=0.03*) | 6.8% | 6.4% | 3.3% | 3.0% |
| ***COL10A1*** | rs7739896 | G | A | 53 (*p=0.0003*) | 6.3% | 6.6% | 2.9% | 3.1% |
| ***EYA4*** | rs2764217 | A | G | 50 (*p=0.03*) | 7.1% | 6.4% | 3.3% | 3.0% |
| ***FOXK1*** | rs28658109 | C | T | 46 (*p=0.02*) | 5.8% | 6.6% | 2.7% | 3.1% |
| ***AGMO*** | rs75674922 | G | A | 54 (*p=0.03*) | 7.8% | 6.5% | 3.8% | 3.1% |
| ***TWIST1*** | rs2390116 | T | C | 62 (*p=0.02*) | 6.3% | 6.6% | 2.9% | 3.1% |
| ***JAZF1*** | rs849135 | A | G | 45 (*p=0.001*) | 6.1% | 6.7% | 2.8% | 3.2% |
| ***Empty*** | rs139203647 | G | GT | 63 (*p=0.02*) | 7.7% | 6.5% | 3.9% | 3.1% |
| ***DYNC1I1*** | rs11769842 | C | T | 57 (*p=0.005*) | 6.8% | 6.2% | 3.2% | 2.9% |
| ***MKX*** | rs2637326 | T | G | 47 (*p=0.008*) | 6.8% | 6.5% | 3.1% | 3.1% |
| ***MPPED2*** | rs607165 | C | T | 55 (*p=0.001*) | 6.9% | 6.4% | 3.3% | 3.0% |
| ***PDE3A*** | rs73067286 | A | T | 52 (*p=0.004*) | 5.8% | 6.6% | 2.8% | 3.1% |
| ***SOX5*** | rs12308843 | C | G | 40 (*p=0.0001*) | 7.0% | 6.5% | 3.4% | 3.1% |
| ***CCDC91*** | rs7958415 | T | C | 46 (*p=0.005*) | 5.8% | 6.6% | 2.6% | 3.1% |
| ***GLI1*** | rs2069506 | A | C | 66 (*p=0.03*) | 6.2% | 6.6% | 2.9% | 3.1% |
| ***USP15*** | rs11174377 | A | G | 57 (*p=0.01*) | 7.0% | 6.5% | 3.3% | 3.1% |
| ***ZCCHC8*** | rs10846978 | C | T | - | 6.4% | 6.6% | 3.1% | 3.0% |
| ***TNFSF11*** | rs9594751 | T | C | 43 (*p=0.002*) | 5.8% | 6.6% | 2.5% | 3.1% |
| ***SLITRK6*** | rs9531779 | G | C | 55 (*p=0.002*) | 6.3% | 6.6% | 3.0% | 3.1% |
| ***MMP14*** | rs1042704 | A | G | 51 (*p=0.008*) | 5.9% | 6.6% | 2.7% | 3.1% |
| ***GPR132*** | rs1022431 | A | C | - | 6.1% | 6.6% | 2.8% | 3.1% |
| ***ALDH1A2*** | rs4646570 | C | G | 56 (*p=0.002*) | 7.0% | 6.4% | 3.5% | 3.0% |
| ***PPIB*** | rs148027154 | A | T | - | 8.1% | 6.5% | 6.3% | 3.1% |
| ***SMAD3*** | rs4776880 | A | G | 40 (*p=6.58e-06*) | 6.0% | 6.6% | 2.8% | 3.1% |
| ***PDE8A*** | rs11629962 | T | C | 54 (*p=0.01*) | 6.0% | 6.6% | 2.9% | 3.1% |
| ***GAN*** | rs2966084 | G | A | 56 (*p=0.01*) | 6.3% | 6.8% | 3.0% | 3.2% |
| ***SOCS7*** | rs113353816 | C | T | 63 (*p=0.04*) | 6.7% | 6.4% | 3.1% | 3.0% |
| ***AXIN2*** | rs16961974 | C | T | 70 (*p=0.03*) | 6.8% | 6.5% | 3.2% | 3.1% |
| ***Empty*** | rs12941198 | C | A | 62 (*p=0.03*) | 6.8% | 6.2% | 3.2% | 2.9% |
| ***DCC*** | rs6508210 | C | A | 45 (*p=0.001*) | 6.9% | 6.4% | 3.2% | 3.0% |
| ***CACTIN*** | rs77733715 | G | A | - | 9.2% | 6.5% | 4.7% | 3.1% |
| ***CRLF1*** | rs2074177 | G | A | 65 (*p=0.03*) | 6.0% | 6.6% | 2.8% | 3.1% |
| ***DDRGK1*** | rs11697820 | T | C | 65 (*p=0.04*) | 6.3% | 6.7% | 3.0% | 3.1% |
| ***ETS2*** | rs111901809 | T | TAGAGGGGCAGGGGGCG | 47 (*p=0.004*) | 6.8% | 6.4% | 3.2% | 3.0% |
| ***OSM*** | rs2285645 | T | C | - | 4.4% | 6.6% | 2.0% | 3.1% |

**Table S9.** Genetic correlations between 517 traits and LSS. LDSC-software[95], was utilised for calculation of the correlations. Traits were extracted from the GWAS database provided by the MRC Integrative Epidemiology Unit (IEU). rg, genetic correlation coefficient value; se, standard error; pFDR, false discovery rate-corrected p-value.

| Trait | GWAS-ID | rg | se | pFDR |
|---|---|---|---|---|
| **Diagnoses - main ICD-10: M54 Dorsalgia** | ukb-a-569 | 0.7543 | 0.06612 | 7.40e-29 |
| **Main ICD-10: M25 Other joint disorders, not elsewhere classified** | ukb-d-M25 | 0.7229 | 0.1138 | 1.21e-09 |
| **Back pain** | ukb-b-9838 | 0.6031 | 0.02683 | 5.05e-109 |
| **Leg pain on walking** | ukb-b-10387 | 0.5652 | 0.04453 | 1.89e-35 |
| **Hip pain** | ukb-b-7289 | 0.5482 | 0.03365 | 1.24e-57 |
| **Main ICD-10: M70 Soft tissue disorders related to use, overuse, and pressure** | ukb-d-M70 | 0.5014 | 0.223 | 0.041 |
| **Back pain for 3+ months** | ukb-b-8463 | 0.4958 | 0.05587 | 6.79e-18 |
| **Main ICD-10: T84 Complications of internal orthopaedic prosthetic devices, implants, and grafts** | ukb-d-T84 | 0.4859 | 0.08538 | 5.54e-08 |
| **Diagnoses - main ICD-10: J22 Unspecified acute lower respiratory infection** | ukb-b-8707 | 0.4761 | 0.381 | 0.278 |
| **Diagnoses - main ICD-10: K30 Dyspepsia** | ukb-b-14814 | 0.468 | 0.1135 | 0.0001 |
| **Number of operations, self-reported** | ukb-b-4733 | 0.458 | 0.02828 | 5.38e-57 |
| **Neck/shoulder pain for 3+ months** | ukb-b-16118 | 0.4546 | 0.06353 | 5.56e-12 |
| **Neck or shoulder pain** | ukb-b-18596 | 0.454 | 0.02897 | 1.97e-53 |
| **Taking other prescription medications** | ukb-b-20292 | 0.4506 | 0.03005 | 6.10e-49 |
| **Diagnoses - main ICD-10: I20 Angina pectoris** | ukb-a-532 | 0.4443 | 0.07324 | 6.50e-09 |
| **Diagnoses - main ICD-10: R07 Pain in throat and chest** | ukb-a-581 | 0.4412 | 0.04621 | 1.56e-20 |
| **Major depressive disorder (ICD-10 coded)** | ebi-a-GCST005903 | 0.4358 | 0.05246 | 8.26e-16 |
| **Long-standing illness, disability, or infirmity** | ukb-b-13764 | 0.4352 | 0.02657 | 3.39e-58 |
| **Number of treatments/medications taken** | ukb-b-3656 | 0.4345 | 0.0243 | 4.26e-69 |
| **Chest pain or discomfort walking normally** | ukb-b-11578 | 0.4327 | 0.0557 | 6.08e-14 |
| **Had major operations** | ukb-b-9937 | 0.4275 | 0.05072 | 3.13e-16 |
| **Diagnoses - main ICD-10: J34 Other disorders of nose and nasal sinuses** | ukb-a-542 | 0.4241 | 0.1592 | 0.014 |
| **Diagnoses - main ICD-10: R11 Nausea and vomiting** | ukb-b-19419 | 0.4234 | 0.1708 | 0.023 |
| **Pain all over the body** | ukb-a-477 | 0.4129 | 0.05168 | 1.08e-14 |
| **Diagnoses - main ICD-10: G56 Mononeuropathies of upper limb** | ukb-a-528 | 0.4106 | 0.05256 | 4.39e-14 |
| **Diagnoses - main ICD-10: R10 Abdominal and pelvic pain** | ukb-a-582 | 0.4106 | 0.05693 | 3.85e-12 |
| **Why reduced smoking: Illness or ill health** | ukb-b-9930 | 0.4094 | 0.1838 | 0.043 |
| **Why reduced smoking: Doctor's advice** | ukb-b-17579 | 0.4067 | 0.2977 | 0.232 |
| **Overall health rating** | ukb-b-6306 | 0.4047 | 0.02328 | 2.25e-65 |
| **Medication for pain relief, constipation, heartburn: Laxatives** | ukb-b-11461 | 0.4041 | 0.04579 | 1.05e-17 |
| **Diagnoses - main ICD-10: K29 Gastritis and duodenitis** | ukb-a-547 | 0.4025 | 0.06778 | 1.36e-08 |
| **Medication for pain relief, constipation, heartburn: Omeprazole** | ukb-b-11605 | 0.3988 | 0.0431 | 2.39e-19 |
| **Lumbar spine bone mineral density** | ieu-a-982 | 0.3955 | 0.04694 | 3.18e-16 |
| **Diagnoses - main ICD-10:R35 Polyuria** | ukb-b-5948 | 0.393 | 0.1248 | 0.003 |
| **Diagnoses - main ICD-10: K52 Other non-infective gastro-enteritis and colitis** | ukb-a-554 | 0.3914 | 0.09023 | 4.20e-05 |
| **Knee pain** | ukb-b-16254 | 0.384 | 0.03448 | 1.40e-27 |
| **Diagnoses - main ICD-10: R69 Unknown and unspecified causes of morbidity** | ukb-b-12510 | 0.3832 | 0.09176 | 8.24e-05 |
| **Knee pain for 3+ months** | ukb-b-8906 | 0.3828 | 0.08275 | 1.15e-05 |
| **Illness, injury, bereavement, stress in the last 2 years: Serious illness, injury, or assault to yourself** | ukb-b-18162 | 0.3817 | 0.04249 | 2.63e-18 |
| **Health satisfaction** | ukb-b-2053 | 0.3817 | 0.03248 | 1.51e-30 |
| **Ever had hysterectomy (womb removed)** | ukb-b-6445 | 0.3769 | 0.05241 | 4.32e-12 |
| **Diagnoses - main ICD-10: M24 Other specific joint derangements** | ukb-a-567 | 0.3766 | 0.1149 | 0.002 |
| **Diagnoses - main ICD-10: M21 Other acquired deformities of limbs** | ukb-a-565 | 0.3727 | 2.404 | 0.894 |
| **Vascular/heart problems diagnosed by doctor: Angina** | ukb-b-8468 | 0.3713 | 0.03715 | 2.17e-22 |
| **Diagnoses - main ICD-10: M17 Gonarthrosis [arthrosis of knee]** | ukb-a-563 | 0.3702 | 0.05174 | 5.56e-12 |
| **Had other major operations** | ukb-b-15378 | 0.3677 | 0.037 | 3.71e-22 |
| **Reason for reducing amount of alcohol drunk: Illness or ill health** | ukb-d-2664_1 | 0.3568 | 0.1052 | 0.002 |
| **Diagnoses - main ICD-10: G47 Sleep disorders** | ukb-a-527 | 0.3524 | 0.07514 | 8.56e-06 |
| **Medication for pain relief, constipation, heartburn: Ranitidine** | ukb-b-8146 | 0.3468 | 0.07108 | 3.50e-06 |
| **Diagnoses - main ICD-10: K62 Other diseases of anus and rectum** | ukb-a-557 | 0.3413 | 0.08907 | 0.0003 |
| **Ever used hormone-replacement therapy (HRT)** | ukb-b-18541 | 0.3382 | 0.0357 | 3.01e-20 |
| **Frequency of tiredness/lethargy in last 2 weeks** | ukb-b-929 | 0.3381 | 0.02681 | 4.86e-35 |
| **Alcohol drinker status: Previous** | ukb-a-227 | 0.3362 | 0.04852 | 2.70e-11 |
| **Medication for pain relief, constipation, heartburn: Paracetamol** | ukb-b-17595 | 0.3308 | 0.02845 | 6.42e-30 |
| **Other serious medical condition/disability diagnosed by doctor** | ukb-b-14961 | 0.3305 | 0.03341 | 5.74e-22 |
| **Ever highly irritable/argumentative for 2 days** | ukb-b-8196 | 0.3302 | 0.04128 | 1.01e-14 |
| **Shortness of breath walking on level ground** | ukb-b-9543 | 0.3279 | 0.04709 | 2.14e-11 |
| **Diagnoses - main ICD-10: M16 Coxarthrosis [arthrosis of hip]** | ukb-a-562 | 0.3266 | 0.0535 | 5.17e-09 |
| **Emphysema/chronic bronchitis** | ukb-b-16207 | 0.325 | 0.05243 | 2.97e-09 |

| | | | | |
|---|---|---|---|---|
| **Diagnoses - main ICD-10: M67 Other disorders of synovium and tendon** | ukb-a-570 | 0.322 | 0.1519 | 0.055 |
| **Frequency of depressed mood in last 2 weeks** | ukb-b-3822 | 0.3216 | 0.03533 | 8.91e-19 |
| **Falls in the last year** | ukb-b-2535 | 0.3212 | 0.03005 | 1.74e-25 |
| **Medication for pain relief, constipation, heartburn: Ibuprofen** | ukb-b-8888 | 0.3184 | 0.03733 | 1.35e-16 |
| **Childhood asthma (age<16)** | ukb-d-ASTHMA_CHILD | 0.3152 | 0.08253 | 0.0003 |
| **Diagnoses - main ICD-10: I25 Chronic ischemic heart disease** | ukb-a-534 | 0.3118 | 0.04279 | 2.26e-12 |
| **Chest pain or discomfort** | ukb-b-10591 | 0.3109 | 0.02984 | 2.87e-24 |
| **Frequency of unenthusiasm/disinterest in the last 2 weeks** | ukb-b-1419 | 0.3097 | 0.03351 | 2.60e-19 |
| **Seen doctor (GP) for nerves, anxiety, tension, or depression** | ukb-b-6991 | 0.3076 | 0.02706 | 1.12e-28 |
| **Diagnoses - main ICD-10: M23 Internal derangement of knee** | ukb-a-566 | 0.3066 | 0.08106 | 0.0004 |
| **Adopted as a child** | ukb-b-13183 | 0.3047 | 0.06779 | 2.11e-05 |
| **Diagnoses - main ICD-10: K21 Gastro-oesophageal reflux disease** | ukb-a-545 | 0.3 | 0.07126 | 7.16e-05 |
| **Transport type for commuting to workplace: Car/motor vehicle** | ukb-b-11025 | 0.2998 | 0.045 | 1.66e-10 |
| **Frequency of tenseness/restlessness in last 2 weeks** | ukb-b-5664 | 0.2979 | 0.03016 | 6.48e-22 |
| **Leg fat mass (left)** | ukb-b-7212 | 0.2976 | 0.02046 | 3.97e-46 |
| **Leg fat mass (right)** | ukb-b-18096 | 0.2957 | 0.02056 | 3.96e-45 |
| **Coronary artery disease** | ebi-a-GCST005195 | 0.294 | 0.02842 | 6.17e-24 |
| **Body mass index (BMI)** | ukb-b-19953 | 0.2928 | 0.01999 | 9.76e-47 |
| **Number of children fathered** | ukb-b-2227 | 0.2927 | 0.04045 | 3.24e-12 |
| **Bilateral oophorectomy (both ovaries removed)** | ukb-a-325 | 0.2926 | 0.05574 | 5.88e-07 |
| **Medication for pain relief, constipation, heartburn: Aspirin** | ukb-b-7137 | 0.292 | 0.034 | 8.31e-17 |
| **Mood swings** | ukb-b-14180 | 0.2895 | 0.02826 | 1.67e-23 |
| **Wheeze or whistling in the chest in the last year** | ukb-b-18335 | 0.2885 | 0.03027 | 1.76e-20 |
| **Vascular/heart problems diagnosed by doctor: Heart attack** | ukb-b-11590 | 0.2884 | 0.04347 | 1.95e-10 |
| **Sleeplessness/insomnia** | ukb-b-3957 | 0.2881 | 0.02539 | 1.38e-28 |
| **Medication for cholesterol, blood pressure, diabetes or take exogenous hormones: Hormone replacement therapy** | ukb-b-8080 | 0.2878 | 0.0591 | 3.68e-06 |
| **Diagnoses - main ICD-10: Z09 Follow-up examination after treatment for conditions other than malignant neoplasms** | ukb-a-594 | 0.2853 | 0.1064 | 0.013 |
| **Medication for cholesterol, blood pressure, diabetes or take exogenous hormones: Cholesterol lowering medication** | ukb-b-17805 | 0.285 | 0.03933 | 3.03e-12 |
| **Maternal smoking around birth** | ukb-b-17685 | 0.2835 | 0.02901 | 1.79e-21 |
| **Noisy workplace** | ukb-b-2091 | 0.2834 | 0.03983 | 7.40e-12 |
| **Waist circumference** | ukb-b-9405 | 0.2818 | 0.02043 | 1.61e-41 |
| **Diagnoses - main ICD-10: J44 Other chronic obstructive pulmonary disease** | ukb-a-543 | 0.279 | 0.09043 | 0.0039943 |
| **Former alcohol drinker** | ukb-b-12654 | 0.279 | 0.05559 | 1.82e-06 |
| **Diagnoses - main ICD-10: J45.9 Asthma, unspecified** | ukb-b-20208 | 0.2786 | 0.1467 | 0.087 |
| **Ever had bowel cancer screening** | ukb-b-17001 | 0.2772 | 0.04815 | 3.83e-08 |
| **Vitamin and mineral supplements: Vitamin B** | ukb-b-10188 | 0.2771 | 0.05369 | 9.11e-07 |
| **Diagnoses - main ICD-10: K20 Oesophagitis** | ukb-b-19354 | 0.2766 | 0.09267 | 0.005 |
| **Weight** | ukb-b-11842 | 0.2762 | 0.02042 | 4.90e-40 |
| **Main ICD-10: N92 Excessive , frequent, and irregular menstruation** | ukb-d-N92 | 0.2752 | 0.06264 | 3.30e-05 |
| **Operation code: bilateral oophorectomy** | ukb-b-1962 | 0.2741 | 0.05391 | 1.34e-06 |
| **Arm fat-free mass (left)** | ukb-b-19925 | 0.2726 | 0.02016 | 4.90e-40 |
| **Bring up phlegm/sputum/mucus on most days** | ukb-b-12841 | 0.2719 | 0.06711 | 0.0001 |
| **Arm fat-free mass (right)** | ukb-b-19520 | 0.2711 | 0.0199 | 1.39e-40 |
| **Major coronary heart disease event** | ukb-d-I9_CHD | 0.268 | 0.04379 | 4.77e-09 |
| **Leg fat percentage** | ukb-b-18377 | 0.2678 | 0.02094 | 6.06e-36 |
| **Arm fat mass (right)** | ukb-b-6704 | 0.2678 | 0.02024 | 1.91e-38 |
| **Arm fat mass (left)** | ukb-b-8338 | 0.2676 | 0.0205 | 2.14e-37 |
| **Depressive symptoms** | ieu-a-1000 | 0.2674 | 0.04703 | 5.68e-08 |
| **Fed-up feelings** | ukb-b-19809 | 0.2673 | 0.02992 | 4.03e-18 |
| **Blood clot in the lung** | ukb-b-13704 | 0.2668 | 0.0736 | 0.0006 |
| **Stomach or abdominal pian** | ukb-b-11413 | 0.2667 | 0.04577 | 2.56e-08 |
| **Diagnoses - main ICD-10: R04 Haemorrhage from respiratory passages** | ukb-a-580 | 0.2656 | 0.1251 | 0.054 |
| **Whole body fat mass** | ukb-b-19393 | 0.2642 | 0.02067 | 6.47e-36 |
| **Smoking status: Current** | ukb-a-225 | 0.2626 | 0.03102 | 2.25e-16 |
| **Diagnoses - secondary ICD-10: M06.99 Rheumatoid arthritis, unspecified (Site unspecified)** | ukb-b-11874 | 0.2622 | 0.08531 | 0.004 |
| **Diagnoses - main ICD-10: K43 Ventral hernia** | ukb-a-550 | 0.2605 | 0.07408 | 0.001 |
| **Leg fat percentage (right)** | ukb-b-20531 | 0.2592 | 0.02091 | 7.00e-34 |
| **Pack years of smoking** | ukb-b-10831 | 0.2577 | 0.03279 | 3.03e-14 |
| **Pulse wave arterial stiffness index** | ukb-b-11971 | 0.2573 | 0.04611 | 1.00e-07 |
| **Ever stopped smoking for 6+ months** | ukb-b-11355 | 0.2542 | 0.08089 | 0.003 |
| **Hip circumference** | ukb-b-15590 | 0.2531 | 0.01998 | 2.49e-35 |
| **Smoking/smokers in household** | ukb-b-960 | 0.253 | 0.04618 | 1.77e-07 |
| **Qualifications: None of the above** | ukb-b-17729 | 0.2525 | 0.02252 | 6.24e-28 |
| **Basal metabolic rate** | ukb-b-16446 | 0.2518 | 0.02022 | 3.54e-34 |

| | | | | |
|---|---|---|---|---|
| **Loneliness, isolation** | ukb-b-8476 | 0.25 | 0.03266 | 1.46e-13 |
| **Other eye problems** | ukb-b-7007 | 0.2495 | 0.04843 | 9.49e-07 |
| **Illness, injury, bereavement, stress in the last 2 years: Financial difficulties** | ukb-b-9667 | 0.2484 | 0.03138 | 1.94e-14 |
| **Ever had stillbirth, spontaneous miscarriage, or termination** | ukb-b-12621 | 0.2474 | 0.04761 | 7.63e-07 |
| **Smoking initiation** | ieu-b-4877 | 0.2471 | 0.0211 | 2.31e-30 |
| **Type 2 diabetes** | ebi-a-GCST006867 | 0.2469 | 0.02953 | 5.53e-16 |
| **Mouth/teeth dental problems: Toothache** | ukb-b-19191 | 0.2447 | 0.06593 | 0.0005 |
| **Job involves shift work** | ukb-b-1712 | 0.2445 | 0.04163 | 1.99e-08 |
| **Blood clot in the leg (DVT)** | ukb-b-11411 | 0.2417 | 0.0562 | 4.87e-05 |
| **Trunk fat mass** | ukb-b-20044 | 0.2414 | 0.02109 | 4.79e-29 |
| **Reason for reducing amount of alcohol drunk: Doctor’s advice** | ukb-d-2664_2 | 0.2413 | 0.1144 | 0.056 |
| **Job involves heavy manual or physical work** | ukb-b-2002 | 0.2395 | 0.02884 | 8.26e-16 |
| **Whole body water mass** | ukb-b-14540 | 0.2394 | 0.02025 | 6.92e-31 |
| **Diagnoses - main ICD-10: K22 Other diseases of oesophagus** | ukb-a-546 | 0.2383 | 0.09374 | 0.019 |
| **Whole body fat-free mass** | ukb-b-13354 | 0.2375 | 0.02024 | 1.87e-30 |
| **Current tobacco smoking** | ukb-b-223 | 0.2367 | 0.02884 | 1.84e-15 |
| **Childhood obesity** | ieu-a-1096 | 0.2352 | 0.04796 | 3.13e-06 |
| **Leg fat free-mass (left)** | ukb-b-16099 | 0.2337 | 0.0206 | 1.42e-28 |
| **Facial pain** | ukb-b-17107 | 0.2337 | 0.07309 | 0.003 |
| **Diagnoses - main ICD-10: I10 Essential (primary) hypertension** | ukb-a-531 | 0.2335 | 0.1284 | 0.102 |
| **Diabetes diagnosed by doctor** | ukb-b-10753 | 0.2332 | 0.03192 | 2.01e-12 |
| **Financial situation satisfaction** | ukb-b-2830 | 0.2328 | 0.03736 | 2.47e-09 |
| **Extreme waist-to-hip ratio** | ieu-a-87 | 0.2324 | 0.07126 | 0.002 |
| **Seen a psychiatrist for nerves, anxiety, tension, or depression** | ukb-b-18336 | 0.2322 | 0.0354 | 3.15e-10 |
| **Average weekly spirits intake** | ukb-b-1707 | 0.2321 | 0.03499 | 1.99e-10 |
| **Qualifications: NVQ or HND or HNC or equivalent** | ukb-b-7926 | 0.2303 | 0.03987 | 3.45e-08 |
| **Leg fat-free mass (right)** | ukb-b-12828 | 0.2279 | 0.02049 | 1.65e-27 |
| **Waist-to-hip ratio** | ieu-a-72 | 0.2278 | 0.03047 | 5.64e-13 |
| **Diagnoses - main ICD-10: I21 Acute myocardial infarction** | ukb-a-533 | 0.2274 | 0.06478 | 0.001 |
| **Miserableness** | ukb-b-18994 | 0.227 | 0.03078 | 1.20e-12 |
| **Trunk fat-free mass** | ukb-b-17409 | 0.2257 | 0.02043 | 3.61e-27 |
| **Body fat percentage** | ukb-b-8909 | 0.2227 | 0.02107 | 5.84e-25 |
| **Number of full brothers** | ukb-b-4263 | 0.222 | 0.04402 | 1.64e-06 |
| **Number of older siblings** | ukb-b-1997 | 0.2196 | 0.07887 | 0.010 |
| **Vascular/heart problems diagnosed by doctor: Stroke** | ukb-b-8714 | 0.2194 | 0.07199 | 0.005 |
| **Arm fat percentage (left)** | ukb-b-20188 | 0.2191 | 0.02083 | 9.77e-25 |
| **Arm fat percentage (right)** | ukb-b-12854 | 0.2182 | 0.0207 | 7.95e-25 |
| **Ever unenthusiastic/disinterested for a whole week** | ukb-b-18674 | 0.2181 | 0.03865 | 7.22e-08 |
| **Types of physical activity in last 4 weeks: None of the above** | ukb-b-15869 | 0.2177 | 0.03735 | 2.56e-08 |
| **Vascular/heart problems diagnosed by doctor: High blood pressure** | ukb-b-14177 | 0.2174 | 0.02334 | 1.36e-19 |
| **Mouth/teeth dental problems: Dentures** | ukb-b-12930 | 0.2173 | 0.03016 | 4.02e-12 |
| **Mouth/teeth dental problems: Painful gums** | ukb-b-11161 | 0.217 | 0.05991 | 0.0007 |
| **Diagnoses - main ICD-10: M20 Acquired deformities of fingers and toes** | ukb-a-564 | 0.2157 | 0.05354 | 0.0001 |
| **Vitamin and mineral supplements: Vitamin A** | ukb-b-9596 | 0.2145 | 0.08409 | 0.019 |
| **Diagnoses - main ICD-10: K44 Diaphragmatic hernia** | ukb-a-551 | 0.214 | 0.07704 | 0.010 |
| **Diagnoses - main ICD-10: R31 Unspecified haematuria** | ukb-b-17885 | 0.2136 | 0.07223 | 0.006 |
| **Cigarettes smoked per day** | ieu-b-142 | 0.2133 | 0.03089 | 3.14e-11 |
| **Cigarettes per day** | ieu-b-25 | 0.2133 | 0.03089 | 3.14e-11 |
| **Diagnoses - main ICD-10: I84 Haemorrhoids** | ukb-a-539 | 0.2131 | 0.05595 | 0.0003 |
| **Exposure to tobacco smoke at home** | ukb-b-4462 | 0.2103 | 0.04221 | 2.15e-06 |
| **Qualifications: CSEs or equivalent** | ukb-b-10505 | 0.2067 | 0.03774 | 1.79e-07 |
| **Mineral and other dietary supplements: Glucosamine** | ukb-b-11535 | 0.2055 | 0.03419 | 9.13e-09 |
| **Tinnitus, Yes, but not now, but have in the past** | ukb-d-4803_14 | 0.2052 | 0.06871 | 0.005 |
| **Number of cigarettes previously smoked daily** | ukb-b-6019 | 0.2036 | 0.03921 | 7.72e-07 |
| **Neuroticism score** | ukb-b-4630 | 0.2006 | 0.03033 | 2.24e-10 |
| **Time spent watching television** | ukb-b-5192 | 0.1964 | 0.0236 | 7.45e-16 |
| **Trunk fat percentage** | ukb-b-16407 | 0.196 | 0.02144 | 6.49e-19 |
| **Handedness (chirality/laterality): Use both right and left hands equally** | ukb-a-37 | 0.1957 | 0.09527 | 0.063 |
| **Frequency of heavy DIY in last 4 weeks** | ukb-b-5053 | 0.1954 | 0.05564 | 0.001 |
| **Fractured bone site(s): Spine** | ukb-b-873 | 0.1952 | 0.1851 | 0.361 |
| **Diagnoses - main ICD-10: N40 Hyperplasia of prostate** | ukb-b-11601 | 0.1951 | 0.06251 | 0.004 |
| **Duration of heavy DIY** | ukb-b-2634 | 0.1947 | 0.04471 | 3.90e-05 |
| **Medication for cholesterol, blood pressure, diabetes or take exogenous hormones: Blood pressure medication** | ukb-b-18009 | 0.1943 | 0.028 | 2.51e-11 |
| **Diagnoses - main ICD-10: K60 Fissure and fistula of anal and rectal regions** | ukb-a-556 | 0.1928 | 0.08046 | 0.028 |
| **Light smokers, at least 100 smokes in lifetime** | ukb-b-8133 | 0.1926 | 0.03768 | 1.16e-06 |
| **Neuroticism** | ebi-a-GCST005232 | 0.1912 | 0.03064 | 2.36e-09 |
| **Job involves mainly walking or standing** | ukb-b-4461 | 0.1886 | 0.02835 | 1.77e-10 |

| | | | | |
|---|---|---|---|---|
| **Eye problems/disorders: Injury or trauma resulting in loss of vision** | ukb-b-9050 | 0.1877 | 0.1193 | 0.163 |
| **Diagnoses - main ICD-10: N81 Female genital prolapse** | ukb-a-577 | 0.1876 | 0.06647 | 0.009 |
| **HOMA-IR** | ieu-b-118 | 0.1872 | 0.07464 | 0.021 |
| **Eye problems/disorders: Diabetes related eye disease** | ukb-b-10181 | 0.1868 | 0.06675 | 0.010 |
| **Ever manic/hyper for 2 days** | ukb-b-15272 | 0.1843 | 0.05674 | 0.002 |
| **Tense/’highly strung’** | ukb-b-10093 | 0.1842 | 0.0309 | 1.21e-08 |
| **Cataract** | bbj-a-94 | 0.1822 | 0.08889 | 0.063 |
| **Illness, injury, bereavement, stress in the last 2 years: Marital separation/divorce** | ukb-b-14624 | 0.181 | 0.0859 | 0.056 |
| **Vitamin and mineral supplements: Vitamin E** | ukb-b-12506 | 0.1799 | 0.06726 | 0.014 |
| **Fractured bone site(s): Other bones** | ukb-b-17738 | 0.1795 | 0.05673 | 0.003 |
| **Diagnoses - main ICD-10: S76 Injury of muscle and tendon at hip and thigh level** | ukb-a-592 | 0.1791 | 0.1568 | 0.321 |
| **Fractured bone site(s): Ankle** | ukb-b-15582 | 0.1766 | 0.07074 | 0.022 |
| **Triglycerides** | ieu-b-111 | 0.1758 | 0.02727 | 6.61e-10 |
| **Hearing difficulty/problems: Yes** | ukb-a-257 | 0.1754 | 0.03368 | 7.23e-07 |
| **Diagnoses - main ICD-10: K80 Cholelithiasis** | ukb-a-559 | 0.171 | 0.06786 | 0.020 |
| **Smoking status: Previous** | ukb-a-224 | 0.1702 | 0.03021 | 7.61e-08 |
| **Diagnoses - main ICD-10: K57 Diverticular disease of intestine** | ukb-a-555 | 0.17 | 0.05136 | 0.002 |
| **Headache** | ukb-b-12181 | 0.1688 | 0.03406 | 2.45e-06 |
| **Diagnoses - main ICD-10: K76 Other diseases of liver** | ukb-a-558 | 0.168 | 0.09632 | 0.118 |
| **Irritability** | ukb-b-13745 | 0.1665 | 0.03492 | 6.00e-06 |
| **Alcohol intake versus 10 years previously** | ukb-b-3460 | 0.1647 | 0.032 | 9.70e-07 |
| **Number of full sisters** | ukb-b-5593 | 0.1629 | 0.04458 | 0.0006 |
| **Duration of moderate activity** | ukb-b-2346 | 0.1609 | 0.03531 | 1.60e-05 |
| **Illness, injury, bereavement, stress in the last 2 years: Death of a close relative** | ukb-b-18712 | 0.1581 | 0.05821 | 0.012 |
| **Types of transport used (excluding work): None of the above** | ukb-b-12386 | 0.1556 | 0.1264 | 0.286 |
| **Prospective memory result** | ukb-b-4282 | 0.1544 | 0.03804 | 0.0001 |
| **Tinnitus, Yes, now a lot of the time** | ukb-d-4803_12 | 0.154 | 0.1353 | 0.322 |
| **Risk taking** | ukb-b-14147 | 0.1538 | 0.0283 | 2.21e-07 |
| **Diagnoses - main ICD-10: I30 Acute pericarditis** | ukb-a-535 | 0.1515 | 0.1111 | 0.233 |
| **Pulse wave reflection index** | ukb-b-11598 | 0.1515 | 0.04294 | 0.0009 |
| **Ever smoked** | ukb-b-20261 | 0.1515 | 0.02426 | 2.32e-09 |
| **Diagnoses - main ICD-10: J33 Nasal polyp** | ukb-a-541 | 0.1493 | 0.1045 | 0.211 |
| **Exposure to tobacco smoke outside home** | ukb-b-6244 | 0.1492 | 0.03656 | 0.0001 |
| **Ever depressed for a whole week** | ukb-b-6848 | 0.1484 | 0.04074 | 0.0006 |
| **Cough on most days** | ukb-b-16858 | 0.1483 | 0.05386 | 0.011 |
| **Diagnoses - main ICD-10: R55 Syncope and collapse** | ukb-b-16187 | 0.148 | 0.0645 | 0.036 |
| **Time spent driving** | ukb-b-3793 | 0.1459 | 0.03543 | 0.0001 |
| **Suffer from ‘nerves’** | ukb-b-19957 | 0.1453 | 0.03408 | 5.66e-05 |
| **Length of working week for main job** | ukb-b-8013 | 0.1435 | 0.04645 | 0.004 |
| **Duration of vigorous activity** | ukb-b-13932 | 0.1426 | 0.0408 | 0.001 |
| **Alcohol intake frequency** | ukb-b-5779 | 0.1426 | 0.02293 | 2.62e-09 |
| **Number of live births** | ukb-b-1209 | 0.1425 | 0.02977 | 5.50e-06 |
| **Frequency of walking for pleasure in last 4 weeks** | ukb-b-4655 | 0.1412 | 0.03298 | 5.30e-05 |
| **Diagnoses - main ICD-10: M10 Gout** | ukb-a-561 | 0.1407 | 0.111 | 0.272 |
| **Fractured/broken bones in the last 5 years** | ukb-b-13346 | 0.138 | 0.03926 | 0.001 |
| **Comparative body size at age 10** | ukb-b-4650 | 0.1374 | 0.02317 | 1.43e-08 |
| **Sensitivity/hurt feelings** | ukb-b-9981 | 0.1371 | 0.02911 | 7.85e-06 |
| **Hearing difficulty/problems with background noise** | ukb-b-18275 | 0.1368 | 0.0274 | 2.04e-06 |
| **Duration of walks** | ukb-b-16998 | 0.1361 | 0.02992 | 1.65e-05 |
| **Vitamin and mineral supplements: Folic acid or Folate (Vitamin B9)** | ukb-b-3563 | 0.1344 | 0.09683 | 0.224 |
| **HOMA-B** | ieu-b-117 | 0.1339 | 0.06558 | 0.064 |
| **Ischemic stroke** | ebi-a-GCST005843 | 0.1334 | 0.04006 | 0.002 |
| **Tinnitus, Yes, now some of the time** | ukb-d-4803_13 | 0.1334 | 0.08961 | 0.190 |
| **Hearing aid user** | ukb-b-19060 | 0.1302 | 0.04883 | 0.014 |
| **Ever had prostate specific antigen (PSA) test** | ukb-b-15695 | 0.128 | 0.0387 | 0.002 |
| **Average weekly intake of other alcoholic drinks** | ukb-b-3831 | 0.1275 | 0.2105 | 0.612 |
| **Diagnoses - main ICD-10: S66 Injury of muscle and tendon at wrist and hand level** | ukb-a-591 | 0.1273 | 0.09657 | 0.252 |
| **Duration of light DIY** | ukb-b-2993 | 0.1268 | 0.03888 | 0.002 |
| **Sodium in urine** | ukb-a-335 | 0.1252 | 0.02768 | 1.86e-05 |
| **Mouth/teeth dental problems: Loose teeth** | ukb-b-12849 | 0.1243 | 0.04745 | 0.016 |
| **Townsend deprivation index at recruitment** | ukb-b-10011 | 0.1235 | 0.03261 | 0.0004 |
| **Squamous cell lung cancer** | ieu-a-967 | 0.1202 | 0.1038 | 0.314 |
| **Weight change compared with 1 year ago** | ukb-b-2094 | 0.1202 | 0.04725 | 0.019 |
| **Diagnoses - main ICD-10: N32 Other disorders of bladder** | ukb-a-575 | 0.1198 | 0.08434 | 0.213 |
| **Fractured bone site(s): Leg** | ukb-b-3798 | 0.1194 | 0.1347 | 0.448 |
| **Creatinine (enzymatic) in the urine** | ukb-a-333 | 0.1178 | 0.02655 | 2.72e-05 |
| **Chronic kidney disease** | ieu-a-1102 | 0.1174 | 0.07639 | 0.175 |
| **Diagnoses - main ICD-10: I80 Phlebitis and thrombophlebitis** | ukb-a-537 | 0.1137 | 0.06562 | 0.121 |

| | | | | |
|---|---|---|---|---|
| **Number of children ever born measurement** | ieu-b-4828 | 0.1126 | 0.06696 | 0.134 |
| **Used an inhaler for chest within last hour** | ukb-b-11188 | 0.1112 | 0.09264 | 0.298 |
| **Forearm bone mineral density** | ieu-a-977 | 0.1098 | 0.1023 | 0.352 |
| **Illness, injury, bereavement, stress in the last 2 years: Serious illness, injury, or assault of a close relative** | ukb-b-9049 | 0.108 | 0.05784 | 0.093 |
| **Daytime dozing/sleeping (narcolepsy)** | ukb-b-5776 | 0.1071 | 0.02811 | 0.0003 |
| **Tinnitus, Yes, now most or all the time** | ukb-d-4803_11 | 0.1071 | 0.05537 | 0.081 |
| **Heel bone mineral density (BMD) T-score, automated** | ukb-b-20124 | 0.104 | 0.02594 | 0.0002 |
| **Doctor diagnosed hayfever or allergic rhinitis** | ukb-b-7178 | 0.1033 | 0.03617 | 0.008 |
| **Guilty feelings** | ukb-b-10169 | 0.1031 | 0.03145 | 0.002 |
| **Why reduced smoking: None of the above** | ukb-b-19752 | 0.1028 | 0.2225 | 0.700 |
| **Mineral and other dietary supplements: Fish oil (including cod liver oil)** | ukb-b-11075 | 0.1024 | 0.0325 | 0.003 |
| **Types of transport used (excluding work): Car/motor vehicle** | ukb-b-16886 | 0.1007 | 0.03278 | 0.004 |
| **Lung adenocarcinoma** | ieu-a-965 | 0.1002 | 0.1299 | 0.512 |
| **Diagnoses - main ICD-10: S09 Other and unspecified injuries of head** | ukb-a-589 | 0.09838 | 0.08634 | 0.322 |
| **Ever taken oral contraceptive pill** | ukb-b-9509 | 0.09814 | 0.04513 | 0.049 |
| **Cancer diagnosed by doctor** | ukb-b-16855 | 0.09713 | 0.05768 | 0.133 |
| **Worrier/anxious feelings** | ukb-b-6519 | 0.09618 | 0.0298 | 0.003 |
| **Headaches for 3+ months** | ukb-b-13092 | 0.09525 | 0.04212 | 0.039 |
| **Non-accidental death in close genetic family** | ukb-b-19722 | 0.09376 | 0.06716 | 0.222 |
| **Diastolic blood pressure, automated reading** | ukb-b-7992 | 0.09332 | 0.02266 | 0.0001 |
| **Mineral and other dietary supplements: Zinc** | ukb-b-13891 | 0.09248 | 0.04737 | 0.078 |
| **Duration of strenuous sports** | ukb-b-1046 | 0.08947 | 0.08951 | 0.390 |
| **Vitamin and mineral supplements: Multivitamins +/- minerals** | ukb-b-16812 | 0.08725 | 0.03723 | 0.032 |
| **Frequency of travelling from home to workplace** | ukb-b-185 | 0.08528 | 0.04487 | 0.087 |
| **Number of cigarettes currently smoked daily (current cigarette smokers)** | ukb-b-469 | 0.08419 | 0.05766 | 0.199 |
| **Sitting height ratio** | ieu-a-1070 | 0.0792 | 0.04958 | 0.156 |
| **Diagnoses - main ICD-10: E04 Other non-toxic goitre** | ukb-a-524 | 0.07919 | 0.07815 | 0.382 |
| **Comparative height size at age 10** | ukb-a-35 | 0.07918 | 0.02399 | 0.002 |
| **Femoral neck bone mineral density** | ieu-a-980 | 0.07917 | 0.04109 | 0.083 |
| **Systemic lupus erythematosus** | ebi-a-GCST003156 | 0.07866 | 0.05133 | 0.176 |
| **Diagnoses - main ICD-10: I48 Atrial fibrillation and flutter** | ukb-b-964 | 0.07756 | 0.04915 | 0.161 |
| **Diagnoses - main ICD-10: L03 Cellulitis** | ukb-a-560 | 0.07584 | 0.09574 | 0.499 |
| **Sitting height** | ukb-b-16881 | 0.07438 | 0.02223 | 0.002 |
| **Reason for reducing amount of alcohol drunk: Financial reasons** | ukb-d-2664_4 | 0.07341 | 0.08587 | 0.464 |
| **Nap during day** | ukb-b-4616 | 0.07316 | 0.02342 | 0.004 |
| **Eye problems/disorders: Cataract** | ukb-b-8329 | 0.07225 | 0.05873 | 0.286 |
| **Loud music exposure frequency** | ukb-b-5436 | 0.06994 | 0.0469 | 0.190 |
| **Mineral and other dietary supplements: Selenium** | ukb-b-19158 | 0.06769 | 0.0702 | 0.409 |
| **Systolic blood pressure, automated reading** | ukb-b-20175 | 0.06732 | 0.02413 | 0.010 |
| **Duration of other exercises** | ukb-b-4512 | 0.06716 | 0.0567 | 0.304 |
| **Multiple sclerosis** | ieu-b-18 | 0.06693 | 0.03586 | 0.093 |
| **Handedness (chirality/laterality): Right-handed** | ukb-d-1707_1 | 0.06594 | 0.05767 | 0.321 |
| **Extreme height** | ieu-a-86 | 0.06538 | 0.03967 | 0.142 |
| **Pulse rate, automated reading** | ukb-b-18103 | 0.06127 | 0.02302 | 0.014 |
| **Medication for cholesterol, blood pressure, diabetes or take exogenous hormones: Insulin** | ukb-b-14070 | 0.06009 | 0.08896 | 0.569 |
| **Vitamin and mineral supplements: Vitamin C** | ukb-b-15175 | 0.06003 | 0.05097 | 0.307 |
| **Had menopause** | ukb-b-18105 | 0.05961 | 0.04721 | 0.272 |
| **Urinary albumin-to-creatinine ratio** | ieu-a-1107 | 0.05954 | 0.07012 | 0.466 |
| **Serum creatinine** | bbj-a-61 | 0.05629 | 0.03415 | 0.142 |
| **Wears glasses or contact lenses** | ukb-b-14576 | 0.05596 | 0.04624 | 0.294 |
| **Primary biliary cirrhosis** | ebi-a-GCST005581 | 0.05485 | 0.09256 | 0.621 |
| **Average weekly beer plus cider intake** | ukb-b-5174 | 0.05434 | 0.03398 | 0.156 |
| **Diagnoses - main ICD-10: H26 Other cataract** | ukb-a-530 | 0.05375 | 0.06039 | 0.448 |
| **Diagnoses - main ICD-10: N20 Calculus of kidney and ureter** | ukb-a-574 | 0.05303 | 0.07094 | 0.525 |
| **Diagnoses - main ICD-10: I83 Varicose veins of lower extremities** | ukb-a-538 | 0.05231 | 0.04598 | 0.322 |
| **Reason for glasses/contact lenses: For a ‘lazy’ eye or an eye with poor vision since childhood (called ‘amblyopia’)** | ukb-b-9139 | 0.05117 | 0.08338 | 0.607 |
| **Circulating leptin levels** | ebi-a-GCST003367 | 0.05094 | 0.05746 | 0.448 |
| **Type of tobacco previously smoked: Manufactured cigarettes** | ukb-d-2877_1 | 0.0508 | 0.07762 | 0.581 |
| **Vitamin and mineral supplements: Vitamin D** | ukb-b-12648 | 0.04979 | 0.06548 | 0.518 |
| **Bipolar disorder** | ieu-b-41 | 0.04951 | 0.02938 | 0.133 |
| **C-reactive protein** | bbj-a-14 | 0.04877 | 0.09683 | 0.674 |
| **Distance between home and workplace** | ukb-b-20455 | 0.04753 | 0.06883 | 0.560 |
| **Platelet count** | ukb-d-30080_irnt | 0.0472 | 0.02128 | 0.044 |
| **Offspring birth weight** | ebi-a-GCST005314 | 0.04685 | 0.04316 | 0.346 |
| **Diagnoses - main ICD-10: H26.9 Cataract, unspecified** | ukb-b-12397 | 0.04672 | 0.05476 | 0.464 |
| **Cerebral aneurysm** | bbj-a-96 | 0.04643 | 0.05856 | 0.499 |
| **Difference in height between childhood and adulthood** | ieu-a-1037 | 0.04638 | 0.06269 | 0.529 |

| | | | | |
|---|---|---|---|---|
| **HbA1c** | ieu-b-4842 | 0.04572 | 0.04479 | 0.378 |
| **Mouth/teeth dental problems: Mouth ulcers** | ukb-b-6458 | 0.04534 | 0.03238 | 0.220 |
| **Main ICD-10: D12 Benign neoplasm of colon, rectum, anus, and anal canal** | ukb-d-D12 | 0.04429 | 0.05291 | 0.473 |
| **Number of unsuccessful stop-smoking attempts** | ukb-b-4801 | 0.04343 | 0.04976 | 0.456 |
| **Diagnoses - main ICD-10: H25 Senile cataract** | ukb-a-529 | 0.04242 | 0.1048 | 0.731 |
| **Diagnoses - main ICD-10: K50 Crohn's disease [regional enteritis]** | ukb-a-552 | 0.04179 | 0.08783 | 0.693 |
| **Platelet crit** | ukb-d-30090_irnt | 0.04057 | 0.02228 | 0.101 |
| **Family relationship satisfaction** | ukb-b-3666 | 0.04006 | 0.03337 | 0.297 |
| **Diagnoses - main ICD-10: D25 Leiomyoma of uterus** | ukb-a-522 | 0.03965 | 0.06031 | 0.579 |
| **Forced expiratory volume in 1-second (FEV1) , predicted** | ukb-b-8428 | 0.03778 | 0.03093 | 0.289 |
| **Nervous feelings** | ukb-b-20544 | 0.0364 | 0.03145 | 0.314 |
| **Number of days/week of moderate physical activity 10+ minutes** | ukb-b-4710 | 0.03618 | 0.03084 | 0.308 |
| **Number of incorrect matches in round** | ukb-b-20498 | 0.03595 | 0.02599 | 0.226 |
| **Diagnoses - main ICD-10: C61 Malignant neoplasm of prostate** | ukb-b-2160 | 0.03195 | 0.076 | 0.722 |
| **Work/job satisfaction** | ukb-b-2105 | 0.03118 | 0.04671 | 0.574 |
| **Inflammatory bowel disease** | ebi-a-GCST004131 | 0.03061 | 0.03342 | 0.436 |
| **Lung function (FEV1/FVC)** | ebi-a-GCST007431 | 0.02682 | 0.02254 | 0.302 |
| **Transferrin** | ieu-a-1052 | 0.02659 | 0.07843 | 0.781 |
| **Alcohol drinker status: Never** | ukb-a-226 | 0.02626 | 0.04579 | 0.631 |
| **Ferritin levels** | ebi-a-GCST005072 | 0.02617 | 0.06198 | 0.722 |
| **Number of pregnancy terminations** | ukb-b-4958 | 0.02551 | 0.04773 | 0.657 |
| **Systolic blood pressure** | ieu-b-38 | 0.02419 | 0.02088 | 0.314 |
| **Frequency of other exercises in last 4 weeks** | ukb-b-3502 | 0.02272 | 0.0467 | 0.687 |
| **Reason for glasses/contact lenses: For a 'squint' or 'turn' in an eye since childhood (called 'strabismus')** | ukb-b-11465 | 0.02246 | 0.07296 | 0.799 |
| **Uric acid** | bbj-a-57 | 0.0221 | 0.02761 | 0.496 |
| **Potassium in urine** | ukb-a-334 | 0.02044 | 0.03224 | 0.594 |
| **Fractured bone site(s): Wrist** | ukb-b-9571 | 0.0188 | 0.06779 | 0.817 |
| **Diastolic blood pressure** | ieu-b-39 | 0.01799 | 0.02164 | 0.477 |
| **Number of days/weeks of vigorous physical activity 10+ minutes** | ukb-b-151 | 0.01773 | 0.02777 | 0.592 |
| **Hand grip strength** | ukb-b-7478 | 0.01671 | 0.02272 | 0.531 |
| **Standing height** | ukb-b-10787 | 0.01637 | 0.02086 | 0.503 |
| **Happiness** | ukb-b-4062 | 0.01437 | 0.03424 | 0.722 |
| **Diagnoses - main ICD-10: K51 Ulcerative colitis** | ukb-a-553 | 0.01424 | 0.07698 | 0.877 |
| **Reason for glasses/contact lenses: For long-sightedness (called 'hypermetropia')** | ukb-b-18189 | 0.01392 | 0.06018 | 0.849 |
| **Eczema** | ieu-a-996 | 0.01207 | 0.06553 | 0.877 |
| **Type of tobacco previously smoked: Hand-rolled cigarettes** | ukb-d-2877_2 | 0.01138 | 0.1056 | 0.926 |
| **Diagnoses - main ICD-10: K40 Inguinal hernia** | ukb-a-549 | 0.01124 | 0.05112 | 0.854 |
| **Reason for reducing amount of alcohol drunk: Other reason** | ukb-d-2664_5 | 0.009846 | 0.04397 | 0.852 |
| **Diagnoses - main ICD-10: M72 Fibroblastic disorders** | ukb-a-572 | 0.007981 | 0.05224 | 0.894 |
| **Underlying (primary) cause of death: ICD-10 C34.9 Bronchus or lung, unspecified** | ukb-d-40001_C349 | 0.007745 | 0.1028 | 0.950 |
| **Worry too long after embarrassment** | ukb-b-13653 | 0.007505 | 0.0289 | 0.829 |
| **Total cholesterol** | ieu-a-301 | 0.006143 | 0.03893 | 0.893 |
| **Diagnoses - main ICD-10: Z47 Other orthopaedic follow-up care** | ukb-a-595 | 0.005589 | 0.08483 | 0.952 |
| **Peak expiratory flow (PEF)** | ukb-b-12019 | 0.005417 | 0.02583 | 0.860 |
| **Apolipoprotein B** | ieu-b-108 | 0.004527 | 0.02631 | 0.885 |
| **Hand grip strength (right)** | ukb-b-10215 | 0.004473 | 0.02308 | 0.871 |
| **Amyotrophic lateral sclerosis** | ebi-a-GCST005647 | 0.002829 | 0.06731 | 0.969 |
| **Frequency of light DIY in last 4 weeks** | ukb-b-4210 | 0.002088 | 0.04373 | 0.966 |
| **Difference in height between adolescence and adulthood** | ieu-a-1040 | 0.001982 | 0.06836 | 0.978 |
| **Number of depression episodes** | ukb-b-1464 | -0.00611 | 0.08342 | 0.950 |
| **Platelet distribution width** | ukb-d-30110_irnt | -0.00867 | 0.02691 | 0.792 |
| **Alzheimer's disease** | ieu-b-2 | -0.01049 | 0.06436 | 0.891 |
| **Birth weight** | ukb-b-13378 | -0.01258 | 0.02386 | 0.659 |
| **Mineral and other dietary supplements: Calcium** | ukb-b-7043 | -0.0161 | 0.0504 | 0.792 |
| **Diagnoses - main ICD-10: E03 Other hyperthyroidism** | ukb-a-523 | -0.01671 | 0.2381 | 0.950 |
| **Diagnoses - main ICD-10: C50 Malignant neoplasm of breast** | ukb-a-519 | -0.01701 | 0.05611 | 0.801 |
| **Reproducibility of spirometry measurement using ERS/ATS criteria** | ukb-b-19066 | -0.01711 | 0.05166 | 0.786 |
| **Friendship satisfaction** | ukb-b-3304 | -0.01741 | 0.03693 | 0.694 |
| **Mean platelet volume** | ebi-a-GCST004599 | -0.01764 | 0.02027 | 0.456 |
| **Number of days/week walked 10+ minutes** | ukb-b-4886 | -0.01806 | 0.03092 | 0.624 |
| **Glaucoma** | bbj-a-121 | -0.01828 | 0.06901 | 0.826 |
| **Schizophrenia** | ieu-a-22 | -0.02027 | 0.02541 | 0.497 |
| **Diagnoses - main ICD-10: F43 Reaction to severe stress and adjustments disorders** | ukb-a-526 | -0.02277 | 0.201 | 0.923 |
| **Birth weight of first child** | ukb-b-3357 | -0.02364 | 0.02856 | 0.478 |
| **Diagnoses - main ICD-10: N19 Unspecified renal failure** | ukb-a-573 | -0.02447 | 0.1021 | 0.843 |
| **Diagnoses -main ICD-10: C44 Other malignant neoplasm of skin** | ukb-a-518 | -0.02504 | 0.05774 | 0.717 |
| **Mean platelet (thrombocyte) volume** | ukb-d-30100_irnt | -0.02601 | 0.01979 | 0.253 |

| | | | | |
|---|---|---|---|---|
| **Infant head circumference** | ieu-a-28 | -0.02848 | 0.06305 | 0.706 |
| **Pallidum volume** | ieu-a-1046 | -0.02874 | 0.06629 | 0.717 |
| **Ferritin** | ieu-a-1050 | -0.02992 | 0.08485 | 0.772 |
| **Forced expiratory volume 1-second (FEV1), Best measure** | ukb-b-11141 | -0.03696 | 0.02462 | 0.186 |
| **Time spent using computer** | ukb-b-4522 | -0.04105 | 0.02322 | 0.113 |
| **Forced expiratory volume in 1-second (FEV1)** | ukb-b-19657 | -0.0428 | 0.02356 | 0.102 |
| **Getting up in the morning** | ukb-b-2772 | -0.04464 | 0.02434 | 0.099 |
| **Neo-conscientiousness** | ieu-a-114 | -0.04494 | 0.1065 | 0.722 |
| **Mouth/teeth dental problems: Bleeding gums** | ukb-b-7872 | -0.04589 | 0.0336 | 0.232 |
| **Type of tobacco previously smoked: Cigars and pipes** | ukb-a-327 | -0.0459 | 0.09557 | 0.690 |
| **Forced vital capacity (FVC), Best measure** | ukb-b-14713 | -0.04689 | 0.02442 | 0.084 |
| **LDL cholesterol** | ieu-b-110 | -0.04904 | 0.02707 | 0.103 |
| **Nucleus accumbens volume** | ieu-a-1042 | -0.04971 | 0.1114 | 0.709 |
| **Forced vital capacity (FVC)** | ukb-b-7953 | -0.05151 | 0.02353 | 0.047 |
| **Mineral and other dietary supplements: Iron** | ukb-b-14863 | -0.05167 | 0.06936 | 0.526 |
| **Autism spectrum disorder** | ieu-a-1185 | -0.05266 | 0.04111 | 0.266 |
| **Fractured bone site(s): Arm** | ukb-b-19255 | -0.05672 | 0.105 | 0.654 |
| **Morning/evening person (chronotype)** | ukb-b-4956 | -0.05691 | 0.02258 | 0.020 |
| **Diagnoses - main ICD-10: K35 Acute appendicitis** | ukb-a-548 | -0.05788 | 0.1269 | 0.703 |
| **Subjective well being** | ieu-a-1009 | -0.06181 | 0.04731 | 0.256 |
| **Forced expiratory volume 1-second (FEV1), predicted percentage** | ukb-b-13405 | -0.06315 | 0.02909 | 0.049 |
| **Putamen volume** | ieu-a-1047 | -0.06383 | 0.05789 | 0.338 |
| **Parkinson’s disease** | ieu-b-7 | -0.06416 | 0.04234 | 0.182 |
| **Thalamus volume** | ieu-a-1048 | -0.06972 | 0.07702 | 0.441 |
| **Anorexia nervosa** | ieu-a-1186 | -0.07211 | 0.05064 | 0.212 |
| **Caudate volume** | ieu-a-1044 | -0.07359 | 0.0623 | 0.306 |
| **Reason for glasses/contact lenses: For just reading/near work as you are getting old (called ‘presbyopia’)** | ukb-b-16235 | -0.07856 | 0.09162 | 0.463 |
| **Age at menarche** | ieu-b-4822 | -0.08198 | 0.04917 | 0.137 |
| **Mean time to correctly identify matches** | ukb-b-16287 | -0.08454 | 0.02531 | 0.002 |
| **Fracture resulting from simple fall** | ukb-b-15251 | -0.08524 | 0.06992 | 0.290 |
| **Length of menstrual cycle** | ukb-b-3278 | -0.09299 | 0.04312 | 0.050 |
| **Diagnoses - main ICD-10: O75 Other complications of labour and delivery not elsewhere classified** | ukb-a-579 | -0.09448 | 0.09706 | 0.405 |
| **Vitamin and mineral supplements: None of the above** | ukb-b-17381 | -0.09521 | 0.03501 | 0.012 |
| **Relative age of first facial hair** | ukb-b-5945 | -0.09538 | 0.02687 | 0.0009 |
| **Fasting blood glucose adjusted for BMI** | ebi-a-GCST007858 | -0.09635 | 0.1085 | 0.448 |
| **Mineral and other dietary supplements: None of the above** | ukb-b-14238 | -0.1002 | 0.03205 | 0.004 |
| **Diagnoses - main ICD-10: Z80 Family history of malignant neoplasm** | ukb-a-596 | -0.1051 | 0.1798 | 0.624 |
| **Celiac disease** | ebi-a-GCST000612 | -0.111 | 0.06049 | 0.099 |
| **Adiponectin** | ieu-a-1 | -0.1113 | 0.09904 | 0.328 |
| **Handedness (chirality/laterality): Left-handed** | ukb-a-36 | -0.1159 | 0.05793 | 0.071 |
| **Primary sclerosing cholangitis** | ieu-a-1112 | -0.116 | 0.06281 | 0.097 |
| **Underlying (primary) cause of death: ICD-10 E85.4 Organ limited amyloidosis** | ukb-a-357 | -0.1165 | 0.1516 | 0.513 |
| **Reason for glasses/contact lenses: For ‘astigmatism’** | ukb-b-10937 | -0.1169 | 0.05444 | 0.051 |
| **Hippocampus volume** | ieu-a-1045 | -0.1174 | 0.07937 | 0.194 |
| **Circulating leptin levels adjusted for BMI** | ebi-a-GCST003368 | -0.1182 | 0.06261 | 0.089 |
| **Types of physical activity in the last 4 weeks: Heavy DIY** | ukb-b-13184 | -0.1209 | 0.03288 | 0.0005 |
| **ICD-10:H40 Glaucoma** | ukb-d-H40 | -0.1261 | 0.09974 | 0.272 |
| **Average weekly red wine intake** | ukb-b-5239 | -0.1266 | 0.03039 | 8.55e-05 |
| **Snoring** | ukb-b-17400 | -0.1289 | 0.02703 | 6.01e-06 |
| **Medication for cholesterol, blood pressure, diabetes or take exogenous hormones: Oral contraceptive pill or minipill** | ukb-b-9547 | -0.1313 | 0.09084 | 0.204 |
| **Target heart rate achieved** | ukb-b-16609 | -0.1325 | 0.06173 | 0.051 |
| **Time from waking to first cigarette** | ukb-b-2732 | -0.1334 | 0.05758 | 0.034 |
| **Average weekly champagne plus white wine intake** | ukb-b-5716 | -0.1334 | 0.03629 | 0.0006 |
| **Smoking cessation** | bbj-a-81 | -0.1379 | 0.05149 | 0.013 |
| **Apolipoprotein A-I** | ieu-b-107 | -0.1414 | 0.02614 | 2.50e-07 |
| **Father still alive** | ukb-b-13160 | -0.1431 | 0.05824 | 0.024 |
| **Breastfed as a baby** | ukb-b-13423 | -0.1435 | 0.03765 | 0.0003 |
| **Age at menarche** | ieu-a-1095 | -0.1444 | 0.02859 | 1.58e-06 |
| **Childhood intelligence** | ieu-a-16 | -0.1468 | 0.06337 | 0.034 |
| **Types of physical activity in the last 4 weeks: Other exercises** | ukb-b-8764 | -0.1468 | 0.02738 | 3.20e-07 |
| **Serum cystatin C (eGFRcys)** | ieu-a-1106 | -0.1475 | 0.06321 | 0.033 |
| **Reason for glasses/contact lenses: For short-sightedness (called ‘myopia’)** | ukb-b-6353 | -0.1485 | 0.03691 | 0.0001 |
| **Frequency of stair climbing in the last 4 weeks** | ukb-b-298 | -0.149 | 0.03157 | 7.49e-06 |
| **Types of physical activity in the last 4 weeks: Light DIY** | ukb-b-11495 | -0.1502 | 0.03017 | 2.19e-06 |
| **Relative age voice broke** | ukb-b-5262 | -0.1512 | 0.03396 | 2.55e-05 |
| **Neo-openness to experience** | ieu-a-117 | -0.1525 | 0.07922 | 0.083 |
| **Diagnoses - main ICD-10: F31 Bipolar affective disorder** | ukb-a-525 | -0.1526 | 0.09155 | 0.137 |

| | | | | |
|---|---|---|---|---|
| **Diagnoses - main ICD-10: S52 Fracture of forearm** | ukb-a-590 | -0.1568 | 0.07432 | 0.056 |
| **Mouth/teeth dental problems: None of the above** | ukb-b-17601 | -0.1584 | 0.03055 | 7.99e-07 |
| **Average weekly fortified wine intake** | ukb-b-1070 | -0.1603 | 0.05517 | 0.007 |
| **Types of transport used (excluding work): Public transport** | ukb-b-15957 | -0.1625 | 0.03697 | 3.25e-05 |
| **Lung cancer** | bbj-a-133 | -0.1646 | 0.1237 | 0.247 |
| **ECG, heart rate** | ukb-b-11361 | -0.1655 | 0.0449 | 0.0005 |
| **Age at menopause (last menstrual period)** | ukb-b-17422 | -0.1659 | 0.02728 | 5.94e-09 |
| **Fluid intelligence score** | ukb-b-5238 | -0.166 | 0.02631 | 1.53e-09 |
| **Sleep duration** | ukb-b-4424 | -0.1675 | 0.02803 | 1.11e-08 |
| **Types of physical activity in last 4 weeks: Walking for pleasure** | ukb-b-7337 | -0.1689 | 0.03378 | 1.99e-06 |
| **Types of physical activity in last 4 weeks: Strenuous sports** | ukb-b-7663 | -0.1703 | 0.03473 | 3.13e-06 |
| **Intelligence** | ebi-a-GCST006250 | -0.1713 | 0.02216 | 8.32e-14 |
| **Tinnitus, No, never** | ukb-d-4803_0 | -0.1768 | 0.04258 | 9.05e-05 |
| **Types of transport used (excluding work): Cycle** | ukb-b-17557 | -0.1785 | 0.03327 | 3.18e-07 |
| **HDL cholesterol** | ieu-b-109 | -0.1824 | 0.02405 | 2.46e-13 |
| **Alcohol usually taken with meals** | ukb-b-16878 | -0.1868 | 0.02924 | 9.45e-10 |
| **Impedance of leg (left)** | ukb-b-14068 | -0.1874 | 0.02057 | 8.50e-19 |
| **Qualifications: Other professional qualifications e.g., nursing, teaching** | ukb-b-13799 | -0.1893 | 0.03094 | 4.77e-09 |
| **Impedance of leg (right)** | ukb-b-7376 | -0.1926 | 0.01997 | 6.12e-21 |
| **Duration walking for pleasure** | ukb-b-703 | -0.198 | 0.03999 | 2.49e-06 |
| **Types of transport used (excluding work): Walk** | ukb-b-14926 | -0.1986 | 0.03351 | 1.46e-08 |
| **Past tobacco smoking** | ukb-b-2134 | -0.2033 | 0.0239 | 1.639e-16 |
| **Reason for glasses/contact lenses: Other eye condition** | ukb-b-15297 | -0.209 | 0.1863 | 0.329 |
| **Transport type for commuting to workplace: Walk** | ukb-b-15005 | -0.2099 | 0.05665 | 0.0005 |
| **Former vs current smoker** | ieu-a-963 | -0.2132 | 0.06222 | 0.001 |
| **Underlying (primary) cause of death: ICD-10 J84.1 Other interstitial pulmonary diseases with fibrosis** | ukb-a-358 | -0.222 | 0.151 | 0.196 |
| **Maximum workload during fitness test** | ukb-b-7551 | -0.222 | 0.05436 | 0.0001 |
| **Alcohol drinker status: Current** | ukb-d-20117_2 | -0.2226 | 0.04012 | 1.21e-07 |
| **Started insulin within one year diagnosis of diabetes** | ukb-b-8388 | -0.2308 | 0.0789 | 0.007 |
| **Transport type for commuting to workplace: Cycle** | ukb-b-15787 | -0.2329 | 0.03618 | 6.97e-10 |
| **Transport type for commuting to workplace: Public transport** | ukb-b-17155 | -0.2347 | 0.03922 | 1.07e-08 |
| **Smoking status: Never** | ukb-d-20116_0 | -0.2386 | 0.02478 | 7.13e-21 |
| **Qualifications: O levels/GCSEs or equivalent** | ukb-b-18099 | -0.2447 | 0.02971 | 1.46e-15 |
| **Vascular/heart problems diagnosed by doctor: None of the above** | ukb-b-13352 | -0.2457 | 0.02317 | 4.30e-25 |
| **Pulse wave peak to peak time** | ukb-b-8778 | -0.2469 | 0.04531 | 2.05e-07 |
| **Impedance of whole body** | ukb-b-19921 | -0.2556 | 0.02064 | 7.70e-34 |
| **Why reduced smoking: Health precaution** | ukb-b-13026 | -0.2562 | 0.1216 | 0.056 |
| **Age of smoking initiation** | ieu-b-24 | -0.259 | 0.03105 | 6.37e-16 |
| **Years of schooling** | ieu-a-1239 | -0.2629 | 0.01929 | 1.39e-40 |
| **College completion** | ieu-a-836 | -0.2667 | 0.04041 | 2.45e-10 |
| **Qualifications: College or University degree** | ukb-b-16489 | -0.2679 | 0.02191 | 5.28e-33 |
| **Mother's age at death** | ukb-b-12687 | -0.2702 | 0.04147 | 4.25e-10 |
| **Illness, injury, bereavement, stress in the last 2 years: None of the above** | ukb-b-17687 | -0.2722 | 0.03142 | 4.36e-17 |
| **Age completed full time education** | ukb-b-6134 | -0.2722 | 0.02826 | 7.04e-21 |
| **Maximum heart rate during fitness test** | ukb-b-14461 | -0.2746 | 0.05061 | 2.33e-07 |
| **Qualifications: A levels/ AS levels or equivalent** | ukb-b-11615 | -0.2759 | 0.02525 | 1.35e-26 |
| **Father's age at death** | ukb-b-11303 | -0.2784 | 0.03921 | 8.12e-12 |
| **Impedance of arm (right)** | ukb-b-7859 | -0.2824 | 0.02113 | 3.82e-39 |
| **Usual walking pace** | ukb-b-4711 | -0.2833 | 0.02477 | 5.32e-29 |
| **Impedance of arm (left)** | ukb-b-19379 | -0.2844 | 0.02141 | 1.03e-38 |
| **Parental longevity** | ebi-a-GCST006702 | -0.2877 | 0.03995 | 4.02e-12 |
| **Medication for cholesterol, blood pressure, diabetes or take exogenous hormones: None of the above** | ukb-b-20379 | -0.2901 | 0.03107 | 1.09e-19 |
| **Duration of fitness test** | ukb-b-6011 | -0.301 | 0.09015 | 0.002 |
| **Age started oral contraceptive pill** | ukb-b-9433 | -0.3324 | 0.03679 | 1.65e-18 |
| **Age at first live birth** | ukb-b-12405 | -0.3432 | 0.02573 | 5.27e-39 |
| **Age at last live birth** | ukb-b-8727 | -0.3554 | 0.03246 | 1.08e-26 |
| **Medication for pain relief, constipation, heartburn: No medication** | ukb-b-11085 | -0.4178 | 0.02462 | 2.02e-62 |
| **Pain types experienced in last month: None of the above** | ukb-b-9130 | -0.5107 | 0.02724 | 7.48e-76 |

**Table S10.** Exposures potentially causal for LSS, and outcomes that LSS was potentially found to be causal for. Mendelian randomization was performed by using TwoSampleMR-database and the phenotype IDs used in the analysis were extracted from the MRC-IEU database. For the analysis, we extracted genetic instruments from FinnGen+EstBB based meta-analysis results. Inverse variance weighted-model was used as a main method, but MR Egger was also utilised as a sensitivity analysis. P<0.05 was used as a threshold for statistical significance. nsnp, number of SNPs; OR, odds ratio, pFDR, false discovery rate-corrected p-value; pHET, p-value for heterogeneity; pPLE, p-value for pleiotropy

| Direction | Trait | Method | nsnp | Causal estimate scale | Causal estimate | pFDR | pHET | Egger intercept | pPLE |
|---|---|---|---|---|---|---|---|---|---|
| **Trait-> SS** | Whole body fat-free mass \|\| id:ukb-b-13354 | IVW | 481 | OR | 1.78 (1.59–1.99) | 2.22e-21 | 1.53e-46 | | |
| | Whole body fat-free mass \|\| id:ukb-b-13354 | MR Egger | 481 | OR | 1.53 (1.17–2.00) | 0.004 | 2.53e-46 | 0.0023 | 0.2255 |
| **Trait-> SS** | Hip circumference \|\| id:ukb-b-15590 | IVW | 369 | OR | 1.51 (1.38–1.64) | 2.62e-18 | 1.05e-26 | | |
| | Hip circumference \|\| id:ukb-b-15590 | MR Egger | 369 | OR | 1.60 (1.26–2.03) | 0.0004 | 8.49e-27 | -0.0012 | 0.6042 |
| **Trait-> SS** | Whole body fat mass \|\| id:ukb-b-19393 | IVW | 383 | OR | 1.48 (1.35–1.63) | 2.82e-15 | 3.46e-35 | | |
| | Whole body fat mass \|\| id:ukb-b-19393 | MR Egger | 383 | OR | 1.66 (1.28–2.16) | 0.0004 | 3.86e-35 | -0.0022 | 0.3585 |
| **Trait-> SS** | Body mass index (BMI) \|\| id:ukb-b-19953 | IVW | 387 | OR | 1.55 (1.42–1.70) | 2.22e-21 | 8.53e-26 | | |
| | Body mass index (BMI) \|\| id:ukb-b-19953 | MR Egger | 387 | OR | 1.60 (1.26–2.03) | 0.0004 | 6.31e-26 | -0.0006 | 0.7846 |
| **SS-> Trait** | Back pain \|\| id:ukb-b-983 | IVW | 33 | Beta | 0.02 (0.01-0.03) | 0.0001 | 2.46e-14 | | |
| | Back pain \|\| id:ukb-b-983 | MR Egger | 33 | Beta | 0.03 (-0.02-0.07) | 0.581 | 1.18e-14 | -0.00017 | 0.9247 |

**Table S11**. Risk factors that were extracted from the GWAS database provided by the MRC Integrative Epidemiology Unit (IEU) (https://gwas.mrcieu.ac.uk/) and were used in the bi-directional mendelian randomization.

| GWAS-ID | Year | Trait | Consortium | Sample size | Number of SNP's |
|---|---|---|---|---|---|
| **ieu-b-35** | 2018 | C-reactive protein level | NA | 204402 | 2414379 |
| **met-d-GlycA** | 2020 | Glycoprotein acetyls | NA | 115078 | 12321875 |
| **ukb-b-13354** | 2018 | Whole body fat-free mass | MRC-IEU | 454850 | 9851867 |
| **ukb-b-15590** | 2018 | Hip circumference | MRC-IEU | 462117 | 9851867 |
| **ukb-b-19393** | 2018 | Whole body fat mass | MRC-IEU | 454137 | 9851867 |
| **ukb-b-19953** | 2018 | Body mass index (BMI) | MRC-IEU | 461460 | 9851867 |
| **ukb-b-3957** | 2018 | Sleeplessness/insomnia | MRC-IEU | 462341 | 9851867 |
| **ukb-b-929** | 2018 | Frequency of tiredness/lethargy in last 2 weeks | MRC-IEU | 449019 | 9851867 |
| **ukb-b-9838** | 2018 | Back pain | MRC-IEU | 461857 | 9851867 |

### References


1. de Leeuw, C. A., Mooij, J. M., Heskes, T. & Posthuma, D. MAGMA: Generalized Gene-Set Analysis of GWAS Data. *PLoS Comput Biol* **11**, e1004219 (2015).
2. Watanabe, K., Taskesen, E., van Bochoven, A. & Posthuma, D. Functional mapping and annotation of genetic associations with FUMA. *Nat Commun* **8**, 1826 (2017).
3. Sakaue, S. *et al.* A cross-population atlas of genetic associations for 220 human phenotypes. *Nat Genet* **53**, 1415–1424 (2021).
4. Suri, P. *et al.* Genome-wide association studies of low back pain and lumbar spinal disorders using electronic health record data identify a locus associated with lumbar spinal stenosis. *Pain* **162**, 2263–2272 (2021).
5. Clark, K., Karsch-Mizrachi, I., Lipman, D. J., Ostell, J. & Sayers, E. W. GenBank. *Nucleic Acids Res* **44**, D67–D72 (2016).
6. UniProt: a worldwide hub of protein knowledge. *Nucleic Acids Res* **47**, D506–D515 (2019).
7. Stanfill, A. G. & Cao, X. Enhancing Research Through the Use of the Genotype-Tissue Expression (GTEx) Database. *Biol Res Nurs* **23**, 533–540 (2021).
8. Cortes, A. *et al.* Identification of multiple risk variants for ankylosing spondylitis through high-density genotyping of immune-related loci. *Nat Genet* **45**, 730–738 (2013).
9. Abe, Y. *et al. Proinflammatory Cytokines Stimulate the Expression of Nerve Growth Factor by Human Intervertebral Disc Cells*. *SPINE* vol. 32.
10. Freemont, A. J. *et al.* Nerve growth factor expression and innervation of the painful intervertebral disc. *Journal of Pathology* **197**, 286–292 (2002).
11. Hu, H. *et al.* OTUD7B controls non-canonical NF-κB activation through deubiquitination of TRAF3. *Nature* **494**, 371–374 (2013).
12. Zhang, H., Liu, D. & Fan, X. Diagnostic and prognostic significance of miR-486-5p in patients who underwent minimally invasive surgery for lumbar spinal stenosis. *European Spine Journal* **33**, 1979–1985 (2024).
13. Russo, S. *et al.* A mutation in the ZNF687 gene that is responsible for the severe form of Paget's disease of bone causes severely altered bone remodeling and promotes hepatocellular carcinoma onset in a knock-in mouse model. *Bone Res* **11**, 16 (2023).
14. Divisato, G. *et al.* ZNF687 Mutations in Severe Paget Disease of Bone Associated with Giant Cell Tumor. *Am J Hum Genet* **98**, 275–86 (2016).
15. Varela, D., Varela, T., Conceição, N. & Cancela, M. L. Regulation of human ZNF687, a gene associated with Paget's disease of bone. *Int J Biochem Cell Biol* **154**, 106332 (2023).
16. Jacobsen, M. *et al.* A point mutation in PTPRC is associated with the development of multiple sclerosis. *Nat Genet* **26**, 495–499 (2000).
17. Dunlock, V.-M. E. *et al.* Tetraspanin CD53 controls T cell immunity through regulation of CD45RO stability, mobility, and function. *Cell Rep* **39**, 111006 (2022).
18. Luo, W., Zhang, G., Wang, Z., Wu, Y. & Xiong, Y. Ubiquitin-specific proteases: Vital regulatory molecules in bone and bone-related diseases. *Int Immunopharmacol* **118**, 110075 (2023).
19. Sun, C., Ma, Q., Yin, J., Zhang, H. & Liu, X. WISP-1 induced by mechanical stress contributes to fibrosis and hypertrophy of the ligamentum flavum through Hedgehog-Gli1 signaling. *Exp Mol Med* **53**, 1068–1079 (2021).
20. Qin, T. *et al.* Abnormally elevated USP37 expression in breast cancer stem cells regulates stemness, epithelial-mesenchymal transition and cisplatin sensitivity. *J Exp Clin Cancer Res* **37**, 287 (2018).

21. Wright, K. M. *et al.* Dystroglycan organizes axon guidance cue localization and axonal pathfinding. *Neuron* **76**, 931–44 (2012).
22. Jahncke, J. N. & Wright, K. M. The many roles of dystroglycan in nervous system development and function. *Developmental Dynamics* **252**, 61–80 (2023).
23. Dai, Y. *et al.* Whole exome sequencing identified a novel DAG1 mutation in a patient with rare, mild and late age of onset muscular dystrophy-dystroglycanopathy. *J Cell Mol Med* **23**, 811–818 (2019).
24. Wang, F. M. *et al.* Aging of the Spine: Characterizing genetic and physiological determinants of spinal curvature. doi:10.1101/2024.02.27.24303450.
25. Veleri, S., Punnakkal, P., Dunbar, G. L. & Maiti, P. Molecular Insights into the Roles of Rab Proteins in Intracellular Dynamics and Neurodegenerative Diseases. *Neuromolecular Med* **20**, 18–36 (2018).
26. Jiang, J. *et al.* Involvement of Rab28 in NF-κB Nuclear Transport in Endothelial Cells. *PLoS One* **8**, (2013).
27. Prunier, C. & Howe, P. H. Disabled-2 (Dab2) Is Required for Transforming Growth Factor β-induced Epithelial to Mesenchymal Transition (EMT). *Journal of Biological Chemistry* **280**, 17540–17548 (2005).
28. Figliuolo da Paz, V., Ghishan, F. K. & Kiela, P. R. Emerging Roles of Disabled Homolog 2 (DAB2) in Immune Regulation. *Frontiers in Immunology* vol. 11 Preprint at https://doi.org/10.3389/fimmu.2020.580302 (2020).
29. Wang, D. *et al.* Cyclic Mechanical Stretch Ameliorates the Degeneration of Nucleus Pulposus Cells through Promoting the ITGA2/PI3K/AKT Signaling Pathway. *Oxid Med Cell Longev* **2021**, 1–11 (2021).
30. Nechanitzky, R. *et al.* Transcription factor EBF1 is essential for the maintenance of B cell identity and prevention of alternative fates in committed cells. *Nat Immunol* **14**, 867–875 (2013).
31. Yang, C.-Y. *et al.* Interaction of CCR4-NOT with EBF1 regulates gene-specific transcription and mRNA stability in B lymphopoiesis. *Genes Dev* **30**, 2310–2324 (2016).
32. Kugimiya, F. *et al.* Involvement of endogenous bone morphogenetic protein (BMP) 2 and BMP6 in bone formation. *Journal of Biological Chemistry* **280**, 35704–35712 (2005).
33. Ebisawa, T. *et al.* Characterization of bone morphogenetic protein-6 signaling pathways in osteoblast differentiation. *J Cell Sci* **112 ( Pt 20)**, 3519–27 (1999).
34. Crews, L. *et al.* Increased BMP6 levels in the brains of Alzheimer's disease patients and APP transgenic mice are accompanied by impaired neurogenesis. *Journal of Neuroscience* **30**, 12252–12262 (2010).
35. Pulkkinen, H. H. *et al.* BMP6/TAZ-Hippo signaling modulates angiogenesis and endothelial cell response to VEGF. *Angiogenesis* **24**, 129–144 (2021).
36. Hu, M.-M. & Shu, H.-B. Multifaceted roles of TRIM38 in innate immune and inflammatory responses. *Cell Mol Immunol* **14**, 331–338 (2017).
37. Hu, S., Li, Y., Wang, B. & Peng, K. TRIM38 protects chondrocytes from IL-1β-induced apoptosis and degeneration via negatively modulating nuclear factor (NF)-κB signaling. *Int Immunopharmacol* **99**, (2021).
38. Koike, Y. *et al.* Genetic insights into ossification of the posterior longitudinal ligament of the spine. *Elife* **12**, (2023).
39. Nakajima, M. *et al.* A genome-wide association study identifies susceptibility loci for ossification of the posterior longitudinal ligament of the spine. *Nat Genet* **46**, 1012–1016 (2014).

40. Jokoji, G. *et al.* CDC5L promotes early chondrocyte differentiation and proliferation by modulating pre-mRNA splicing of SOX9, COL2A1, and WEE1. *Journal of Biological Chemistry* **297**, 100994 (2021).
41. Kim, S. K. Identification of 613 new loci associated with heel bone mineral density and a polygenic risk score for bone mineral density, osteoporosis and fracture. *PLoS One* **13**, e0200785 (2018).
42. Kemp, J. P., Medina-Gomez, C., Tobias, J. H., Rivadeneira, F. & Evans, D. M. The case for genome-wide association studies of bone acquisition in paediatric and adolescent populations. *Bonekey Rep* **5**, 796 (2016).
43. Estrada, K. *et al.* Genome-wide meta-analysis identifies 56 bone mineral density loci and reveals 14 loci associated with risk of fracture. *Nat Genet* **44**, 491–501 (2012).
44. Bian, Q. *et al.* Excessive Activation of TGFβ by Spinal Instability Causes Vertebral Endplate Sclerosis. *Sci Rep* **6**, 27093 (2016).
45. Hegewald, A. A. *et al.* Effects of initial boost with TGF-beta 1 and grade of intervertebral disc degeneration on 3D culture of human annulus fibrosus cells. *J Orthop Surg Res* **9**, 73 (2014).
46. Pham, D., Vincentz, J. W., Firulli, A. B. & Kaplan, M. H. Twist1 Regulates *Ifng* Expression in Th1 Cells by Interfering with Runx3 Function. *The Journal of Immunology* **189**, 832–840 (2012).
47. Salo, V. *et al.* Genome-wide meta-analysis conducted in three large biobanks expands the genetic landscape of lumbar disc herniations. doi:10.1101/2023.10.15.23296916.
48. Bjornsdottir, G. *et al.* Rare SLC13A1 variants associate with intervertebral disc disorder highlighting role of sulfate in disc pathology. *Nat Commun* **13**, 634 (2022).
49. Soranzo, N. *et al.* Meta-Analysis of Genome-Wide Scans for Human Adult Stature Identifies Novel Loci and Associations with Measures of Skeletal Frame Size. *PLoS Genet* **5**, e1000445 (2009).
50. Johansson, Å. *et al.* Common variants in the JAZF1 gene associated with height identified by linkage and genome-wide association analysis. *Hum Mol Genet* **18**, 373–380 (2009).
51. Nakamichi, R. & Asahara, H. The transcription factors regulating intervertebral disc development. *JOR Spine* **3**, (2020).
52. Nakamichi, R. *et al.* Mohawk promotes the maintenance and regeneration of the outer annulus fibrosus of intervertebral discs. *Nat Commun* **7**, 12503 (2016).
53. Shen, L. *et al.* miR-448 downregulates MPPED2 to promote cancer proliferation and inhibit apoptosis in oral squamous cell carcinoma. *Exp Ther Med* **12**, 2747–2752 (2016).
54. Schimunek, L. *et al.* MPPED2 Polymorphism Is Associated With Altered Systemic Inflammation and Adverse Trauma Outcomes. *Front Genet* **10**, (2019).
55. Liu, C. F. & Lefebvre, V. The transcription factors SOX9 and SOX5/SOX6 cooperate genome-wide through super-enhancers to drive chondrogenesis. *Nucleic Acids Res* **43**, 8183–8203 (2015).
56. Lefebvre, V., Li, P. & de Crombrugghe, B. A new long form of Sox5 (L-Sox5), Sox6 and Sox9 are coexpressed in chondrogenesis and cooperatively activate the type II collagen gene. *EMBO J* **17**, 5718–33 (1998).
57. Han, Y. & Lefebvre, V. L-Sox5 and Sox6 Drive Expression of the Aggrecan Gene in Cartilage by Securing Binding of Sox9 to a Far-Upstream Enhancer. *Mol Cell Biol* **28**, 4999–5013 (2008).
58. Nakajima, M., Koido, M., Guo, L., Terao, C. & Ikegawa, S. A novel CCDC91 isoform associated with ossification of the posterior longitudinal ligament of the spine works as

a non-coding RNA to regulate osteogenic genes. *The American Journal of Human Genetics* **110**, 638–647 (2023).
59. Tan, Z. *et al.* Synergistic co-regulation and competition by a SOX9-GLI-FOXA phasic transcriptional network coordinate chondrocyte differentiation transitions. *PLoS Genet* **14**, e1007346 (2018).
60. Inui, M. *et al.* USP15 is a deubiquitylating enzyme for receptor-activated SMADs. *Nat Cell Biol* **13**, 1368–1375 (2011).
61. Meola, N. *et al.* Identification of a Nuclear Exosome Decay Pathway for Processed Transcripts. *Mol Cell* **64**, 520–533 (2016).
62. Najmabadi, H. *et al.* Deep sequencing reveals 50 novel genes for recessive cognitive disorders. *Nature* **478**, 57–63 (2011).
63. Revy, P., Kannengiesser, C. & Bertuch, A. A. Genetics of human telomere biology disorders. *Nat Rev Genet* **24**, 86–108 (2023).
64. Itoh, Y. *et al.* A common SNP risk variant MT1-MMP causative for Dupuytren's disease has a specific defect in collagenolytic activity. *Matrix Biology* **97**, 20–39 (2021).
65. Kosaka, H. *et al.* Pathomechanism of Loss of Elasticity and Hypertrophy of Lumbar Ligamentum Flavum in Elderly Patients With Lumbar Spinal Canal Stenosis. *Spine (Phila Pa 1976)* **32**, 2805–2811 (2007).
66. Osthues, T. *et al.* The Lipid Receptor G2A (GPR132) Mediates Macrophage Migration in Nerve Injury-Induced Neuropathic Pain. *Cells* **9**, (2020).
67. Styrkarsdottir, U. *et al.* Severe osteoarthritis of the hand associates with common variants within the ALDH1A2 gene and with rare variants at 1p31. *Nat Genet* **46**, 498–502 (2014).
68. Chu, M. *et al.* The rs4238326 polymorphism in ALDH1A2 gene potentially associated with non-post traumatic knee osteoarthritis susceptibility: a two-stage population-based study. *Osteoarthritis Cartilage* **25**, 1062–1067 (2017).
69. Zhu, L. *et al.* Variants in *ALDH1A2* reveal an anti-inflammatory role for retinoic acid and a new class of disease-modifying drugs in osteoarthritis. *Sci Transl Med* **14**, (2022).
70. Smith, T., Ferreira, L. R., Hebert, C., Norris, K. & Sauk, J. J. Hsp47 and cyclophilin B traverse the endoplasmic reticulum with procollagen into pre-Golgi intermediate vesicles. A role for Hsp47 and cyclophilin B in the export of procollagen from the endoplasmic reticulum. *J Biol Chem* **270**, 18323–8 (1995).
71. van Dijk, F. S. *et al.* PPIB Mutations Cause Severe Osteogenesis Imperfecta. *The American Journal of Human Genetics* **85**, 521–527 (2009).
72. Epstein, P. M., Basole, C. & Brocke, S. The Role of PDE8 in T Cell Recruitment and Function in Inflammation. *Front Cell Dev Biol* **9**, (2021).
73. Wang, H. *et al.* Kinetic and structural studies of phosphodiesterase-8A and implication on the inhibitor selectivity. *Biochemistry* **47**, 12760–8 (2008).
74. Allen, E. *et al.* Gigaxonin-controlled degradation of MAP1B light chain is critical to neuronal survival. *Nature* **438**, 224–228 (2005).
75. Martens, N. *et al.* Suppressor of Cytokine Signaling 7 Inhibits Prolactin, Growth Hormone, and Leptin Signaling by Interacting with STAT5 or STAT3 and Attenuating Their Nuclear Translocation. *Journal of Biological Chemistry* **280**, 13817–13823 (2005).
76. Jho, E. *et al.* Wnt/beta-catenin/Tcf signaling induces the transcription of Axin2, a negative regulator of the signaling pathway. *Mol Cell Biol* **22**, 1172–83 (2002).
77. Houschyar, K. S. *et al.* Wnt Pathway in Bone Repair and Regeneration - What Do We Know So Far. *Front Cell Dev Biol* **6**, 170 (2018).

78. Li, X. *et al.* The adaptor protein Nck-1 couples the netrin-1 receptor DCC (deleted in colorectal cancer) to the activation of the small GTPase Racl through an atypical mechanism. *Journal of Biological Chemistry* **277**, 37788–37797 (2002).
79. Bu, G. *et al.* Increased expression of netrin-1 and its deleted in colorectal cancer receptor in human diseased lumbar intervertebral disc compared with autopsy control. *Spine (Phila Pa 1976)* **37**, 2074–2081 (2012).
80. Cousminer, D. L. *et al.* Genome-wide association study implicates novel loci and reveals candidate effector genes for longitudinal pediatric bone accrual. *Genome Biol* **22**, 1 (2021).
81. Zheng, Z. *et al.* CRLF1 Is a Key Regulator in the Ligamentum Flavum Hypertrophy. *Front Cell Dev Biol* **8**, (2020).
82. Xi, P., Ding, D., Zhou, J., Wang, M. & Cong, Y.-S. DDRGK1 regulates NF-κB activity by modulating IκBα stability. *PLoS One* **8**, e64231 (2013).
83. Egunsola, A. T. *et al.* Loss of DDRGK1 modulates SOX9 ubiquitination in spondyloepimetaphyseal dysplasia. *J Clin Invest* **127**, 1475–1484 (2017).
84. Raouf, A. & Seth, A. Ets transcription factors and targets in osteogenesis. *Oncogene* **19**, 6455–6463 (2000).
85. Malik, N. *et al.* Molecular cloning, sequence analysis, and functional expression of a novel growth regulator, oncostatin M. *Mol Cell Biol* **9**, 2847–53 (1989).
86. Sims, N. A. & Quinn, J. M. W. Osteoimmunology: oncostatin M as a pleiotropic regulator of bone formation and resorption in health and disease. *Bonekey Rep* **3**, (2014).
87. Liu, T. *et al.* Chordin-Like 1 Improves Osteogenesis of Bone Marrow Mesenchymal Stem Cells Through Enhancing BMP4-SMAD Pathway. *Front Endocrinol (Lausanne)* **10**, (2019).
88. Gao, W.-L., Zhang, S.-Q., Zhang, H., Wan, B. & Yin, Z.-S. Chordin-like protein 1 promotes neuronal differentiation by inhibiting bone morphogenetic protein-4 in neural stem cells. *Mol Med Rep* **7**, 1143–8 (2013).
89. Kurakazu, I. *et al.* FOXO1 transcription factor regulates chondrogenic differentiation through transforming growth factor β1 signaling. *Journal of Biological Chemistry* **294**, 17555–17569 (2019).
90. Yang, J., Lee, S. H., Goddard, M. E. & Visscher, P. M. GCTA: a tool for genome-wide complex trait analysis. *Am J Hum Genet* **88**, 76–82 (2011).
91. Bakshi, A. *et al.* Fast set-based association analysis using summary data from GWAS identifies novel gene loci for human complex traits. *Sci Rep* **6**, (2016).
92. Giambartolomei, C. *et al.* Bayesian test for colocalisation between pairs of genetic association studies using summary statistics. *PLoS Genet* **10**, e1004383 (2014).
93. Martin, F. J. *et al.* Ensembl 2023. *Nucleic Acids Res* **51**, D933–D941 (2023).
94. Sliz, E. *et al.* Evidence of a causal effect of genetic tendency to gain muscle mass on uterine leiomyomata. *Nat Commun* **14**, 542 (2023).
95. Bulik-Sullivan, B. K. *et al.* LD Score regression distinguishes confounding from polygenicity in genome-wide association studies. *Nat Genet* **47**, 291–295 (2015).
